# Metabolomic Risk Predictors of Diabetic Foot Complications: a longitudinal observational study in Type 1 Diabetes

**DOI:** 10.1101/2022.05.16.22275166

**Authors:** Jonas A. Andersen, Tommi Suvitaival, Kajetan Trošt, María José Romero-Lado, Simone Theilade, Ismo Mattila, Marie Frimodt-Møller, Anne Rasmussen, Peter Rossing, Cristina Legido-Quigley, Tarunveer S. Ahluwalia

**Affiliations:** Steno Diabetes Center Copenhagen, Herlev, Denmark; Department of Medicine, Herlev-Gentofte Hospital, Copenhagen, Denmark; Department of Clinical Medicine, University of Copenhagen, Copenhagen, Denmark; The Bioinformatics Center, Department of Biology, University of Copenhagen, Copenhagen, Denmark; Department of Science and Environment, Roskilde University, Roskilde, Denmark

**Keywords:** Metabolomics, diabetic foot complications, diabetic foot ulcer, amputation, Charcot

## Abstract

Diabetic foot complications (DFCs) comprising diabetic foot ulcer (DFU), charcot’s neuroarthropathy and amputations are a collective term used for the ailments of the foot that individuals with diabetes incur. Despite implementation of national and international guidelines, DFCs are still a growing challenge to the individual and society. Novel markers for the treatment and prevention of DFCs are thus needed. The aim of this study was to investigate circulating metabolites associated with the prevalence and incidence of DFCs in persons with type 1 diabetes (T1D). A panel of non-targeted serum metabolites (n=75) were analyzed using mass spectrometry in 637 individuals with T1D with a median follow up time of 10 years. Cross sectional associations between metabolites and DFCs were analysed by linear regression models at baseline, Cox proportional hazards model at follow-up and adjusted for relevant confounders (age, sex, Hb_A1c_, systolic blood pressure, bmi, smoking, statin use, total cholesterol, plasma triglycerides, and renal function). The median (interquartile range) age was 55 (47, 64) years, diabetes duration of 35 (25, 44) years and HbA_1c_ levels 64 (8%) (56, 72(7.3%, 8.7%)) mmol/mol. In the adjusted model, four amino acids (Proline, Threonine, Valine, and Leucine) were associated with a decreased incidence of Charcot’s arthropathy at baseline (*p*<0.05). In addition, circulating ribonic acid levels were associated with an increased risk of DFUs during follow-up (HR 1.38(1.06-1.8); p<0.05) which were validated in an independent cross-sectional T1D cohort (p<0.05). This study identifies novel circulating metabolites, as potential biomarkers for risk stratification of diabetic foot complications.

## Introduction

The complications that afflict the feet of individuals with diabetes are often referred to under the common term diabetic foot complications. Generally, the term covers four ailments that afflict the diabetic foot; diabetic foot ulcers (DFU), infections of the diabetic foot, amputations incurred by individuals with diabetes and Charcot’s arthropathy. The ailments encompassed in the term are closely linked to each other; and with a lifetime risk of 19-34% the DFU is the most common diabetic foot complication incurred by individuals with diabetes(1). The DFU is often the triggering event for infections and amputations of the diabetic foot(1) and has been associated to risk of Charcot’s arthropathy(2). Apart from the connection between the individual ailments of the diabetic foot, the diabetic foot borrows from both the macro and microvascular complications of diabetes with connections to neuropathy, peripheral arterial disease and retinopathy among others(3). To put the challenge in perspective an individual with diabetes is subjected to amputation of the lower limb or part of a lower limb every 30 sec. on a global scale(4). In addition to the burden incurred by the individual, the diabetic foot complication poses a significant economic challenge to all societies in the world. The cost of treating the diabetic foot complication is substantial, with a 5.4 times higher cost of treating an individual with a diabetic foot complication compared to an individual with diabetes without a foot complication(5). On a larger scale 1% of the total health-care budget in England was spend on treating the diabetic foot(6). These challenges have led to several national and international strategies with the intend of reducing the incidence of diabetic foot complications(7). This has, however, not lead to a significant reduction in the incidence or cost of diabetic foot complications in general. In addition, the projected increase in size and lifespan of the global diabetic population, new treatment strategies are needed to incur reductions in or even prevention of prevention of diabetic foot complications(3).

One of the emerging frontiers in prevention and treatment of the diabetic foot complication is metabolomics. Metabolomics has shown promise in identifying pathways and biomarkers associated with risk of developing diabetes(8; 9). Furthermore, specific metabolites have been associated with increased risk of developing complications associated with diabetes(10). Despite the increased focus on the relationship between metabolomics and diabetic complications, studies on this topic are still lacking.

Thus, the aim of the current study was to investigate the association between a non-targeted panel of circulating serum metabolites and three severe forms of diabetic foot complications (DFU, amputations and Charcot’s arthropathy (form here on referred to as diabetic foot complications or DFCs)) among adult persons with type 1 diabetes (T1D).

## Research Design and Methods

637 individuals with T1D and a wide range of albuminuria were recruited from the outpatient clinic at Steno Diabetes Center Copenhagen (SDCC).

This is a longitudinal observational cohort recruited between 2009 and 2011. Details of this cohort have been previously described (11).

Another cross-sectional observational cohort with 161 T1D individuals being followed at SDCC and recruited during 2016-2017, also described in detail previously (12), was used as the validation cohort.

All participants were subdivided into different albuminuria stages, normoalbuminuria (<3.39 mg/mmol corresponding to <30 mg/24 h or mg/g), moderately increased albuminuria (3.39–33.79 mg/mmol corresponding to 30–299 mg/24 h or mg/g), and severely increased albuminuria (≥33.90 mg/mmol corresponding to ≥300 mg/24 h or mg/g). Participants with end stage kidney disease defined as receiving dialysis, kidney transplantation, estimated glomerular filtration rate (eGFR)< 15ml/min/1.73m^2^ at baseline were excluded.

In the current study plasma metabolomics data along with information on (with or without) all foot complications (diabetic foot ulcer (DFU), amputation, and Charcot’s arthropathy) was available for 637 individuals in the primary study whereas 126 individuals with T1D and peripheral neuropathy had information available only on DFUs (validation cohort).

The study was conducted according to the Helsinki Declaration on ethical principles for medical research. The study was approved by the Danish Ethical Committee for the Capital Region of Denmark (Hillerød, Denmark), Danish Patient Safety Authorities and Danish Data Protection Agency. All participants provided informed written consent.

### Baseline Characteristics and Diagnosis Codes

All information on clinical characteristics and diabetic foot complications was extracted from the patient’s electronic health records. Three diabetic foot complications including DFUs, amputations, and Charcot’s arthropathy were investigated in the current study. All information on diabetic foot complications were attained in accordance with the international classification of diseases, tenth revision(13). A DFU was defined as a lesion of the skin on the foot of the person with type 1 diabetes (14). All information on procedures (amputations) were attained in accordance with Danish version of the Nordic Classification of Surgical Procedures(15). Date of diabetic foot complications diagnosis was cross referenced with date of participant blood sampling for metabolomic and biochemical analysis (or baseline). The diagnosis was then registered as “at baseline” if date of diagnosis was made prior to or at baseline and as during follow-up if date of diagnosis was after baseline. eGFR was calculated using the CKD-EPI 2012 equation (16).

Participant baseline characteristics (Table 1) refer to the time point where blood collections were made for metabolomics analyses. Longitudinal data was acquired until 31^st^ Dec 2020. Data regarding potential changes in medication during follow-up were not available.

**Table 1.**
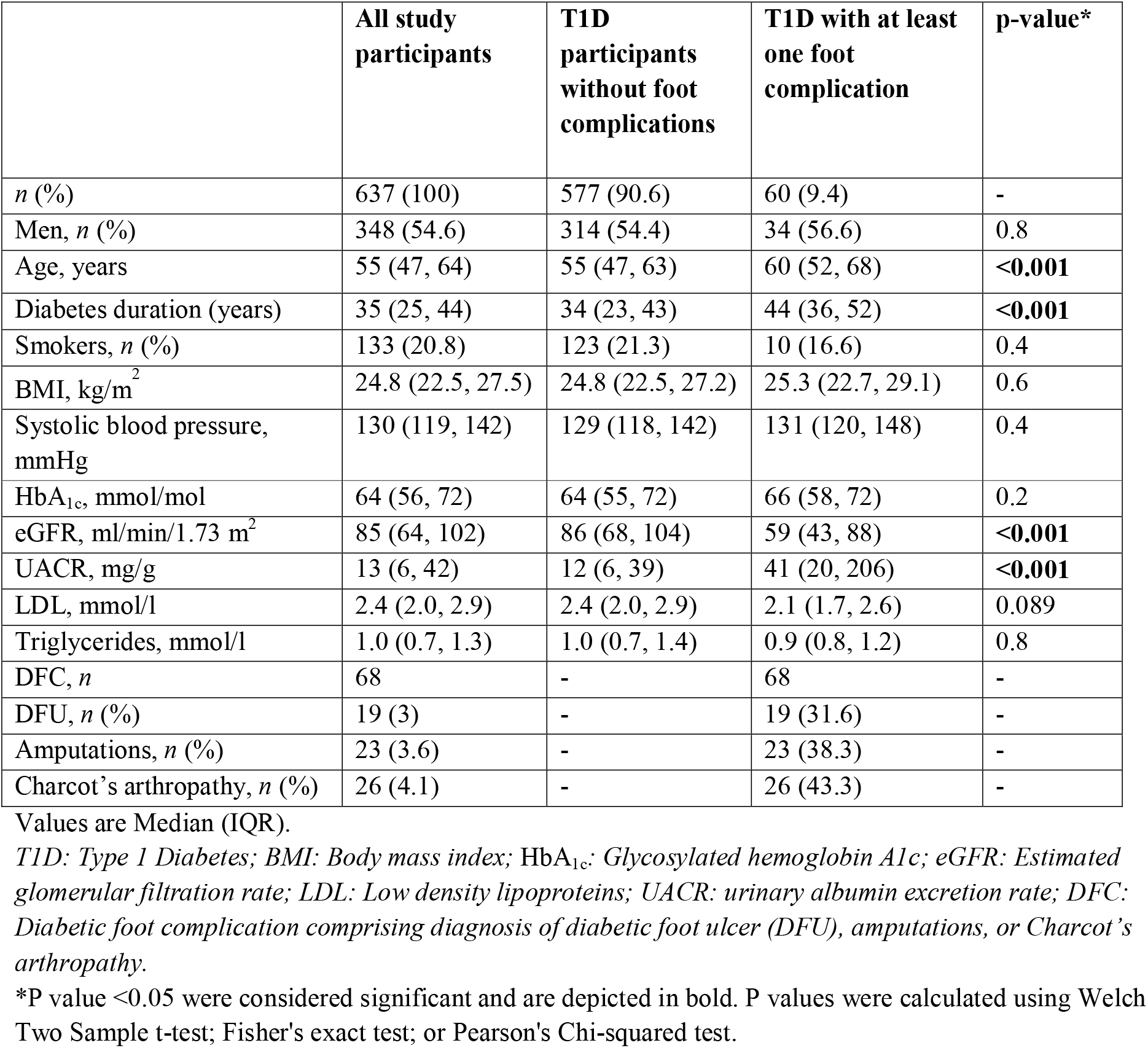
Clinical characteristics of the study participants at baseline.

|  | All study participants | T1D participants without foot complications | T1D with at least one foot complication | p-value* |
| --- | --- | --- | --- | --- |
| *n* (%) | 637 (100) | 577 (90.6) | 60 (9.4) | - |
| Men, *n* (%) | 348 (54.6) | 314 (54.4) | 34 (56.6) | 0.8 |
| Age, years | 55 (47, 64) | 55 (47, 63) | 60 (52, 68) | **<0.001** |
| Diabetes duration (years) | 35 (25, 44) | 34 (23, 43) | 44 (36, 52) | **<0.001** |
| Smokers, *n* (%) | 133 (20.8) | 123 (21.3) | 10 (16.6) | 0.4 |
| BMI, kg/m<sup>2</sup> | 24.8 (22.5, 27.5) | 24.8 (22.5, 27.2) | 25.3 (22.7, 29.1) | 0.6 |
| Systolic blood pressure, mmHg | 130 (119, 142) | 129 (118, 142) | 131 (120, 148) | 0.4 |
| HbA<sub>1c</sub>, mmol/mol | 64 (56, 72) | 64 (55, 72) | 66 (58, 72) | 0.2 |
| eGFR, ml/min/1.73 m<sup>2</sup> | 85 (64, 102) | 86 (68, 104) | 59 (43, 88) | **<0.001** |
| UACR, mg/g | 13 (6, 42) | 12 (6, 39) | 41 (20, 206) | **<0.001** |
| LDL, mmol/l | 2.4 (2.0, 2.9) | 2.4 (2.0, 2.9) | 2.1 (1.7, 2.6) | 0.089 |
| Triglycerides, mmol/l | 1.0 (0.7, 1.3) | 1.0 (0.7, 1.4) | 0.9 (0.8, 1.2) | 0.8 |
| DFC, *n* | 68 | - | 68 | - |
| DFU, *n* (%) | 19 (3) | - | 19 (31.6) | - |
| Amputations, *n* (%) | 23 (3.6) | - | 23 (38.3) | - |
| Charcot's arthropathy, *n* (%) | 26 (4.1) | - | 26 (43.3) | - |

Values are Median (IQR).
T1D: Type 1 Diabetes; BMI: Body mass index; HbA<sub>1c</sub>: Glycosylated hemoglobin A1c; eGFR: Estimated glomerular filtration rate; LDL: Low density lipoproteins; UACR: urinary albumin excretion rate; DFC: Diabetic foot complication comprising diagnosis of diabetic foot ulcer (DFU), amputations, or Charcot's arthropathy.
\*P value <0.05 were considered significant and are depicted in bold. P values were calculated using Welch Two Sample t-test; Fisher's exact test; or Pearson's Chi-squared test.

### Serum Metabolomics Analyses

The method used has previously been described in detail by Tofte et. al(17). In brief, the metabolomic measurements were performed using serum stored at -80° C. Leco Pegasus four-dimensional gas chromatography with time off flight mass spectrometry (4D GC×GC TOFMS) instrument (Leco Corp. MI, USA) was used for metabolomics measurements. Using ChromaTOF raw data were assessed using peak-picking and resulting data were processed (alignment and normalization) with Guinea(18) and postprocessed in R-software. Inclusion of metabolites in the following analysis was based on certainty of the identification and level of technical precision. No restrictions for prior hypothesis or known pathways were implemented. A total of 75 metabolites (ESM Table 1, Supplementary table 4.1.1.3) were measured and included in analyses. These included amino acids, free fatty acids and compounds from the polyol and energy metabolite pathways.

Metabolites with missing or undetectable values less than or equal to 20%, underwent imputation using the k-nearest neighbor algorithm and were log2-transformed as previously described (17; 19).

## Statistical analysis

All statistical analysis and subsequent data visualization were carried out using the R-software. Continuous variables are given as median (interquartile range Q1-Q3 or IQR). Skewed variables, including all metabolites were log transformed before association testing. Categorical variables are given as total number followed by %. Clinical characteristics at baseline between two groups with and without diabetes foot complications were compared using Welch two sample t-test, Fisher’s exact test or Chi-squared test for continuous or categorical variables, respectively. Circulating metabolites and specific diabetic foot complication outcome were evaluated as follows: Cross sectional associations between each metabolite and diabetic foot complication outcome at baseline were assessed using multivariate linear regression models adjusted for relevant clinical variables.

The following models were used: *Crude model:* unadjusted, *M1 model* adjusted for age, gender, body mass index, systolic blood pressure, cholesterol, HbA_1c_, smoking, statin use, and triglycerides, also referred to as adjusted model. *M2 model* was M1 with additional adjustment for renal function markers (eGFR and/or UACR) also referred to as “fully adjusted” model. Multiple testing correction for p values were perfomed using the Benjamini-Hochberg method (p_adjusted_) throughout the analyses.

Cox proportional hazards model was used to assess longitudinal follow-up end points. The hazard ratios (HR) are reported per doubling of specified metabolites level. Significant or top performing metabolites with P_adjusted_ <0.05 in the adjusted (M1) longitudinal cox proportional hazards model were included in survival analysis with the fully adjusted cox proportional hazards (M2) model for end points. The R-package “survminer” was used for constructing a) Kaplan-Meier plots for end points during longitudinal follow-up with stratification on median levels of the top-performing metabolite at baseline, and b) forest plot of the hazard ratios of the top-performing metabolite and clinical covariates. Integration of the results from all the crude models was visualized with the R-package “circlize”. Significance was defined as p<0.05 (before multiple testing) and p_adjusted_<0.05 (after multiple testing) unless other is noted. Significance for validation cohort was defined as p<0.05.

## Results

### Clinical characteristics of study population

The primary study included 637 participants (54.6% males) with T1D, a median (IQR) age of 55 (47-64) years, diabetes duration of 35 (25-44) years (Table 1). In the following the participants are divided in two groups; first group without any diabetic foot complication at baseline (n=577) and the second group with at least one foot complication at baseline (*n*=60).

Age, diabetes duration, and urinary albumin creatinine ratio (UACR) were significantly higher in T1D with foot complications compared to the T1D without foot complications group. The estimated glomerular filtration rate (eGFR) was significantly lower in the two groups with foot complications compared to the group without foot complications (Table 1).

All T1D individuals from baseline that did not incur a diabetic foot complication during a median follow up time of 10 years formed the no-foot complications group (*n*=488) whereas those who developed at least one foot complication during a median follow up (n=89) formed the foot complications group, for longitudinal analysis (Extra supplementary material, ESM Table 2). A total of 68 diagnoses of diabetic foot complications were registered at baseline (Table 1), while 149 new diagnoses were made during follow-up (ESM Table 2).

### Cross-sectional analysis

All 75 circulating metabolites were included in a linear regression analysis at baseline (Supplementary file Table 4.1.1.3).

In the crude model, 20 metabolites were associated with Charcot’s arthropathy at baseline before multiple testing correction (p<0.05) but only 9 remained significant after correction (p_adjusted_<0.05; Figure 1, Supplementary Figure 4.1.1.1 and Table 4.1.1.2). In the M1 four amino acids (Proline, Threonine, Valine, and Leucine) were negatively while six other metabolites were positively associated with Charcot’s arthropathy at baseline (p_adjusted_<0.05; Figure 1, Supplementary Figure 4.1.2.1, and Table 4.1.2.2). In the M2 model, only proline remained associated (p_adjusted_<0.05) with decreased incidence of Charcot’s arthropathy (Figure 1, Supplementary figure 4.1.4.1 and Table 4.1.4.2).

**Figure 1.**
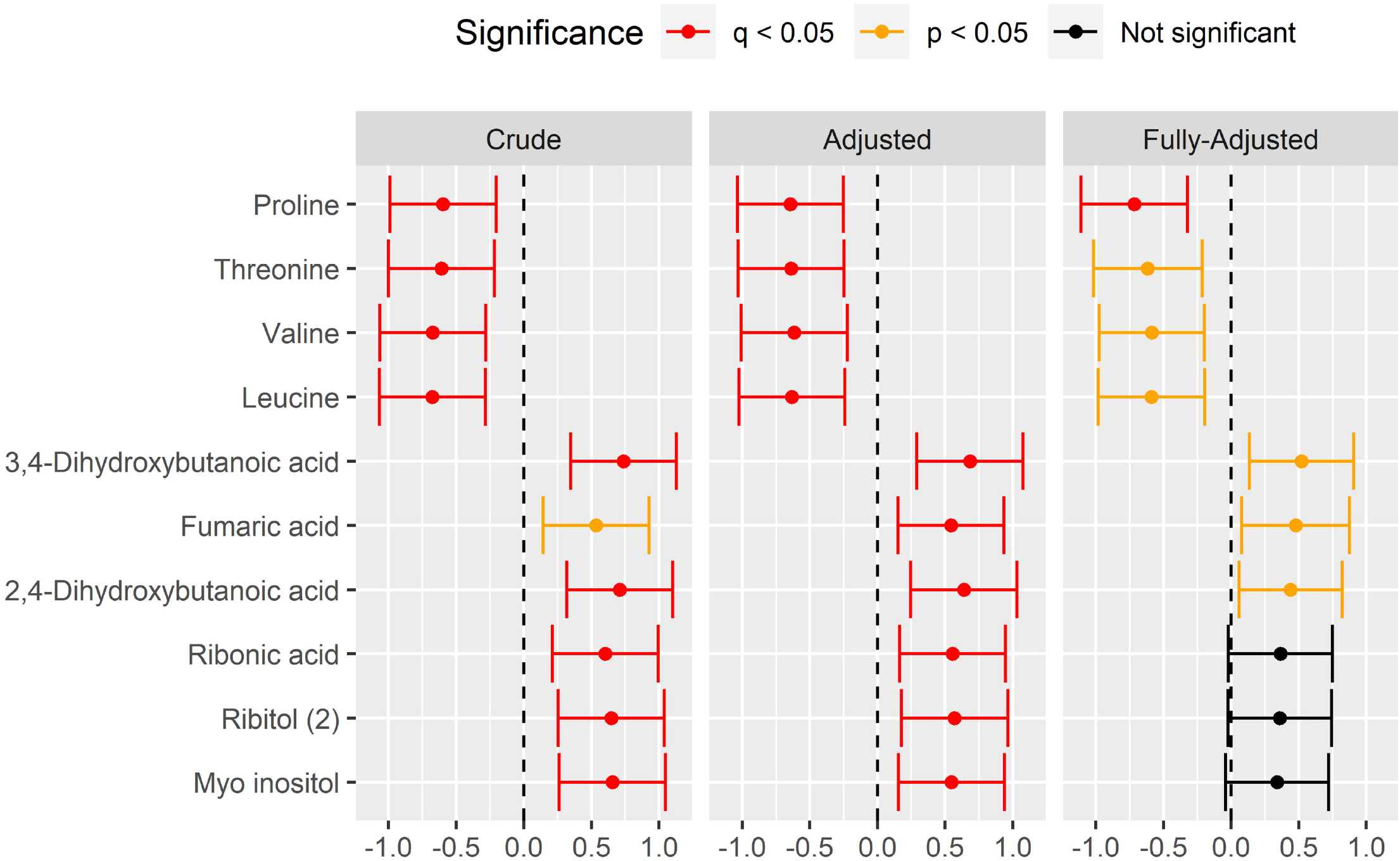
Cross-sectional analysis -*Charcot’s Arthropathy* Cross-sectional analysis of association between baseline metabolites level and Charcot’s arthropathy at baseline. Divided in crude (with correction for multiple testing), adjusted (further adjusted for age, gender, body mass index, systolic blood pressure, cholesterol, glycosylated hemoglobin A1c, smoking, statin and triglycerides) and fully-adjusted (further adjusted for estimated glomerular filtration rate (eGFR) and urinary-albumin creatinine ratio), with 95% confidence intervals. Significance was defined as *p<0*.*05*. In this figure an significant *p*-value not adjusted for multiple testing is marked in yellow (p). A *p*-value that is significant after adjustment for multiple testing is marked in red (q). Finally, a *p*-value that is not-significant before multiple testing is marked in black.

**Figure 2.**
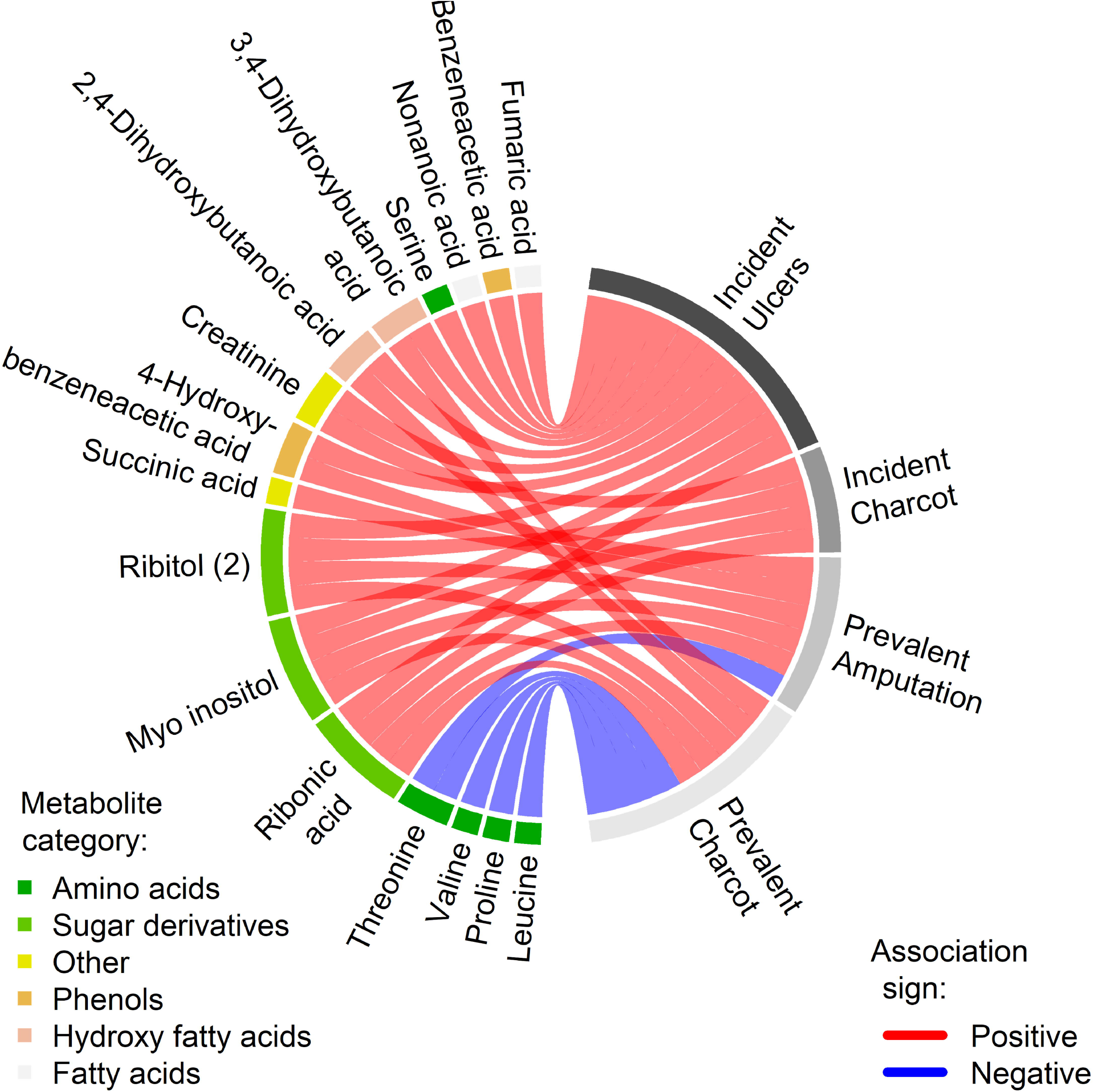
Survival analysis – *diabetic foot ulcer-free days* Kaplan-Meier analysis of ulcer-free days stratified on median of plasma levels of Ribonic acid at baseline.

Seventeen metabolites showed an association with amputations at baseline before multiple testing (p<0.05, Supplementary Table 5.1.1.2) in the crude model, while six remained significant after correction (p_adjusted_<0.05). Of the six metabolites, five were positively associated (Ribonic acid,

Myo Inositol, 4-Hydroxybenzeneacetic acid, Ribitol, and Succinic acid) while amino acid Threonine was negatively correlated with amputations (Supplementary Figure 5.1.1.1). In the M1 model three metabolites showed a significant association with amputations at baseline before but none after the multiple testing correction (Supplementary Figure 5.1.2.1 and Table 5.1.2.2). None of the metabolites were associated with the incidence of DFUs at baseline after correcting for multiple testing in the crude or adjusted models (p_adjusted_>0.05; Supplementary Tables 6.1.1.3, 6.1.2.3, and 6.1.3.3).

### Longitudinal analysis

All plasma metabolites used for cross-sectional analysis were examined for longitudinal analyses. Albeit 17 circulating metabolites associated with the risk of incurring Charcot’s arthropathy during follow-up in the crude model, only four metabolites (Ribonic acid, Ribitol, Creatinine, Myo Inositol) remained significant (Supplementary Figure 4.2.1.1 and Table 4.2.1.2) after multiple testing correction. In the M1 model out of 16 metabolites associated to risk of incurring Charcot’s arthropathy during follow up (p<0.05, Supplementary Table 4.2.2.2), none retained after multiple testing correction. Similarly, no metabolites remained significant in M3.

None of the tested metabolites were associated with risk of incurring an amputation during follow-up in crude or adjusted models (p_adjusted_>0.05; supplementary Tables 5.2.1.2, 5.2.2.2, and 5.2.3.2). In the crude model 11 metabolites were significantly associated (p_adjusted_<0.05) to the risk of incurring a DFU during follow-up (Supplementary Figure 6.2.1.1 and Table 6.2.1.2). In the M1 model two metabolites (Ribonic acid and Tyrosine) remained significantly associated with risk of incurring DFU during follow-up (Supplementary Figure 6.2.2.1 and Table 6.2.2.2). Ribonic acid retained a significant association with risk of incurring a DFU during follow-up with a hazard ratio of 1.39 (1.08-1.8 (*p*=0.012) (Supplementary Section 6.2.3.3 and Figure 6.2.3.3.1)) after adjustment for renal function.

**Figure 3.**
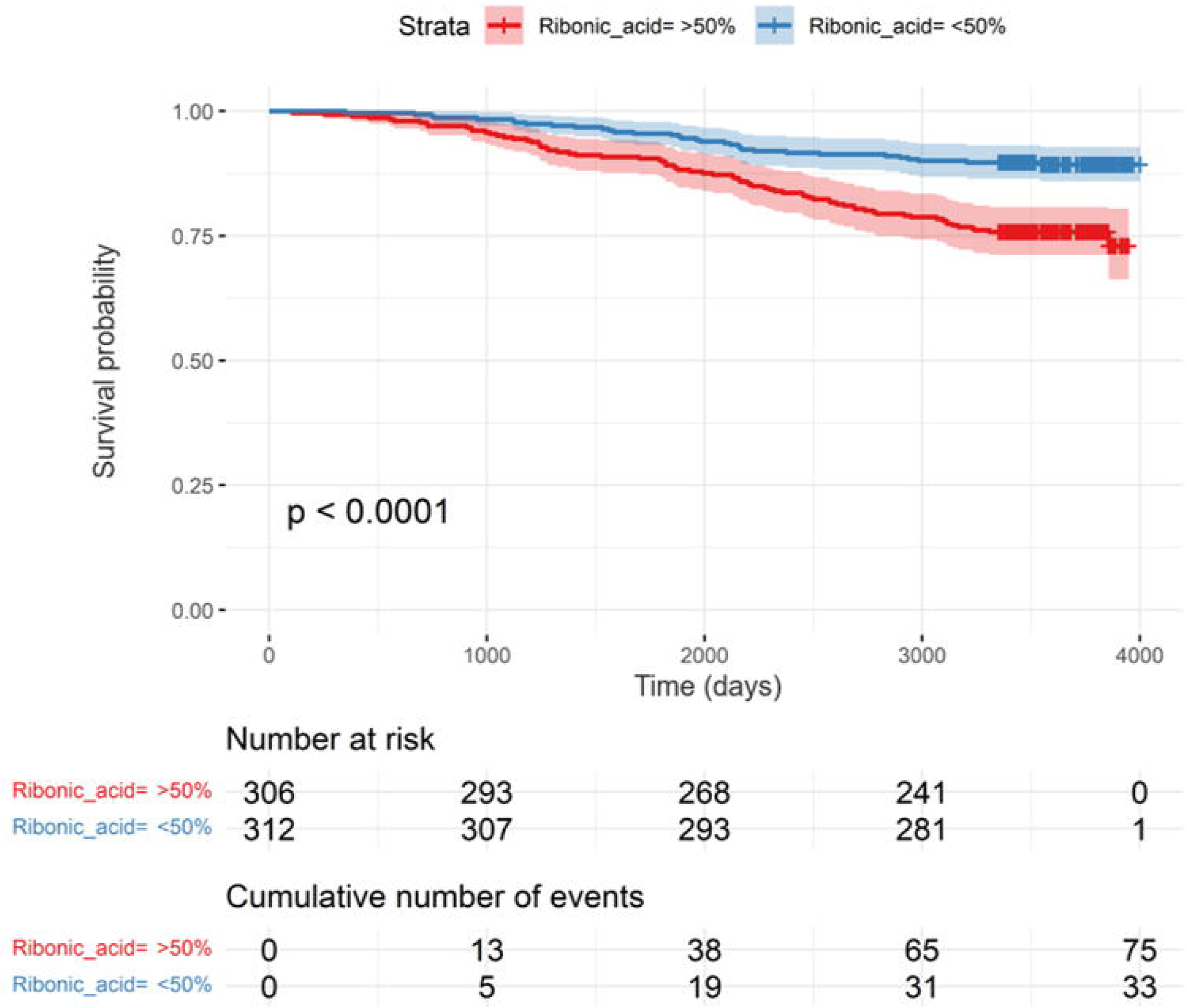
Overview of serum metabolite associations with diabetic foot complications (incident and prevalent; crude model). Chord diagram of crude associations between the metabolome (left) and foot complications (right) at baseline (prevalent cases) and during the follow-up (incident cases). A curve between a metabolite and a complication depicts a significant crude association (red: positive; blue: negative) after correction for multiple testing. The category of each metabolite is indicated by the color aside the metabolite name. Amino acids Leucine, Proline, Threonine and Valine are negatively associated with prevalent Charcot’s arthropathy (blue curves), whereas most of the other associations are positive (red curves).

A survival analysis of DFU-free days stratified on median plasma levels of Ribonic acid in the study population showed a significantly higher number of DFU-free days in the group with the lowest levels of Ribonic acid in serum at baseline (*p*<0.0001) (Figure 2).

Overall associations of significant metabolites and diabetic foot complication outcomes from the crude model have been depicted in Figure 3 chord diagram.

### Validation

We replicated Ribonic acid association with DFU in the cross-sectional validation study with 126 T1D individuals: 27 with DFU and 99 without any DFU (ESM Table 3).

Higher circulating Ribonic acid levels were significantly associated with DFU risk in the crude (Beta: 0.57; p = 0.013), M1 (Beta: 0.57; p=0.019) and M2 (Beta: 0.53; p=0.024) models, respectively (ESM Table 4; ESM figure 1).

## Discussion

This is one of the first prospective studies to examine a possible correlation between circulating plasma metabolites and diabetic foot complications. Several metabolites with association to incidence and risk of incurring diabetic foot complications are identified in the current study. Perhaps the most interesting being Ribonic acid also known as Ribonate, a derivate of the sugar Ribose and part of the pentose pathway. A significant association between risk of incurring a DFU and plasma levels of Ribonic acid. Prior studies have associated Ribonic acid with the incidence and future risk of retinopathy and nephropathy in the same cohort examined here(17; 19). This may hint at general correlation between complications associated with diabetes and plasma levels of Ribonic acid in this one cohort. While this may still be true, a study by Chen et al. identified an association between plasma levels of Ribonic acid and retinopathy when comparing a population with diabetes and retinopathy to a population with diabetes without retinopathy(20). Similarly, Hu et al. found that all-cause mortality was linked to plasma levels of Ribonic acid in a population with chronic kidney disease(21). A possible role for Ribonic acid and/or the pentose pathway, as a marker of diabetes, is further supported by animal models that have shown that Ribonic acid levels in urine and plasma are elevated when diabetes is induced(22; 23). Although the proposed role of the pentose pathway and Ribonic acid is still unclear, it is a candidate for future research both when looking for potential biomarker and metabolomic pathways to target, in treatment and prevention of diabetic foot complications.

In addition to the connection between Ribonic acid and development of DFUs, an inverse association between plasma levels of four amino acids (Threonine, Leucine, Valine, and Proline) and incidence Charcot’s arthropathy at baseline was found. Like Ribonic acid the role of circulating amino acids in diabetes complications is unclear. There have been studies showing a vital role for amino acids in human metabolic pathways including gluconeogenesis(24-26) and the secretion of insulin and glucagon(27). In regard to diabetic complications and circulating amino acids the published data is sparse for foot related complications albeit associations to microvascular complications (nephropathy and autonomic neuropathy) have been shown (10; 28-30). A recent study by Hung et al. found an association between DFU healing and plasma levels of four amino acids. These amino acids included Threonine and Leucine, which were also found in our study(31). An association between amino acids and healing of pressure, trauma and burn ulcers have been proposed in other studies(32; 33). Prospective randomized studies have even hinted at beneficial effects on pressure-ulcer healing of nutritional supplements including specific amino acids(34). However, the beneficial effects are still uncertain due to the limited available data(35). Similar results have been seen when adding nutritional supplements to treatment of individuals with diabetes; smaller retrospective and prospective studies have shown promise(36; 37), but the combined data is insufficient to conclude if there is a beneficial effect(38). While the role of diet in diabetic foot complications is still unclear, the role of diet in diabetes in general is not up for debate(39). We need further studies on the association between diabetic foot complications and amino acids to identify the most potent biomarkers and targets for treatment and prevention of diabetic foot complications.

Study limitations included missing information on medication. Consequently, information on dosage and changes in dosage during the study is unavailable. In addition, information on some of the known clinical risk factors (toe blood pressure and foot deformities) in development of diabetic foot complications were unavailable at the time of data analysis. Finally, information on diet, which may affect the composition of the individual participants metabolome, was unavailable. On the other hand, the strengths of this study are, the large size of the cohort with T1D, long follow-up, high throughput metabolomic measurements using mass spectrometry and high qualitative bioinformatic data analyses methods.

A recent editorial from our institute highlights the importance of including longitudinal follow-up, as presented in this study, in metabolomic and similar studies on diabetes and its complications(40). This study adds to the growing evidence base for specific metabolites associated to complications of diabetes, specifically DFU and Charcot’s arthropathy. The identified circulating metabolites have already been described in the literature, suggesting associations with diabetes and microvascular complications. Plasma metabolomics in diabetic foot complications is an intriguing prospect, as risk profiling promises the opportunity to prevent rather than treat these debilitating ailments. In addition, metabolomics can help uncover molecular pathways of foot complication development, that are also potential targets for developing novel therapeutics.

## Acknowledgments

We thank the participants of this study. Jonas A. Andersen, and Tarunveer S. Ahluwalia are the guarantors of this study.

## Funding

TSA was supported by funding from the Novo Nordisk Foundation NNF18OC0052457 and Steno Diabetes Center Copenhagen.

## Author Contributions

Conceptualization: JAA, PR, TSA

Methodology: TS, KT, IM, CLQ, TSA

Investigation: JAA, TS, MJR-L, TSA

Visualization: TS

Funding acquisition: ST, PR, CLQ, TSA

Project administration: ST, AR, MFM, PR, TSA

Supervision: TSA

Writing – original draft: JAA, TS, TSA

Writing – review & editing: JAA, TS, ST, AR, MFM, PR, CLQ, TSA

All authors read and approved the final version of the manuscript.

## Competing interests

None of the authors have any competing interest connected to this study. *Outside of this study:* PR has received consultancy and/or speaking fees (to SDCC) from AbbVie, Astellas, AstraZeneca, Bayer, Boehringer Ingelheim, Gilead, Eli Lilly, MSD, Novo Nordisk and Sanofi Aventis, and research grants from Novo Nordisk and Astra Zeneca.

MFM has received speaking fees from Boehringer Ingelheim, Novartis, Baxter and Sanofi

## Data availability

The data sets used in this study are available from the corresponding author after acquiring required permissions from the relevant regulatory authorities.

## Notes

### Funding Statement

This study PI was funded by the Novo Nordisk Foundation.

### Author Declarations

The study was conducted according to the Helsinki Declaration on ethical principles for medical research. The study was approved by the Danish Ethical Committee, Danish Patient Safety Authorities and Danish Data Protection Agency.

### Summary of Updates

The revised manuscript includes validation of original results in an additional Type 1 Diabetes cohort comprising information on Diabetic Foot ulcers and top metabolite measures. It also involves one additional co author (Maria Jose Romero Lado) who was involved in data analysis for validation part.

## References

1. Armstrong DG, Swerdlow MA, Armstrong AA, Conte MS, Padula WV, Bus SA. Five year mortality and direct costs of care for people with diabetic foot complications are comparable to cancer. J Foot Ankle Res 2020;13:16

2. Fauzi AA, Chung TY, Latif LA. Risk factors of diabetic foot Charcot arthropathy: a case-control study at a Malaysian tertiary care centre. Singapore Med J 2016;57:198–203

3. Edmonds M, Manu C, Vas P. The current burden of diabetic foot disease. J Clin Orthop Trauma 2021;17:88–93

4. IDF Diabetes Atlas [article online], 2021. Available from https://diabetesatlas.org/. Accessed 22/11-2021 2021

5. Williams R, Karuranga S, Malanda B, Saeedi P, Basit A, Besancon S, Bommer C, Esteghamati A, Ogurtsova K, Zhang P, Colagiuri S. Global and regional estimates and projections of diabetes-related health expenditure: Results from the International Diabetes Federation Diabetes Atlas, 9th edition. Diabetes Res Clin Pract 2020;162:108072

6. Kerr M, Barron E, Chadwick P, Evans T, Kong WM, Rayman G, Sutton-Smith M, Todd G, Young B, Jeffcoate WJ. The cost of diabetic foot ulcers and amputations to the National Health Service in England. Diabet Med 2019;36:995–1002

7. Parker CN, Van Netten JJ, Parker TJ, Jia L, Corcoran H, Garrett M, Kwok CF, Nather A, Que MT, Srisawasdi G, Wraight P, Lazzarini PA. Differences between national and international guidelines for the management of diabetic foot disease. Diabetes Metab Res Rev 2019;35:e3101

8. Long J, Yang Z, Wang L, Han Y, Peng C, Yan C, Yan D. Metabolite biomarkers of type 2 diabetes mellitus and pre-diabetes: a systematic review and meta-analysis. BMC Endocr Disord 2020;20:174

9. Arneth B, Arneth R, Shams M. Metabolomics of Type 1 and Type 2 Diabetes. Int J Mol Sci 2019;20

10. Jin Q, Ma RCW. Metabolomics in Diabetes and Diabetic Complications: Insights from Epidemiological Studies. Cells 2021;10

11. Tofte N, Suvitaival T, Ahonen L, Winther SA, Theilade S, Frimodt-Moller M, Ahluwalia TS, Rossing P. Lipidomic analysis reveals sphingomyelin and phosphatidylcholine species associated with renal impairment and all-cause mortality in type 1 diabetes. Sci Rep 2019;9:16398

12. Clos-Garcia M, Ahluwalia TS, Winther SA, Henriksen P, Ali M, Fan Y, Stankevic E, Lyu L, Vogt JK, Hansen T, Legido-Quigley C, Rossing P, Pedersen O. Multiomics signatures of type 1 diabetes with and without albuminuria. Front Endocrinol (Lausanne) 2022;13:1015557

13. ICD-10 Version:2010 [article online], 2010. Available from https://icd.who.int/browse10/2010/en. Accessed 09/02-2022 2022

14. DNBo. H. Diabetic Foot Ulcers -a health technology assessment. Copenhagen: Danish National Board of Health, 2011 2011;

15. Sundhedsvæsenets Klassifikations System (SKS) [article online], 2021. Available from https://sundhedsdatastyrelsen.dk/da/rammer-og-retningslinjer/om-klassifikationer/sks-klassifikationer. Accessed 09/02-2022 2022

16. Inker LA, Schmid CH, Tighiouart H, Eckfeldt JH, Feldman HI, Greene T, Kusek JW, Manzi J, Van Lente F, Zhang YL, Coresh J, Levey AS, Investigators C-E. Estimating glomerular filtration rate from serum creatinine and cystatin C. N Engl J Med 2012;367:20–29

17. Tofte N, Suvitaival T, Trost K, Mattila IM, Theilade S, Winther SA, Ahluwalia TS, Frimodt-Møller M, Legido-Quigley C, Rossing P. Metabolomic Assessment Reveals Alteration in Polyols and Branched Chain Amino Acids Associated With Present and Future Renal Impairment in a Discovery Cohort of 637 Persons With Type 1 Diabetes. Front Endocrinol (Lausanne) 2019;10:818

18. Castillo S, Mattila I, Miettinen J, Orešič M, Hyötyläinen T. Data analysis tool for comprehensive two-dimensional gas chromatography/time-of-flight mass spectrometry. Anal Chem 2011;83:3058–3067

19. Curovic VR, Suvitaival T, Mattila I, Ahonen L, Trost K, Theilade S, Hansen TW, Legido-Quigley C, Rossing P. Circulating Metabolites and Lipids Are Associated to Diabetic Retinopathy in Individuals With Type 1 Diabetes. Diabetes 2020;69:2217–2226

20. Chen L, Cheng CY, Choi H, Ikram MK, Sabanayagam C, Tan GS, Tian D, Zhang L, Venkatesan G, Tai ES, Wang JJ, Mitchell P, Cheung CM, Beuerman RW, Zhou L, Chan EC, Wong TY. Plasma Metabonomic Profiling of Diabetic Retinopathy. Diabetes 2016;65:1099–1108

21. Hu JR, Coresh J, Inker LA, Levey AS, Zheng Z, Rebholz CM, Tin A, Appel LJ, Chen J, Sarnak MJ, Grams ME. Serum metabolites are associated with all-cause mortality in chronic kidney disease. Kidney Int 2018;94:381–389

22. Jing L, Chengji W. GC/MS-based metabolomics strategy to analyze the effect of exercise intervention in diabetic rats. Endocr Connect 2019;8:654–660

23. Steer KA, Sochor M, McLean P. Renal hypertrophy in experimental diabetes. Changes in pentose phosphate pathway activity. Diabetes 1985;34:485–490

24. Schutz Y. Protein turnover, ureagenesis and gluconeogenesis. Int J Vitam Nutr Res 2011;81:101–107

25. Basu R, Chandramouli V, Dicke B, Landau B, Rizza R. Obesity and type 2 diabetes impair insulin-induced suppression of glycogenolysis as well as gluconeogenesis. Diabetes 2005;54:1942–1948

26. Chung ST, Hsia DS, Chacko SK, Rodriguez LM, Haymond MW. Increased gluconeogenesis in youth with newly diagnosed type 2 diabetes. Diabetologia 2015;58:596–603

27. Gannon MC, Nuttall FQ. Amino acid ingestion and glucose metabolism--a review. IUBMB Life 2010;62:660–668

28. Mathew AV, Jaiswal M, Ang L, Michailidis G, Pennathur S, Pop-Busui R. Impaired Amino Acid and TCA Metabolism and Cardiovascular Autonomic Neuropathy Progression in Type 1 Diabetes. Diabetes 2019;68:2035–2044

29. Hansen CS, Suvitaival T, Theilade S, Mattila I, Lajer M, Trost K, Ahonen L, Hansen TW, Legido-Quigley C, Rossing P, Ahluwalia TS. Cardiovascular Autonomic Neuropathy in Type 1 Diabetes Is Associated With Disturbances in TCA, Lipid, and Glucose Metabolism. Front Endocrinol (Lausanne) 2022;13:831793

30. Tofte N, Vogelzangs N, Mook-Kanamori D, Brahimaj A, Nano J, Ahmadizar F, van Dijk KW, Frimodt-Moller M, Arts I, Beulens JWJ, Rutters F, van der Heijden AA, Kavousi M, Stehouwer CDA, Nijpels G, van Greevenbroek MMJ, van der Kallen CJH, Rossing P, Ahluwalia TS, Hart LM. Plasma Metabolomics Identifies Markers of Impaired Renal Function: A Meta-analysis of 3089 Persons with Type 2 Diabetes. J Clin Endocrinol Metab 2020;105

31. Hung SY, Tsai JS, Yeh JT, Chen KH, Lin CN, Yang HM, Lin CW, Chen HY, Huang CH, Huang YY. Amino acids and wound healing in people with limb-threatening diabetic foot ulcers. J Diabetes Complications 2019;33:107403

32. Wong A, Chew A, Wang CM, Ong L, Zhang SH, Young S. The use of a specialised amino acid mixture for pressure ulcers: a placebo-controlled trial. J Wound Care 2014;23:259–260, 262-254, 266-259

33. Stechmiller JK. Understanding the role of nutrition and wound healing. Nutr Clin Pract 2010;25:61–68 34.

34. Cereda E, Neyens JCL, Caccialanza R, Rondanelli M, Schols J. Efficacy of a Disease-Specific Nutritional Support for Pressure Ulcer Healing: A Systematic Review and Meta-Analysis. J Nutr Health Aging 2017;21:655–661

35. Langer G, Fink A. Nutritional interventions for preventing and treating pressure ulcers. Cochrane Database Syst Rev 2014:Cd003216

36. Armstrong DG, Hanft JR, Driver VR, Smith AP, Lazaro-Martinez JL, Reyzelman AM, Furst GJ, Vayser DJ, Cervantes HL, Snyder RJ, Moore MF, May PE, Nelson JL, Baggs GE, Voss AC. Effect of oral nutritional supplementation on wound healing in diabetic foot ulcers: a prospective randomized controlled trial. Diabet Med 2014;31:1069–1077

37. Sipahi S, Gungor O, Gunduz M, Cilci M, Demirci MC, Tamer A. The effect of oral supplementation with a combination of beta-hydroxy-beta-methylbutyrate, arginine and glutamine on wound healing: a retrospective analysis of diabetic haemodialysis patients. BMC Nephrol 2013;14:8

38. Moore ZE, Corcoran MA, Patton D. Nutritional interventions for treating foot ulcers in people with diabetes. Cochrane Database Syst Rev 2020;7:CD011378

39. Diabetes Prevention [article online], 2022. Available from https://www.idf.org/aboutdiabetes/prevention.html. Accessed 09/02-2022 2022

40. Ahluwalia TS, Kilpelainen TO, Singh S, Rossing P. Editorial: Novel Biomarkers for Type 2 Diabetes. Front Endocrinol (Lausanne) 2019;10:649

