## Supplementary Data File for "Metabolomic Risk Predictors of Diabetic Foot Complications: a longitudinal observational study in Type 1 Diabetes"

### PROFIL Foot Complications Metabolomics

Tommi Suvitaival,, Steno Diabetes Center Copenhagen

January 20, 2023

### 1 Settings

```
## [1] TRUE
```

#### 2 Dates

### 3 Event-Times

#### 3.1 Counts and Ranges

```
table( data$"Charcot.at.DATE" )
```

```
##
## NEJ  JA
## 611  26
```

```
table( data$"Charcot.from.DATE" )
```

```
##
## NEJ  JA
## 598  13
```

```
table( data$"Ulcer.diagnosis.at.DATE" )
```

```
##
## NEJ  JA
## 618  19
```

```
table( data$"Ulcer.diagnosis.from.DATE" )
```

```
##
## NEJ  JA
## 510 108
```

```
table( data$"Amputation.at.DATE" )
```

```
##
## NEJ  JA
## 614  23
```

```
table( data$"Amputation.from.DATE" )
```

```
##
## NEJ  JA
## 586  28
```

```
range( data$"Charcot_dt", na.rm = TRUE )
```

```
## [1] "1995-11-07" "2018-06-08"
```

```
tapply(
  X = as.numeric( data$"Charcot.tdiff" ),
  INDEX = data$"Charcot.from.DATE",
  FUN = summary
)
```

```
## $NEJ
##    Min. 1st Qu.  Median    Mean 3rd Qu.    Max.
##    2537    2685    2837    2841    2982    3185
##
## $JA
##    Min. 1st Qu.  Median    Mean 3rd Qu.    Max.
##     509    1076    1995    1805    2563    2924
```

```
range( data$"Sår_diagnose_dt", na.rm = TRUE )
```

```
## [1] "2001-12-11" "2020-09-01"
```

```
tapply(
  X = as.numeric( data$"Ulcer.diagnosis.tdiff" ),
  INDEX = data$"Ulcer.diagnosis.from.DATE",
  FUN = summary
)
```

```
## $NEJ
##    Min. 1st Qu.  Median    Mean 3rd Qu.    Max.
##    3353    3492    3653    3658    3815    4001
##
## $JA
##    Min. 1st Qu.  Median    Mean 3rd Qu.    Max.
##     106    1230    1948    1904    2539    3857
```

```
range( data$"Amputation_dt", na.rm = TRUE )
```

```
## [1] "1998-01-01" "2020-02-04"
```

```
tapply(
  X = as.numeric( data$"Amputation.tdiff" ),
  INDEX = data$"Amputation.from.DATE",
  FUN = summary
)
```

```
## $NEJ
##    Min. 1st Qu.  Median    Mean 3rd Qu.    Max.
##    3143    3295    3442    3449    3591    3791
##
## $JA
##    Min. 1st Qu.  Median    Mean 3rd Qu.    Max.
##   259.0   956.5  2138.0  2074.1  2941.2  3468.0
```

#### 3.2 Histograms

##### 3.2.1 Time From Baseline

```
## Don't know how to automatically pick scale for object of type difftime. Defaulting to continuous.

## `stat_bin()` using `bins = 30`. Pick better value with `binwidth`.

## Warning: Removed 17 rows containing non-finite values (stat_bin).
```

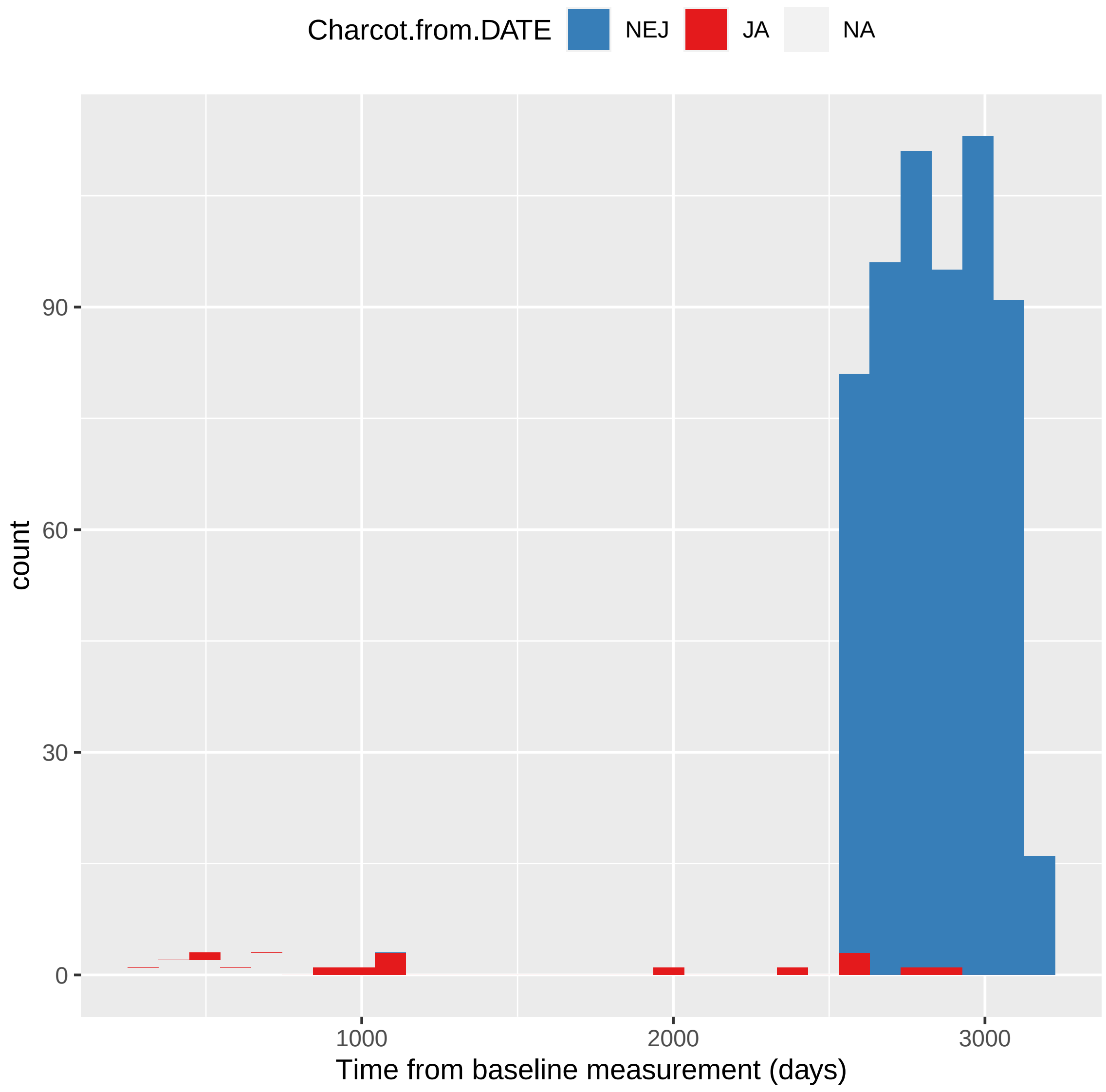

```
## Don't know how to automatically pick scale for object of type difftime. Defaulting to continuous.
## `stat_bin()` using `bins = 30`. Pick better value with `binwidth`.

## Warning: Removed 23 rows containing non-finite values (stat_bin).
```

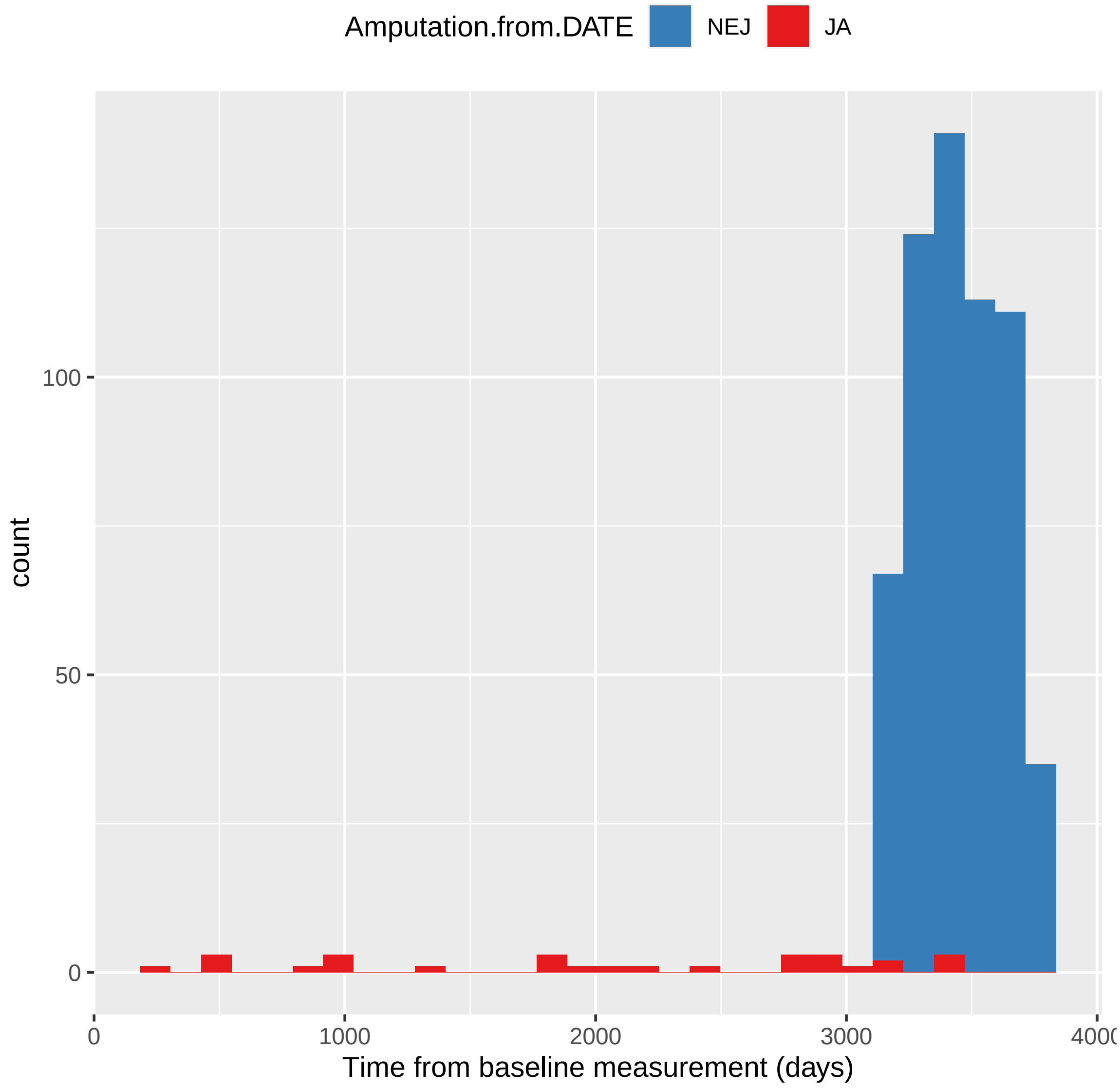

```
## Don't know how to automatically pick scale for object of type difftime. Defaulting to continuous.
## `stat_bin()` using `bins = 30`. Pick better value with `binwidth`.

## Warning: Removed 19 rows containing non-finite values (stat_bin).
```

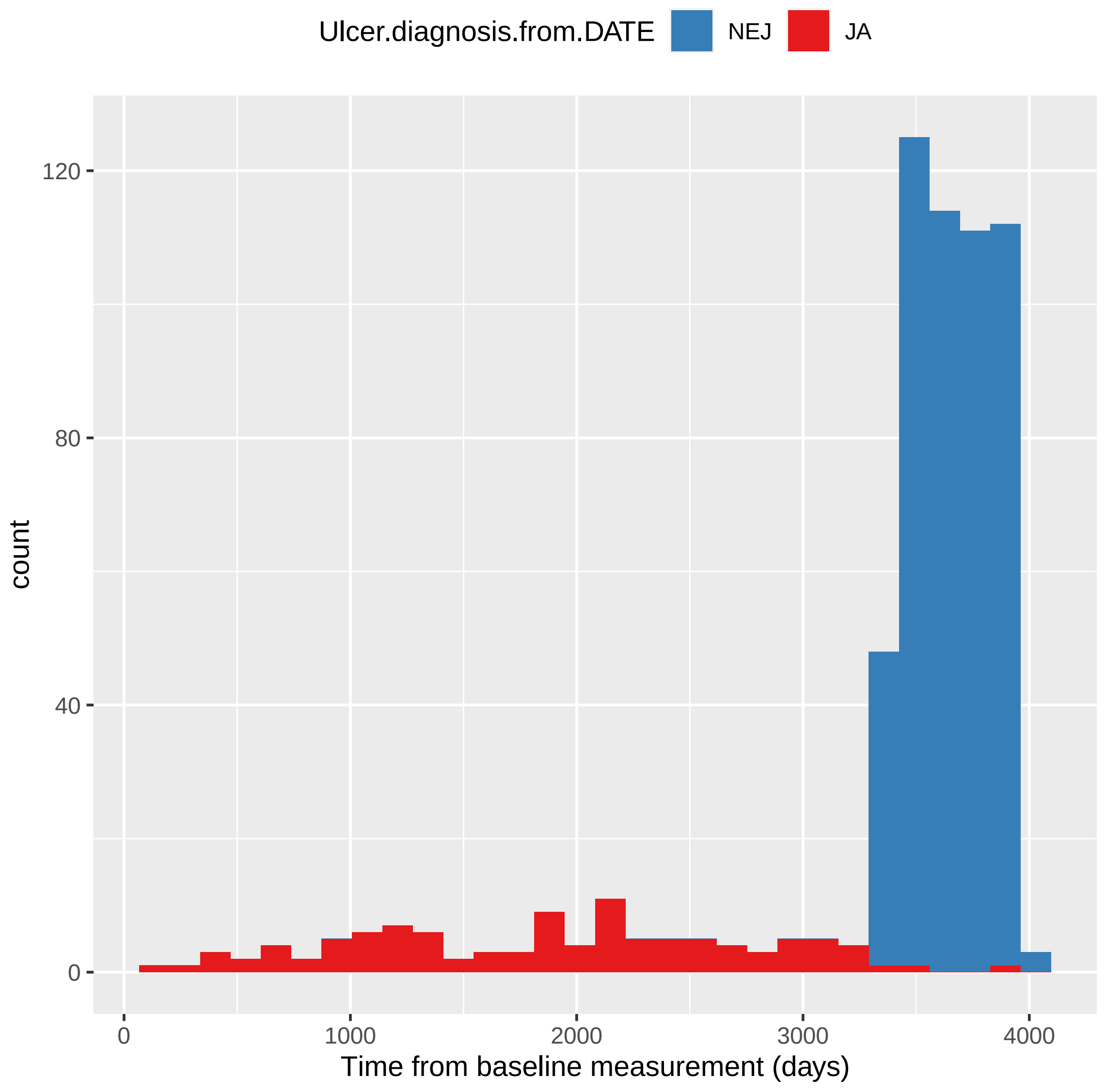
Ulcer.diagnosis.from.DATE
NEJ
JA
120
80
40
0
count
0
1000
2000
3000
4000
Time from baseline measurement (days)

##### 3.2.2 Events v. Baseline

```
## `stat_bin()` using `bins = 30`. Pick better value with `binwidth`.
```

```
## Warning: Removed 598 rows containing non-finite values (stat_bin).
```

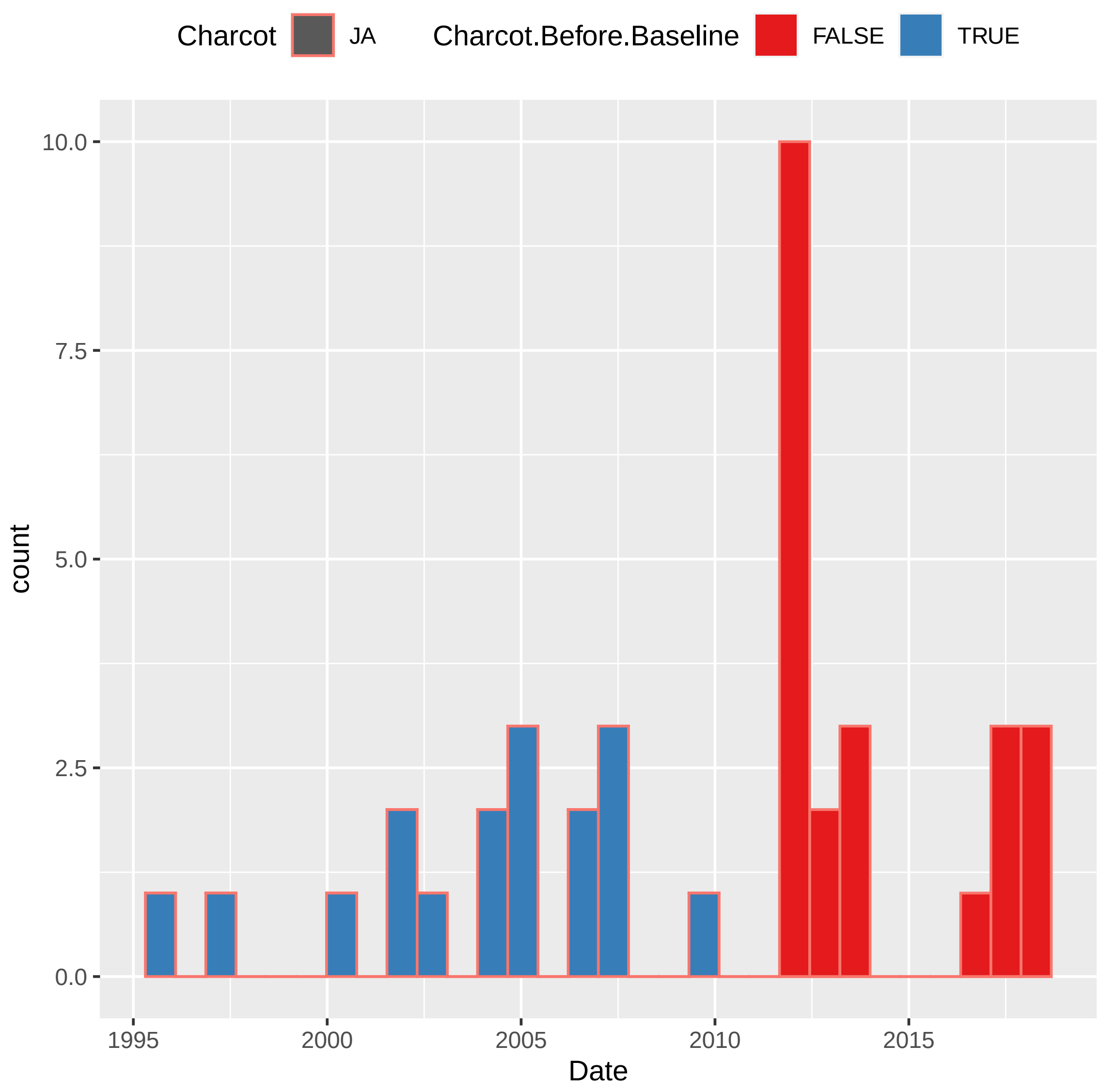

```
## `stat_bin()` using `bins = 30`. Pick better value with `binwidth`.
```

```
## Warning: Removed 586 rows containing non-finite values (stat_bin).
```

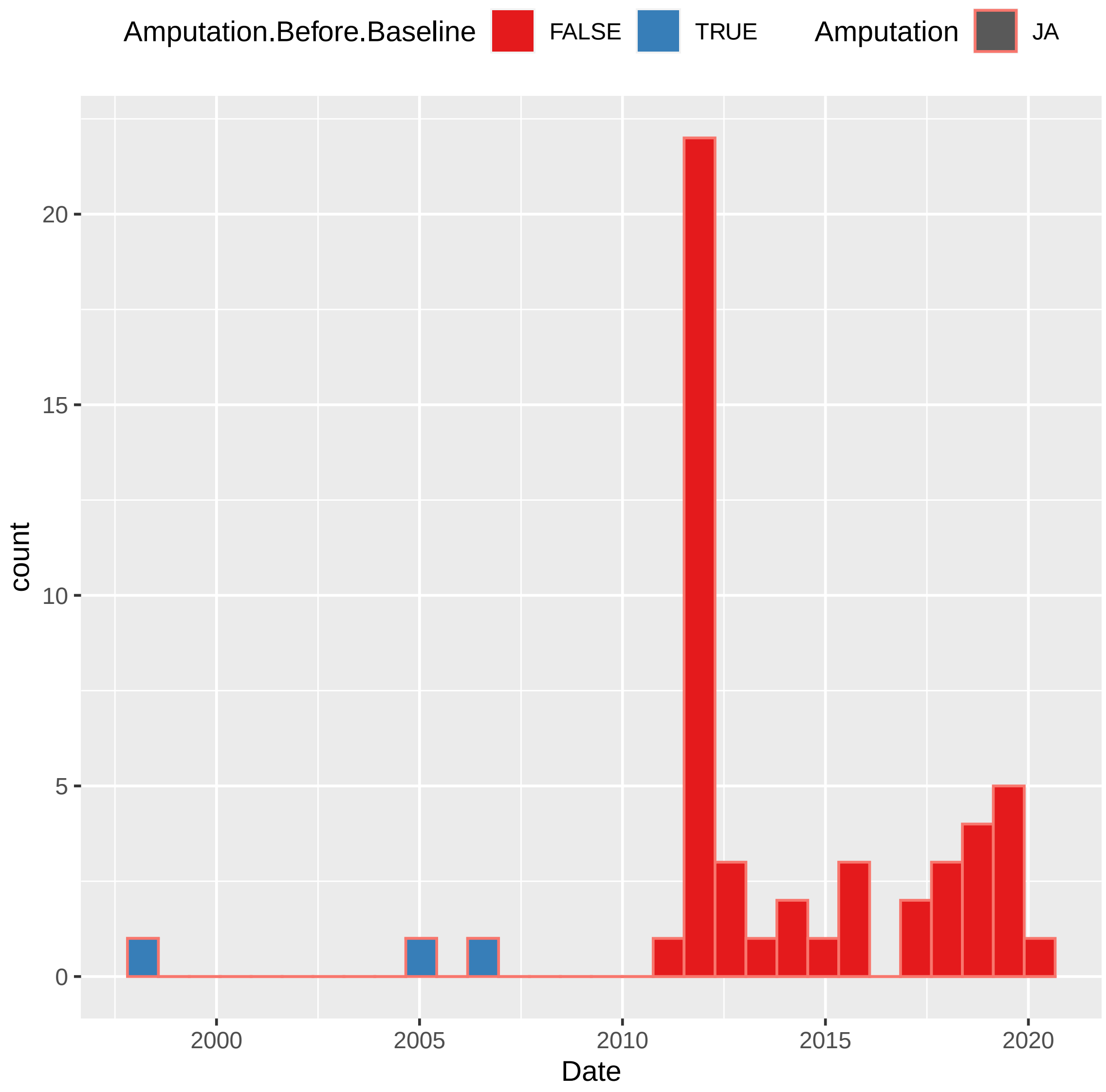

```
## `stat_bin()` using `bins = 30`. Pick better value with `binwidth`.

## Warning: Removed 510 rows containing non-finite values (stat_bin).
```

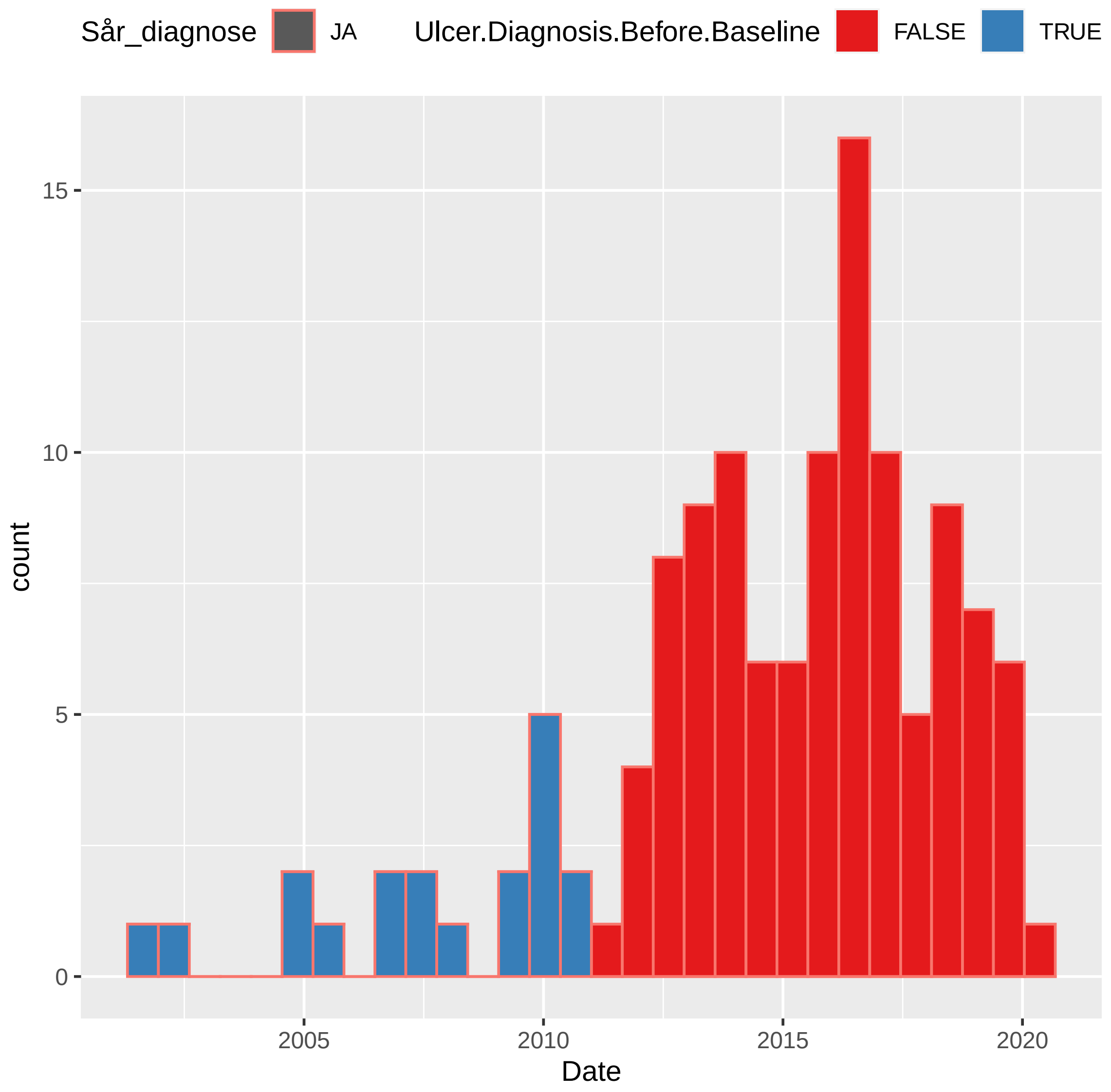
Sår_diagnose
JA
Ulcer.Diagnosis.Before.Baseline
FALSE
TRUE
count
15
10
5
0
2005
2010
2015
2020
Date

### 4 Charcot

#### 4.1 Charcot at DATE

##### 4.1.1 Crude Model

```
## [1] "Fitting models:"
## [1] "~ Charcot.at.DATE"
## [1] ""
```

###### 4.1.1.1 Forest Plot of Model Coefficients

```
## Warning: Ignoring unknown aesthetics: x
```

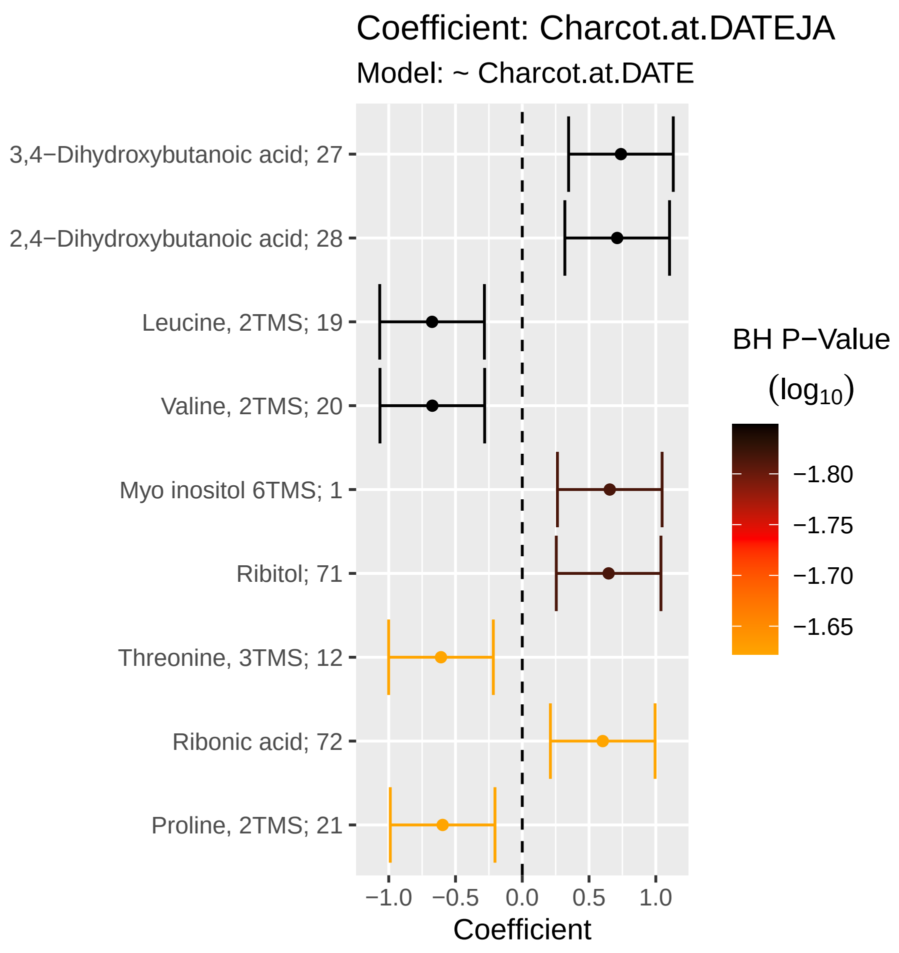

###### 4.1.1.2 Tables of Model Coefficients

```
## [1] ""
## [1] "Table: Charcot.at.DATEJA"
## [1] "  (from model: "
## [1] "   ~ Charcot.at.DATE)"
## [1] ""
##                              Name Coefficient   CI.L   CI.R  p.value  p.adj
## 1 3,4-Dihydroxybutanoic acid; 27       0.739  0.347  1.130 0.000220 0.0142
## 2 2,4-Dihydroxybutanoic acid; 28       0.711  0.319  1.100 0.000379 0.0142
## 3             Leucine, 2TMS; 19      -0.675 -1.070 -0.284 0.000731 0.0142
## 4              Valine, 2TMS; 20      -0.674 -1.070 -0.282 0.000756 0.0142
## 5           Myo inositol 6TMS; 1       0.655  0.263  1.050 0.001050 0.0153
## 6                    Ribitol; 71       0.647  0.255  1.040 0.001220 0.0153
## 7           Threonine, 3TMS; 12      -0.609 -1.000 -0.217 0.002340 0.0239
## 8              Ribonic acid; 72       0.603  0.211  0.995 0.002580 0.0239
## 9             Proline, 2TMS; 21      -0.596 -0.988 -0.204 0.002860 0.0239
```

###### 4.1.1.3 Table with All Metabolites

```
## [1] ""
## [1] "Table: Charcot.at.DATEJA"
## [1] "  (from model: "
## [1] "   ~ Charcot.at.DATE)"
## [1] ""
##                                Name Coefficient     CI.L    CI.R  p.value  p.adj
## 1  3,4-Dihydroxybutanoic acid; 27     0.73900  0.34700  1.1300 0.000220 0.0142
## 2  2,4-Dihydroxybutanoic acid; 28     0.71100  0.31900  1.1000 0.000379 0.0142
## 3               Leucine, 2TMS; 19    -0.67500 -1.07000 -0.2840 0.000731 0.0142
## 4                Valine, 2TMS; 20    -0.67400 -1.07000 -0.2820 0.000756 0.0142
## 5             Myo inositol 6TMS; 1     0.65500  0.26300  1.0500 0.001050 0.0153
## 6                      Ribitol; 71     0.64700  0.25500  1.0400 0.001220 0.0153
## 7             Threonine, 3TMS; 12    -0.60900 -1.00000 -0.2170 0.002340 0.0239
## 8                Ribonic acid; 72     0.60300  0.21100  0.9950 0.002580 0.0239
## 9               Proline, 2TMS; 21    -0.59600 -0.98800 -0.2040 0.002860 0.0239
## 10          Fumaric acid, 2TMS; 9     0.53400  0.14200  0.9260 0.007540 0.0566
## 11              Glycine, 3TMS; 17     0.51600  0.12400  0.9080 0.009900 0.0675
## 12                 Creatinine; 50     0.48900  0.09720  0.8810 0.014500 0.0776
## 13                    Ribitol; 70     0.48900  0.09710  0.8810 0.014500 0.0776
## 14           Methionine, 2TMS; 16    -0.48900 -0.88100 -0.0970 0.014500 0.0776
## 15          Pyroglutamic acid; 69     0.47700  0.08520  0.8690 0.017000 0.0852
## 16           Isoleucine, 2TMS; 18    -0.43700 -0.82900 -0.0449 0.028900 0.1260
## 17               Serine, 3TMS; 14    -0.43500 -0.82700 -0.0430 0.029600 0.1260
## 18           Citric acid, 4TMS; 6     0.43300  0.04110  0.8250 0.030400 0.1260
## 19 4-Hydroxybenzeneacetic acid; 4     0.39500  0.00346  0.7870 0.048000 0.1840
## 20         Benzeneacetic acid; 47     0.39300  0.00141  0.7850 0.049200 0.1840
## 21         Glyceryl-glycoside; 59     0.38900 -0.00317  0.7810 0.051900 0.1850
## 22             Tartronic acid; 73     0.36600 -0.02550  0.7580 0.066900 0.2240
## 23             Arabinopyranose; 51     0.36400 -0.02770  0.7560 0.068600 0.2240
## 24           Malic acid, 3TMS; 11     0.33800 -0.05410  0.7300 0.091200 0.2850
## 25              Glyceric acid; 30     0.32200 -0.06980  0.7140 0.107000 0.3210
## 26        3-Indoleacetic acid; 40     0.30600 -0.08600  0.6980 0.126000 0.3610
## 27            Dodecanoic acid; 54     0.30300 -0.08910  0.6950 0.130000 0.3610
## 28          Aminomalonic acid; 45     0.29600 -0.09580  0.6880 0.139000 0.3710
## 29           Myristoleic acid; 65     0.28000 -0.11200  0.6720 0.162000 0.4190
## 30         Succinic acid, 2TMS; 7     0.26300 -0.12900  0.6550 0.188000 0.4570
## 31       4-Deoxytetronic acid; 33    -0.26300 -0.65500  0.1290 0.189000 0.4570
## 32      Arachidonic acid, TMS; 24     0.24500 -0.14700  0.6370 0.221000 0.5130
## 33      Eicosapentaenoic acid; 55     0.24200 -0.15000  0.6340 0.226000 0.5130
## 34       4-Deoxytetronic acid; 32     0.23600 -0.15600  0.6280 0.238000 0.5250
## 35           Stearic acid, TMS; 2     0.22800 -0.16400  0.6200 0.254000 0.5440
## 36            1,3-Propanediol; 34     0.22300 -0.16900  0.6150 0.265000 0.5530
## 37                1-Dodecanol; 36     0.21300 -0.17900  0.6050 0.286000 0.5800
## 38                Bisphenol A; 48     0.20600 -0.18600  0.5980 0.303000 0.5990
## 39              Decanoic acid; 52     0.18000 -0.21200  0.5720 0.368000 0.6870
## 40           alpha-Tocopherol; 26     0.18000 -0.21200  0.5720 0.368000 0.6870
## 41           Cholesterol, TMS; 23     0.17200 -0.22000  0.5640 0.390000 0.6870
## 42     3-Indolepropionic acid; 41    -0.17100 -0.56300  0.2210 0.393000 0.6870
## 43             Oleic acid, TMS; 3     0.16900 -0.22200  0.5610 0.397000 0.6870
## 44                   Tyrosine; 75    -0.16700 -0.55900  0.2250 0.403000 0.6870
## 45         11-Eicosenoic acid; 35     0.14700 -0.24500  0.5390 0.464000 0.7580
## 46        Phenylalanine, 2TMS; 13    -0.14500 -0.53700  0.2470 0.470000 0.7580
```

```
## 47                  Glycerol; 57     0.14300 -0.24900  0.5350 0.476000 0.7580
## 48 3-Hydroxybutyric acid, 2TMS; 1    -0.14000 -0.53200  0.2520 0.485000 0.7580
## 49 4-Hydroxyphenyllactic acid; 44     0.13600 -0.25600  0.5270 0.498000 0.7620
## 50          Tridecanoic acid; 74     0.11900 -0.27300  0.5110 0.551000 0.8090
## 51               Campesterol; 49     0.11800 -0.27400  0.5100 0.555000 0.8090
## 52         Palmitic acid, TMS; 5     0.11600 -0.27600  0.5080 0.561000 0.8090
## 53              Ethanolamine; 56    -0.09800 -0.49000  0.2940 0.624000 0.8500
## 54        2-Palmitoylglycerol; 39    -0.09710 -0.48900  0.2950 0.627000 0.8500
## 55              Pyruvic acid; 31     0.09330 -0.29900  0.4850 0.641000 0.8500
## 56      Docosahexaenoic acid; 53     0.09120 -0.30100  0.4830 0.648000 0.8500
## 57              Hydroxylamine; 62    -0.09050 -0.48200  0.3010 0.651000 0.8500
## 58              Nonanoic acid; 67     0.08860 -0.30300  0.4810 0.658000 0.8500
## 59 2-Hydroxybutyric acid, 2TMS; 2    -0.08530 -0.47700  0.3070 0.670000 0.8510
## 60         Linoleic acid, TMS; 4    -0.07820 -0.47000  0.3140 0.696000 0.8700
## 61              Octanoic acid; 68    -0.06480 -0.45700  0.3270 0.746000 0.8720
## 62        Heptadecanoic acid; 60     0.06170 -0.33000  0.4540 0.758000 0.8720
## 63               Lactic acid; 29     0.06130 -0.33100  0.4530 0.759000 0.8720
## 64        Glutamic acid, 3TMS; 8    -0.06050 -0.45200  0.3310 0.762000 0.8720
## 65     4-Hydroxybutanoic acid; 43    -0.05620 -0.44800  0.3360 0.779000 0.8720
## 66                  Glycerol; 58     0.05610 -0.33600  0.4480 0.779000 0.8720
## 67 alpha-ketoglutaric acid, TMS M     0.05280 -0.33900  0.4450 0.792000 0.8720
## 68             Hydroxyproline; 64     0.04770 -0.34400  0.4400 0.811000 0.8720
## 69  2-hydroxy Isovaleric acid; 38     0.04720 -0.34500  0.4390 0.813000 0.8720
## 70         Nonadecanoic acid; 66     0.04710 -0.34500  0.4390 0.814000 0.8720
## 71        Heptadecanoic acid; 61     0.03480 -0.35700  0.4270 0.862000 0.9110
## 72             L-5-Oxoproline; 63     0.01620 -0.37600  0.4080 0.936000 0.9750
## 73              Alanine, 2TMS; 25    -0.00625 -0.39800  0.3860 0.975000 0.9810
## 74             Arachidic acid; 46    -0.00534 -0.39700  0.3870 0.979000 0.9810
## 75             1-Monopalmitin; 37     0.00470 -0.38700  0.3970 0.981000 0.9810
```

###### 4.1.2 Adjusted Model

```
## [1] "Fitting models:"
## [1] "~ Charcot.at.DATE + Age.x + Gender.x + Hba1c_baseline + CALSBP + bmi + Smoking + Statin + log_B
## [1] ""
```

###### 4.1.2.1 Forest Plot of Model Coefficients

```
## Warning: Ignoring unknown aesthetics: x
## Ignoring unknown aesthetics: x
## Ignoring unknown aesthetics: x
## Ignoring unknown aesthetics: x
## Ignoring unknown aesthetics: x
## Ignoring unknown aesthetics: x
## Ignoring unknown aesthetics: x
## Ignoring unknown aesthetics: x
## Ignoring unknown aesthetics: x
```

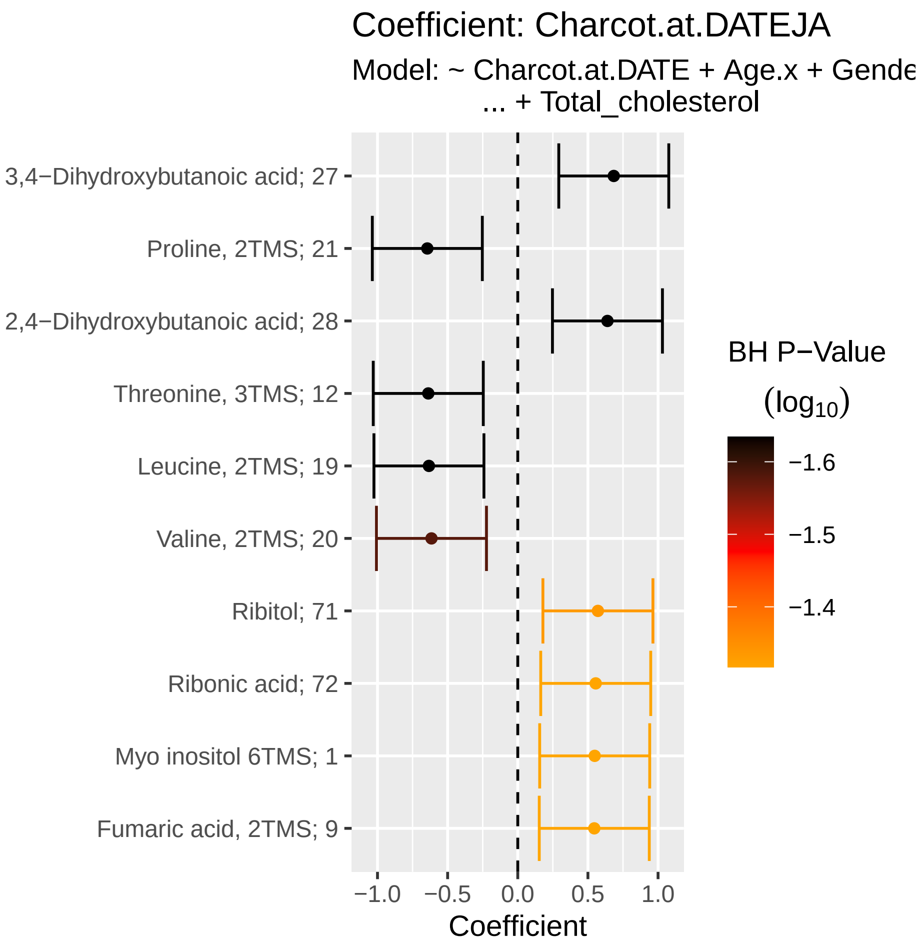

###### 4.1.2.2 Tables of Model Coefficients

```
## [1] ""
## [1] "Table: Charcot.at.DATEJA"
## [1] "  (from model: "
## [1] "   ~ Charcot.at.DATE + Age.x + Gender.x + Hba1c_baseline"
## [1] "     + CALSBP + bmi + Smoking + Statin + log_Blood_TGA +"
## [1] "     Total_cholesterol)"
## [1] ""
##                              Name Coefficient   CI.L   CI.R  p.value  p.adj
## 1  3,4-Dihydroxybutanoic acid; 27       0.684  0.292  1.080 0.000625 0.0232
## 2               Proline, 2TMS; 21      -0.645 -1.040 -0.253 0.001270 0.0232
## 3  2,4-Dihydroxybutanoic acid; 28       0.639  0.247  1.030 0.001400 0.0232
## 4             Threonine, 3TMS; 12      -0.638 -1.030 -0.246 0.001430 0.0232
## 5               Leucine, 2TMS; 19      -0.633 -1.030 -0.241 0.001550 0.0232
## 6                Valine, 2TMS; 20      -0.615 -1.010 -0.223 0.002110 0.0264
## 7                     Ribitol; 71       0.571  0.179  0.963 0.004300 0.0460
## 8                Ribonic acid; 72       0.556  0.164  0.948 0.005480 0.0481
## 9            Myo inositol 6TMS; 1       0.548  0.156  0.940 0.006130 0.0481
## 10          Fumaric acid, 2TMS; 9       0.545  0.153  0.937 0.006420 0.0481
## [1] ""
## [1] "Table: Age.x"
## [1] "  (from model: "
## [1] "   ~ Charcot.at.DATE + Age.x + Gender.x + Hba1c_baseline"
## [1] "     + CALSBP + bmi + Smoking + Statin + log_Blood_TGA +"
## [1] "     Total_cholesterol)"
## [1] ""
##                              Name Coefficient    CI.L   CI.R  p.value    p.adj
## 1       Eicosapentaenoic acid; 55     0.02180 0.01520 0.0285 1.60e-10 1.20e-08
## 2  4-Hydroxybenzeneacetic acid; 4     0.01650 0.00982 0.0232 1.33e-06 4.72e-05
## 3  2,4-Dihydroxybutanoic acid; 28     0.01630 0.00958 0.0230 1.89e-06 4.72e-05
## 4            Myo inositol 6TMS; 1     0.01440 0.00772 0.0211 2.44e-05 4.58e-04
## 5                     Ribitol; 71     0.01380 0.00708 0.0205 5.55e-05 8.33e-04
## 6        Docosahexaenoic acid; 53     0.01330 0.00660 0.0200 9.99e-05 1.25e-03
## 7          3-Indoleacetic acid; 40     0.01300 0.00631 0.0197 1.41e-04 1.51e-03
## 8                Ribonic acid; 72     0.01140 0.00470 0.0181 8.54e-04 8.01e-03
## 9           Aminomalonic acid; 45     0.01090 0.00425 0.0176 1.35e-03 1.13e-02
## 10 alpha-ketoglutaric acid, TMS M     0.01070 0.00404 0.0174 1.67e-03 1.25e-02
## 11           Malic acid, 3TMS; 11     0.01020 0.00354 0.0169 2.73e-03 1.86e-02
## 12               Pyruvic acid; 31     0.01000 0.00335 0.0167 3.27e-03 1.94e-02
## 13                    Ribitol; 70     0.00997 0.00328 0.0167 3.51e-03 1.94e-02
## 14 4-Hydroxyphenyllactic acid; 44     0.00994 0.00324 0.0166 3.63e-03 1.94e-02
## 15              Decanoic acid; 52     0.00983 0.00313 0.0165 4.02e-03 2.01e-02
## 16           Citric acid, 4TMS; 6     0.00963 0.00294 0.0163 4.80e-03 2.25e-02
## 17 3,4-Dihydroxybutanoic acid; 27     0.00951 0.00281 0.0162 5.38e-03 2.38e-02
## 18           alpha-Tocopherol; 26     0.00942 0.00273 0.0161 5.82e-03 2.39e-02
## 19          Fumaric acid, 2TMS; 9     0.00938 0.00268 0.0161 6.06e-03 2.39e-02
## 20         Succinic acid, 2TMS; 7     0.00925 0.00256 0.0159 6.74e-03 2.53e-02
## 21              Glyceric acid; 30     0.00917 0.00248 0.0159 7.24e-03 2.59e-02
## 22              Glycine, 3TMS; 17     0.00880 0.00210 0.0155 1.00e-02 3.29e-02
## 23              Alanine, 2TMS; 25     0.00879 0.00209 0.0155 1.01e-02 3.29e-02
## 24         11-Eicosenoic acid; 35     0.00855 0.00185 0.0152 1.23e-02 3.85e-02
## 25          Pyroglutamic acid; 69     0.00826 0.00157 0.0150 1.56e-02 4.67e-02
## [1] ""
```

```
## [1] "Table: Gender.x"
## [1] "  (from model: "
## [1] "   ~ Charcot.at.DATE + Age.x + Gender.x + Hba1c_baseline"
## [1] "     + CALSBP + bmi + Smoking + Statin + log_Blood_TGA +"
## [1] "     Total_cholesterol)"
## [1] ""
##                              Name Coefficient   CI.L    CI.R  p.value    p.adj
## 1           Citric acid, 4TMS; 6      -0.390 -0.548 -0.2330 1.21e-06 9.08e-05
## 2           Methionine, 2TMS; 16       0.369  0.212  0.5270 4.31e-06 1.62e-04
## 3               Valine, 2TMS; 20       0.358  0.201  0.5160 8.34e-06 2.05e-04
## 4           Myristoleic acid; 65      -0.353 -0.511 -0.1960 1.09e-05 2.05e-04
## 5              Proline, 2TMS; 21       0.349  0.192  0.5070 1.40e-05 2.09e-04
## 6              Glycine, 3TMS; 17      -0.339 -0.496 -0.1810 2.47e-05 2.96e-04
## 7              Leucine, 2TMS; 19       0.336  0.178  0.4930 2.93e-05 2.96e-04
## 8             Tartronic acid; 73      -0.334 -0.492 -0.1770 3.23e-05 2.96e-04
## 9           Isoleucine, 2TMS; 18       0.332  0.175  0.4900 3.55e-05 2.96e-04
## 10             Glyceric acid; 30      -0.326 -0.483 -0.1680 5.04e-05 3.78e-04
## 11           Dodecanoic acid; 54      -0.323 -0.480 -0.1650 5.92e-05 4.04e-04
## 12      4-Deoxytetronic acid; 33       0.305  0.148  0.4630 1.46e-04 9.14e-04
## 13            Oleic acid, TMS; 3      -0.296 -0.454 -0.1390 2.26e-04 1.30e-03
## 14         Aminomalonic acid; 45      -0.292 -0.449 -0.1340 2.82e-04 1.51e-03
## 15 2-hydroxy Isovaleric acid; 38       0.290  0.132  0.4470 3.15e-04 1.57e-03
## 16           Cholesterol, TMS; 23      -0.270 -0.428 -0.1130 7.63e-04 3.58e-03
## 17         Glutamic acid, 3TMS; 8       0.256  0.099  0.4140 1.41e-03 6.17e-03
## 18      Docosahexaenoic acid; 53      -0.255 -0.412 -0.0970 1.54e-03 6.17e-03
## 19           Tridecanoic acid; 74      -0.253 -0.411 -0.0959 1.62e-03 6.17e-03
## 20         Heptadecanoic acid; 60      -0.253 -0.410 -0.0955 1.65e-03 6.17e-03
## 21              Decanoic acid; 52      -0.247 -0.405 -0.0896 2.11e-03 7.53e-03
## 22         Heptadecanoic acid; 61      -0.245 -0.403 -0.0877 2.28e-03 7.77e-03
## 23           Myo inositol 6TMS; 1      -0.241 -0.399 -0.0837 2.69e-03 8.77e-03
## 24         Succinic acid, 2TMS; 7      -0.238 -0.396 -0.0808 3.02e-03 9.44e-03
## 25          Nonadecanoic acid; 66      -0.221 -0.378 -0.0631 6.04e-03 1.81e-02
## 26          Palmitic acid, TMS; 5      -0.205 -0.363 -0.0478 1.06e-02 3.06e-02
## 27           Stearic acid, TMS; 2      -0.196 -0.354 -0.0386 1.47e-02 4.08e-02
## 28         Benzeneacetic acid; 47      -0.190 -0.347 -0.0322 1.82e-02 4.88e-02
## [1] ""
## [1] "Table: Hba1c_baseline"
## [1] "  (from model: "
## [1] "   ~ Charcot.at.DATE + Age.x + Gender.x + Hba1c_baseline"
## [1] "     + CALSBP + bmi + Smoking + Statin + log_Blood_TGA +"
## [1] "     Total_cholesterol)"
## [1] ""
##                         Name Coefficient    CI.L    CI.R  p.value  p.adj
## 1      Tridecanoic acid; 74      -0.131 -0.2020 -0.0590 0.000348 0.0145
## 2 Eicosapentaenoic acid; 55      -0.127 -0.1990 -0.0560 0.000476 0.0145
## 3          Valine, 2TMS; 20       0.122  0.0503  0.1930 0.000844 0.0145
## 4         Glyceric acid; 30      -0.120 -0.1920 -0.0489 0.000967 0.0145
## 5          Ethanolamine; 56       0.119  0.0477  0.1910 0.001080 0.0145
## 6       Arabinopyranose; 51       0.119  0.0470  0.1900 0.001160 0.0145
## 7  Docosahexaenoic acid; 53      -0.116 -0.1870 -0.0440 0.001550 0.0166
## 8         Alanine, 2TMS; 25       0.110  0.0380  0.1810 0.002680 0.0251
## [1] ""
## [1] "Table: CALSBP"
## [1] "  (from model: "
```

```
## [1] "   ~ Charcot.at.DATE + Age.x + Gender.x + Hba1c_baseline"
## [1] "     + CALSBP + bmi + Smoking + Statin + log_Blood_TGA +"
## [1] "     Total_cholesterol)"
## [1] ""
## [1] "No significant associations at p.adj < 0.05"
## [1] ""
## [1] "Table: bmi"
## [1] "  (from model: "
## [1] "   ~ Charcot.at.DATE + Age.x + Gender.x + Hba1c_baseline"
## [1] "     + CALSBP + bmi + Smoking + Statin + log_Blood_TGA +"
## [1] "     Total_cholesterol)"
## [1] ""
##                             Name Coefficient     CI.L     CI.R  p.value
## 1         Glutamic acid, 3TMS; 8      0.0458  0.02520  0.06630 1.31e-05
## 2 2-Hydroxybutyric acid, 2TMS; 2      0.0403  0.01970  0.06090 1.24e-04
## 3                Campesterol; 49     -0.0400 -0.06060 -0.01940 1.39e-04
## 4              Decanoic acid; 52     -0.0312 -0.05180 -0.01060 2.95e-03
## 5               Pyruvic acid; 31     -0.0310 -0.05160 -0.01040 3.17e-03
## 6            1,3-Propanediol; 34     -0.0305 -0.05110 -0.00992 3.68e-03
## 7             Arachidic acid; 46     -0.0300 -0.05050 -0.00939 4.32e-03
## 8                Lactic acid; 29      0.0294  0.00878  0.04990 5.17e-03
##      p.adj
## 1 0.000983
## 2 0.003470
## 3 0.003470
## 4 0.046000
## 5 0.046000
## 6 0.046000
## 7 0.046300
## 8 0.048500
## [1] ""
## [1] "Table: Smoking"
## [1] "  (from model: "
## [1] "   ~ Charcot.at.DATE + Age.x + Gender.x + Hba1c_baseline"
## [1] "     + CALSBP + bmi + Smoking + Statin + log_Blood_TGA +"
## [1] "     Total_cholesterol)"
## [1] ""
##                          Name Coefficient   CI.L    CI.R  p.value   p.adj
## 1      Glutamic acid, 3TMS; 8       0.385  0.192  0.5770 9.13e-05 0.00538
## 2    Docosahexaenoic acid; 53      -0.362 -0.555 -0.1690 2.30e-04 0.00538
## 3  3-Indolepropionic acid; 41      -0.361 -0.554 -0.1680 2.40e-04 0.00538
## 4          Tartronic acid; 73      -0.356 -0.549 -0.1640 2.87e-04 0.00538
## 5           Glyceric acid; 30      -0.329 -0.522 -0.1370 8.08e-04 0.01210
## 6        Citric acid, 4TMS; 6      -0.313 -0.506 -0.1210 1.43e-03 0.01780
## 7        alpha-Tocopherol; 26      -0.296 -0.489 -0.1040 2.57e-03 0.02760
## 8      Benzeneacetic acid; 47      -0.286 -0.479 -0.0935 3.60e-03 0.03370
## 9            Ribonic acid; 72      -0.279 -0.472 -0.0868 4.47e-03 0.03720
## 10       Malic acid, 3TMS; 11      -0.271 -0.463 -0.0780 5.89e-03 0.04350
## 11            Campesterol; 49      -0.268 -0.461 -0.0754 6.38e-03 0.04350
## [1] ""
## [1] "Table: Statin"
## [1] "  (from model: "
## [1] "   ~ Charcot.at.DATE + Age.x + Gender.x + Hba1c_baseline"
## [1] "     + CALSBP + bmi + Smoking + Statin + log_Blood_TGA +"
```

```
## [1] "     Total_cholesterol)"
## [1] ""
##                 Name Coefficient   CI.L   CI.R  p.value  p.adj
## 1 L-5-Oxoproline; 63      -0.335 -0.508 -0.162 0.000143 0.0107
## [1] ""
## [1] "Table: log_Blood_TGA"
## [1] "  (from model: "
## [1] "   ~ Charcot.at.DATE + Age.x + Gender.x + Hba1c_baseline"
## [1] "     + CALSBP + bmi + Smoking + Statin + log_Blood_TGA +"
## [1] "     Total_cholesterol)"
## [1] ""
##                              Name Coefficient   CI.L  CI.R  p.value    p.adj
## 1             Dodecanoic acid; 54       0.287 0.1570 0.416 1.43e-05 0.000915
## 2           Palmitic acid, TMS; 5       0.279 0.1490 0.408 2.44e-05 0.000915
## 3            Stearic acid, TMS; 2       0.264 0.1340 0.393 6.59e-05 0.001370
## 4               Decanoic acid; 52       0.262 0.1330 0.392 7.29e-05 0.001370
## 5  3,4-Dihydroxybutanoic acid; 27       0.253 0.1230 0.382 1.29e-04 0.001690
## 6              Arachidic acid; 46       0.252 0.1230 0.382 1.35e-04 0.001690
## 7                Ribonic acid; 72       0.239 0.1090 0.368 3.01e-04 0.002920
## 8               Octanoic acid; 68       0.238 0.1090 0.368 3.11e-04 0.002920
## 9          Glyceryl-glycoside; 59       0.234 0.1050 0.364 3.91e-04 0.003260
## 10 4-Hydroxybenzeneacetic acid; 4       0.223 0.0934 0.352 7.41e-04 0.005560
## 11                    Ribitol; 71       0.216 0.0864 0.345 1.08e-03 0.007020
## 12             Oleic acid, TMS; 3       0.215 0.0858 0.345 1.12e-03 0.007020
## 13           Arabinopyranose; 51       0.204 0.0745 0.333 2.02e-03 0.011700
## 14 2,4-Dihydroxybutanoic acid; 28       0.193 0.0637 0.323 3.46e-03 0.018500
## 15         Heptadecanoic acid; 60       0.187 0.0571 0.316 4.73e-03 0.023700
## 16          Fumaric acid, 2TMS; 9       0.183 0.0531 0.312 5.72e-03 0.026800
## 17 2-Hydroxybutyric acid, 2TMS; 2       0.179 0.0497 0.309 6.69e-03 0.029500
## 18            Myo inositol 6TMS; 1       0.174 0.0447 0.304 8.37e-03 0.034600
## 19                 Lactic acid; 29       0.173 0.0437 0.303 8.77e-03 0.034600
## 20         Heptadecanoic acid; 61       0.170 0.0401 0.299 1.03e-02 0.038500
## 21           Myristoleic acid; 65       0.165 0.0360 0.295 1.23e-02 0.043800
## 22               Pyruvic acid; 31       0.164 0.0342 0.293 1.32e-02 0.045100
## 23           Malic acid, 3TMS; 11       0.162 0.0327 0.292 1.41e-02 0.045900
## 24         Glutamic acid, 3TMS; 8       0.160 0.0303 0.289 1.56e-02 0.048800
## [1] ""
## [1] "Table: Total_cholesterol"
## [1] "  (from model: "
## [1] "   ~ Charcot.at.DATE + Age.x + Gender.x + Hba1c_baseline"
## [1] "     + CALSBP + bmi + Smoking + Statin + log_Blood_TGA +"
## [1] "     Total_cholesterol)"
## [1] ""
##                              Name Coefficient    CI.L    CI.R  p.value    p.adj
## 1            Cholesterol, TMS; 23       0.464  0.3680  0.5600 3.57e-21 2.68e-19
## 2                 Campesterol; 49       0.372  0.2750  0.4680 3.75e-14 1.41e-12
## 3            alpha-Tocopherol; 26       0.330  0.2340  0.4260 1.85e-11 4.63e-10
## 4          Benzeneacetic acid; 47      -0.190 -0.2860 -0.0938 1.09e-04 2.04e-03
## 5      4-Hydroxybutanoic acid; 43      -0.184 -0.2810 -0.0881 1.73e-04 2.44e-03
## 6               Proline, 2TMS; 21      -0.183 -0.2790 -0.0867 1.95e-04 2.44e-03
## 7           Linoleic acid, TMS; 4       0.181  0.0844  0.2770 2.33e-04 2.50e-03
## 8              L-5-Oxoproline; 63      -0.166 -0.2620 -0.0697 7.28e-04 6.61e-03
## 9               Glycine, 3TMS; 17      -0.165 -0.2610 -0.0685 7.93e-04 6.61e-03
## 10 2,4-Dihydroxybutanoic acid; 28      -0.162 -0.2580 -0.0659 9.63e-04 7.22e-03
```

```
## 11           Isoleucine, 2TMS; 18      -0.156 -0.2520 -0.0594 1.53e-03 1.04e-02
## 12 4-Hydroxybenzeneacetic acid; 4      -0.154 -0.2500 -0.0577 1.72e-03 1.07e-02
## 13      Eicosapentaenoic acid; 55       0.152  0.0560  0.2480 1.93e-03 1.12e-02
## 14       Docosahexaenoic acid; 53       0.148  0.0513  0.2440 2.66e-03 1.42e-02
## 15                   Tyrosine; 75      -0.146 -0.2420 -0.0495 3.00e-03 1.50e-02
## 16              Alanine, 2TMS; 25      -0.144 -0.2410 -0.0481 3.28e-03 1.52e-02
## 17            Threonine, 3TMS; 12      -0.144 -0.2400 -0.0474 3.45e-03 1.52e-02
## 18           Methionine, 2TMS; 16      -0.142 -0.2380 -0.0460 3.76e-03 1.57e-02
## 19         Glyceryl-glycoside; 59      -0.141 -0.2370 -0.0444 4.17e-03 1.65e-02
## 20            Arabinopyranose; 51      -0.136 -0.2330 -0.0402 5.46e-03 2.00e-02
## 21        2-Palmitoylglycerol; 39       0.136  0.0398  0.2320 5.60e-03 2.00e-02
## 22                    Ribitol; 71      -0.135 -0.2310 -0.0384 6.10e-03 2.08e-02
## 23        3-Indoleacetic acid; 40      -0.127 -0.2230 -0.0308 9.68e-03 3.16e-02
## 24              Hydroxylamine; 62      -0.126 -0.2220 -0.0296 1.04e-02 3.24e-02
## 25               Serine, 3TMS; 14      -0.121 -0.2170 -0.0250 1.36e-02 4.04e-02
## 26               Ribonic acid; 72      -0.121 -0.2170 -0.0244 1.40e-02 4.04e-02
```

###### 4.1.2.3 Table with All Metabolites

```
## [1] ""
## [1] "Table: Charcot.at.DATEJA"
## [1] "  (from model: "
## [1] "   ~ Charcot.at.DATE + Age.x + Gender.x + Hba1c_baseline"
## [1] "     + CALSBP + bmi + Smoking + Statin + log_Blood_TGA +"
## [1] "     Total_cholesterol)"
## [1] ""
##                               Name Coefficient     CI.L    CI.R  p.value  p.adj
## 1  3,4-Dihydroxybutanoic acid; 27     0.68400  0.29200  1.0800 0.000625 0.0232
## 2                Proline, 2TMS; 21    -0.64500 -1.04000 -0.2530 0.001270 0.0232
## 3  2,4-Dihydroxybutanoic acid; 28     0.63900  0.24700  1.0300 0.001400 0.0232
## 4              Threonine, 3TMS; 12    -0.63800 -1.03000 -0.2460 0.001430 0.0232
## 5                Leucine, 2TMS; 19    -0.63300 -1.03000 -0.2410 0.001550 0.0232
## 6                 Valine, 2TMS; 20    -0.61500 -1.01000 -0.2230 0.002110 0.0264
## 7                      Ribitol; 71     0.57100  0.17900  0.9630 0.004300 0.0460
## 8                 Ribonic acid; 72     0.55600  0.16400  0.9480 0.005480 0.0481
## 9             Myo inositol 6TMS; 1     0.54800  0.15600  0.9400 0.006130 0.0481
## 10          Fumaric acid, 2TMS; 9     0.54500  0.15300  0.9370 0.006420 0.0481
## 11                  Creatinine; 50     0.51000  0.11800  0.9030 0.010700 0.0686
## 12                Serine, 3TMS; 14    -0.50900 -0.90100 -0.1170 0.011000 0.0686
## 13            Methionine, 2TMS; 16    -0.47500 -0.86700 -0.0825 0.017700 0.0958
## 14                     Ribitol; 70     0.47400  0.08170  0.8660 0.017900 0.0958
## 15           Pyroglutamic acid; 69     0.43300  0.04090  0.8250 0.030400 0.1520
## 16            Isoleucine, 2TMS; 18    -0.41400 -0.80600 -0.0216 0.038700 0.1810
## 17          Glyceryl-glycoside; 59     0.39700  0.00506  0.7890 0.047100 0.2080
## 18              Tartronic acid; 73     0.36200 -0.03030  0.7540 0.070500 0.2860
## 19              Arabinopyranose; 51     0.35900 -0.03270  0.7520 0.072400 0.2860
## 20               Glycine, 3TMS; 17     0.34500 -0.04660  0.7380 0.084200 0.3030
## 21            Citric acid, 4TMS; 6     0.34500 -0.04730  0.7370 0.084800 0.3030
## 22            Stearic acid, TMS; 2     0.33200 -0.06020  0.7240 0.097100 0.3310
## 23              Dodecanoic acid; 54     0.32600 -0.06570  0.7190 0.103000 0.3350
## 24            Cholesterol, TMS; 23     0.32200 -0.07060  0.7140 0.108000 0.3380
## 25              1,3-Propanediol; 34     0.31700 -0.07540  0.7090 0.113000 0.3400
## 26            Malic acid, 3TMS; 11     0.30500 -0.08710  0.6970 0.127000 0.3670
## 27               Glyceric acid; 30     0.29300 -0.09910  0.6850 0.143000 0.3720
## 28            alpha-Tocopherol; 26     0.29000 -0.10200  0.6820 0.147000 0.3720
## 29       Arachidonic acid, TMS; 24     0.28900 -0.10300  0.6810 0.148000 0.3720
## 30           Benzeneacetic acid; 47     0.28900 -0.10300  0.6810 0.149000 0.3720
## 31                 1-Dodecanol; 36     0.28500 -0.10700  0.6780 0.154000 0.3720
## 32 4-Hydroxybenzeneacetic acid; 4     0.27600 -0.11600  0.6680 0.168000 0.3930
## 33            Myristoleic acid; 65     0.26500 -0.12700  0.6570 0.185000 0.4150
## 34        4-Deoxytetronic acid; 32     0.26000 -0.13200  0.6530 0.193000 0.4150
## 35          3-Indoleacetic acid; 40     0.25800 -0.13500  0.6500 0.198000 0.4150
## 36               Decanoic acid; 52     0.25500 -0.13700  0.6480 0.202000 0.4150
## 37                    Tyrosine; 75    -0.25400 -0.64600  0.1380 0.205000 0.4150
## 38          Succinic acid, 2TMS; 7     0.24800 -0.14400  0.6400 0.215000 0.4240
## 39                 Campesterol; 49     0.24100 -0.15200  0.6330 0.229000 0.4410
## 40         Phenylalanine, 2TMS; 13    -0.22100 -0.61300  0.1710 0.269000 0.5050
## 41                 Bisphenol A; 48     0.20800 -0.18400  0.6000 0.299000 0.5460
## 42       Eicosapentaenoic acid; 55     0.19500 -0.19700  0.5870 0.330000 0.5900
## 43             Oleic acid, TMS; 3     0.19100 -0.20200  0.5830 0.341000 0.5940
## 44        4-Deoxytetronic acid; 33    -0.18100 -0.57400  0.2110 0.364000 0.6210
```

```
## 45          Palmitic acid, TMS; 5     0.17500 -0.21700  0.5670 0.383000 0.6250
## 46          Aminomalonic acid; 45     0.17400 -0.21800  0.5670 0.383000 0.6250
## 47           Tridecanoic acid; 74     0.17000 -0.22200  0.5620 0.396000 0.6320
## 48              Hydroxylamine; 62    -0.16300 -0.55500  0.2290 0.416000 0.6380
## 49  2-hydroxy Isovaleric acid; 38     0.16200 -0.23000  0.5550 0.417000 0.6380
## 50               Ethanolamine; 56    -0.14600 -0.53900  0.2460 0.464000 0.6910
## 51              Nonanoic acid; 67     0.14200 -0.25000  0.5340 0.478000 0.6910
## 52    3-Indolepropionic acid; 41    -0.14200 -0.53400  0.2500 0.479000 0.6910
## 53 4-Hydroxyphenyllactic acid; 44     0.11900 -0.27300  0.5110 0.552000 0.7680
## 54               Pyruvic acid; 31     0.11900 -0.27300  0.5110 0.553000 0.7680
## 55              Alanine, 2TMS; 25    -0.11600 -0.50800  0.2760 0.563000 0.7680
## 56 3-Hydroxybutyric acid, 2TMS; 1    -0.11000 -0.50200  0.2830 0.584000 0.7820
## 57             Arachidic acid; 46     0.10300 -0.28900  0.4960 0.605000 0.7940
## 58         11-Eicosenoic acid; 35     0.09700 -0.29500  0.4890 0.628000 0.7940
## 59                   Glycerol; 57     0.09550 -0.29700  0.4880 0.633000 0.7940
## 60         Heptadecanoic acid; 60     0.08850 -0.30400  0.4810 0.658000 0.7940
## 61       Docosahexaenoic acid; 53     0.08830 -0.30400  0.4800 0.659000 0.7940
## 62 alpha-ketoglutaric acid, TMS M     0.08710 -0.30500  0.4790 0.663000 0.7940
## 63             1-Monopalmitin; 37     0.08600 -0.30600  0.4780 0.667000 0.7940
## 64              Hydroxyproline; 64     0.07640 -0.31600  0.4690 0.702000 0.8220
## 65          Nonadecanoic acid; 66     0.07090 -0.32100  0.4630 0.723000 0.8220
## 66 2-Hydroxybutyric acid, 2TMS; 2    -0.07090 -0.46300  0.3210 0.723000 0.8220
## 67              Octanoic acid; 68     0.06630 -0.32600  0.4580 0.740000 0.8290
## 68                Lactic acid; 29     0.04970 -0.34200  0.4420 0.804000 0.8690
## 69             L-5-Oxoproline; 63    -0.04640 -0.43800  0.3460 0.817000 0.8690
## 70     4-Hydroxybutanoic acid; 43    -0.04530 -0.43700  0.3470 0.821000 0.8690
## 71        2-Palmitoylglycerol; 39    -0.04470 -0.43700  0.3470 0.823000 0.8690
## 72         Heptadecanoic acid; 61     0.03620 -0.35600  0.4280 0.857000 0.8870
## 73          Linoleic acid, TMS; 4     0.03320 -0.35900  0.4250 0.868000 0.8870
## 74         Glutamic acid, 3TMS; 8    -0.03140 -0.42400  0.3610 0.875000 0.8870
## 75                   Glycerol; 58     0.00871 -0.38300  0.4010 0.965000 0.9650
```

###### 4.1.3 Adjusted Model with eGFR

```
## [1] "Fitting models:"
## [1] "~ Charcot.at.DATE + Age.x + Gender.x + Hba1c_baseline + CALSBP + bmi + Smoking + Statin + log_B[illegible]
## [1] ""
```

###### 4.1.3.1 Forest Plot of Model Coefficients

```
## Warning: Ignoring unknown aesthetics: x
## Ignoring unknown aesthetics: x
## Ignoring unknown aesthetics: x
## Ignoring unknown aesthetics: x
## Ignoring unknown aesthetics: x
## Ignoring unknown aesthetics: x
## Ignoring unknown aesthetics: x
## Ignoring unknown aesthetics: x
## Ignoring unknown aesthetics: x
```

```
## NULL
```

###### 4.1.3.2 Tables of Model Coefficients

```
## [1] ""
## [1] "Table: Charcot.at.DATEJA"
## [1] "  (from model: "
## [1] "   ~ Charcot.at.DATE + Age.x + Gender.x + Hba1c_baseline"
## [1] "     + CALSBP + bmi + Smoking + Statin + log_Blood_TGA +"
## [1] "     Total_cholesterol + egfr)"
## [1] ""
## [1] "No significant associations at p.adj < 0.05"
## [1] ""
## [1] "Table: Age.x"
## [1] "  (from model: "
## [1] "   ~ Charcot.at.DATE + Age.x + Gender.x + Hba1c_baseline"
## [1] "     + CALSBP + bmi + Smoking + Statin + log_Blood_TGA +"
## [1] "     Total_cholesterol + egfr)"
## [1] ""
##                              Name Coefficient    CI.L   CI.R  p.value    p.adj
## 1       Eicosapentaenoic acid; 55     0.02430 0.01760 0.0311 3.04e-12 2.28e-10
## 2        Docosahexaenoic acid; 53     0.01510 0.00825 0.0219 1.58e-05 5.92e-04
## 3               Glyceric acid; 30     0.01160 0.00478 0.0184 8.68e-04 1.63e-02
## 4                Pyruvic acid; 31     0.01140 0.00444 0.0183 1.31e-03 1.63e-02
## 5            alpha-Tocopherol; 26     0.01110 0.00430 0.0179 1.41e-03 1.63e-02
## 6               Decanoic acid; 52     0.01120 0.00432 0.0181 1.44e-03 1.63e-02
## 7  alpha-ketoglutaric acid, TMS M     0.01120 0.00430 0.0182 1.52e-03 1.63e-02
## 8  4-Hydroxybenzeneacetic acid; 4     0.00989 0.00313 0.0167 4.17e-03 3.83e-02
## 9           Aminomalonic acid; 45     0.00995 0.00307 0.0168 4.59e-03 3.83e-02
## 10        3-Indoleacetic acid; 40     0.00961 0.00274 0.0165 6.15e-03 4.62e-02
## [1] ""
## [1] "Table: Gender.x"
## [1] "  (from model: "
## [1] "   ~ Charcot.at.DATE + Age.x + Gender.x + Hba1c_baseline"
## [1] "     + CALSBP + bmi + Smoking + Statin + log_Blood_TGA +"
## [1] "     Total_cholesterol + egfr)"
## [1] ""
##                             Name Coefficient    CI.L    CI.R  p.value    p.adj
## 1       4-Deoxytetronic acid; 33       0.425  0.2670  0.5830 1.38e-07 1.04e-05
## 2              Glyceric acid; 30      -0.370 -0.5280 -0.2120 4.54e-06 1.33e-04
## 3           Myristoleic acid; 65      -0.370 -0.5290 -0.2110 5.33e-06 1.33e-04
## 4              Proline, 2TMS; 21       0.352  0.1940  0.5110 1.38e-05 2.58e-04
## 5             Tartronic acid; 73      -0.344 -0.5020 -0.1850 2.20e-05 3.30e-04
## 6            Dodecanoic acid; 54      -0.333 -0.4930 -0.1740 4.19e-05 5.24e-04
## 7           Cholesterol, TMS; 23      -0.311 -0.4660 -0.1570 8.07e-05 8.64e-04
## 8              Oleic acid, TMS; 3      -0.313 -0.4720 -0.1530 1.27e-04 1.19e-03
## 9           Methionine, 2TMS; 16       0.309  0.1500  0.4680 1.42e-04 1.19e-03
## 10          Citric acid, 4TMS; 6      -0.305 -0.4630 -0.1470 1.58e-04 1.19e-03
## 11      Docosahexaenoic acid; 53      -0.290 -0.4480 -0.1320 3.33e-04 2.27e-03
## 12             Glycine, 3TMS; 17      -0.287 -0.4460 -0.1290 3.95e-04 2.47e-03
## 13             Leucine, 2TMS; 19       0.272  0.1140  0.4310 7.56e-04 4.17e-03
## 14              Valine, 2TMS; 20       0.267  0.1110  0.4240 8.30e-04 4.17e-03
## 15         Aminomalonic acid; 45      -0.271 -0.4300 -0.1120 8.46e-04 4.17e-03
## 16             Decanoic acid; 52      -0.270 -0.4290 -0.1110 8.90e-04 4.17e-03
## 17            Hydroxyproline; 64       0.258  0.0975  0.4180 1.63e-03 7.06e-03
## 18          Stearic acid, TMS; 2      -0.256 -0.4160 -0.0963 1.69e-03 7.06e-03
```

```
## 19          Tridecanoic acid; 74      -0.255 -0.4160 -0.0951 1.82e-03 7.17e-03
## 20         Palmitic acid, TMS; 5      -0.250 -0.4100 -0.0913 2.06e-03 7.41e-03
## 21        Heptadecanoic acid; 61      -0.252 -0.4130 -0.0916 2.09e-03 7.41e-03
## 22        Heptadecanoic acid; 60      -0.251 -0.4120 -0.0907 2.17e-03 7.41e-03
## 23        Glutamic acid, 3TMS; 8       0.239  0.0811  0.3970 3.02e-03 9.83e-03
## 24 2-hydroxy Isovaleric acid; 38       0.237  0.0768  0.3970 3.75e-03 1.17e-02
## 25          Isoleucine, 2TMS; 18       0.231  0.0737  0.3880 4.01e-03 1.20e-02
## 26                  Tyrosine; 75      -0.227 -0.3870 -0.0666 5.55e-03 1.57e-02
## 27         Nonadecanoic acid; 66      -0.227 -0.3880 -0.0664 5.64e-03 1.57e-02
## 28        Succinic acid, 2TMS; 7      -0.217 -0.3780 -0.0567 8.03e-03 2.15e-02
## [1] ""
## [1] "Table: Hba1c_baseline"
## [1] "  (from model: "
## [1] "   ~ Charcot.at.DATE + Age.x + Gender.x + Hba1c_baseline"
## [1] "     + CALSBP + bmi + Smoking + Statin + log_Blood_TGA +"
## [1] "     Total_cholesterol + egfr)"
## [1] ""
##                         Name Coefficient    CI.L    CI.R  p.value   p.adj
## 1 Eicosapentaenoic acid; 55      -0.137 -0.2070 -0.0664 0.000140 0.00928
## 2      Tridecanoic acid; 74      -0.134 -0.2060 -0.0625 0.000247 0.00928
## 3  Docosahexaenoic acid; 53      -0.124 -0.1940 -0.0530 0.000608 0.01330
## 4         Glyceric acid; 30      -0.122 -0.1920 -0.0514 0.000709 0.01330
## 5         Alanine, 2TMS; 25       0.119  0.0472  0.1900 0.001150 0.01590
## 6       Arabinopyranose; 51       0.116  0.0450  0.1880 0.001410 0.01590
## 7          Valine, 2TMS; 20       0.113  0.0429  0.1830 0.001580 0.01590
## 8          Ethanolamine; 56       0.115  0.0433  0.1870 0.001700 0.01590
## 9         Decanoic acid; 52      -0.101 -0.1720 -0.0298 0.005420 0.04520
## [1] ""
## [1] "Table: CALSBP"
## [1] "  (from model: "
## [1] "   ~ Charcot.at.DATE + Age.x + Gender.x + Hba1c_baseline"
## [1] "     + CALSBP + bmi + Smoking + Statin + log_Blood_TGA +"
## [1] "     Total_cholesterol + egfr)"
## [1] ""
## [1] "No significant associations at p.adj < 0.05"
## [1] ""
## [1] "Table: bmi"
## [1] "  (from model: "
## [1] "   ~ Charcot.at.DATE + Age.x + Gender.x + Hba1c_baseline"
## [1] "     + CALSBP + bmi + Smoking + Statin + log_Blood_TGA +"
## [1] "     Total_cholesterol + egfr)"
## [1] ""
##                              Name Coefficient    CI.L     CI.R  p.value    p.adj
## 1        Glutamic acid, 3TMS; 8      0.0462  0.0260  0.06640 7.70e-06 0.000577
## 2 2-Hydroxybutyric acid, 2TMS; 2      0.0402  0.0201  0.06040 9.52e-05 0.003150
## 3               Campesterol; 49     -0.0396 -0.0598 -0.01940 1.26e-04 0.003150
## 4             Decanoic acid; 52     -0.0308 -0.0512 -0.01040 3.08e-03 0.045800
## 5              Pyruvic acid; 31     -0.0306 -0.0511 -0.01010 3.48e-03 0.045800
## 6           1,3-Propanediol; 34     -0.0304 -0.0510 -0.00982 3.83e-03 0.045800
## 7               Lactic acid; 29      0.0299  0.0094  0.05040 4.28e-03 0.045800
## 8           Arachidic acid; 46     -0.0294 -0.0500 -0.00885 5.07e-03 0.047500
## [1] ""
## [1] "Table: Smoking"
## [1] "  (from model: "
```

```
## [1] "   ~ Charcot.at.DATE + Age.x + Gender.x + Hba1c_baseline"
## [1] "     + CALSBP + bmi + Smoking + Statin + log_Blood_TGA +"
## [1] "     Total_cholesterol + egfr)"
## [1] ""
##                          Name Coefficient   CI.L    CI.R  p.value   p.adj
## 1    Docosahexaenoic acid; 53      -0.384 -0.574 -0.1940 7.49e-05 0.00373
## 2      Glutamic acid, 3TMS; 8       0.374  0.185  0.5640 1.12e-04 0.00373
## 3  3-Indolepropionic acid; 41      -0.372 -0.564 -0.1800 1.49e-04 0.00373
## 4          Tartronic acid; 73      -0.354 -0.545 -0.1640 2.71e-04 0.00489
## 5           Glyceric acid; 30      -0.348 -0.537 -0.1580 3.26e-04 0.00489
## 6        alpha-Tocopherol; 26      -0.309 -0.499 -0.1200 1.38e-03 0.01690
## 7             Valine, 2TMS; 20      -0.304 -0.492 -0.1150 1.58e-03 0.01690
## 8        Citric acid, 4TMS; 6      -0.269 -0.459 -0.0795 5.45e-03 0.04830
## 9             Campesterol; 49      -0.264 -0.454 -0.0745 6.35e-03 0.04830
## 10     Benzeneacetic acid; 47      -0.267 -0.459 -0.0750 6.44e-03 0.04830
## [1] ""
## [1] "Table: Statin"
## [1] "  (from model: "
## [1] "   ~ Charcot.at.DATE + Age.x + Gender.x + Hba1c_baseline"
## [1] "     + CALSBP + bmi + Smoking + Statin + log_Blood_TGA +"
## [1] "     Total_cholesterol + egfr)"
## [1] ""
##                 Name Coefficient   CI.L   CI.R  p.value  p.adj
## 1 L-5-Oxoproline; 63      -0.313 -0.488 -0.138 0.000456 0.0342
## [1] ""
## [1] "Table: log_Blood_TGA"
## [1] "  (from model: "
## [1] "   ~ Charcot.at.DATE + Age.x + Gender.x + Hba1c_baseline"
## [1] "     + CALSBP + bmi + Smoking + Statin + log_Blood_TGA +"
## [1] "     Total_cholesterol + egfr)"
## [1] ""
##                              Name Coefficient    CI.L    CI.R  p.value    p.adj
## 1           Palmitic acid, TMS; 5       0.300  0.1710  0.4290 5.43e-06 0.000248
## 2             Dodecanoic acid; 54       0.293  0.1640  0.4220 9.00e-06 0.000248
## 3            Stearic acid, TMS; 2       0.293  0.1630  0.4220 9.92e-06 0.000248
## 4               Decanoic acid; 52       0.272  0.1430  0.4010 3.62e-05 0.000679
## 5               Octanoic acid; 68       0.269  0.1390  0.3990 5.06e-05 0.000759
## 6              Arachidic acid; 46       0.254  0.1240  0.3840 1.35e-04 0.001680
## 7  2-Hydroxybutyric acid, 2TMS; 2       0.230  0.1020  0.3580 4.21e-04 0.004510
## 8              Oleic acid, TMS; 3       0.224  0.0940  0.3530 7.27e-04 0.006820
## 9             Arabinopyranose; 51       0.206  0.0763  0.3360 1.87e-03 0.015600
## 10         Glyceryl-glycoside; 59       0.199  0.0697  0.3280 2.57e-03 0.019300
## 11           Isoleucine, 2TMS; 18       0.191  0.0637  0.3190 3.32e-03 0.022600
## 12 3,4-Dihydroxybutanoic acid; 27       0.184  0.0574  0.3100 4.37e-03 0.027300
## 13         Heptadecanoic acid; 60       0.184  0.0537  0.3140 5.64e-03 0.031300
## 14               Valine, 2TMS; 20       0.179  0.0517  0.3060 5.84e-03 0.031300
## 15                Lactic acid; 29       0.181  0.0508  0.3110 6.42e-03 0.032100
## 16           Myristoleic acid; 65       0.174  0.0449  0.3030 8.25e-03 0.038700
## 17         Heptadecanoic acid; 61       0.172  0.0415  0.3020 9.77e-03 0.039900
## 18          Glutamic acid, 3TMS; 8       0.168  0.0402  0.2960 1.00e-02 0.039900
## 19               Pyruvic acid; 31       0.171  0.0406  0.3010 1.01e-02 0.039900
## 20               Ribonic acid; 72       0.161  0.0354  0.2860 1.20e-02 0.043400
## 21 4-Hydroxybenzeneacetic acid; 4       0.162  0.0355  0.2890 1.21e-02 0.043400
## 22          Aminomalonic acid; 45      -0.161 -0.2900 -0.0317 1.46e-02 0.049900
```

```
## [1] ""
## [1] "Table: Total_cholesterol"
## [1] "  (from model: "
## [1] "   ~ Charcot.at.DATE + Age.x + Gender.x + Hba1c_baseline"
## [1] "     + CALSBP + bmi + Smoking + Statin + log_Blood_TGA +"
## [1] "     Total_cholesterol + egfr)"
## [1] ""
##                               Name Coefficient    CI.L    CI.R  p.value    p.adj
## 1            Cholesterol, TMS; 23       0.456  0.3630  0.5480 1.67e-21 1.25e-19
## 2                 Campesterol; 49       0.371  0.2770  0.4660 2.23e-14 8.37e-13
## 3            alpha-Tocopherol; 26       0.325  0.2300  0.4200 2.09e-11 5.23e-10
## 4          Benzeneacetic acid; 47      -0.183 -0.2790 -0.0876 1.78e-04 2.94e-03
## 5               Proline, 2TMS; 21      -0.179 -0.2740 -0.0837 2.32e-04 2.94e-03
## 6           Linoleic acid, TMS; 4       0.179  0.0828  0.2740 2.65e-04 2.94e-03
## 7      4-Hydroxybutanoic acid; 43      -0.179 -0.2750 -0.0828 2.74e-04 2.94e-03
## 8            Isoleucine, 2TMS; 18      -0.173 -0.2670 -0.0787 3.27e-04 3.07e-03
## 9               L-5-Oxoproline; 63      -0.171 -0.2670 -0.0750 4.93e-04 4.11e-03
## 10              Glycine, 3TMS; 17      -0.157 -0.2520 -0.0621 1.21e-03 9.06e-03
## 11                   Tyrosine; 75      -0.155 -0.2510 -0.0588 1.60e-03 1.09e-02
## 12           Methionine, 2TMS; 16      -0.152 -0.2470 -0.0565 1.81e-03 1.13e-02
## 13            Threonine, 3TMS; 12      -0.146 -0.2420 -0.0497 2.97e-03 1.71e-02
## 14      Eicosapentaenoic acid; 55       0.141  0.0471  0.2360 3.31e-03 1.77e-02
## 15 2,4-Dihydroxybutanoic acid; 28      -0.137 -0.2300 -0.0450 3.58e-03 1.79e-02
## 16       Docosahexaenoic acid; 53       0.139  0.0445  0.2340 4.00e-03 1.87e-02
## 17            Arabinopyranose; 51      -0.138 -0.2340 -0.0425 4.70e-03 2.07e-02
## 18 4-Hydroxybenzeneacetic acid; 4      -0.134 -0.2280 -0.0403 5.09e-03 2.08e-02
## 19              Alanine, 2TMS; 25      -0.137 -0.2320 -0.0407 5.28e-03 2.08e-02
## 20               Serine, 3TMS; 14      -0.132 -0.2280 -0.0362 6.93e-03 2.60e-02
## 21        2-Palmitoylglycerol; 39       0.132  0.0356  0.2280 7.31e-03 2.61e-02
## 22          Glyceryl-glycoside; 59      -0.127 -0.2230 -0.0319 8.98e-03 3.06e-02
## 23              Hydroxylamine; 62      -0.125 -0.2220 -0.0291 1.08e-02 3.51e-02
## [1] ""
## [1] "Table: egfr"
## [1] "  (from model: "
## [1] "   ~ Charcot.at.DATE + Age.x + Gender.x + Hba1c_baseline"
## [1] "     + CALSBP + bmi + Smoking + Statin + log_Blood_TGA +"
## [1] "     Total_cholesterol + egfr)"
## [1] ""
##                               Name Coefficient      CI.L      CI.R  p.value
## 1            Myo inositol 6TMS; 1    -0.01620 -0.019100 -0.013300 6.96e-27
## 2                     Ribitol; 71    -0.01550 -0.018500 -0.012600 1.07e-24
## 3                  Creatinine; 50    -0.01510 -0.018100 -0.012100 7.54e-23
## 4                Ribonic acid; 72    -0.01460 -0.017500 -0.011600 7.91e-22
## 5  2,4-Dihydroxybutanoic acid; 28    -0.01440 -0.017400 -0.011500 1.43e-21
## 6  3,4-Dihydroxybutanoic acid; 27    -0.01270 -0.015700 -0.009780 5.84e-17
## 7  4-Hydroxybenzeneacetic acid; 4    -0.01130 -0.014300 -0.008320 1.43e-13
## 8        4-Deoxytetronic acid; 33    -0.01130 -0.014300 -0.008300 2.15e-13
## 9        4-Deoxytetronic acid; 32    -0.01020 -0.013200 -0.007200 4.47e-11
## 10           Isoleucine, 2TMS; 18     0.00926  0.006270  0.012200 1.45e-09
## 11 2-Hydroxybutyric acid, 2TMS; 2     0.00925  0.006260  0.012200 1.60e-09
## 12               Valine, 2TMS; 20     0.00832  0.005350  0.011300 4.70e-08
## 13           Citric acid, 4TMS; 6    -0.00796 -0.011000 -0.004960 2.19e-07
## 14          Pyroglutamic acid; 69    -0.00789 -0.010900 -0.004870 3.49e-07
## 15              Hydroxyproline; 64    -0.00774 -0.010800 -0.004700 6.53e-07
```

```
## 16        3-Indoleacetic acid; 40    -0.00691 -0.009930 -0.003890 7.53e-06
## 17         Glyceryl-glycoside; 59    -0.00663 -0.009660 -0.003610 1.81e-05
## 18 4-Hydroxyphenyllactic acid; 44    -0.00635 -0.009380 -0.003320 4.13e-05
## 19               Serine, 3TMS; 14     0.00587  0.002830  0.008900 1.54e-04
## 20              Octanoic acid; 68     0.00583  0.002790  0.008870 1.74e-04
## 21              Leucine, 2TMS; 19     0.00575  0.002740  0.008750 1.83e-04
## 22           Stearic acid, TMS; 2     0.00565  0.002610  0.008680 2.70e-04
## 23           Methionine, 2TMS; 16     0.00526  0.002240  0.008280 6.56e-04
## 24                   Tyrosine; 75     0.00510  0.002060  0.008150 1.03e-03
## 25              Glycine, 3TMS; 17    -0.00481 -0.007830 -0.001800 1.77e-03
## 26  2-hydroxy Isovaleric acid; 38     0.00485  0.001800  0.007890 1.81e-03
## 27      Eicosapentaenoic acid; 55     0.00473  0.001750  0.007720 1.90e-03
## 28          Fumaric acid, 2TMS; 9    -0.00456 -0.007600 -0.001520 3.29e-03
## 29           Cholesterol, TMS; 23     0.00424  0.001300  0.007170 4.66e-03
## 30          Palmitic acid, TMS; 5     0.00417  0.001150  0.007200 6.84e-03
## 31           Malic acid, 3TMS; 11    -0.00406 -0.007100 -0.001020 8.95e-03
## 32              Glyceric acid; 30     0.00379  0.000794  0.006790 1.32e-02
## 33       Docosahexaenoic acid; 53     0.00373  0.000729  0.006730 1.49e-02
## 34                   Glycerol; 57     0.00379  0.000735  0.006850 1.51e-02
## 35              Alanine, 2TMS; 25    -0.00365 -0.006690 -0.000610 1.86e-02
## 36         Benzeneacetic acid; 47    -0.00355 -0.006590 -0.000519 2.17e-02
##       p.adj
## 1  5.22e-25
## 2  4.03e-23
## 3  1.88e-21
## 4  1.48e-20
## 5  2.15e-20
## 6  7.30e-16
## 7  1.54e-12
## 8  2.01e-12
## 9  3.73e-10
## 10 1.09e-08
## 11 1.09e-08
## 12 2.94e-07
## 13 1.27e-06
## 14 1.87e-06
## 15 3.27e-06
## 16 3.53e-05
## 17 7.98e-05
## 18 1.72e-04
## 19 6.07e-04
## 20 6.52e-04
## 21 6.54e-04
## 22 9.19e-04
## 23 2.14e-03
## 24 3.23e-03
## 25 5.22e-03
## 26 5.22e-03
## 27 5.28e-03
## 28 8.82e-03
## 29 1.21e-02
## 30 1.71e-02
## 31 2.17e-02
## 32 3.09e-02
```

```
## 33 3.33e-02
## 34 3.33e-02
## 35 3.99e-02
## 36 4.53e-02
```

###### 4.1.3.3 Table with All Metabolites

```
## [1] ""
## [1] "Table: Charcot.at.DATEJA"
## [1] "  (from model: "
## [1] "   ~ Charcot.at.DATE + Age.x + Gender.x + Hba1c_baseline"
## [1] "     + CALSBP + bmi + Smoking + Statin + log_Blood_TGA +"
## [1] "     Total_cholesterol + egfr)"
## [1] ""
##                              Name Coefficient     CI.L    CI.R  p.value  p.adj
## 1               Proline, 2TMS; 21    -0.66300 -1.05000 -0.2750 0.000826 0.0619
## 2             Threonine, 3TMS; 12    -0.62500 -1.02000 -0.2320 0.001820 0.0684
## 3               Leucine, 2TMS; 19    -0.56000 -0.94800 -0.1730 0.004650 0.1160
## 4  3,4-Dihydroxybutanoic acid; 27     0.51000  0.12900  0.8910 0.008760 0.1460
## 5                Valine, 2TMS; 20    -0.50600 -0.89000 -0.1220 0.009760 0.1460
## 6           Fumaric acid, 2TMS; 9     0.48500  0.09260  0.8770 0.015400 0.1930
## 7                      Ribitol; 70     0.45800  0.06680  0.8500 0.021800 0.2060
## 8  2,4-Dihydroxybutanoic acid; 28     0.44100  0.06380  0.8190 0.022000 0.2060
## 9                Serine, 3TMS; 14    -0.43300 -0.82400 -0.0417 0.030100 0.2510
## 10           Methionine, 2TMS; 16    -0.40900 -0.79900 -0.0191 0.039800 0.2900
## 11           Stearic acid, TMS; 2     0.40500  0.01370  0.7960 0.042500 0.2900
## 12           Cholesterol, TMS; 23     0.38100  0.00281  0.7600 0.048300 0.3020
## 13                    Ribitol; 71     0.36400 -0.01340  0.7420 0.058700 0.3160
## 14               Ribonic acid; 72     0.35800 -0.02120  0.7370 0.064300 0.3160
## 15             Tartronic acid; 73     0.36400 -0.02440  0.7520 0.066200 0.3160
## 16            Arabinopyranose; 51     0.36500 -0.02630  0.7570 0.067500 0.3160
## 17              Glyceric acid; 30     0.34300 -0.04300  0.7300 0.081500 0.3360
## 18            Dodecanoic acid; 54     0.34400 -0.04630  0.7340 0.084000 0.3360
## 19            Myo inositol 6TMS; 1     0.32700 -0.04930  0.7030 0.088500 0.3360
## 20       4-Deoxytetronic acid; 33    -0.33400 -0.72100  0.0518 0.089600 0.3360
## 21           alpha-Tocopherol; 26     0.32500 -0.06110  0.7110 0.098900 0.3520
## 22          Pyroglutamic acid; 69     0.32400 -0.06590  0.7150 0.103000 0.3520
## 23            1,3-Propanediol; 34     0.32000 -0.07410  0.7140 0.111000 0.3630
## 24                 Creatinine; 50     0.30500 -0.07800  0.6880 0.118000 0.3680
## 25         Glyceryl-glycoside; 59     0.30500 -0.08550  0.6950 0.126000 0.3680
## 26      Arachidonic acid, TMS; 24     0.30600 -0.08770  0.7000 0.128000 0.3680
## 27           Isoleucine, 2TMS; 18    -0.29400 -0.67900  0.0916 0.135000 0.3760
## 28                1-Dodecanol; 36     0.29300 -0.10100  0.6880 0.145000 0.3880
## 29           Myristoleic acid; 65     0.28600 -0.10400  0.6750 0.150000 0.3880
## 30              Decanoic acid; 52     0.28100 -0.10900  0.6710 0.157000 0.3910
## 31              Glycine, 3TMS; 17     0.27800 -0.11100  0.6660 0.162000 0.3910
## 32      Eicosapentaenoic acid; 55     0.25800 -0.12700  0.6430 0.190000 0.4440
## 33        Phenylalanine, 2TMS; 13    -0.25200 -0.64600  0.1420 0.210000 0.4730
## 34           Malic acid, 3TMS; 11     0.24800 -0.14400  0.6410 0.215000 0.4730
## 35          Benzeneacetic acid; 47     0.24100 -0.15000  0.6330 0.226000 0.4850
## 36           Citric acid, 4TMS; 6     0.23500 -0.15200  0.6220 0.234000 0.4870
## 37                Campesterol; 49     0.22800 -0.15900  0.6150 0.248000 0.4890
## 38          Palmitic acid, TMS; 5     0.22800 -0.16200  0.6180 0.251000 0.4890
## 39  2-hydroxy Isovaleric acid; 38     0.22700 -0.16600  0.6190 0.258000 0.4890
## 40         Succinic acid, 2TMS; 7     0.22600 -0.16800  0.6190 0.261000 0.4890
## 41                Bisphenol A; 48     0.22000 -0.17500  0.6160 0.275000 0.5020
## 42             Oleic acid, TMS; 3     0.21200 -0.17900  0.6030 0.288000 0.5150
## 43                   Tyrosine; 75    -0.18800 -0.58100  0.2050 0.348000 0.6070
## 44           Tridecanoic acid; 74     0.17600 -0.21700  0.5690 0.379000 0.6340
```

```
## 45              Hydroxylamine; 62    -0.17600 -0.57000  0.2180 0.380000 0.6340
## 46              Alanine, 2TMS; 25    -0.16800 -0.56000  0.2240 0.400000 0.6530
## 47        3-Indoleacetic acid; 40     0.15900 -0.23000  0.5490 0.422000 0.6730
## 48          Aminomalonic acid; 45     0.14900 -0.24100  0.5380 0.455000 0.7050
## 49              Octanoic acid; 68     0.14400 -0.24800  0.5360 0.470000 0.7050
## 50                   Glycerol; 57     0.14500 -0.25000  0.5390 0.472000 0.7050
## 51       Docosahexaenoic acid; 53     0.14000 -0.24800  0.5270 0.480000 0.7050
## 52               Pyruvic acid; 31     0.13300 -0.26000  0.5250 0.507000 0.7180
## 53              Nonanoic acid; 67     0.13300 -0.26100  0.5280 0.508000 0.7180
## 54 4-Hydroxybenzeneacetic acid; 4     0.12500 -0.25800  0.5080 0.523000 0.7180
## 55               Ethanolamine; 56    -0.12700 -0.52100  0.2670 0.526000 0.7180
## 56       4-Deoxytetronic acid; 32     0.11900 -0.27200  0.5090 0.551000 0.7380
## 57 3-Hydroxybutyric acid, 2TMS; 1    -0.11300 -0.50800  0.2830 0.576000 0.7540
## 58     3-Indolepropionic acid; 41    -0.11000 -0.50100  0.2820 0.583000 0.7540
## 59             Arachidic acid; 46     0.10700 -0.28600  0.5000 0.594000 0.7550
## 60 alpha-ketoglutaric acid, TMS M     0.09440 -0.29900  0.4880 0.638000 0.7950
## 61             1-Monopalmitin; 37     0.09240 -0.30300  0.4870 0.646000 0.7950
## 62         11-Eicosenoic acid; 35     0.08740 -0.30600  0.4810 0.663000 0.8020
## 63         Heptadecanoic acid; 60     0.08010 -0.31300  0.4730 0.689000 0.8210
## 64          Nonadecanoic acid; 66     0.07160 -0.32200  0.4650 0.721000 0.8450
## 65                Lactic acid; 29     0.06840 -0.32400  0.4610 0.732000 0.8450
## 66 2-Hydroxybutyric acid, 2TMS; 2     0.05810 -0.32800  0.4440 0.768000 0.8720
## 67          Linoleic acid, TMS; 4     0.04900 -0.34300  0.4410 0.806000 0.9010
## 68     4-Hydroxybutanoic acid; 43    -0.04640 -0.44000  0.3470 0.817000 0.9010
## 69         Heptadecanoic acid; 61     0.04090 -0.35200  0.4340 0.838000 0.9110
## 70                   Glycerol; 58    -0.03390 -0.43000  0.3620 0.867000 0.9280
## 71             Hydroxyproline; 64    -0.02660 -0.41900  0.3650 0.894000 0.9380
## 72 4-Hydroxyphenyllactic acid; 44     0.02480 -0.36600  0.4160 0.901000 0.9380
## 73             L-5-Oxoproline; 63    -0.02190 -0.41500  0.3710 0.913000 0.9380
## 74         2-Palmitoylglycerol; 39    -0.01480 -0.40900  0.3790 0.941000 0.9540
## 75          Glutamic acid, 3TMS; 8    -0.00921 -0.39600  0.3770 0.963000 0.9630
```

###### 4.1.4 Fully-Adjusted Model

```
## [1] "Fitting models:"
## [1] "~ Charcot.at.DATE + Age.x + Gender.x + Hba1c_baseline + CALSBP + bmi + Smoking + Statin + log_B
## [1] ""
```

###### 4.1.4.1 Forest Plot of Model Coefficients

```
## Warning: Ignoring unknown aesthetics: x
## Ignoring unknown aesthetics: x
## Ignoring unknown aesthetics: x
## Ignoring unknown aesthetics: x
## Ignoring unknown aesthetics: x
## Ignoring unknown aesthetics: x
## Ignoring unknown aesthetics: x
## Ignoring unknown aesthetics: x
## Ignoring unknown aesthetics: x
## Ignoring unknown aesthetics: x
```

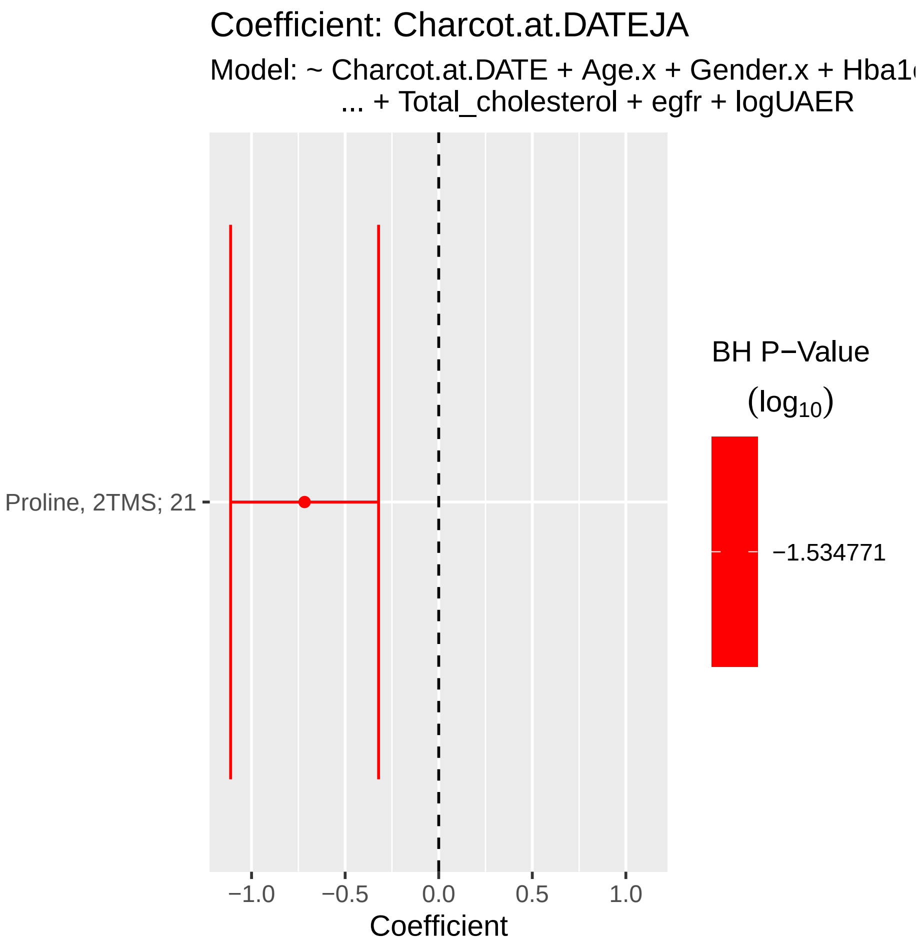

###### 4.1.4.2 Tables of Model Coefficients

```
## [1] ""
## [1] "Table: Charcot.at.DATEJA"
## [1] "  (from model: "
## [1] "   ~ Charcot.at.DATE + Age.x + Gender.x + Hba1c_baseline"
## [1] "     + CALSBP + bmi + Smoking + Statin + log_Blood_TGA +"
## [1] "     Total_cholesterol + egfr + logUAER)"
## [1] ""
##                Name Coefficient  CI.L   CI.R  p.value  p.adj
## 1 Proline, 2TMS; 21      -0.717 -1.11 -0.321 0.000389 0.0292
## [1] ""
## [1] "Table: Age.x"
## [1] "  (from model: "
## [1] "   ~ Charcot.at.DATE + Age.x + Gender.x + Hba1c_baseline"
## [1] "     + CALSBP + bmi + Smoking + Statin + log_Blood_TGA +"
## [1] "     Total_cholesterol + egfr + logUAER)"
## [1] ""
##                             Name Coefficient    CI.L   CI.R  p.value    p.adj
## 1      Eicosapentaenoic acid; 55      0.0252 0.01790 0.0326 2.35e-11 1.76e-09
## 2       Docosahexaenoic acid; 53      0.0137 0.00634 0.0211 2.77e-04 1.04e-02
## 3 alpha-ketoglutaric acid, TMS M      0.0124 0.00491 0.0200 1.22e-03 3.04e-02
## 4               Pyruvic acid; 31      0.0118 0.00427 0.0193 2.15e-03 4.03e-02
## [1] ""
## [1] "Table: Gender.x"
## [1] "  (from model: "
## [1] "   ~ Charcot.at.DATE + Age.x + Gender.x + Hba1c_baseline"
## [1] "     + CALSBP + bmi + Smoking + Statin + log_Blood_TGA +"
## [1] "     Total_cholesterol + egfr + logUAER)"
## [1] ""
##                             Name Coefficient    CI.L    CI.R  p.value    p.adj
## 1       4-Deoxytetronic acid; 33       0.399  0.2350  0.5640 2.01e-06 0.000151
## 2             Tartronic acid; 73      -0.379 -0.5450 -0.2140 7.53e-06 0.000282
## 3              Glyceric acid; 30      -0.358 -0.5220 -0.1940 2.07e-05 0.000490
## 4          Methionine, 2TMS; 16       0.353  0.1870  0.5190 3.17e-05 0.000490
## 5             Proline, 2TMS; 21       0.348  0.1830  0.5140 3.78e-05 0.000490
## 6          Myristoleic acid; 65      -0.350 -0.5160 -0.1830 3.92e-05 0.000490
## 7              Valine, 2TMS; 20       0.327  0.1640  0.4900 8.57e-05 0.000918
## 8          Cholesterol, TMS; 23      -0.295 -0.4560 -0.1350 3.24e-04 0.002810
## 9             Leucine, 2TMS; 19       0.303  0.1370  0.4680 3.37e-04 0.002810
## 10           Oleic acid, TMS; 3      -0.295 -0.4620 -0.1280 5.41e-04 0.004060
## 11         Citric acid, 4TMS; 6      -0.284 -0.4490 -0.1190 7.50e-04 0.004840
## 12          Dodecanoic acid; 54      -0.286 -0.4530 -0.1200 7.75e-04 0.004840
## 13            Glycine, 3TMS; 17      -0.275 -0.4410 -0.1090 1.17e-03 0.006730
## 14         Isoleucine, 2TMS; 18       0.261  0.0965  0.4250 1.87e-03 0.009640
## 15           Hydroxyproline; 64       0.265  0.0977  0.4320 1.93e-03 0.009640
## 16 2-hydroxy Isovaleric acid; 38       0.258  0.0903  0.4250 2.58e-03 0.012100
## 17        Aminomalonic acid; 45      -0.248 -0.4140 -0.0822 3.41e-03 0.015100
## 18     Docosahexaenoic acid; 53      -0.243 -0.4080 -0.0787 3.81e-03 0.015800
## 19            Decanoic acid; 52      -0.245 -0.4110 -0.0783 4.00e-03 0.015800
## 20         Stearic acid, TMS; 2      -0.239 -0.4060 -0.0718 5.11e-03 0.019200
## 21       Glutamic acid, 3TMS; 8       0.224  0.0598  0.3890 7.55e-03 0.027000
## 22       Succinic acid, 2TMS; 7      -0.221 -0.3890 -0.0531 9.90e-03 0.033500
## 23        Palmitic acid, TMS; 5      -0.218 -0.3840 -0.0516 1.03e-02 0.033500
```

```
## 24          Tridecanoic acid; 74      -0.218 -0.3860 -0.0499 1.11e-02 0.034600
## 25         Nonadecanoic acid; 66      -0.205 -0.3730 -0.0375 1.65e-02 0.049600
## [1] ""
## [1] "Table: Hba1c_baseline"
## [1] "  (from model: "
## [1] "   ~ Charcot.at.DATE + Age.x + Gender.x + Hba1c_baseline"
## [1] "     + CALSBP + bmi + Smoking + Statin + log_Blood_TGA +"
## [1] "     Total_cholesterol + egfr + logUAER)"
## [1] ""
##                         Name Coefficient    CI.L    CI.R  p.value   p.adj
## 1          Valine, 2TMS; 20       0.147  0.0740  0.2210 8.46e-05 0.00634
## 2          Ethanolamine; 56       0.137  0.0615  0.2130 3.88e-04 0.01020
## 3         Alanine, 2TMS; 25       0.133  0.0581  0.2090 5.25e-04 0.01020
## 4 Eicosapentaenoic acid; 55      -0.130 -0.2040 -0.0566 5.43e-04 0.01020
## 5       Arabinopyranose; 51       0.118  0.0433  0.1940 2.03e-03 0.03050
## 6  Docosahexaenoic acid; 53      -0.114 -0.1880 -0.0398 2.62e-03 0.03270
## [1] ""
## [1] "Table: CALSBP"
## [1] "  (from model: "
## [1] "   ~ Charcot.at.DATE + Age.x + Gender.x + Hba1c_baseline"
## [1] "     + CALSBP + bmi + Smoking + Statin + log_Blood_TGA +"
## [1] "     Total_cholesterol + egfr + logUAER)"
## [1] ""
## [1] "No significant associations at p.adj < 0.05"
## [1] ""
## [1] "Table: bmi"
## [1] "  (from model: "
## [1] "   ~ Charcot.at.DATE + Age.x + Gender.x + Hba1c_baseline"
## [1] "     + CALSBP + bmi + Smoking + Statin + log_Blood_TGA +"
## [1] "     Total_cholesterol + egfr + logUAER)"
## [1] ""
##                              Name Coefficient    CI.L    CI.R  p.value   p.adj
## 1         Glutamic acid, 3TMS; 8      0.0450  0.0245  0.0656 1.81e-05 0.00136
## 2 2-Hydroxybutyric acid, 2TMS; 2      0.0415  0.0210  0.0620 7.67e-05 0.00259
## 3                Campesterol; 49     -0.0407 -0.0613 -0.0202 1.04e-04 0.00259
## 4              Decanoic acid; 52     -0.0366 -0.0574 -0.0158 5.75e-04 0.01080
## [1] ""
## [1] "Table: Smoking"
## [1] "  (from model: "
## [1] "   ~ Charcot.at.DATE + Age.x + Gender.x + Hba1c_baseline"
## [1] "     + CALSBP + bmi + Smoking + Statin + log_Blood_TGA +"
## [1] "     Total_cholesterol + egfr + logUAER)"
## [1] ""
##                         Name Coefficient   CI.L   CI.R  p.value  p.adj
## 1 3-Indolepropionic acid; 41      -0.351 -0.550 -0.153 0.000538 0.0156
## 2         Tartronic acid; 73      -0.341 -0.538 -0.145 0.000684 0.0156
## 3   Docosahexaenoic acid; 53      -0.339 -0.535 -0.143 0.000700 0.0156
## 4     Glutamic acid, 3TMS; 8       0.334  0.138  0.529 0.000831 0.0156
## 5           Valine, 2TMS; 20      -0.301 -0.495 -0.107 0.002350 0.0302
## 6          Glyceric acid; 30      -0.299 -0.494 -0.103 0.002750 0.0302
## 7       alpha-Tocopherol; 26      -0.298 -0.494 -0.103 0.002820 0.0302
## [1] ""
## [1] "Table: Statin"
## [1] "  (from model: "
```

```
## [1] "   ~ Charcot.at.DATE + Age.x + Gender.x + Hba1c_baseline"
## [1] "     + CALSBP + bmi + Smoking + Statin + log_Blood_TGA +"
## [1] "     Total_cholesterol + egfr + logUAER)"
## [1] ""
## [1] "No significant associations at p.adj < 0.05"
## [1] ""
## [1] "Table: log_Blood_TGA"
## [1] "  (from model: "
## [1] "   ~ Charcot.at.DATE + Age.x + Gender.x + Hba1c_baseline"
## [1] "     + CALSBP + bmi + Smoking + Statin + log_Blood_TGA +"
## [1] "     Total_cholesterol + egfr + logUAER)"
## [1] ""
##                               Name Coefficient   CI.L  CI.R  p.value   p.adj
## 1          Palmitic acid, TMS; 5       0.293 0.1610 0.426 1.52e-05 0.00110
## 2            Dodecanoic acid; 54       0.277 0.1450 0.410 4.43e-05 0.00110
## 3           Stearic acid, TMS; 2       0.278 0.1450 0.411 4.46e-05 0.00110
## 4              Octanoic acid; 68       0.273 0.1400 0.406 5.88e-05 0.00110
## 5             Arachidic acid; 46       0.267 0.1330 0.401 9.37e-05 0.00140
## 6              Decanoic acid; 52       0.252 0.1190 0.385 2.01e-04 0.00252
## 7  2-Hydroxybutyric acid, 2TMS; 2       0.244 0.1130 0.375 2.58e-04 0.00276
## 8             Oleic acid, TMS; 3       0.224 0.0913 0.357 9.65e-04 0.00905
## 9                Lactic acid; 29       0.205 0.0713 0.338 2.65e-03 0.02210
## 10         Glyceryl-glycoside; 59       0.200 0.0673 0.333 3.15e-03 0.02360
## 11           Arabinopyranose; 51       0.188 0.0548 0.321 5.69e-03 0.03880
## 12 3,4-Dihydroxybutanoic acid; 27       0.174 0.0460 0.303 7.80e-03 0.04240
## 13         Heptadecanoic acid; 60       0.182 0.0478 0.315 7.84e-03 0.04240
## 14               Ribonic acid; 72       0.172 0.0439 0.300 8.51e-03 0.04240
## 15           Myristoleic acid; 65       0.177 0.0444 0.309 8.91e-03 0.04240
## 16           Isoleucine, 2TMS; 18       0.174 0.0434 0.305 9.05e-03 0.04240
## 17 4-Hydroxybenzeneacetic acid; 4       0.171 0.0411 0.301 9.95e-03 0.04390
## 18          Fumaric acid, 2TMS; 9       0.173 0.0401 0.306 1.08e-02 0.04490
## [1] ""
## [1] "Table: Total_cholesterol"
## [1] "  (from model: "
## [1] "   ~ Charcot.at.DATE + Age.x + Gender.x + Hba1c_baseline"
## [1] "     + CALSBP + bmi + Smoking + Statin + log_Blood_TGA +"
## [1] "     Total_cholesterol + egfr + logUAER)"
## [1] ""
##                               Name Coefficient    CI.L    CI.R  p.value    p.adj
## 1           Cholesterol, TMS; 23       0.456  0.3620  0.5510 7.87e-21 5.90e-19
## 2                Campesterol; 49       0.380  0.2830  0.4760 1.87e-14 7.01e-13
## 3           alpha-Tocopherol; 26       0.329  0.2320  0.4250 3.10e-11 7.75e-10
## 4         Benzeneacetic acid; 47      -0.185 -0.2830 -0.0872 2.16e-04 4.05e-03
## 5          Linoleic acid, TMS; 4       0.182  0.0843  0.2800 2.72e-04 4.09e-03
## 6           Isoleucine, 2TMS; 18      -0.170 -0.2660 -0.0734 5.56e-04 5.58e-03
## 7      4-Hydroxybutanoic acid; 43      -0.174 -0.2720 -0.0751 5.65e-04 5.58e-03
## 8              Proline, 2TMS; 21      -0.170 -0.2670 -0.0731 5.95e-04 5.58e-03
## 9             L-5-Oxoproline; 63      -0.168 -0.2660 -0.0694 8.43e-04 7.03e-03
## 10     Eicosapentaenoic acid; 55       0.158  0.0615  0.2540 1.32e-03 9.59e-03
## 11          Methionine, 2TMS; 16      -0.158 -0.2560 -0.0611 1.43e-03 9.59e-03
## 12                  Tyrosine; 75      -0.159 -0.2570 -0.0607 1.53e-03 9.59e-03
## 13           Threonine, 3TMS; 12      -0.153 -0.2510 -0.0542 2.38e-03 1.32e-02
## 14       Docosahexaenoic acid; 53       0.149  0.0527  0.2460 2.47e-03 1.32e-02
## 15             Glycine, 3TMS; 17      -0.146 -0.2430 -0.0485 3.33e-03 1.67e-02
```

```
## 16             Alanine, 2TMS; 25      -0.142 -0.2400 -0.0441 4.52e-03 2.12e-02
## 17 4-Hydroxybenzeneacetic acid; 4      -0.129 -0.2250 -0.0334 8.21e-03 3.62e-02
## 18        Glyceryl-glycoside; 59      -0.128 -0.2250 -0.0303 1.03e-02 4.08e-02
## 19           Arabinopyranose; 51      -0.127 -0.2250 -0.0291 1.10e-02 4.08e-02
## 20              Serine, 3TMS; 14      -0.127 -0.2250 -0.0290 1.11e-02 4.08e-02
## 21             Hydroxylamine; 62      -0.127 -0.2260 -0.0287 1.14e-02 4.08e-02
## 22          Malic acid, 3TMS; 11      -0.125 -0.2230 -0.0271 1.24e-02 4.10e-02
## 23       2-Palmitoylglycerol; 39       0.126  0.0270  0.2240 1.26e-02 4.10e-02
## 24 2,4-Dihydroxybutanoic acid; 28      -0.116 -0.2090 -0.0219 1.56e-02 4.88e-02
## [1] ""
## [1] "Table: egfr"
## [1] "  (from model: "
## [1] "   ~ Charcot.at.DATE + Age.x + Gender.x + Hba1c_baseline"
## [1] "     + CALSBP + bmi + Smoking + Statin + log_Blood_TGA +"
## [1] "     Total_cholesterol + egfr + logUAER)"
## [1] ""
##                               Name Coefficient      CI.L     CI.R  p.value
## 1           Myo inositol 6TMS; 1    -0.01600 -0.019200 -0.01290 9.69e-23
## 2                    Ribitol; 71    -0.01500 -0.018100 -0.01180 6.87e-20
## 3                 Creatinine; 50    -0.01490 -0.018100 -0.01160 4.67e-19
## 4  2,4-Dihydroxybutanoic acid; 28    -0.01440 -0.017600 -0.01120 1.32e-18
## 5               Ribonic acid; 72    -0.01330 -0.016500 -0.01010 4.81e-16
## 6       4-Deoxytetronic acid; 33    -0.01140 -0.014700 -0.00817 8.43e-12
## 7  3,4-Dihydroxybutanoic acid; 27    -0.01100 -0.014200 -0.00779 2.15e-11
## 8  4-Hydroxybenzeneacetic acid; 4    -0.01110 -0.014300 -0.00785 2.54e-11
## 9           Isoleucine, 2TMS; 18     0.00951  0.006260  0.01280 1.12e-08
## 10      4-Deoxytetronic acid; 32    -0.00895 -0.012200 -0.00567 1.02e-07
## 11          Citric acid, 4TMS; 6    -0.00863 -0.011900 -0.00536 2.57e-07
## 12         Pyroglutamic acid; 69    -0.00869 -0.012000 -0.00539 2.66e-07
## 13 2-Hydroxybutyric acid, 2TMS; 2     0.00802  0.004770  0.01130 1.43e-06
## 14 4-Hydroxyphenyllactic acid; 44    -0.00776 -0.011100 -0.00446 4.30e-06
## 15       3-Indoleacetic acid; 40    -0.00739 -0.010700 -0.00410 1.13e-05
## 16              Valine, 2TMS; 20     0.00720  0.003970  0.01040 1.31e-05
## 17            Hydroxyproline; 64    -0.00651 -0.009820 -0.00319 1.22e-04
## 18        Glyceryl-glycoside; 59    -0.00628 -0.009580 -0.00298 1.94e-04
## 19             Leucine, 2TMS; 19     0.00584  0.002570  0.00912 4.75e-04
## 20              Serine, 3TMS; 14     0.00591  0.002600  0.00922 4.76e-04
## 21             Glycine, 3TMS; 17    -0.00571 -0.009000 -0.00243 6.65e-04
## 22         Fumaric acid, 2TMS; 9    -0.00550 -0.008810 -0.00219 1.14e-03
## 23     Eicosapentaenoic acid; 55     0.00527  0.002020  0.00851 1.49e-03
## 24          Stearic acid, TMS; 2     0.00530  0.001990  0.00861 1.73e-03
## 25          Methionine, 2TMS; 16     0.00467  0.001390  0.00796 5.37e-03
## 26         Palmitic acid, TMS; 5     0.00401  0.000715  0.00731 1.71e-02
##       p.adj
## 1  7.27e-21
## 2  2.57e-18
## 3  1.17e-17
## 4  2.47e-17
## 5  7.21e-15
## 6  1.05e-10
## 7  2.31e-10
## 8  2.38e-10
## 9  9.35e-08
## 10 7.64e-07
```

```
## 11 1.66e-06
## 12 1.66e-06
## 13 8.25e-06
## 14 2.30e-05
## 15 5.67e-05
## 16 6.12e-05
## 17 5.38e-04
## 18 8.09e-04
## 19 1.79e-03
## 20 1.79e-03
## 21 2.37e-03
## 22 3.90e-03
## 23 4.86e-03
## 24 5.42e-03
## 25 1.61e-02
## 26 4.94e-02
## [1] ""
## [1] "Table: logUAER"
## [1] "  (from model: "
## [1] "   ~ Charcot.at.DATE + Age.x + Gender.x + Hba1c_baseline"
## [1] "     + CALSBP + bmi + Smoking + Statin + log_Blood_TGA +"
## [1] "     Total_cholesterol + egfr + logUAER)"
## [1] ""
##                              Name Coefficient   CI.L  CI.R p.value  p.adj
## 1 3,4-Dihydroxybutanoic acid; 27      0.0714 0.0316 0.111 0.00045 0.0338
```

###### 4.1.4.3 Table with All Metabolites

```
## [1] ""
## [1] "Table: Charcot.at.DATEJA"
## [1] "  (from model: "
## [1] "   ~ Charcot.at.DATE + Age.x + Gender.x + Hba1c_baseline"
## [1] "     + CALSBP + bmi + Smoking + Statin + log_Blood_TGA +"
## [1] "     Total_cholesterol + egfr + logUAER)"
## [1] ""
##                               Name Coefficient    CI.L     CI.R  p.value  p.adj
## 1               Proline, 2TMS; 21     -0.7170 -1.1100 -0.32100 0.000389 0.0292
## 2             Threonine, 3TMS; 12     -0.6170 -1.0200 -0.21600 0.002580 0.0638
## 3                Valine, 2TMS; 20     -0.5870 -0.9760 -0.19700 0.003190 0.0638
## 4               Leucine, 2TMS; 19     -0.5910 -0.9860 -0.19600 0.003400 0.0638
## 5  3,4-Dihydroxybutanoic acid; 27      0.5210  0.1360  0.90600 0.008090 0.1210
## 6             Malic acid, 3TMS; 11      0.4770  0.0782  0.87600 0.019100 0.1790
## 7             Cholesterol, TMS; 23      0.4590  0.0750  0.84400 0.019200 0.1790
## 8            Fumaric acid, 2TMS; 9      0.4770  0.0776  0.87600 0.019300 0.1790
## 9                Serine, 3TMS; 14     -0.4690 -0.8680 -0.06910 0.021500 0.1790
## 10 2,4-Dihydroxybutanoic acid; 28      0.4390  0.0571  0.82200 0.024300 0.1820
## 11                     Ribitol; 70      0.4430  0.0435  0.84300 0.029800 0.1900
## 12           Stearic acid, TMS; 2      0.4410  0.0418  0.84100 0.030400 0.1900
## 13            Methionine, 2TMS; 16     -0.3920 -0.7890  0.00478 0.052800 0.2900
## 14             Arabinopyranose; 51      0.3870 -0.0122  0.78700 0.057400 0.2900
## 15                Ribonic acid; 72      0.3640 -0.0198  0.74800 0.063100 0.2900
## 16                     Ribitol; 71      0.3600 -0.0224  0.74200 0.065000 0.2900
## 17             Dodecanoic acid; 54      0.3740 -0.0244  0.77300 0.065800 0.2900
## 18               Glyceric acid; 30      0.3620 -0.0316  0.75500 0.071500 0.2920
## 19       Arachidonic acid, TMS; 24      0.3610 -0.0400  0.76300 0.077600 0.2920
## 20            Myo inositol 6TMS; 1      0.3410 -0.0393  0.72200 0.078800 0.2920
## 21          Succinic acid, 2TMS; 7      0.3560 -0.0454  0.75700 0.082100 0.2920
## 22              Tartronic acid; 73      0.3470 -0.0492  0.74300 0.086100 0.2920
## 23             1,3-Propanediol; 34      0.3490 -0.0538  0.75100 0.089500 0.2920
## 24            Pyroglutamic acid; 69     0.3320 -0.0665  0.73000 0.102000 0.3120
## 25            Isoleucine, 2TMS; 18     -0.3250 -0.7170  0.06680 0.104000 0.3120
## 26            alpha-Tocopherol; 26      0.3130 -0.0800  0.70600 0.118000 0.3240
## 27            Myristoleic acid; 65      0.3160 -0.0812  0.71400 0.119000 0.3240
## 28                  Creatinine; 50      0.3080 -0.0814  0.69700 0.121000 0.3240
## 29               Decanoic acid; 52      0.3020 -0.0959  0.70100 0.137000 0.3530
## 30        4-Deoxytetronic acid; 33     -0.2820 -0.6750  0.11000 0.159000 0.3900
## 31              Oleic acid, TMS; 3      0.2860 -0.1140  0.68500 0.161000 0.3900
## 32           Palmitic acid, TMS; 5      0.2760 -0.1220  0.67300 0.174000 0.3910
## 33         Phenylalanine, 2TMS; 13     -0.2740 -0.6760  0.12900 0.183000 0.3910
## 34                1-Dodecanol; 36       0.2740 -0.1290  0.67700 0.183000 0.3910
## 35                 Campesterol; 49      0.2650 -0.1280  0.65700 0.186000 0.3910
## 36               Glycine, 3TMS; 17      0.2660 -0.1300  0.66300 0.188000 0.3910
## 37          Glyceryl-glycoside; 59      0.2470 -0.1510  0.64500 0.224000 0.4540
## 38               Nonanoic acid; 67      0.2440 -0.1590  0.64700 0.236000 0.4540
## 39            Citric acid, 4TMS; 6      0.2380 -0.1560  0.63300 0.236000 0.4540
## 40  2-hydroxy Isovaleric acid; 38      0.2370 -0.1630  0.63800 0.245000 0.4600
## 41                    Tyrosine; 75     -0.2180 -0.6180  0.18300 0.286000 0.5240
## 42          Benzeneacetic acid; 47      0.2040 -0.1950  0.60400 0.316000 0.5640
## 43      Eicosapentaenoic acid; 55       0.1960 -0.1960  0.58800 0.326000 0.5680
## 44                 Lactic acid; 29      0.1890 -0.2110  0.58900 0.353000 0.6020
```

```
## 45          Tridecanoic acid; 74      0.1780 -0.2240  0.57900 0.385000 0.6260
## 46                  Glycerol; 57      0.1780 -0.2250  0.58100 0.387000 0.6260
## 47             Alanine, 2TMS; 25     -0.1740 -0.5740  0.22500 0.392000 0.6260
## 48             Hydroxylamine; 62     -0.1630 -0.5650  0.24000 0.428000 0.6580
## 49              Pyruvic acid; 31      0.1610 -0.2400  0.56200 0.432000 0.6580
## 50             Octanoic acid; 68      0.1580 -0.2420  0.55700 0.439000 0.6580
## 51 alpha-ketoglutaric acid, TMS M      0.1560 -0.2460  0.55700 0.447000 0.6580
## 52               Bisphenol A; 48      0.1520 -0.2520  0.55600 0.461000 0.6650
## 53           Aminomalonic acid; 45      0.1450 -0.2520  0.54200 0.474000 0.6660
## 54 2-Hydroxybutyric acid, 2TMS; 2      0.1410 -0.2510  0.53400 0.480000 0.6660
## 55       Docosahexaenoic acid; 53      0.1380 -0.2560  0.53200 0.492000 0.6700
## 56        3-Indoleacetic acid; 40      0.1360 -0.2610  0.53300 0.500000 0.6700
## 57           Linoleic acid, TMS; 4      0.1150 -0.2850  0.51500 0.572000 0.7420
## 58 3-Hydroxybutyric acid, 2TMS; 1     -0.1160 -0.5200  0.28800 0.574000 0.7420
## 59     3-Indolepropionic acid; 41     -0.1040 -0.5030  0.29600 0.610000 0.7760
## 60              Arachidic acid; 46      0.0964 -0.3050  0.49800 0.638000 0.7880
## 61 4-Hydroxybenzeneacetic acid; 4      0.0929 -0.2970  0.48300 0.641000 0.7880
## 62               Ethanolamine; 56     -0.0907 -0.4930  0.31100 0.658000 0.7950
## 63             1-Monopalmitin; 37      0.0884 -0.3150  0.49200 0.667000 0.7950
## 64       4-Deoxytetronic acid; 32      0.0766 -0.3190  0.47300 0.704000 0.8250
## 65          Heptadecanoic acid; 61     -0.0669 -0.4690  0.33500 0.744000 0.8550
## 66         11-Eicosenoic acid; 35      0.0612 -0.3410  0.46400 0.766000 0.8550
## 67          Nonadecanoic acid; 66      0.0601 -0.3410  0.46200 0.769000 0.8550
## 68 4-Hydroxyphenyllactic acid; 44      0.0581 -0.3400  0.45600 0.775000 0.8550
## 69              L-5-Oxoproline; 63      0.0508 -0.3500  0.45200 0.804000 0.8740
## 70                  Glycerol; 58      0.0388 -0.3650  0.44300 0.851000 0.9110
## 71              Hydroxyproline; 64     -0.0343 -0.4340  0.36600 0.866000 0.9130
## 72        2-Palmitoylglycerol; 39     -0.0290 -0.4310  0.37300 0.888000 0.9130
## 73          Heptadecanoic acid; 60      0.0287 -0.3730  0.43000 0.888000 0.9130
## 74          Glutamic acid, 3TMS; 8     -0.0223 -0.4150  0.37100 0.911000 0.9200
## 75     4-Hydroxybutanoic acid; 43     -0.0207 -0.4230  0.38200 0.920000 0.9200
```

##### 4.1.5 Compilation

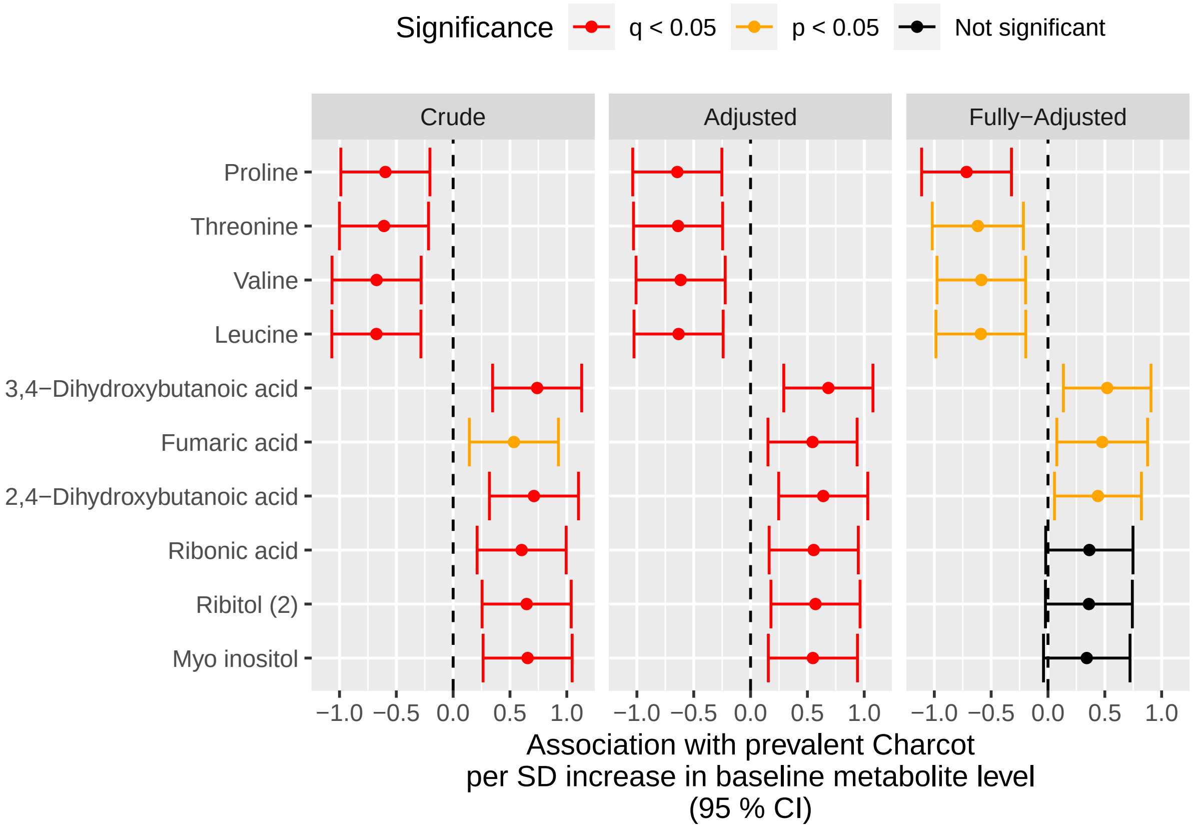

#### 4.2 Charcot from DATE

##### 4.2.1 Crude Model

###### 4.2.1.1 Forest Plot of Model Coefficients

```
## Warning: Ignoring unknown aesthetics: x
```

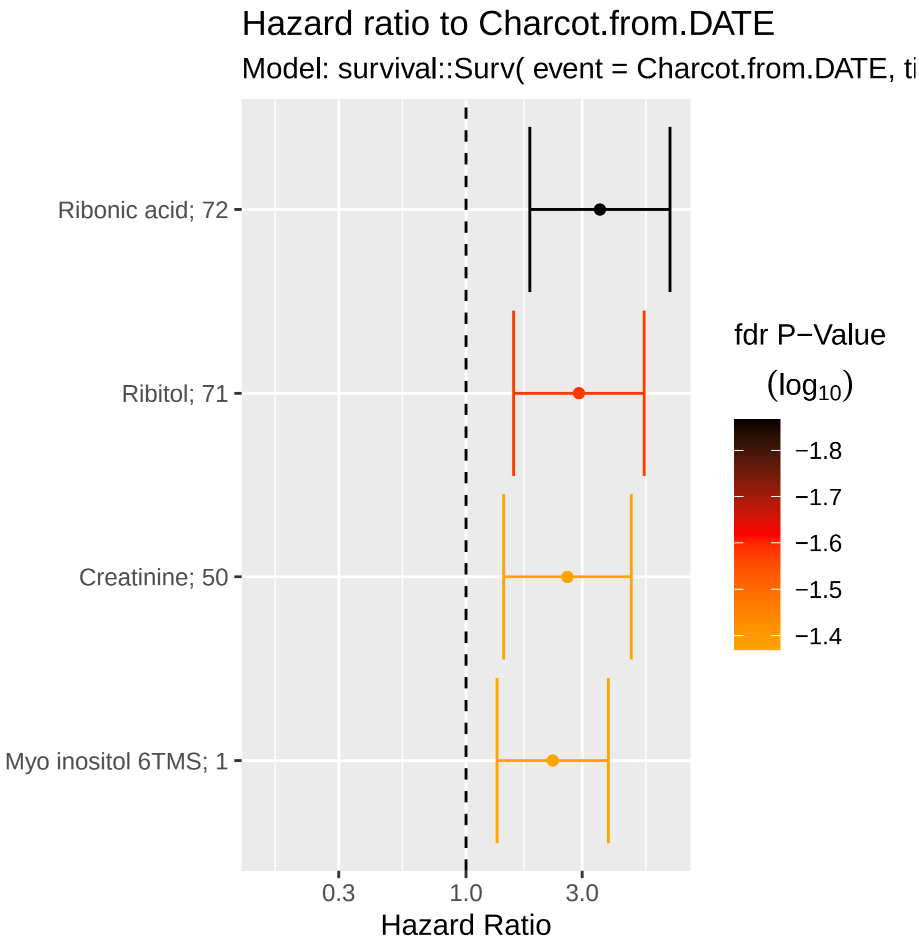

###### 4.2.1.2 Table with All Metabolites

| Name | exp(coef) | Lower 95 % | Upper 95 % | Pr(>\|z\|) | p.adj |
|---|---|---|---|---|---|
| Ribonic acid | 3.55 | 1.83 | 6.89 | 0.000182 | 0.0136 |
| Ribitol (2) | 2.91 | 1.57 | 5.4 | 0.000706 | 0.0265 |
| Creatinine | 2.61 | 1.43 | 4.77 | 0.00184 | 0.0427 |
| Myo inositol | 2.27 | 1.34 | 3.84 | 0.00228 | 0.0427 |
| Tyrosine | 0.698 | 0.545 | 0.895 | 0.00455 | 0.0655 |
| 2,4-Dihydroxybutanoic acid | 2.18 | 1.26 | 3.77 | 0.00524 | 0.0655 |
| Malic acid | 2.06 | 1.2 | 3.54 | 0.00868 | 0.093 |
| 3,4-Dihydroxybutanoic acid | 2.05 | 1.18 | 3.56 | 0.0106 | 0.099 |
| Citric acid | 2.1 | 1.18 | 3.76 | 0.012 | 0.1 |
| 4-Hydroxybenzeneacetic acid | 2.2 | 1.17 | 4.14 | 0.0146 | 0.109 |
| Valine | 0.597 | 0.389 | 0.918 | 0.0187 | 0.128 |
| Tartronic acid | 2.28 | 1.13 | 4.62 | 0.022 | 0.137 |
| Benzeneacetic acid | 2.15 | 1.07 | 4.34 | 0.0324 | 0.177 |
| Leucine | 0.729 | 0.546 | 0.975 | 0.0333 | 0.177 |
| Hydroxylamine | 1.92 | 1.05 | 3.54 | 0.0354 | 0.177 |
| Isoleucine | 0.727 | 0.531 | 0.996 | 0.0473 | 0.213 |
| 4-Deoxytetronic acid (1) | 1.82 | 1 | 3.29 | 0.0484 | 0.213 |
| Ethanolamine | 0.599 | 0.356 | 1.01 | 0.0536 | 0.223 |
| 2-hydroxy Isovaleric acid | 0.662 | 0.423 | 1.03 | 0.0702 | 0.277 |
| Methionine | 0.696 | 0.453 | 1.07 | 0.0982 | 0.368 |
| Serine | 0.696 | 0.443 | 1.09 | 0.115 | 0.411 |
| Pyruvic acid | 0.697 | 0.436 | 1.12 | 0.132 | 0.436 |
| Glycerol (2) | 1.57 | 0.87 | 2.84 | 0.134 | 0.436 |
| Linoleic acid | 1.55 | 0.832 | 2.87 | 0.168 | 0.526 |
| Octanoic acid | 0.815 | 0.6 | 1.11 | 0.191 | 0.532 |
| Fumaric acid | 1.44 | 0.824 | 2.53 | 0.2 | 0.532 |
| Glyceric acid | 1.47 | 0.796 | 2.73 | 0.217 | 0.532 |
| Stearic acid | 1.37 | 0.83 | 2.27 | 0.218 | 0.532 |
| 4-Deoxytetronic acid (2) | 1.49 | 0.789 | 2.81 | 0.22 | 0.532 |
| L-5-Oxoproline | 1.45 | 0.797 | 2.64 | 0.224 | 0.532 |
| Glycerol (1) | 1.59 | 0.744 | 3.38 | 0.232 | 0.532 |
| Nonanoic acid | 1.42 | 0.796 | 2.55 | 0.234 | 0.532 |
| 4-Hydroxybutanoic acid | 1.4 | 0.799 | 2.44 | 0.241 | 0.532 |
| Alanine | 0.749 | 0.463 | 1.21 | 0.241 | 0.532 |
| Glyceryl-glycoside | 1.48 | 0.732 | 3 | 0.274 | 0.557 |
| 4-Hydroxyphenyllactic acid | 1.44 | 0.749 | 2.76 | 0.275 | 0.557 |
| Proline | 0.76 | 0.462 | 1.25 | 0.28 | 0.557 |
| alpha-ketoglutaric acid | 0.837 | 0.602 | 1.16 | 0.289 | 0.557 |
| Arabinopyranose | 1.45 | 0.72 | 2.9 | 0.3 | 0.557 |
| Succinic acid | 1.34 | 0.768 | 2.34 | 0.303 | 0.557 |
| Pyroglutamic acid | 1.44 | 0.72 | 2.87 | 0.305 | 0.557 |
| Tridecanoic acid | 1.41 | 0.72 | 2.76 | 0.317 | 0.566 |
| 11-Eicosenoic acid | 1.38 | 0.722 | 2.63 | 0.33 | 0.575 |
| 3-Hydroxybutyric acid | 0.767 | 0.443 | 1.33 | 0.345 | 0.587 |
| Glutamic acid | 0.802 | 0.5 | 1.28 | 0.358 | 0.597 |
| Threonine | 0.845 | 0.582 | 1.23 | 0.378 | 0.615 |
| Docosahexaenoic acid | 1.33 | 0.697 | 2.55 | 0.386 | 0.615 |
| Lactic acid | 0.818 | 0.506 | 1.32 | 0.411 | 0.623 |
| Glycine | 1.25 | 0.729 | 2.14 | 0.418 | 0.623 |
| 2-Hydroxybutyric acid | 0.819 | 0.496 | 1.35 | 0.437 | 0.623 |

| Name | exp(coef) | Lower 95 % | Upper 95 % | Pr(>\|z\|) | p.adj |
|---|---|---|---|---|---|
| 3-Indoleacetic acid | 0.825 | 0.503 | 1.35 | 0.447 | 0.623 |
| 3-Indolepropionic acid | 0.86 | 0.574 | 1.29 | 0.464 | 0.623 |
| 1,3-Propanediol | 1.27 | 0.661 | 2.46 | 0.469 | 0.623 |
| Arachidonic acid | 1.25 | 0.684 | 2.28 | 0.471 | 0.623 |
| Nonadecanoic acid | 1.29 | 0.641 | 2.61 | 0.474 | 0.623 |
| Arachidic acid | 1.2 | 0.729 | 1.96 | 0.478 | 0.623 |
| Ribitol (1) | 1.22 | 0.703 | 2.11 | 0.481 | 0.623 |
| Decanoic acid | 0.842 | 0.521 | 1.36 | 0.482 | 0.623 |
| Eicosapentaenoic acid | 0.846 | 0.51 | 1.4 | 0.515 | 0.655 |
| Hydroxyproline | 1.19 | 0.677 | 2.09 | 0.546 | 0.682 |
| Myristoleic acid | 1.13 | 0.64 | 1.99 | 0.675 | 0.823 |
| alpha-Tocopherol | 1.13 | 0.623 | 2.07 | 0.681 | 0.823 |
| Bisphenol A | 1.1 | 0.641 | 1.9 | 0.724 | 0.862 |
| Oleic acid | 1.08 | 0.632 | 1.86 | 0.769 | 0.901 |
| Aminomalonic acid | 1.07 | 0.617 | 1.87 | 0.802 | 0.925 |
| Cholesterol | 0.939 | 0.547 | 1.61 | 0.82 | 0.927 |
| Heptadecanoic acid (2) | 1.06 | 0.606 | 1.86 | 0.835 | 0.927 |
| Phenylalanine | 0.946 | 0.554 | 1.62 | 0.84 | 0.927 |
| 1-Monopalmitin | 1.05 | 0.605 | 1.83 | 0.854 | 0.928 |
| Palmitic acid | 1.04 | 0.601 | 1.81 | 0.88 | 0.939 |
| 2-Palmitoylglycerol | 0.963 | 0.563 | 1.65 | 0.889 | 0.939 |
| Dodecanoic acid | 1.03 | 0.601 | 1.77 | 0.913 | 0.951 |
| Campesterol | 1.02 | 0.598 | 1.75 | 0.932 | 0.957 |
| 1-Dodecanol | 0.994 | 0.573 | 1.73 | 0.984 | 0.99 |
| Heptadecanoic acid (1) | 0.997 | 0.573 | 1.73 | 0.99 | 0.99 |

###### 4.2.1.3 Top-Metabolite

```
## Call:
## survival::coxph(formula = survival::Surv(time = Charcot.tdiff,
##     event = Charcot.from.DATE) ~ Ribonic_acid, data = data.km)
##
##   n= 611, number of events= 13
##    (26 observations deleted due to missingness)
##
##               coef exp(coef) se(coef)     z Pr(>|z|)
## Ribonic_acid 1.288     3.625    0.344 3.743 0.000182 ***
## ---
## Signif. codes:  0 '***' 0.001 '**' 0.01 '*' 0.05 '.' 0.1 ' ' 1
##
##              exp(coef) exp(-coef) lower .95 upper .95
## Ribonic_acid     3.625     0.2759     1.847     7.114
##
## Concordance= 0.833  (se = 0.036 )
## Likelihood ratio test= 15.28  on 1 df,   p=9e-05
## Wald test            = 14.01  on 1 df,   p=2e-04
## Score (logrank) test = 12.37  on 1 df,   p=4e-04
```

###### 4.2.1.4 Kaplan-Maier Curve with Median Cutpoint

- Top metabolite

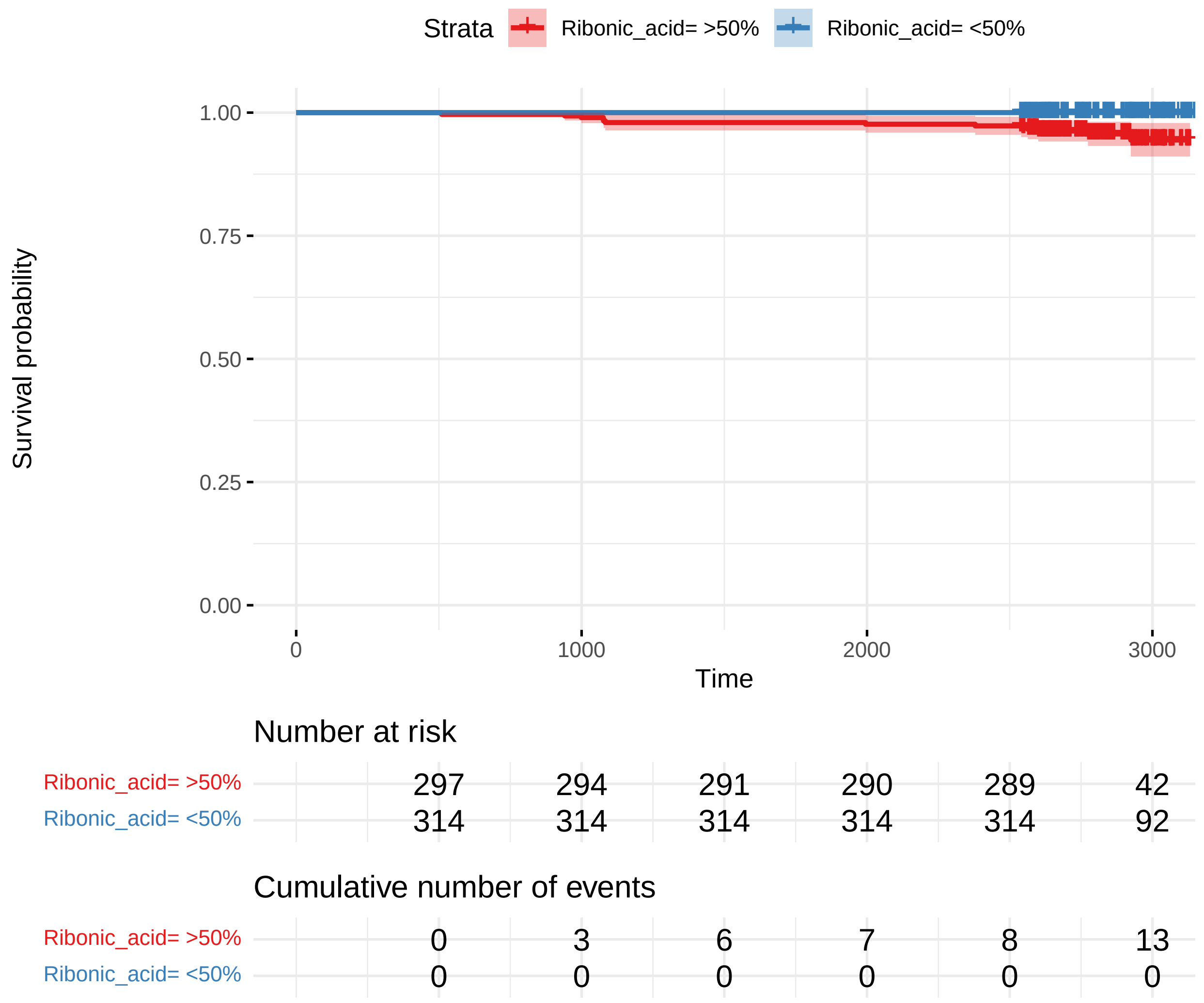

##### 4.2.2 Adusted Model

###### 4.2.2.1 Forest Plot of Model Coefficients

```
## NULL
```

###### 4.2.2.2 Table with All Metabolites

| Name | exp(coef) | Lower 95 % | Upper 95 % | Pr(>\|z\|) | p.adj |
|---|---|---|---|---|---|
| Ribonic acid | 3.13 | 1.58 | 6.19 | 0.00106 | 0.0796 |
| Ribitol (2) | 2.57 | 1.34 | 4.91 | 0.00435 | 0.128 |
| Creatinine | 2.38 | 1.3 | 4.38 | 0.00511 | 0.128 |
| Valine | 0.533 | 0.331 | 0.857 | 0.0094 | 0.128 |
| Tyrosine | 0.711 | 0.548 | 0.923 | 0.0103 | 0.128 |
| Myo inositol | 2.05 | 1.17 | 3.61 | 0.0126 | 0.128 |
| 2,4-Dihydroxybutanoic acid | 2.11 | 1.17 | 3.8 | 0.0128 | 0.128 |
| Citric acid | 2.19 | 1.17 | 4.11 | 0.0149 | 0.128 |
| Tartronic acid | 2.31 | 1.17 | 4.56 | 0.0153 | 0.128 |
| Hydroxylamine | 2.15 | 1.11 | 4.16 | 0.0226 | 0.155 |
| Isoleucine | 0.694 | 0.506 | 0.95 | 0.0228 | 0.155 |
| 3,4-Dihydroxybutanoic acid | 1.92 | 1.06 | 3.48 | 0.0313 | 0.196 |
| Leucine | 0.733 | 0.55 | 0.979 | 0.0351 | 0.202 |
| Benzeneacetic acid | 2.23 | 1.04 | 4.8 | 0.0403 | 0.202 |
| Ethanolamine | 0.56 | 0.322 | 0.975 | 0.0404 | 0.202 |
| 4-Hydroxybenzeneacetic acid | 1.99 | 1.01 | 3.94 | 0.047 | 0.215 |
| Malic acid | 1.78 | 0.999 | 3.17 | 0.0504 | 0.215 |
| Pyruvic acid | 0.604 | 0.363 | 1 | 0.0518 | 0.215 |
| 4-Deoxytetronic acid (1) | 1.83 | 0.989 | 3.38 | 0.0544 | 0.215 |
| 2-hydroxy Isovaleric acid | 0.65 | 0.411 | 1.03 | 0.0664 | 0.249 |
| Methionine | 0.642 | 0.383 | 1.07 | 0.0918 | 0.328 |
| Glycerol (2) | 1.72 | 0.905 | 3.28 | 0.0977 | 0.333 |
| Octanoic acid | 0.687 | 0.435 | 1.09 | 0.108 | 0.352 |
| Alanine | 0.68 | 0.416 | 1.11 | 0.125 | 0.355 |
| 3-Indoleacetic acid | 0.642 | 0.364 | 1.13 | 0.125 | 0.355 |
| alpha-ketoglutaric acid | 0.748 | 0.515 | 1.08 | 0.125 | 0.355 |
| Linoleic acid | 1.65 | 0.867 | 3.13 | 0.128 | 0.355 |
| Glutamic acid | 0.695 | 0.427 | 1.13 | 0.144 | 0.385 |
| Proline | 0.677 | 0.398 | 1.15 | 0.149 | 0.386 |
| Glyceric acid | 1.58 | 0.821 | 3.03 | 0.171 | 0.42 |
| L-5-Oxoproline | 1.58 | 0.818 | 3.05 | 0.174 | 0.42 |
| 2-Hydroxybutyric acid | 0.72 | 0.441 | 1.18 | 0.189 | 0.443 |
| 3-Hydroxybutyric acid | 0.704 | 0.41 | 1.21 | 0.204 | 0.454 |
| Lactic acid | 0.739 | 0.463 | 1.18 | 0.206 | 0.454 |
| Nonanoic acid | 1.45 | 0.792 | 2.66 | 0.228 | 0.49 |
| Serine | 0.738 | 0.445 | 1.22 | 0.237 | 0.494 |
| 4-Hydroxybutanoic acid | 1.39 | 0.779 | 2.49 | 0.263 | 0.533 |
| Eicosapentaenoic acid | 0.768 | 0.47 | 1.25 | 0.291 | 0.575 |
| Glycerol (1) | 1.51 | 0.684 | 3.31 | 0.309 | 0.585 |
| Glycine | 1.34 | 0.758 | 2.38 | 0.312 | 0.585 |
| Decanoic acid | 0.795 | 0.498 | 1.27 | 0.338 | 0.613 |
| Pyroglutamic acid | 1.39 | 0.696 | 2.79 | 0.349 | 0.613 |
| Stearic acid | 1.31 | 0.745 | 2.29 | 0.351 | 0.613 |
| 4-Deoxytetronic acid (2) | 1.33 | 0.701 | 2.54 | 0.379 | 0.638 |
| Tridecanoic acid | 1.35 | 0.687 | 2.66 | 0.383 | 0.638 |
| Glyceryl-glycoside | 1.35 | 0.649 | 2.81 | 0.421 | 0.687 |
| Fumaric acid | 1.25 | 0.71 | 2.19 | 0.444 | 0.708 |
| Arabinopyranose | 1.3 | 0.618 | 2.72 | 0.493 | 0.744 |
| 11-Eicosenoic acid | 1.26 | 0.649 | 2.45 | 0.494 | 0.744 |
| 1,3-Propanediol | 1.24 | 0.649 | 2.38 | 0.512 | 0.744 |

| Name | exp(coef) | Lower 95 % | Upper 95 % | Pr(>\|z\|) | p.adj |
|---|---|---|---|---|---|
| Nonadecanoic acid | 1.28 | 0.604 | 2.73 | 0.515 | 0.744 |
| Docosahexaenoic acid | 1.24 | 0.647 | 2.38 | 0.516 | 0.744 |
| Threonine | 0.864 | 0.547 | 1.37 | 0.531 | 0.744 |
| Bisphenol A | 1.19 | 0.683 | 2.07 | 0.539 | 0.744 |
| 4-Hydroxyphenyllactic acid | 1.22 | 0.634 | 2.34 | 0.554 | 0.744 |
| Arachidic acid | 1.17 | 0.697 | 1.95 | 0.556 | 0.744 |
| Aminomalonic acid | 1.19 | 0.653 | 2.18 | 0.566 | 0.744 |
| Succinic acid | 1.17 | 0.651 | 2.12 | 0.593 | 0.766 |
| Phenylalanine | 0.884 | 0.522 | 1.5 | 0.648 | 0.823 |
| 3-Indolepropionic acid | 0.91 | 0.574 | 1.44 | 0.69 | 0.863 |
| Arachidonic acid | 1.12 | 0.623 | 2.01 | 0.707 | 0.869 |
| Hydroxyproline | 1.11 | 0.624 | 1.98 | 0.721 | 0.872 |
| Ribitol (1) | 1.09 | 0.625 | 1.89 | 0.765 | 0.909 |
| Palmitic acid | 0.926 | 0.546 | 1.57 | 0.775 | 0.909 |
| Dodecanoic acid | 0.934 | 0.516 | 1.69 | 0.82 | 0.934 |
| Heptadecanoic acid (1) | 0.942 | 0.541 | 1.64 | 0.834 | 0.934 |
| Cholesterol | 0.935 | 0.499 | 1.75 | 0.835 | 0.934 |
| 1-Monopalmitin | 1.06 | 0.561 | 2.01 | 0.856 | 0.944 |
| alpha-Tocopherol | 1.05 | 0.56 | 1.96 | 0.884 | 0.961 |
| Oleic acid | 0.965 | 0.544 | 1.71 | 0.903 | 0.968 |
| 2-Palmitoylglycerol | 0.977 | 0.569 | 1.68 | 0.933 | 0.975 |
| Campesterol | 1.02 | 0.561 | 1.85 | 0.952 | 0.975 |
| Heptadecanoic acid (2) | 0.983 | 0.569 | 1.7 | 0.952 | 0.975 |
| Myristoleic acid | 0.987 | 0.553 | 1.76 | 0.964 | 0.975 |
| 1-Dodecanol | 1.01 | 0.569 | 1.79 | 0.975 | 0.975 |

###### 4.2.2.3 Top-Metabolite

```
## Call:
## survival::coxph(formula = survival::Surv(time = Charcot.tdiff,
##     event = Charcot.from.DATE) ~ Ribonic_acid + Age.x + Gender.x +
##     Hba1c_baseline + CALSBP + bmi + Smoking + Statin + log_Blood_TGA +
##     Total_cholesterol, data = data.km)
##
##   n= 596, number of events= 13
##    (41 observations deleted due to missingness)
##
##                        coef exp(coef)  se(coef)      z Pr(>|z|)
## Ribonic_acid       1.159583  3.188605  0.354197  3.274  0.00106 **
## Age.x              0.005852  1.005870  0.026693  0.219  0.82645
## Gender.x          -0.080659  0.922508  0.573072 -0.141  0.88807
## Hba1c_baseline    -0.160816  0.851449  0.285865 -0.563  0.57373
## CALSBP             0.004624  1.004635  0.016462  0.281  0.77877
## bmi                0.040727  1.041568  0.070670  0.576  0.56441
## Smoking           -0.960873  0.382559  1.055937 -0.910  0.36284
## Statin             0.264468  1.302738  0.731946  0.361  0.71786
## log_Blood_TGA      0.543023  1.721203  0.463491  1.172  0.24136
## Total_cholesterol -0.126826  0.880887  0.343617 -0.369  0.71206
## ---
## Signif. codes:  0 '***' 0.001 '**' 0.01 '*' 0.05 '.' 0.1 ' ' 1
##
##                   exp(coef) exp(-coef) lower .95 upper .95
## Ribonic_acid         3.1886     0.3136   1.59260     6.384
## Age.x                1.0059     0.9942   0.95460     1.060
## Gender.x             0.9225     1.0840   0.30003     2.836
## Hba1c_baseline       0.8514     1.1745   0.48622     1.491
## CALSBP               1.0046     0.9954   0.97274     1.038
## bmi                  1.0416     0.9601   0.90685     1.196
## Smoking              0.3826     2.6140   0.04829     3.030
## Statin               1.3027     0.7676   0.31033     5.469
## log_Blood_TGA        1.7212     0.5810   0.69392     4.269
## Total_cholesterol    0.8809     1.1352   0.44919     1.727
##
## Concordance= 0.854  (se = 0.038 )
## Likelihood ratio test= 19.52  on 10 df,   p=0.03
## Wald test            = 17.25  on 10 df,   p=0.07
## Score (logrank) test = 16.92  on 10 df,   p=0.08
```

###### 4.2.2.3.1 Forest Plot with Clinical Variables

- Top metabolite

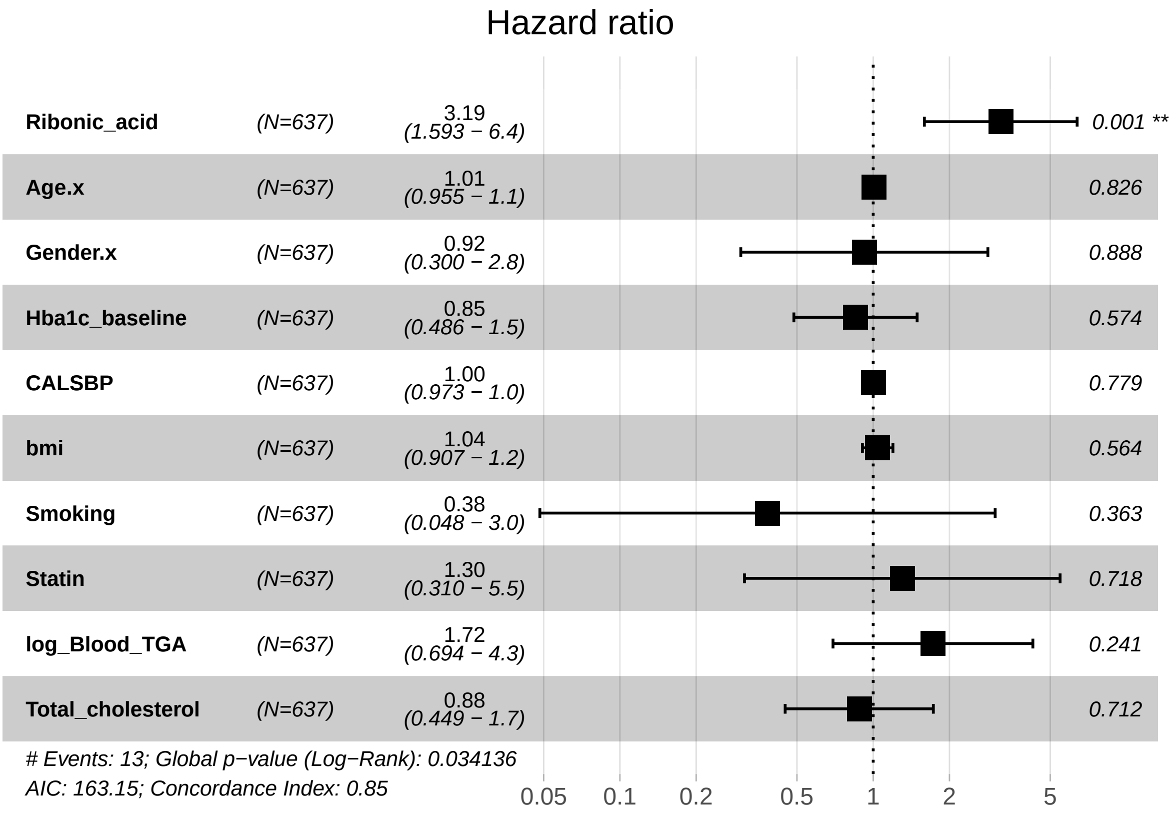

##### 4.2.3 Adusted Model with eGFR

###### 4.2.3.1 Forest Plot of Model Coefficients

```
## NULL
```

###### 4.2.3.2 Table with All Metabolites

| Name | exp(coef) | Lower 95 % | Upper 95 % | Pr(>\|z\|) | p.adj |
|---|---|---|---|---|---|
| 3-Indoleacetic acid | 0.482 | 0.26 | 0.891 | 0.02 | 0.499 |
| Tyrosine | 0.739 | 0.561 | 0.973 | 0.0313 | 0.499 |
| Tartronic acid | 1.95 | 1.03 | 3.68 | 0.0394 | 0.499 |
| Ethanolamine | 0.576 | 0.332 | 0.998 | 0.0491 | 0.499 |
| Hydroxylamine | 1.99 | 0.98 | 4.05 | 0.057 | 0.499 |
| Pyruvic acid | 0.624 | 0.384 | 1.02 | 0.0583 | 0.499 |
| Ribonic acid | 2.11 | 0.937 | 4.74 | 0.0716 | 0.499 |
| Citric acid | 1.79 | 0.949 | 3.39 | 0.072 | 0.499 |
| Glyceric acid | 1.83 | 0.941 | 3.57 | 0.0751 | 0.499 |
| Proline | 0.628 | 0.37 | 1.06 | 0.0842 | 0.499 |
| Malic acid | 1.72 | 0.913 | 3.24 | 0.0933 | 0.499 |
| Valine | 0.659 | 0.403 | 1.08 | 0.0972 | 0.499 |
| Linoleic acid | 1.73 | 0.905 | 3.32 | 0.0973 | 0.499 |
| Alanine | 0.68 | 0.43 | 1.07 | 0.0981 | 0.499 |
| alpha-ketoglutaric acid | 0.701 | 0.46 | 1.07 | 0.0997 | 0.499 |
| Stearic acid | 1.56 | 0.891 | 2.75 | 0.12 | 0.537 |
| 2-hydroxy Isovaleric acid | 0.657 | 0.386 | 1.12 | 0.122 | 0.537 |
| Benzeneacetic acid | 1.74 | 0.847 | 3.56 | 0.132 | 0.549 |
| 3-Hydroxybutyric acid | 0.669 | 0.383 | 1.17 | 0.156 | 0.615 |
| Octanoic acid | 0.743 | 0.489 | 1.13 | 0.164 | 0.615 |
| Creatinine | 1.6 | 0.791 | 3.24 | 0.19 | 0.68 |
| Ribitol (2) | 1.61 | 0.775 | 3.34 | 0.202 | 0.682 |
| Decanoic acid | 0.736 | 0.452 | 1.2 | 0.216 | 0.682 |
| L-5-Oxoproline | 1.54 | 0.772 | 3.07 | 0.221 | 0.682 |
| Glycerol (2) | 1.51 | 0.775 | 2.93 | 0.227 | 0.682 |
| Glutamic acid | 0.737 | 0.435 | 1.25 | 0.256 | 0.739 |
| Glycerol (1) | 1.54 | 0.711 | 3.32 | 0.274 | 0.762 |
| 4-Hydroxybutanoic acid | 1.36 | 0.771 | 2.38 | 0.291 | 0.78 |
| Arabinopyranose | 1.46 | 0.696 | 3.06 | 0.317 | 0.809 |
| Leucine | 0.862 | 0.64 | 1.16 | 0.326 | 0.809 |
| Methionine | 0.778 | 0.449 | 1.35 | 0.371 | 0.809 |
| Isoleucine | 0.854 | 0.604 | 1.21 | 0.373 | 0.809 |
| Lactic acid | 0.838 | 0.567 | 1.24 | 0.375 | 0.809 |
| Nonanoic acid | 1.32 | 0.705 | 2.47 | 0.386 | 0.809 |
| Docosahexaenoic acid | 1.37 | 0.666 | 2.8 | 0.394 | 0.809 |
| Bisphenol A | 1.3 | 0.713 | 2.36 | 0.395 | 0.809 |
| Myo inositol | 1.34 | 0.678 | 2.64 | 0.402 | 0.809 |
| Arachidic acid | 1.23 | 0.739 | 2.05 | 0.424 | 0.809 |
| Arachidonic acid | 1.29 | 0.687 | 2.42 | 0.429 | 0.809 |
| Nonadecanoic acid | 1.33 | 0.636 | 2.8 | 0.445 | 0.809 |
| 2,4-Dihydroxybutanoic acid | 1.31 | 0.647 | 2.66 | 0.45 | 0.809 |
| 1,3-Propanediol | 1.3 | 0.655 | 2.58 | 0.453 | 0.809 |
| 4-Hydroxybenzeneacetic acid | 1.33 | 0.616 | 2.86 | 0.47 | 0.812 |
| Tridecanoic acid | 1.25 | 0.674 | 2.33 | 0.476 | 0.812 |
| 3,4-Dihydroxybutanoic acid | 1.28 | 0.633 | 2.58 | 0.492 | 0.814 |
| Serine | 0.84 | 0.506 | 1.39 | 0.499 | 0.814 |
| 11-Eicosenoic acid | 1.23 | 0.649 | 2.35 | 0.521 | 0.831 |
| 4-Deoxytetronic acid (1) | 1.23 | 0.62 | 2.45 | 0.549 | 0.858 |
| Eicosapentaenoic acid | 0.852 | 0.483 | 1.5 | 0.581 | 0.89 |
| Phenylalanine | 0.869 | 0.501 | 1.51 | 0.619 | 0.928 |

| Name | exp(coef) | Lower 95 % | Upper 95 % | Pr(>\|z\|) | p.adj |
|---|---|---|---|---|---|
| Threonine | 0.9 | 0.568 | 1.43 | 0.653 | 0.958 |
| Aminomalonic acid | 1.15 | 0.611 | 2.16 | 0.664 | 0.958 |
| Dodecanoic acid | 0.887 | 0.48 | 1.64 | 0.7 | 0.964 |
| 4-Deoxytetronic acid (2) | 0.894 | 0.49 | 1.63 | 0.717 | 0.964 |
| Hydroxyproline | 0.896 | 0.492 | 1.63 | 0.72 | 0.964 |
| Succinic acid | 1.12 | 0.591 | 2.12 | 0.732 | 0.964 |
| Fumaric acid | 1.1 | 0.627 | 1.94 | 0.733 | 0.964 |
| Heptadecanoic acid (1) | 0.919 | 0.537 | 1.57 | 0.757 | 0.978 |
| Glycine | 1.06 | 0.58 | 1.96 | 0.84 | 0.992 |
| 1-Monopalmitin | 1.07 | 0.566 | 2.01 | 0.842 | 0.992 |
| Palmitic acid | 1.06 | 0.59 | 1.91 | 0.843 | 0.992 |
| alpha-Tocopherol | 1.06 | 0.581 | 1.93 | 0.849 | 0.992 |
| Pyroglutamic acid | 1.06 | 0.544 | 2.07 | 0.862 | 0.992 |
| 2-Hydroxybutyric acid | 0.958 | 0.568 | 1.61 | 0.871 | 0.992 |
| 2-Palmitoylglycerol | 1.04 | 0.573 | 1.87 | 0.909 | 0.992 |
| 4-Hydroxyphenyllactic acid | 0.964 | 0.511 | 1.82 | 0.91 | 0.992 |
| 1-Dodecanol | 1.03 | 0.571 | 1.86 | 0.921 | 0.992 |
| Myristoleic acid | 1.02 | 0.616 | 1.7 | 0.928 | 0.992 |
| Cholesterol | 1.02 | 0.555 | 1.88 | 0.944 | 0.992 |
| Oleic acid | 1.02 | 0.597 | 1.74 | 0.945 | 0.992 |
| Campesterol | 0.985 | 0.544 | 1.78 | 0.96 | 0.992 |
| Heptadecanoic acid (2) | 1.01 | 0.581 | 1.77 | 0.963 | 0.992 |
| Ribitol (1) | 0.992 | 0.569 | 1.73 | 0.976 | 0.992 |
| 3-Indolepropionic acid | 1.01 | 0.641 | 1.58 | 0.979 | 0.992 |
| Glyceryl-glycoside | 1 | 0.515 | 1.94 | 0.999 | 0.999 |

###### 4.2.3.3 Top-Metabolite from Adjusted Model

```
## Call:
## survival::coxph(formula = survival::Surv(time = Charcot.tdiff,
##     event = Charcot.from.DATE) ~ Ribonic_acid + Age.x + Gender.x +
##     Hba1c_baseline + CALSBP + bmi + Smoking + Statin + log_Blood_TGA +
##     Total_cholesterol + egfr, data = data.km)
##
##   n= 594, number of events= 13
##    (43 observations deleted due to missingness)
##
##                        coef exp(coef)  se(coef)      z Pr(>|z|)
## Ribonic_acid       0.757418  2.132763  0.420427  1.802   0.0716 .
## Age.x              0.003861  1.003869  0.027314  0.141   0.8876
## Gender.x           0.023550  1.023829  0.572240  0.041   0.9672
## Hba1c_baseline    -0.151465  0.859448  0.285140 -0.531   0.5953
## CALSBP             0.002568  1.002571  0.016434  0.156   0.8758
## bmi                0.047709  1.048865  0.071347  0.669   0.5037
## Smoking           -0.821791  0.439644  1.069596 -0.768   0.4423
## Statin             0.120647  1.128227  0.737486  0.164   0.8701
## log_Blood_TGA      0.470718  1.601143  0.469502  1.003   0.3161
## Total_cholesterol -0.128937  0.879029  0.330540 -0.390   0.6965
## egfr              -0.020804  0.979411  0.013434 -1.549   0.1215
## ---
## Signif. codes:  0 '***' 0.001 '**' 0.01 '*' 0.05 '.' 0.1 ' ' 1
##
##                   exp(coef) exp(-coef) lower .95 upper .95
## Ribonic_acid         2.1328     0.4689   0.93557     4.862
## Age.x                1.0039     0.9961   0.95154     1.059
## Gender.x             1.0238     0.9767   0.33353     3.143
## Hba1c_baseline       0.8594     1.1635   0.49148     1.503
## CALSBP               1.0026     0.9974   0.97079     1.035
## bmi                  1.0489     0.9534   0.91199     1.206
## Smoking              0.4396     2.2746   0.05403     3.577
## Statin               1.1282     0.8863   0.26586     4.788
## log_Blood_TGA        1.6011     0.6246   0.63795     4.019
## Total_cholesterol    0.8790     1.1376   0.45988     1.680
## egfr                 0.9794     1.0210   0.95396     1.006
##
## Concordance= 0.873  (se = 0.027 )
## Likelihood ratio test= 21.97  on 11 df,   p=0.02
## Wald test            = 18.89  on 11 df,   p=0.06
## Score (logrank) test = 22.45  on 11 df,   p=0.02
```

###### 4.2.3.3.1 Forest Plot with Clinical Variables

- Top metabolite from adjusted model

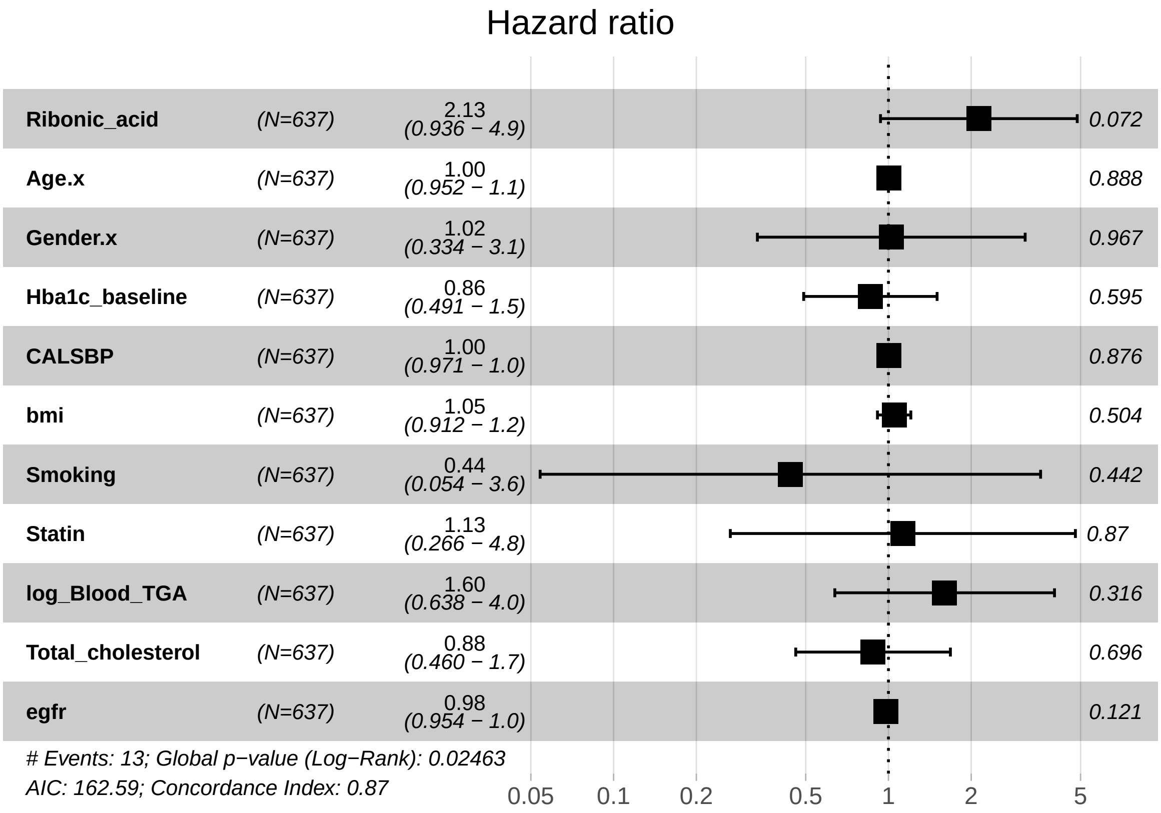

##### 4.2.4 Fully Adusted Model

###### 4.2.4.1 Forest Plot of Model Coefficients

```
## NULL
```

###### 4.2.4.2 Table with All Metabolites

| Name | exp(coef) | Lower 95 % | Upper 95 % | Pr(>\|z\|) | p.adj |
|---|---|---|---|---|---|
| Proline | 0.475 | 0.271 | 0.832 | 0.00933 | 0.583 |
| Tyrosine | 0.701 | 0.523 | 0.938 | 0.0169 | 0.583 |
| Ethanolamine | 0.509 | 0.276 | 0.941 | 0.0313 | 0.583 |
| Glyceric acid | 2.26 | 1.04 | 4.92 | 0.0402 | 0.583 |
| Alanine | 0.626 | 0.394 | 0.996 | 0.0483 | 0.583 |
| Citric acid | 1.9 | 0.996 | 3.63 | 0.0513 | 0.583 |
| 3-Indoleacetic acid | 0.528 | 0.276 | 1.01 | 0.0544 | 0.583 |
| Stearic acid | 1.76 | 0.954 | 3.24 | 0.0705 | 0.622 |
| Tartronic acid | 1.81 | 0.93 | 3.53 | 0.0806 | 0.622 |
| Ribitol (2) | 2.08 | 0.909 | 4.78 | 0.083 | 0.622 |
| Hydroxylamine | 1.89 | 0.879 | 4.09 | 0.103 | 0.65 |
| Glutamic acid | 0.615 | 0.343 | 1.11 | 0.104 | 0.65 |
| Benzeneacetic acid | 1.82 | 0.849 | 3.92 | 0.124 | 0.667 |
| Leucine | 0.749 | 0.519 | 1.08 | 0.125 | 0.667 |
| 3-Hydroxybutyric acid | 0.625 | 0.332 | 1.18 | 0.145 | 0.678 |
| Valine | 0.687 | 0.415 | 1.14 | 0.145 | 0.678 |
| Ribonic acid | 1.9 | 0.776 | 4.65 | 0.16 | 0.678 |
| 4-Hydroxybenzeneacetic acid | 1.82 | 0.786 | 4.2 | 0.163 | 0.678 |
| Decanoic acid | 0.697 | 0.395 | 1.23 | 0.212 | 0.699 |
| Arachidonic acid | 1.51 | 0.765 | 2.97 | 0.235 | 0.699 |
| L-5-Oxoproline | 1.58 | 0.737 | 3.41 | 0.239 | 0.699 |
| Nonadecanoic acid | 1.66 | 0.672 | 4.11 | 0.272 | 0.699 |
| 2-hydroxy Isovaleric acid | 0.72 | 0.4 | 1.29 | 0.272 | 0.699 |
| Pyruvic acid | 0.752 | 0.446 | 1.27 | 0.285 | 0.699 |
| 2,4-Dihydroxybutanoic acid | 1.5 | 0.711 | 3.16 | 0.287 | 0.699 |
| Docosahexaenoic acid | 1.57 | 0.673 | 3.65 | 0.297 | 0.699 |
| Octanoic acid | 0.796 | 0.517 | 1.23 | 0.3 | 0.699 |
| Malic acid | 1.41 | 0.736 | 2.69 | 0.302 | 0.699 |
| Glycerol (1) | 1.61 | 0.651 | 3.99 | 0.302 | 0.699 |
| Isoleucine | 0.791 | 0.503 | 1.24 | 0.31 | 0.699 |
| Linoleic acid | 1.4 | 0.729 | 2.69 | 0.313 | 0.699 |
| Lactic acid | 0.824 | 0.564 | 1.2 | 0.318 | 0.699 |
| 4-Hydroxybutanoic acid | 1.39 | 0.711 | 2.7 | 0.338 | 0.699 |
| Myo inositol | 1.43 | 0.689 | 2.95 | 0.339 | 0.699 |
| Threonine | 0.796 | 0.491 | 1.29 | 0.356 | 0.699 |
| Nonanoic acid | 1.38 | 0.693 | 2.77 | 0.357 | 0.699 |
| Heptadecanoic acid (1) | 0.767 | 0.428 | 1.37 | 0.373 | 0.699 |
| Serine | 0.793 | 0.47 | 1.34 | 0.385 | 0.699 |
| Glycerol (2) | 1.37 | 0.67 | 2.8 | 0.389 | 0.699 |
| Tridecanoic acid | 1.32 | 0.698 | 2.51 | 0.39 | 0.699 |
| Creatinine | 1.37 | 0.669 | 2.79 | 0.391 | 0.699 |
| Arabinopyranose | 1.41 | 0.644 | 3.08 | 0.391 | 0.699 |
| 11-Eicosenoic acid | 1.3 | 0.647 | 2.61 | 0.462 | 0.805 |
| alpha-ketoglutaric acid | 0.813 | 0.448 | 1.48 | 0.497 | 0.832 |
| 3-Indolepropionic acid | 1.19 | 0.717 | 1.98 | 0.499 | 0.832 |
| Aminomalonic acid | 1.26 | 0.617 | 2.57 | 0.527 | 0.85 |
| 3,4-Dihydroxybutanoic acid | 1.29 | 0.578 | 2.86 | 0.538 | 0.85 |
| Pyroglutamic acid | 1.26 | 0.587 | 2.71 | 0.552 | 0.85 |
| 1,3-Propanediol | 1.26 | 0.582 | 2.74 | 0.555 | 0.85 |
| Dodecanoic acid | 0.832 | 0.434 | 1.59 | 0.578 | 0.867 |

| Name | exp(coef) | Lower 95 % | Upper 95 % | Pr(>\|z\|) | p.adj |
|---|---|---|---|---|---|
| Methionine | 0.851 | 0.451 | 1.61 | 0.62 | 0.911 |
| 4-Deoxytetronic acid (2) | 0.86 | 0.453 | 1.63 | 0.646 | 0.931 |
| Campesterol | 1.16 | 0.602 | 2.23 | 0.659 | 0.932 |
| 4-Deoxytetronic acid (1) | 1.18 | 0.539 | 2.58 | 0.679 | 0.943 |
| Eicosapentaenoic acid | 0.893 | 0.474 | 1.68 | 0.726 | 0.97 |
| Fumaric acid | 1.1 | 0.603 | 1.99 | 0.763 | 0.97 |
| 2-Hydroxybutyric acid | 0.921 | 0.523 | 1.62 | 0.776 | 0.97 |
| Palmitic acid | 1.08 | 0.588 | 1.97 | 0.809 | 0.97 |
| Cholesterol | 0.93 | 0.497 | 1.74 | 0.82 | 0.97 |
| Hydroxyproline | 1.07 | 0.558 | 2.05 | 0.839 | 0.97 |
| Phenylalanine | 1.07 | 0.552 | 2.08 | 0.84 | 0.97 |
| 1-Dodecanol | 1.06 | 0.56 | 2.03 | 0.849 | 0.97 |
| Bisphenol A | 1.06 | 0.576 | 1.94 | 0.858 | 0.97 |
| Glycine | 1.06 | 0.546 | 2.06 | 0.861 | 0.97 |
| Arachidic acid | 1.05 | 0.585 | 1.9 | 0.862 | 0.97 |
| 4-Hydroxyphenyllactic acid | 0.938 | 0.458 | 1.92 | 0.862 | 0.97 |
| Succinic acid | 0.945 | 0.488 | 1.83 | 0.867 | 0.97 |
| Glyceryl-glycoside | 0.956 | 0.474 | 1.93 | 0.9 | 0.977 |
| Oleic acid | 1.04 | 0.6 | 1.78 | 0.901 | 0.977 |
| Heptadecanoic acid (2) | 1.03 | 0.579 | 1.82 | 0.925 | 0.977 |
| Ribitol (1) | 1.03 | 0.569 | 1.85 | 0.934 | 0.977 |
| 1-Monopalmitin | 0.969 | 0.438 | 2.15 | 0.938 | 0.977 |
| Myristoleic acid | 1.01 | 0.608 | 1.69 | 0.957 | 0.982 |
| 2-Palmitoylglycerol | 1.01 | 0.53 | 1.93 | 0.972 | 0.982 |
| alpha-Tocopherol | 1.01 | 0.555 | 1.83 | 0.982 | 0.982 |

###### 4.2.4.3 Top-Metabolite from Adjusted Model

```
## Call:
## survival::coxph(formula = survival::Surv(time = Charcot.tdiff,
##     event = Charcot.from.DATE) ~ Ribonic_acid + Age.x + Gender.x +
##     Hba1c_baseline + CALSBP + bmi + Smoking + Statin + log_Blood_TGA +
##     Total_cholesterol + egfr + logUAER, data = data.km)
##
##   n= 561, number of events= 12
##    (76 observations deleted due to missingness)
##
##                        coef exp(coef)  se(coef)      z Pr(>|z|)
## Ribonic_acid       0.652055  1.919482  0.464544  1.404   0.1604
## Age.x              0.031762  1.032272  0.031573  1.006   0.3144
## Gender.x          -0.718644  0.487413  0.670599 -1.072   0.2839
## Hba1c_baseline    -0.270277  0.763168  0.302165 -0.894   0.3711
## CALSBP            -0.007227  0.992799  0.018765 -0.385   0.7001
## bmi                0.029357  1.029793  0.074984  0.392   0.6954
## Smoking           -1.451550  0.234207  1.125927 -1.289   0.1973
## Statin            -0.459668  0.631493  0.774852 -0.593   0.5530
## log_Blood_TGA      0.729517  2.074079  0.461130  1.582   0.1136
## Total_cholesterol -0.264300  0.767743  0.341581 -0.774   0.4391
## egfr              -0.010075  0.989975  0.014653 -0.688   0.4917
## logUAER            0.350711  1.420077  0.158507  2.213   0.0269 *
## ---
## Signif. codes:  0 '***' 0.001 '**' 0.01 '*' 0.05 '.' 0.1 ' ' 1
##
##                   exp(coef) exp(-coef) lower .95 upper .95
## Ribonic_acid         1.9195     0.5210   0.77226     4.771
## Age.x                1.0323     0.9687   0.97033     1.098
## Gender.x             0.4874     2.0516   0.13094     1.814
## Hba1c_baseline       0.7632     1.3103   0.42210     1.380
## CALSBP               0.9928     1.0073   0.95695     1.030
## bmi                  1.0298     0.9711   0.88904     1.193
## Smoking              0.2342     4.2697   0.02578     2.128
## Statin               0.6315     1.5835   0.13830     2.884
## log_Blood_TGA        2.0741     0.4821   0.84006     5.121
## Total_cholesterol    0.7677     1.3025   0.39306     1.500
## egfr                 0.9900     1.0101   0.96195     1.019
## logUAER              1.4201     0.7042   1.04086     1.937
##
## Concordance= 0.873  (se = 0.051 )
## Likelihood ratio test= 26.37  on 12 df,   p=0.01
## Wald test            = 22.11  on 12 df,   p=0.04
## Score (logrank) test = 28.74  on 12 df,   p=0.004
```

###### 4.2.4.3.1 Forest Plot with Clinical Variables

- Top metabolite from adjusted model

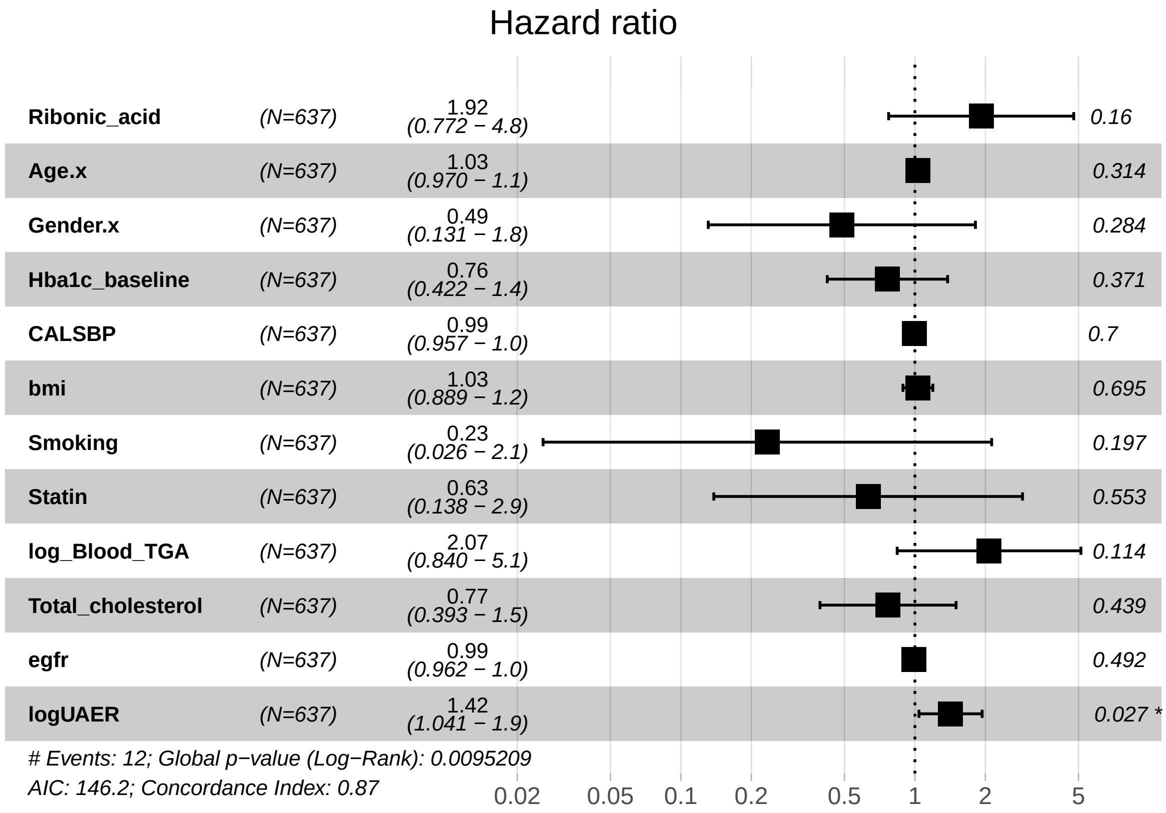

### 5 Amputation

#### 5.1 Amputation at DATE

##### 5.1.1 Crude Model

```
## [1] "Fitting models:"
## [1] "~ Amputation.at.DATE"
## [1] ""
```

###### 5.1.1.1 Forest Plot of Model Coefficients

```
## Warning: Ignoring unknown aesthetics: x
```

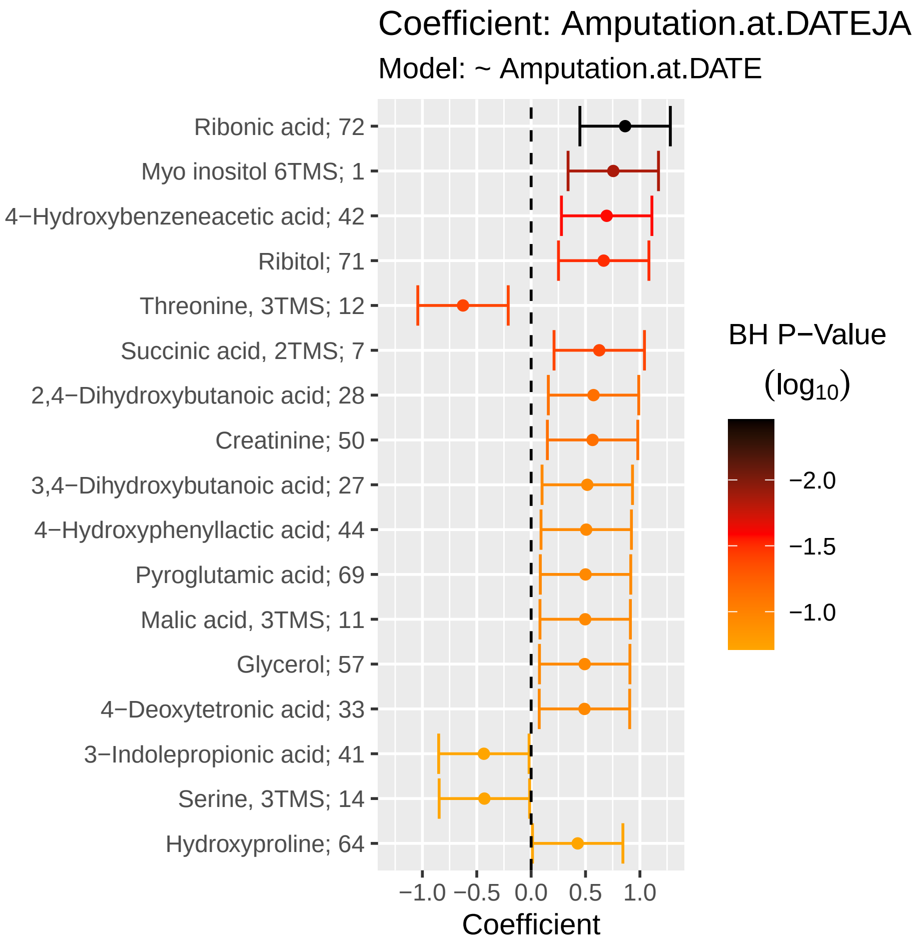

###### 5.1.1.2 Tables of Model Coefficients

```
## [1] ""
## [1] "Table: Amputation.at.DATEJA"
## [1] "  (from model: "
## [1] "   ~ Amputation.at.DATE)"
## [1] ""
##                               Name Coefficient    CI.L    CI.R  p.value   p.adj
## 1                 Ribonic acid; 72       0.864  0.4480  1.2800 4.65e-05 0.00349
## 2             Myo inositol 6TMS; 1       0.755  0.3390  1.1700 3.72e-04 0.01390
## 3  4-Hydroxybenzeneacetic acid; 4       0.695  0.2790  1.1100 1.06e-03 0.02650
## 4                      Ribitol; 71       0.667  0.2510  1.0800 1.66e-03 0.03120
## 5             Threonine, 3TMS; 12      -0.627 -1.0400 -0.2110 3.15e-03 0.03960
## 6           Succinic acid, 2TMS; 7       0.626  0.2100  1.0400 3.16e-03 0.03960
## 7  2,4-Dihydroxybutanoic acid; 28       0.574  0.1580  0.9890 6.86e-03 0.07280
## 8                   Creatinine; 50       0.565  0.1490  0.9810 7.76e-03 0.07280
## 9  3,4-Dihydroxybutanoic acid; 27       0.516  0.1000  0.9320 1.50e-02 0.11200
## 10 4-Hydroxyphenyllactic acid; 44       0.506  0.0906  0.9220 1.70e-02 0.11200
## 11          Pyroglutamic acid; 69       0.500  0.0842  0.9160 1.84e-02 0.11200
## 12           Malic acid, 3TMS; 11       0.497  0.0810  0.9130 1.92e-02 0.11200
## 13                    Glycerol; 57       0.492  0.0765  0.9080 2.03e-02 0.11200
## 14       4-Deoxytetronic acid; 33       0.490  0.0744  0.9060 2.09e-02 0.11200
## 15     3-Indolepropionic acid; 41      -0.435 -0.8500 -0.0189 4.05e-02 0.19300
## 16                Serine, 3TMS; 14      -0.430 -0.8460 -0.0143 4.26e-02 0.19300
## 17              Hydroxyproline; 64       0.428  0.0121  0.8440 4.37e-02 0.19300
```

###### 5.1.1.3 Table with All Metabolites

```
## [1] ""
## [1] "Table: Amputation.at.DATEJA"
## [1] "  (from model: "
## [1] "   ~ Amputation.at.DATE)"
## [1] ""
##                               Name Coefficient    CI.L    CI.R p.value   p.adj
## 1               Ribonic acid; 72     0.86400  0.4480  1.2800 4.65e-05 0.00349
## 2           Myo inositol 6TMS; 1     0.75500  0.3390  1.1700 3.72e-04 0.01390
## 3  4-Hydroxybenzeneacetic acid; 4    0.69500  0.2790  1.1100 1.06e-03 0.02650
## 4                    Ribitol; 71     0.66700  0.2510  1.0800 1.66e-03 0.03120
## 5            Threonine, 3TMS; 12    -0.62700 -1.0400 -0.2110 3.15e-03 0.03960
## 6          Succinic acid, 2TMS; 7    0.62600  0.2100  1.0400 3.16e-03 0.03960
## 7  2,4-Dihydroxybutanoic acid; 28    0.57400  0.1580  0.9890 6.86e-03 0.07280
## 8                 Creatinine; 50     0.56500  0.1490  0.9810 7.76e-03 0.07280
## 9  3,4-Dihydroxybutanoic acid; 27    0.51600  0.1000  0.9320 1.50e-02 0.11200
## 10 4-Hydroxyphenyllactic acid; 44    0.50600  0.0906  0.9220 1.70e-02 0.11200
## 11          Pyroglutamic acid; 69    0.50000  0.0842  0.9160 1.84e-02 0.11200
## 12           Malic acid, 3TMS; 11    0.49700  0.0810  0.9130 1.92e-02 0.11200
## 13                  Glycerol; 57     0.49200  0.0765  0.9080 2.03e-02 0.11200
## 14       4-Deoxytetronic acid; 33    0.49000  0.0744  0.9060 2.09e-02 0.11200
## 15     3-Indolepropionic acid; 41   -0.43500 -0.8500 -0.0189 4.05e-02 0.19300
## 16              Serine, 3TMS; 14    -0.43000 -0.8460 -0.0143 4.26e-02 0.19300
## 17             Hydroxyproline; 64    0.42800  0.0121  0.8440 4.37e-02 0.19300
## 18        3-Indoleacetic acid; 40    0.37700 -0.0384  0.7930 7.53e-02 0.31400
## 19                  Tyrosine; 75    -0.36900 -0.7850  0.0470 8.21e-02 0.31700
## 20              Valine, 2TMS; 20    -0.36600 -0.7820  0.0500 8.46e-02 0.31700
## 21          Methionine, 2TMS; 16    -0.35900 -0.7750  0.0564 9.03e-02 0.32200
## 22             Leucine, 2TMS; 19    -0.35300 -0.7690  0.0629 9.63e-02 0.32800
## 23         Glyceryl-glycoside; 59    0.34400 -0.0715  0.7600 1.05e-01 0.34100
## 24       4-Deoxytetronic acid; 32    0.33900 -0.0765  0.7550 1.10e-01 0.34300
## 25         Heptadecanoic acid; 61    0.32900 -0.0871  0.7440 1.21e-01 0.36400
## 26        Phenylalanine, 2TMS; 13   -0.30400 -0.7190  0.1120 1.52e-01 0.42500
## 27         Glutamic acid, 3TMS; 8   -0.30300 -0.7190  0.1130 1.53e-01 0.42500
## 28     4-Hydroxybutanoic acid; 43    0.28900 -0.1270  0.7040 1.74e-01 0.46500
## 29                   Ribitol; 70     0.26200 -0.1540  0.6780 2.17e-01 0.56200
## 30            Myristoleic acid; 65    0.24500 -0.1700  0.6610 2.47e-01 0.61800
## 31               Campesterol; 49    -0.23600 -0.6520  0.1790 2.65e-01 0.62300
## 32             Alanine, 2TMS; 25     0.23100 -0.1850  0.6470 2.76e-01 0.62300
## 33           Stearic acid, TMS; 2    0.23000 -0.1860  0.6450 2.79e-01 0.62300
## 34         Heptadecanoic acid; 60    0.22800 -0.1880  0.6440 2.83e-01 0.62300
## 35          Citric acid, 4TMS; 6     0.22100 -0.1950  0.6370 2.98e-01 0.62600
## 36           Dodecanoic acid; 54     0.21900 -0.1970  0.6340 3.03e-01 0.62600
## 37        2-Palmitoylglycerol; 39   -0.21600 -0.6320  0.2000 3.09e-01 0.62600
## 38          Cholesterol, TMS; 23    -0.19500 -0.6110  0.2200 3.57e-01 0.68900
## 39          Fumaric acid, 2TMS; 9    0.19400 -0.2220  0.6090 3.62e-01 0.68900
## 40  2-hydroxy Isovaleric acid; 38   -0.18600 -0.6020  0.2290 3.80e-01 0.68900
## 41          Aminomalonic acid; 45   -0.18500 -0.6000  0.2310 3.84e-01 0.68900
## 42               Pyruvic acid; 31    0.18200 -0.2340  0.5980 3.91e-01 0.68900
## 43         11-Eicosenoic acid; 35    0.18100 -0.2350  0.5960 3.95e-01 0.68900
## 44              Hydroxylamine; 62    0.15600 -0.2600  0.5720 4.62e-01 0.78300
## 45     Eicosapentaenoic acid; 55    -0.15300 -0.5690  0.2630 4.70e-01 0.78300
## 46          Benzeneacetic acid; 47    0.14600 -0.2700  0.5620 4.92e-01 0.79700
```

```
## 47               Proline, 2TMS; 21    -0.14300 -0.5590  0.2730 5.00e-01 0.79700
## 48          alpha-Tocopherol; 26    -0.13600 -0.5510  0.2800 5.23e-01 0.80800
## 49            1-Monopalmitin; 37     0.13300 -0.2830  0.5480 5.32e-01 0.80800
## 50             Decanoic acid; 52     0.13100 -0.2850  0.5460 5.38e-01 0.80800
## 51 2-Hydroxybutyric acid, 2TMS; 2     0.12500 -0.2910  0.5410 5.56e-01 0.81700
## 52         Palmitic acid, TMS; 5     0.12100 -0.2950  0.5370 5.68e-01 0.81900
## 53          Tridecanoic acid; 74     0.11700 -0.2990  0.5320 5.83e-01 0.82500
## 54     Arachidonic acid, TMS; 24     0.11000 -0.3060  0.5260 6.04e-01 0.83700
## 55          Isoleucine, 2TMS; 18    -0.10700 -0.5230  0.3090 6.14e-01 0.83700
## 56              Ethanolamine; 56     0.09870 -0.3170  0.5150 6.42e-01 0.83700
## 57           Arabinopyranose; 51    -0.09410 -0.5100  0.3220 6.57e-01 0.83700
## 58               Lactic acid; 29     0.09380 -0.3220  0.5100 6.58e-01 0.83700
## 59         Nonadecanoic acid; 66     0.09020 -0.3260  0.5060 6.71e-01 0.83700
## 60             Glyceric acid; 30    -0.08710 -0.5030  0.3290 6.81e-01 0.83700
## 61               1-Dodecanol; 36     0.08220 -0.3340  0.4980 6.98e-01 0.83700
## 62 alpha-ketoglutaric acid, TMS M     0.08110 -0.3350  0.4970 7.02e-01 0.83700
## 63         Linoleic acid, TMS; 4    -0.08100 -0.4970  0.3350 7.03e-01 0.83700
## 64            Arachidic acid; 46     0.07690 -0.3390  0.4930 7.17e-01 0.84000
## 65 3-Hydroxybutyric acid, 2TMS; 1     0.06500 -0.3510  0.4810 7.59e-01 0.85600
## 66           1,3-Propanediol; 34     0.06460 -0.3510  0.4800 7.61e-01 0.85600
## 67            Tartronic acid; 73    -0.06350 -0.4790  0.3520 7.65e-01 0.85600
## 68             Octanoic acid; 68     0.05990 -0.3560  0.4760 7.78e-01 0.85800
## 69                  Glycerol; 58     0.04750 -0.3680  0.4630 8.23e-01 0.89400
## 70               Bisphenol A; 48    -0.04260 -0.4580  0.3730 8.41e-01 0.90100
## 71            L-5-Oxoproline; 63    -0.03410 -0.4500  0.3820 8.72e-01 0.92200
## 72             Glycine, 3TMS; 17     0.01970 -0.3960  0.4360 9.26e-01 0.96400
## 73             Nonanoic acid; 67    -0.01080 -0.4270  0.4050 9.59e-01 0.97500
## 74             Oleic acid, TMS; 3     0.01010 -0.4060  0.4260 9.62e-01 0.97500
## 75       Docosahexaenoic acid; 53     0.00511 -0.4110  0.4210 9.81e-01 0.98100
```

##### 5.1.2 Adjusted Model

```
## [1] "Fitting models:"
## [1] "~ Amputation.at.DATE + Age.x + Gender.x + Hba1c_baseline + CALSBP + bmi + Smoking + Statin + log
## [1] ""
```

###### 5.1.2.1 Forest Plot of Model Coefficients

```
## Warning: Ignoring unknown aesthetics: x
## Ignoring unknown aesthetics: x
## Ignoring unknown aesthetics: x
## Ignoring unknown aesthetics: x
## Ignoring unknown aesthetics: x
## Ignoring unknown aesthetics: x
## Ignoring unknown aesthetics: x
## Ignoring unknown aesthetics: x
## Ignoring unknown aesthetics: x
## Ignoring unknown aesthetics: x
```

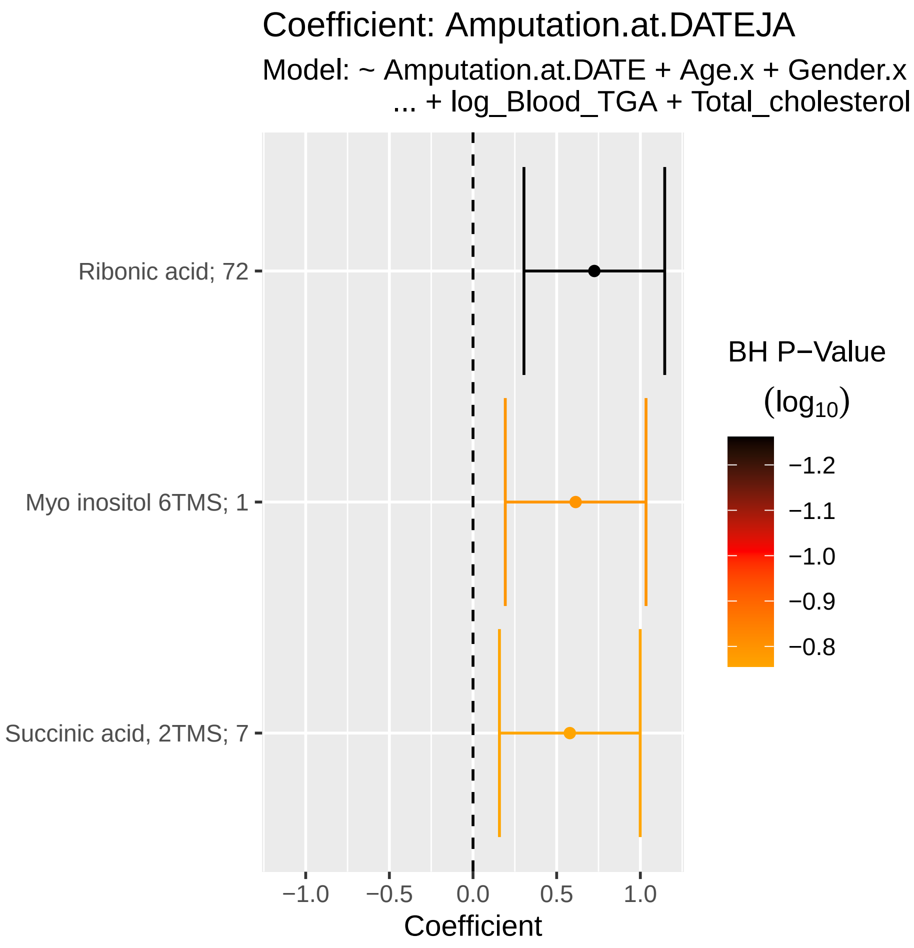

###### 5.1.2.2 Tables of Model Coefficients

```
## [1] ""
## [1] "Table: Amputation.at.DATEJA"
## [1] "  (from model: "
## [1] "   ~ Amputation.at.DATE + Age.x + Gender.x + Hba1c_baseline"
## [1] "     + CALSBP + bmi + Smoking + Statin + log_Blood_TGA +"
## [1] "     Total_cholesterol)"
## [1] ""
##                     Name Coefficient  CI.L  CI.R p.value  p.adj
## 1       Ribonic acid; 72       0.725 0.304 1.150 0.00073 0.0547
## 2   Myo inositol 6TMS; 1       0.613 0.193 1.030 0.00427 0.1600
## 3 Succinic acid, 2TMS; 7       0.579 0.158 0.999 0.00702 0.1760
## [1] ""
## [1] "Table: Age.x"
## [1] "  (from model: "
## [1] "   ~ Amputation.at.DATE + Age.x + Gender.x + Hba1c_baseline"
## [1] "     + CALSBP + bmi + Smoking + Statin + log_Blood_TGA +"
## [1] "     Total_cholesterol)"
## [1] ""
##                              Name Coefficient      CI.L      CI.R  p.value
## 1       Eicosapentaenoic acid; 55     0.02240  1.57e-02  2.91e-02 5.20e-11
## 2  2,4-Dihydroxybutanoic acid; 28     0.01680  1.01e-02  2.35e-02 8.61e-07
## 3  4-Hydroxybenzeneacetic acid; 4     0.01630  9.57e-03  2.30e-02 1.90e-06
## 4            Myo inositol 6TMS; 1     0.01450  7.80e-03  2.12e-02 2.21e-05
## 5                     Ribitol; 71     0.01410  7.41e-03  2.08e-02 3.66e-05
## 6        Docosahexaenoic acid; 53     0.01350  6.77e-03  2.02e-02 8.08e-05
## 7          3-Indoleacetic acid; 40     0.01330  6.58e-03  2.00e-02 1.01e-04
## 8           Aminomalonic acid; 45     0.01150  4.80e-03  1.82e-02 7.61e-04
## 9                Ribonic acid; 72     0.01130  4.63e-03  1.80e-02 9.11e-04
## 10 alpha-ketoglutaric acid, TMS M     0.01100  4.29e-03  1.77e-02 1.29e-03
## 11                    Ribitol; 70     0.01060  3.87e-03  1.73e-02 1.98e-03
## 12           Malic acid, 3TMS; 11     0.01030  3.57e-03  1.70e-02 2.65e-03
## 13               Pyruvic acid; 31     0.01020  3.52e-03  1.69e-02 2.77e-03
## 14          Fumaric acid, 2TMS; 9     0.01010  3.45e-03  1.68e-02 2.97e-03
## 15 3,4-Dihydroxybutanoic acid; 27     0.01010  3.43e-03  1.68e-02 3.04e-03
## 16           Citric acid, 4TMS; 6     0.01010  3.41e-03  1.68e-02 3.10e-03
## 17           alpha-Tocopherol; 26     0.01010  3.40e-03  1.68e-02 3.13e-03
## 18              Decanoic acid; 52     0.01010  3.36e-03  1.67e-02 3.23e-03
## 19              Glyceric acid; 30     0.00983  3.14e-03  1.65e-02 4.00e-03
## 20 4-Hydroxyphenyllactic acid; 44     0.00964  2.95e-03  1.63e-02 4.76e-03
## 21              Glycine, 3TMS; 17     0.00932  2.63e-03  1.60e-02 6.35e-03
## 22         Succinic acid, 2TMS; 7     0.00888  2.19e-03  1.56e-02 9.30e-03
## 23         11-Eicosenoic acid; 35     0.00849  1.80e-03  1.52e-02 1.29e-02
## 24          Pyroglutamic acid; 69     0.00840  1.70e-03  1.51e-02 1.39e-02
## 25              Alanine, 2TMS; 25     0.00831  1.61e-03  1.50e-02 1.50e-02
## 26                 Creatinine; 50     0.00751  8.18e-04  1.42e-02 2.78e-02
## 27        Phenylalanine, 2TMS; 13     0.00737  6.78e-04  1.41e-02 3.09e-02
## 28               Valine, 2TMS; 20    -0.00733 -1.40e-02 -6.36e-04 3.19e-02
## 29              Leucine, 2TMS; 19    -0.00726 -1.40e-02 -5.68e-04 3.35e-02
## 30           Myristoleic acid; 65     0.00706  3.68e-04  1.38e-02 3.87e-02
## 31         Heptadecanoic acid; 61     0.00688  1.90e-04  1.36e-02 4.38e-02
## 32         Glutamic acid, 3TMS; 8     0.00688  1.86e-04  1.36e-02 4.40e-02
## 33            Dodecanoic acid; 54     0.00687  1.81e-04  1.36e-02 4.41e-02
```

```
## 34          Isoleucine, 2TMS; 18    -0.00676 -1.34e-02 -6.36e-05 4.79e-02
## 35            Tartronic acid; 73     0.00668 -9.95e-06  1.34e-02 5.03e-02
## 36           Oleic acid, TMS; 3     0.00647 -2.25e-04  1.32e-02 5.82e-02
## 37        Benzeneacetic acid; 47     0.00640 -2.90e-04  1.31e-02 6.08e-02
## 38             Hydroxylamine; 62    -0.00626 -1.30e-02  4.32e-04 6.67e-02
## 39               1-Dodecanol; 36    -0.00621 -1.29e-02  4.85e-04 6.91e-02
## 40          Nonadecanoic acid; 66     0.00608 -6.10e-04  1.28e-02 7.49e-02
## 41                  Tyrosine; 75     0.00606 -6.32e-04  1.28e-02 7.59e-02
## 42        Heptadecanoic acid; 60     0.00590 -7.93e-04  1.26e-02 8.41e-02
## 43          Palmitic acid, TMS; 5     0.00585 -8.45e-04  1.25e-02 8.68e-02
## 44            L-5-Oxoproline; 63     0.00578 -9.14e-04  1.25e-02 9.06e-02
##       p.adj
## 1  3.90e-09
## 2  3.23e-05
## 3  4.76e-05
## 4  4.14e-04
## 5  5.48e-04
## 6  1.01e-03
## 7  1.08e-03
## 8  7.14e-03
## 9  7.60e-03
## 10 9.70e-03
## 11 1.35e-02
## 12 1.35e-02
## 13 1.35e-02
## 14 1.35e-02
## 15 1.35e-02
## 16 1.35e-02
## 17 1.35e-02
## 18 1.35e-02
## 19 1.58e-02
## 20 1.79e-02
## 21 2.27e-02
## 22 3.17e-02
## 23 4.20e-02
## 24 4.36e-02
## 25 4.50e-02
## 26 8.03e-02
## 27 8.53e-02
## 28 8.53e-02
## 29 8.66e-02
## 30 9.67e-02
## 31 1.00e-01
## 32 1.00e-01
## 33 1.00e-01
## 34 1.06e-01
## 35 1.08e-01
## 36 1.21e-01
## 37 1.23e-01
## 38 1.32e-01
## 39 1.33e-01
## 40 1.39e-01
## 41 1.39e-01
## 42 1.50e-01
```

```
## 43 1.51e-01
## 44 1.54e-01
## [1] ""
## [1] "Table: Gender.x"
## [1] "  (from model: "
## [1] "   ~ Amputation.at.DATE + Age.x + Gender.x + Hba1c_baseline"
## [1] "     + CALSBP + bmi + Smoking + Statin + log_Blood_TGA +"
## [1] "     Total_cholesterol)"
## [1] ""
##                             Name Coefficient     CI.L     CI.R  p.value
## 1           Citric acid, 4TMS; 6      -0.401 -0.55800 -0.24300 5.97e-07
## 2           Methionine, 2TMS; 16       0.389  0.23200  0.54700 1.25e-06
## 3               Valine, 2TMS; 20       0.381  0.22400  0.53800 2.09e-06
## 4              Proline, 2TMS; 21       0.372  0.21400  0.52900 3.66e-06
## 5           Myristoleic acid; 65      -0.365 -0.52200 -0.20700 5.55e-06
## 6              Leucine, 2TMS; 19       0.359  0.20200  0.51600 7.80e-06
## 7              Glycine, 3TMS; 17      -0.349 -0.50600 -0.19200 1.37e-05
## 8           Isoleucine, 2TMS; 18       0.346  0.18900  0.50400 1.62e-05
## 9             Tartronic acid; 73      -0.342 -0.50000 -0.18500 2.00e-05
## 10           Dodecanoic acid; 54      -0.336 -0.49300 -0.17800 2.89e-05
## 11             Glyceric acid; 30      -0.332 -0.48900 -0.17500 3.55e-05
## 12      4-Deoxytetronic acid; 33       0.306  0.14900  0.46300 1.39e-04
## 13            Oleic acid, TMS; 3      -0.302 -0.45900 -0.14500 1.68e-04
## 14         Aminomalonic acid; 45      -0.294 -0.45100 -0.13600 2.53e-04
## 15 2-hydroxy Isovaleric acid; 38       0.289  0.13200  0.44700 3.14e-04
## 16          Cholesterol, TMS; 23      -0.280 -0.43700 -0.12200 4.97e-04
## 17           Myo inositol 6TMS; 1      -0.266 -0.42300 -0.10800 9.31e-04
## 18        Glutamic acid, 3TMS; 8       0.265  0.10700  0.42200 9.83e-04
## 19          Tridecanoic acid; 74      -0.261 -0.41800 -0.10300 1.18e-03
## 20        Heptadecanoic acid; 60      -0.258 -0.41600 -0.10100 1.29e-03
## 21      Docosahexaenoic acid; 53      -0.257 -0.41400 -0.09930 1.39e-03
## 22             Decanoic acid; 52      -0.256 -0.41400 -0.09900 1.41e-03
## 23        Succinic acid, 2TMS; 7      -0.254 -0.41100 -0.09670 1.56e-03
## 24        Heptadecanoic acid; 61      -0.251 -0.40800 -0.09350 1.78e-03
## 25         Nonadecanoic acid; 66      -0.223 -0.38100 -0.06610 5.38e-03
## 26         Palmitic acid, TMS; 5      -0.212 -0.36900 -0.05470 8.25e-03
## 27          Stearic acid, TMS; 2      -0.209 -0.36600 -0.05150 9.29e-03
## 28        Benzeneacetic acid; 47      -0.198 -0.35500 -0.04080 1.36e-02
## 29            Hydroxyproline; 64       0.176  0.01840  0.33300 2.86e-02
## 30              Ribonic acid; 72      -0.175 -0.33200 -0.01770 2.92e-02
## 31        11-Eicosenoic acid; 35      -0.173 -0.33100 -0.01600 3.08e-02
## 32            1-Monopalmitin; 37       0.173  0.01560  0.33000 3.12e-02
## 33             Nonanoic acid; 67      -0.161 -0.31800 -0.00324 4.55e-02
## 34                  Tyrosine; 75      -0.156 -0.31300  0.00147 5.22e-02
## 35              Pyruvic acid; 31       0.150 -0.00692  0.30800 6.10e-02
## 36                  Glycerol; 57      -0.145 -0.30300  0.01200 7.03e-02
## 37     Arachidonic acid, TMS; 24      -0.144 -0.30100  0.01360 7.34e-02
##       p.adj
## 1  4.47e-05
## 2  4.68e-05
## 3  5.24e-05
## 4  6.87e-05
## 5  8.32e-05
## 6  9.75e-05
```

```
## 7  1.47e-04
## 8  1.52e-04
## 9  1.66e-04
## 10 2.17e-04
## 11 2.42e-04
## 12 8.70e-04
## 13 9.68e-04
## 14 1.36e-03
## 15 1.57e-03
## 16 2.33e-03
## 17 4.10e-03
## 18 4.10e-03
## 19 4.64e-03
## 20 4.80e-03
## 21 4.80e-03
## 22 4.80e-03
## 23 5.08e-03
## 24 5.57e-03
## 25 1.61e-02
## 26 2.38e-02
## 27 2.58e-02
## 28 3.64e-02
## 29 7.30e-02
## 30 7.30e-02
## 31 7.31e-02
## 32 7.31e-02
## 33 1.03e-01
## 34 1.15e-01
## 35 1.31e-01
## 36 1.46e-01
## 37 1.49e-01
## [1] ""
## [1] "Table: Hba1c_baseline"
## [1] "  (from model: "
## [1] "   ~ Amputation.at.DATE + Age.x + Gender.x + Hba1c_baseline"
## [1] "     + CALSBP + bmi + Smoking + Statin + log_Blood_TGA +"
## [1] "     Total_cholesterol)"
## [1] ""
##                          Name Coefficient     CI.L     CI.R  p.value  p.adj
## 1        Tridecanoic acid; 74     -0.1280 -0.19900 -0.05670 0.000427 0.0163
## 2         Arabinopyranose; 51      0.1280  0.05660  0.19900 0.000434 0.0163
## 3   Eicosapentaenoic acid; 55     -0.1220 -0.19300 -0.05120 0.000753 0.0188
## 4            Ethanolamine; 56      0.1160  0.04490  0.18700 0.001390 0.0221
## 5    Docosahexaenoic acid; 53     -0.1140 -0.18500 -0.04240 0.001750 0.0221
## 6           Glyceric acid; 30     -0.1130 -0.18500 -0.04230 0.001770 0.0221
## 7            Valine, 2TMS; 20      0.1110  0.03980  0.18200 0.002240 0.0240
## 8           Alanine, 2TMS; 25      0.1060  0.03470  0.17700 0.003530 0.0331
## 9           Decanoic acid; 52     -0.0949 -0.16600 -0.02380 0.008930 0.0745
## 10 4-Hydroxybutanoic acid; 43     -0.0917 -0.16300 -0.02060 0.011500 0.0861
## 11     11-Eicosenoic acid; 35     -0.0831 -0.15400 -0.01200 0.022100 0.1500
## 12            Lactic acid; 29      0.0795  0.00836  0.15100 0.028500 0.1780
## 13       Myristoleic acid; 65     -0.0771 -0.14800 -0.00599 0.033600 0.1940
## 14         Tartronic acid; 73     -0.0750 -0.14600 -0.00388 0.038800 0.2000
## 15          Leucine, 2TMS; 19      0.0746  0.00344  0.14600 0.039900 0.2000
```

```
## [1] ""
## [1] "Table: CALSBP"
## [1] "  (from model: "
## [1] "   ~ Amputation.at.DATE + Age.x + Gender.x + Hba1c_baseline"
## [1] "     + CALSBP + bmi + Smoking + Statin + log_Blood_TGA +"
## [1] "     Total_cholesterol)"
## [1] ""
##                   Name Coefficient    CI.L   CI.R  p.value  p.adj
## 1 Myristoleic acid; 65     0.00805 0.00338 0.0127 0.000719 0.0539
## [1] ""
## [1] "Table: bmi"
## [1] "  (from model: "
## [1] "   ~ Amputation.at.DATE + Age.x + Gender.x + Hba1c_baseline"
## [1] "     + CALSBP + bmi + Smoking + Statin + log_Blood_TGA +"
## [1] "     Total_cholesterol)"
## [1] ""
##                               Name Coefficient      CI.L      CI.R  p.value
## 1          Glutamic acid, 3TMS; 8      0.0452  0.024700  0.065700 1.58e-05
## 2  2-Hydroxybutyric acid, 2TMS; 2      0.0401  0.019500  0.060600 1.31e-04
## 3                  Campesterol; 49     -0.0392 -0.059700 -0.018700 1.82e-04
## 4                Pyruvic acid; 31     -0.0305 -0.051000 -0.009940 3.62e-03
## 5               Decanoic acid; 52     -0.0300 -0.050600 -0.009500 4.14e-03
## 6                 Lactic acid; 29      0.0296  0.009100  0.050200 4.67e-03
## 7              Arachidic acid; 46     -0.0294 -0.050000 -0.008910 4.94e-03
## 8             1,3-Propanediol; 34     -0.0292 -0.049700 -0.008630 5.37e-03
## 9            alpha-Tocopherol; 26     -0.0266 -0.047200 -0.006110 1.10e-02
## 10          Pyroglutamic acid; 69     -0.0240 -0.044500 -0.003490 2.18e-02
## 11                    Ribitol; 70      0.0240  0.003430  0.044500 2.22e-02
## 12 2,4-Dihydroxybutanoic acid; 28     -0.0238 -0.044400 -0.003310 2.28e-02
## 13 alpha-ketoglutaric acid, TMS M     -0.0226 -0.043100 -0.002080 3.09e-02
## 14          Citric acid, 4TMS; 6     -0.0215 -0.042000 -0.000972 4.01e-02
## 15         11-Eicosenoic acid; 35      0.0211  0.000569  0.041600 4.40e-02
## 16             Tartronic acid; 73     -0.0210 -0.041500 -0.000484 4.48e-02
## 17          Nonadecanoic acid; 66     -0.0210 -0.041500 -0.000469 4.50e-02
##      p.adj
## 1  0.00118
## 2  0.00455
## 3  0.00455
## 4  0.05040
## 5  0.05040
## 6  0.05040
## 7  0.05040
## 8  0.05040
## 9  0.09160
## 10 0.14300
## 11 0.14300
## 12 0.14300
## 13 0.17800
## 14 0.19900
## 15 0.19900
## 16 0.19900
## 17 0.19900
## [1] ""
## [1] "Table: Smoking"
```

```
## [1] "  (from model: "
## [1] "   ~ Amputation.at.DATE + Age.x + Gender.x + Hba1c_baseline"
## [1] "     + CALSBP + bmi + Smoking + Statin + log_Blood_TGA +"
## [1] "     Total_cholesterol)"
## [1] ""
##                               Name Coefficient    CI.L     CI.R  p.value   p.adj
## 1          Glutamic acid, 3TMS; 8       0.372  0.1790  0.56500 0.000159 0.00571
## 2      3-Indolepropionic acid; 41      -0.369 -0.5620 -0.17600 0.000177 0.00571
## 3        Docosahexaenoic acid; 53      -0.362 -0.5550 -0.16900 0.000241 0.00571
## 4              Tartronic acid; 73      -0.356 -0.5490 -0.16300 0.000305 0.00571
## 5               Glyceric acid; 30      -0.330 -0.5230 -0.13700 0.000817 0.01220
## 6            Citric acid, 4TMS; 6      -0.308 -0.5010 -0.11500 0.001760 0.02200
## 7            alpha-Tocopherol; 26      -0.297 -0.4900 -0.10400 0.002560 0.02740
## 8          Benzeneacetic acid; 47      -0.283 -0.4760 -0.08970 0.004100 0.03840
## 9                Valine, 2TMS; 20      -0.276 -0.4690 -0.08270 0.005120 0.04260
## 10                Campesterol; 49      -0.272 -0.4650 -0.07880 0.005780 0.04340
## 11           Malic acid, 3TMS; 11      -0.258 -0.4510 -0.06540 0.008690 0.05930
## 12               Ribonic acid; 72      -0.255 -0.4480 -0.06190 0.009640 0.06030
## 13        3-Indoleacetic acid; 40      -0.234 -0.4270 -0.04080 0.017600 0.10200
## 14 4-Hydroxyphenyllactic acid; 44      -0.218 -0.4110 -0.02530 0.026600 0.13600
## 15              Leucine, 2TMS; 19      -0.218 -0.4110 -0.02450 0.027200 0.13600
## 16                    Ribitol; 70       0.210  0.0175  0.40300 0.032600 0.15300
## 17         Heptadecanoic acid; 60      -0.202 -0.3950 -0.00851 0.040700 0.18000
## [1] ""
## [1] "Table: Statin"
## [1] "  (from model: "
## [1] "   ~ Amputation.at.DATE + Age.x + Gender.x + Hba1c_baseline"
## [1] "     + CALSBP + bmi + Smoking + Statin + log_Blood_TGA +"
## [1] "     Total_cholesterol)"
## [1] ""
##                          Name Coefficient    CI.L    CI.R  p.value  p.adj
## 1         L-5-Oxoproline; 63      -0.334 -0.5070 -0.1610 0.000156 0.0117
## 2            Campesterol; 49       0.252  0.0794  0.4250 0.004250 0.1590
## 3       Cholesterol, TMS; 23      -0.222 -0.3950 -0.0485 0.012100 0.1770
## 4     Glutamic acid, 3TMS; 8       0.212  0.0391  0.3850 0.016300 0.1770
## 5  Arachidonic acid, TMS; 24       0.211  0.0376  0.3840 0.017000 0.1770
## 6         Arachidic acid; 46      -0.208 -0.3810 -0.0351 0.018400 0.1770
## 7     Heptadecanoic acid; 61      -0.205 -0.3780 -0.0320 0.020200 0.1770
## 8      Aminomalonic acid; 45      -0.205 -0.3780 -0.0317 0.020400 0.1770
## 9        Dodecanoic acid; 54      -0.201 -0.3740 -0.0283 0.022600 0.1770
## 10               Ribitol; 71       0.197  0.0244  0.3700 0.025300 0.1770
## 11     Linoleic acid, TMS; 4      -0.197 -0.3700 -0.0236 0.025900 0.1770
## 12               Ribitol; 70       0.193  0.0199  0.3660 0.028800 0.1800
## [1] ""
## [1] "Table: log_Blood_TGA"
## [1] "  (from model: "
## [1] "   ~ Amputation.at.DATE + Age.x + Gender.x + Hba1c_baseline"
## [1] "     + CALSBP + bmi + Smoking + Statin + log_Blood_TGA +"
## [1] "     Total_cholesterol)"
## [1] ""
##                               Name Coefficient     CI.L     CI.R  p.value
## 1             Dodecanoic acid; 54       0.273  0.14400  0.40200 3.44e-05
## 2           Palmitic acid, TMS; 5       0.271  0.14200  0.40000 3.70e-05
## 3               Decanoic acid; 52       0.252  0.12300  0.38100 1.31e-04
```

```
## 4              Stearic acid, TMS; 2       0.250  0.12100  0.37900 1.48e-04
## 5                Arachidic acid; 46       0.247  0.11800  0.37600 1.70e-04
## 6                 Octanoic acid; 68       0.235  0.10600  0.36400 3.53e-04
## 7  3,4-Dihydroxybutanoic acid; 27       0.225  0.09620  0.35400 6.23e-04
## 8            Glyceryl-glycoside; 59       0.218  0.08950  0.34700 8.98e-04
## 9                  Ribonic acid; 72       0.212  0.08270  0.34100 1.30e-03
## 10               Oleic acid, TMS; 3       0.208  0.07940  0.33700 1.55e-03
## 11 4-Hydroxybenzeneacetic acid; 4       0.208  0.07870  0.33700 1.60e-03
## 12              Arabinopyranose; 51       0.194  0.06550  0.32300 3.12e-03
## 13                      Ribitol; 71       0.191  0.06220  0.32000 3.67e-03
## 14          Heptadecanoic acid; 60       0.181  0.05250  0.31000 5.82e-03
## 15 2-Hydroxybutyric acid, 2TMS; 2       0.181  0.05160  0.31000 6.06e-03
## 16                  Lactic acid; 29       0.171  0.04160  0.30000 9.54e-03
## 17 2,4-Dihydroxybutanoic acid; 28       0.167  0.03780  0.29600 1.13e-02
## 18          Glutamic acid, 3TMS; 8       0.166  0.03710  0.29500 1.16e-02
## 19          Heptadecanoic acid; 61       0.165  0.03600  0.29400 1.22e-02
## 20            Fumaric acid, 2TMS; 9       0.163  0.03370  0.29200 1.34e-02
## 21                 Valine, 2TMS; 20       0.160  0.03150  0.28900 1.48e-02
## 22                 Pyruvic acid; 31       0.159  0.03040  0.28800 1.54e-02
## 23            Isoleucine, 2TMS; 18       0.159  0.02970  0.28800 1.59e-02
## 24           Aminomalonic acid; 45      -0.154 -0.28300 -0.02550 1.89e-02
## 25            Myristoleic acid; 65       0.153  0.02450  0.28200 1.97e-02
## 26      4-Hydroxybutanoic acid; 43       0.153  0.02430  0.28200 1.99e-02
## 27 alpha-ketoglutaric acid, TMS M       0.152  0.02340  0.28100 2.06e-02
## 28        4-Deoxytetronic acid; 33       0.151  0.02170  0.28000 2.20e-02
## 29             Myo inositol 6TMS; 1       0.149  0.01960  0.27700 2.40e-02
## 30             Malic acid, 3TMS; 11       0.148  0.01880  0.27700 2.47e-02
## 31                Leucine, 2TMS; 19       0.141  0.01240  0.27000 3.17e-02
## 32      3-Indolepropionic acid; 41      -0.139 -0.26800 -0.00987 3.49e-02
## 33                Proline, 2TMS; 21       0.138  0.00901  0.26700 3.60e-02
## 34                Glyceric acid; 30      -0.136 -0.26500 -0.00727 3.84e-02
## 35       Arachidonic acid, TMS; 24       0.134  0.00531  0.26300 4.13e-02
## 36          Succinic acid, 2TMS; 7       0.132  0.00274  0.26100 4.53e-02
## 37           Nonadecanoic acid; 66       0.123 -0.00623  0.25200 6.21e-02
## 38              Tartronic acid; 73      -0.122 -0.25100  0.00716 6.42e-02
## 39             1,3-Propanediol; 34       0.116 -0.01310  0.24500 7.82e-02
## 40 3-Hydroxybutyric acid, 2TMS; 1       0.114 -0.01470  0.24300 8.24e-02
## 41             Tridecanoic acid; 74       0.112 -0.01700  0.24100 8.90e-02
## 42                Serine, 3TMS; 14      -0.112 -0.24100  0.01710 8.90e-02
## 43          3-Indoleacetic acid; 40       0.111 -0.01770  0.24000 9.08e-02
## 44                Nonanoic acid; 67       0.105 -0.02360  0.23400 1.09e-01
##      p.adj
## 1  0.00139
## 2  0.00139
## 3  0.00254
## 4  0.00254
## 5  0.00254
## 6  0.00442
## 7  0.00668
## 8  0.00842
## 9  0.01080
## 10 0.01090
## 11 0.01090
## 12 0.01950
```

```
## 13 0.02120
## 14 0.03030
## 15 0.03030
## 16 0.04470
## 17 0.04800
## 18 0.04800
## 19 0.04800
## 20 0.05040
## 21 0.05190
## 22 0.05190
## 23 0.05190
## 24 0.05730
## 25 0.05730
## 26 0.05730
## 27 0.05730
## 28 0.05910
## 29 0.06170
## 30 0.06170
## 31 0.07670
## 32 0.08170
## 33 0.08180
## 34 0.08470
## 35 0.08850
## 36 0.09450
## 37 0.12600
## 38 0.12700
## 39 0.15000
## 40 0.15400
## 41 0.15800
## 42 0.15800
## 43 0.15800
## 44 0.18600
## [1] ""
## [1] "Table: Total_cholesterol"
## [1] "  (from model: "
## [1] "    ~ Amputation.at.DATE + Age.x + Gender.x + Hba1c_baseline"
## [1] "      + CALSBP + bmi + Smoking + Statin + log_Blood_TGA +"
## [1] "      Total_cholesterol)"
## [1] ""
##                               Name Coefficient     CI.L     CI.R  p.value
## 1            Cholesterol, TMS; 23      0.4590  0.36200  0.55500 1.02e-20
## 2                 Campesterol; 49      0.3640  0.26800  0.46000 1.23e-13
## 3            alpha-Tocopherol; 26      0.3230  0.22700  0.41900 4.95e-11
## 4          Benzeneacetic acid; 47     -0.1950 -0.29100 -0.09830 7.46e-05
## 5           Linoleic acid, TMS; 4      0.1800  0.08400  0.27700 2.42e-04
## 6      4-Hydroxybutanoic acid; 43     -0.1800 -0.27600 -0.08330 2.56e-04
## 7               Proline, 2TMS; 21     -0.1770 -0.27300 -0.08040 3.23e-04
## 8               Glycine, 3TMS; 17     -0.1700 -0.26600 -0.07370 5.39e-04
## 9  2,4-Dihydroxybutanoic acid; 28     -0.1670 -0.26300 -0.07070 6.73e-04
## 10              L-5-Oxoproline; 63     -0.1660 -0.26200 -0.06970 7.30e-04
## 11           Isoleucine, 2TMS; 18     -0.1510 -0.24700 -0.05470 2.11e-03
## 12 4-Hydroxybenzeneacetic acid; 4     -0.1510 -0.24700 -0.05460 2.14e-03
## 13                   Tyrosine; 75     -0.1480 -0.24400 -0.05190 2.56e-03
## 14             Arabinopyranose; 51     -0.1460 -0.24300 -0.05010 2.89e-03
```

```
## 15      Eicosapentaenoic acid; 55      0.1460  0.04990  0.24200 2.92e-03
## 16       Docosahexaenoic acid; 53      0.1460  0.04950  0.24200 3.00e-03
## 17         Glyceryl-glycoside; 59     -0.1440 -0.24100 -0.04820 3.26e-03
## 18            Threonine, 3TMS; 12     -0.1410 -0.23700 -0.04460 4.14e-03
## 19           Methionine, 2TMS; 16     -0.1410 -0.23700 -0.04450 4.16e-03
## 20              Alanine, 2TMS; 25     -0.1390 -0.23500 -0.04300 4.58e-03
## 21                    Ribitol; 71     -0.1370 -0.23400 -0.04120 5.12e-03
## 22        2-Palmitoylglycerol; 39      0.1340  0.03740  0.23000 6.49e-03
## 23        3-Indoleacetic acid; 40     -0.1300 -0.22600 -0.03340 8.29e-03
## 24              Hydroxylamine; 62     -0.1210 -0.21700 -0.02440 1.40e-02
## 25               Ribonic acid; 72     -0.1190 -0.21500 -0.02290 1.52e-02
## 26               Serine, 3TMS; 14     -0.1170 -0.21300 -0.02090 1.70e-02
## 27        Phenylalanine, 2TMS; 13     -0.1150 -0.21100 -0.01890 1.90e-02
## 28 3,4-Dihydroxybutanoic acid; 27     -0.1130 -0.20900 -0.01680 2.14e-02
## 29           Malic acid, 3TMS; 11     -0.1010 -0.19700 -0.00444 4.03e-02
## 30               Pyruvic acid; 31     -0.1000 -0.19600 -0.00391 4.14e-02
## 31          Palmitic acid, TMS; 5      0.0994  0.00314  0.19600 4.30e-02
## 32 2-Hydroxybutyric acid, 2TMS; 2      0.0913 -0.00497  0.18800 6.31e-02
## 33           Myo inositol 6TMS; 1     -0.0889 -0.18500  0.00739 7.04e-02
## 34            1,3-Propanediol; 34     -0.0825 -0.17900  0.01370 9.29e-02
## 35     3-Indolepropionic acid; 41      0.0821 -0.01410  0.17800 9.44e-02
## 36              Leucine, 2TMS; 19     -0.0820 -0.17800  0.01430 9.51e-02
##       p.adj
## 1  7.65e-19
## 2  4.63e-12
## 3  1.24e-09
## 4  1.40e-03
## 5  3.20e-03
## 6  3.20e-03
## 7  3.46e-03
## 8  5.05e-03
## 9  5.48e-03
## 10 5.48e-03
## 11 1.34e-02
## 12 1.34e-02
## 13 1.41e-02
## 14 1.41e-02
## 15 1.41e-02
## 16 1.41e-02
## 17 1.44e-02
## 18 1.64e-02
## 19 1.64e-02
## 20 1.72e-02
## 21 1.83e-02
## 22 2.21e-02
## 23 2.70e-02
## 24 4.37e-02
## 25 4.57e-02
## 26 4.92e-02
## 27 5.29e-02
## 28 5.72e-02
## 29 1.03e-01
## 30 1.03e-01
## 31 1.04e-01
```

```
## 32 1.48e-01
## 33 1.60e-01
## 34 1.98e-01
## 35 1.98e-01
## 36 1.98e-01
```

###### 5.1.2.3 Table with All Metabolites

```
## [1] ""
## [1] "Table: Amputation.at.DATEJA"
## [1] "  (from model: "
## [1] "   ~ Amputation.at.DATE + Age.x + Gender.x + Hba1c_baseline"
## [1] "     + CALSBP + bmi + Smoking + Statin + log_Blood_TGA +"
## [1] "     Total_cholesterol)"
## [1] ""
##                              Name Coefficient     CI.L     CI.R p.value  p.adj
## 1                Ribonic acid; 72     0.72500  0.30400  1.15000 0.00073 0.0547
## 2             Myo inositol 6TMS; 1     0.61300  0.19300  1.03000 0.00427 0.1600
## 3          Succinic acid, 2TMS; 7     0.57900  0.15800  0.99900 0.00702 0.1760
## 4             Threonine, 3TMS; 12    -0.53500 -0.95600 -0.11500 0.01260 0.2230
## 5  4-Hydroxybenzeneacetic acid; 4     0.52300  0.10200  0.94300 0.01490 0.2230
## 6          Glutamic acid, 3TMS; 8    -0.47300 -0.89400 -0.05280 0.02740 0.2790
## 7                  Creatinine; 50     0.46200  0.04150  0.88300 0.03130 0.2790
## 8                    Tyrosine; 75    -0.45400 -0.87500 -0.03350 0.03430 0.2790
## 9                     Ribitol; 71     0.45300  0.03270  0.87400 0.03470 0.2790
## 10          Methionine, 2TMS; 16    -0.43700 -0.85700 -0.01600 0.04190 0.2790
## 11                   Glycerol; 57     0.43400  0.01350  0.85500 0.04310 0.2790
## 12           Pyroglutamic acid; 69     0.42900  0.00845  0.85000 0.04560 0.2790
## 13       Phenylalanine, 2TMS; 13    -0.42400 -0.84400 -0.00314 0.04830 0.2790
## 14 2,4-Dihydroxybutanoic acid; 28     0.38800 -0.03270  0.80900 0.07070 0.3550
## 15              Hydroxyproline; 64     0.38700 -0.03320  0.80800 0.07100 0.3550
## 16 3,4-Dihydroxybutanoic acid; 27     0.37900 -0.04200  0.79900 0.07770 0.3640
## 17                Valine, 2TMS; 20    -0.37100 -0.79100  0.05010 0.08420 0.3690
## 18 4-Hydroxyphenyllactic acid; 44     0.36500 -0.05520  0.78600 0.08860 0.3690
## 19               Leucine, 2TMS; 19    -0.35900 -0.78000  0.06130 0.09410 0.3710
## 20            Malic acid, 3TMS; 11     0.35100 -0.06950  0.77200 0.10200 0.3820
## 21          Heptadecanoic acid; 61     0.30600 -0.11500  0.72700 0.15400 0.5150
## 22                Serine, 3TMS; 14    -0.30500 -0.72600  0.11600 0.15500 0.5150
## 23               Proline, 2TMS; 21    -0.30300 -0.72400  0.11800 0.15800 0.5150
## 24        4-Deoxytetronic acid; 33     0.28700 -0.13400  0.70800 0.18100 0.5390
## 25        4-Deoxytetronic acid; 32     0.28600 -0.13500  0.70700 0.18300 0.5390
## 26  2-hydroxy Isovaleric acid; 38    -0.28300 -0.70400  0.13700 0.18700 0.5390
## 27     4-Hydroxybutanoic acid; 43     0.27300 -0.14800  0.69400 0.20300 0.5600
## 28             Arabinopyranose; 51    -0.26600 -0.68700  0.15400 0.21500 0.5600
## 29     3-Indolepropionic acid; 41    -0.26500 -0.68600  0.15500 0.21700 0.5600
## 30            Myristoleic acid; 65     0.25900 -0.16200  0.68000 0.22800 0.5600
## 31             Dodecanoic acid; 54     0.25700 -0.16400  0.67800 0.23100 0.5600
## 32                 Campesterol; 49    -0.23600 -0.65700  0.18400 0.27100 0.6340
## 33            Stearic acid, TMS; 2     0.22900 -0.19200  0.64900 0.28700 0.6510
## 34               Alanine, 2TMS; 25     0.21600 -0.20500  0.63600 0.31500 0.6950
## 35        2-Palmitoylglycerol; 39    -0.20800 -0.62900  0.21200 0.33200 0.7120
## 36           Aminomalonic acid; 45    -0.19400 -0.61400  0.22700 0.36700 0.7360
## 37          Heptadecanoic acid; 60     0.19200 -0.22800  0.61300 0.37000 0.7360
## 38          Glyceryl-glycoside; 59     0.18900 -0.23100  0.61000 0.37800 0.7360
## 39      Eicosapentaenoic acid; 55    -0.18700 -0.60800  0.23300 0.38300 0.7360
## 40               Hydroxylamine; 62     0.16600 -0.25400  0.58700 0.43800 0.8140
## 41            Tridecanoic acid; 74     0.16000 -0.26100  0.58000 0.45700 0.8140
## 42          11-Eicosenoic acid; 35     0.15900 -0.26200  0.58000 0.45900 0.8140
## 43            Isoleucine, 2TMS; 18    -0.15600 -0.57700  0.26400 0.46700 0.8140
## 44            alpha-Tocopherol; 26    -0.14300 -0.56400  0.27800 0.50500 0.8440
```

```
## 45             Decanoic acid; 52     0.14200 -0.27900  0.56200 0.50900 0.8440
## 46                   Ribitol; 70     0.13900 -0.28200  0.56000 0.51800 0.8440
## 47             Glyceric acid; 30    -0.12900 -0.54900  0.29200 0.54800 0.8750
## 48         Palmitic acid, TMS; 5     0.12500 -0.29600  0.54600 0.56100 0.8760
## 49            1-Monopalmitin; 37     0.11700 -0.30400  0.53700 0.58700 0.8790
## 50       3-Indoleacetic acid; 40     0.11100 -0.31000  0.53100 0.60700 0.8790
## 51            Tartronic acid; 73    -0.11000 -0.53100  0.31000 0.60700 0.8790
## 52            Arachidic acid; 46     0.10900 -0.31200  0.53000 0.61200 0.8790
## 53               1-Dodecanol; 36     0.10600 -0.31500  0.52700 0.62100 0.8790
## 54 2-Hydroxybutyric acid, 2TMS; 2     0.09480 -0.32600  0.51500 0.65900 0.9010
## 55         Fumaric acid, 2TMS; 9     0.09430 -0.32600  0.51500 0.66000 0.9010
## 56               Lactic acid; 29     0.08210 -0.33900  0.50300 0.70200 0.9210
## 57             Octanoic acid; 68     0.07920 -0.34100  0.50000 0.71200 0.9210
## 58 alpha-ketoglutaric acid, TMS M    -0.07920 -0.50000  0.34100 0.71200 0.9210
## 59          Citric acid, 4TMS; 6     0.07570 -0.34500  0.49600 0.72400 0.9210
## 60         Nonadecanoic acid; 66     0.05690 -0.36400  0.47800 0.79100 0.9560
## 61            L-5-Oxoproline; 63    -0.05560 -0.47600  0.36500 0.79600 0.9560
## 62             Nonanoic acid; 67     0.05470 -0.36600  0.47500 0.79900 0.9560
## 63                  Glycerol; 58     0.05240 -0.36800  0.47300 0.80700 0.9560
## 64               Bisphenol A; 48    -0.04820 -0.46900  0.37200 0.82200 0.9560
## 65      Arachidonic acid, TMS; 24     0.03780 -0.38300  0.45800 0.86000 0.9560
## 66             Glycine, 3TMS; 17     0.03440 -0.38600  0.45500 0.87200 0.9560
## 67            Oleic acid, TMS; 3     0.03000 -0.39100  0.45100 0.88900 0.9560
## 68              Ethanolamine; 56     0.02680 -0.39400  0.44700 0.90100 0.9560
## 69 3-Hydroxybutyric acid, 2TMS; 1     0.02420 -0.39600  0.44500 0.91000 0.9560
## 70          1,3-Propanediol; 34     0.02310 -0.39800  0.44400 0.91400 0.9560
## 71        Benzeneacetic acid; 47     0.01990 -0.40100  0.44100 0.92600 0.9560
## 72              Pyruvic acid; 31     0.01910 -0.40200  0.44000 0.92900 0.9560
## 73      Docosahexaenoic acid; 53    -0.01870 -0.43900  0.40200 0.93000 0.9560
## 74         Linoleic acid, TMS; 4     0.01280 -0.40800  0.43300 0.95300 0.9640
## 75          Cholesterol, TMS; 23     0.00965 -0.41100  0.43000 0.96400 0.9640
```

###### 5.1.3 Adjusted Model with eGFR

```
## [1] "Fitting models:"
## [1] "~ Amputation.at.DATE + Age.x + Gender.x + Hba1c_baseline + CALSBP + bmi + Smoking + Statin + lo
## [1] ""
```

###### 5.1.3.1 Forest Plot of Model Coefficients

```
## Warning: Ignoring unknown aesthetics: x
## Ignoring unknown aesthetics: x
## Ignoring unknown aesthetics: x
## Ignoring unknown aesthetics: x
## Ignoring unknown aesthetics: x
## Ignoring unknown aesthetics: x
## Ignoring unknown aesthetics: x
## Ignoring unknown aesthetics: x
## Ignoring unknown aesthetics: x
## Ignoring unknown aesthetics: x
```

```
## NULL
```

###### 5.1.3.2 Tables of Model Coefficients

```
## [1] ""
## [1] "Table: Amputation.at.DATEJA"
## [1] "  (from model: "
## [1] "   ~ Amputation.at.DATE + Age.x + Gender.x + Hba1c_baseline"
## [1] "     + CALSBP + bmi + Smoking + Statin + log_Blood_TGA +"
## [1] "     Total_cholesterol + egfr)"
## [1] ""
## [1] "No significant associations at p.adj < 0.2"
## [1] ""
## [1] "Table: Age.x"
## [1] "  (from model: "
## [1] "   ~ Amputation.at.DATE + Age.x + Gender.x + Hba1c_baseline"
## [1] "     + CALSBP + bmi + Smoking + Statin + log_Blood_TGA +"
## [1] "     Total_cholesterol + egfr)"
## [1] ""
##                               Name Coefficient      CI.L     CI.R  p.value
## 1       Eicosapentaenoic acid; 55     0.02470  1.79e-02 0.031500 1.45e-12
## 2        Docosahexaenoic acid; 53     0.01520  8.38e-03 0.022000 1.33e-05
## 3               Glyceric acid; 30     0.01200  5.22e-03 0.018900 5.50e-04
## 4           alpha-Tocopherol; 26     0.01160  4.74e-03 0.018400 9.00e-04
## 5                Pyruvic acid; 31     0.01150  4.58e-03 0.018400 1.13e-03
## 6               Decanoic acid; 52     0.01140  4.53e-03 0.018300 1.15e-03
## 7  alpha-ketoglutaric acid, TMS M     0.01140  4.47e-03 0.018300 1.28e-03
## 8           Aminomalonic acid; 45     0.01030  3.42e-03 0.017200 3.35e-03
## 9  4-Hydroxybenzeneacetic acid; 4     0.00987  3.10e-03 0.016600 4.28e-03
## 10        3-Indoleacetic acid; 40     0.00984  2.97e-03 0.016700 5.01e-03
## 11                   Tyrosine; 75     0.00917  2.25e-03 0.016100 9.38e-03
## 12 2,4-Dihydroxybutanoic acid; 28     0.00874  2.06e-03 0.015400 1.04e-02
## 13                   Ribitol; 70     0.00901  2.10e-03 0.015900 1.06e-02
## 14          Palmitic acid, TMS; 5     0.00845  1.57e-03 0.015300 1.61e-02
## 15            Malic acid, 3TMS; 11     0.00814  1.23e-03 0.015100 2.10e-02
## 16         11-Eicosenoic acid; 35     0.00816  1.23e-03 0.015100 2.11e-02
## 17            Myristoleic acid; 65     0.00800  1.14e-03 0.014900 2.24e-02
## 18         Succinic acid, 2TMS; 7     0.00797  1.05e-03 0.014900 2.41e-02
## 19         Glutamic acid, 3TMS; 8     0.00759  7.85e-04 0.014400 2.88e-02
## 20         Heptadecanoic acid; 61     0.00765  7.25e-04 0.014600 3.04e-02
## 21          Fumaric acid, 2TMS; 9     0.00741  4.93e-04 0.014300 3.58e-02
## 22             Dodecanoic acid; 54     0.00730  4.23e-04 0.014200 3.75e-02
## 23             Oleic acid, TMS; 3     0.00727  3.72e-04 0.014200 3.89e-02
## 24             Tartronic acid; 73     0.00703  1.73e-04 0.013900 4.45e-02
## 25                   Glycerol; 57     0.00706  1.19e-04 0.014000 4.62e-02
## 26              Alanine, 2TMS; 25     0.00701  9.85e-05 0.013900 4.68e-02
## 27             L-5-Oxoproline; 63     0.00688 -3.89e-05 0.013800 5.13e-02
## 28              Octanoic acid; 68     0.00683 -7.90e-05 0.013700 5.27e-02
## 29              Hydroxylamine; 62    -0.00677 -1.37e-02 0.000172 5.60e-02
## 30              Glycine, 3TMS; 17     0.00662 -2.35e-04 0.013500 5.84e-02
## 31 4-Hydroxyphenyllactic acid; 44     0.00662 -2.74e-04 0.013500 5.98e-02
## 32        Phenylalanine, 2TMS; 13     0.00660 -3.33e-04 0.013500 6.21e-02
## 33        2-Palmitoylglycerol; 39     0.00660 -3.40e-04 0.013500 6.23e-02
## 34             Hydroxyproline; 64    -0.00654 -1.35e-02 0.000363 6.33e-02
## 35           Nonadecanoic acid; 66     0.00654 -3.90e-04 0.013500 6.43e-02
## 36            Methionine, 2TMS; 16     0.00645 -4.26e-04 0.013300 6.60e-02
```

```
## 37          Stearic acid, TMS; 2     0.00617 -7.25e-04 0.013100 7.94e-02
## 38        Heptadecanoic acid; 60     0.00612 -8.02e-04 0.013000 8.31e-02
## 39          Myo inositol 6TMS; 1     0.00557 -1.08e-03 0.012200 1.01e-01
##       p.adj
## 1  1.08e-10
## 2  5.00e-04
## 3  1.37e-02
## 4  1.37e-02
## 5  1.37e-02
## 6  1.37e-02
## 7  1.37e-02
## 8  3.14e-02
## 9  3.57e-02
## 10 3.76e-02
## 11 6.11e-02
## 12 6.11e-02
## 13 6.11e-02
## 14 8.62e-02
## 15 9.87e-02
## 16 9.87e-02
## 17 9.88e-02
## 18 1.00e-01
## 19 1.14e-01
## 20 1.14e-01
## 21 1.27e-01
## 22 1.27e-01
## 23 1.27e-01
## 24 1.35e-01
## 25 1.35e-01
## 26 1.35e-01
## 27 1.37e-01
## 28 1.37e-01
## 29 1.37e-01
## 30 1.37e-01
## 31 1.37e-01
## 32 1.37e-01
## 33 1.37e-01
## 34 1.37e-01
## 35 1.37e-01
## 36 1.37e-01
## 37 1.61e-01
## 38 1.64e-01
## 39 1.94e-01
## [1] ""
## [1] "Table: Gender.x"
## [1] "  (from model: "
## [1] "   ~ Amputation.at.DATE + Age.x + Gender.x + Hba1c_baseline"
## [1] "     + CALSBP + bmi + Smoking + Statin + log_Blood_TGA +"
## [1] "     Total_cholesterol + egfr)"
## [1] ""
##                              Name Coefficient     CI.L     CI.R  p.value
## 1       4-Deoxytetronic acid; 33       0.431  0.27200  0.58900 1.06e-07
## 2           Myristoleic acid; 65      -0.384 -0.54300 -0.22500 2.46e-06
## 3              Glyceric acid; 30      -0.375 -0.53400 -0.21700 3.61e-06
```

```
## 4              Proline, 2TMS; 21       0.374  0.21500  0.53400 4.34e-06
## 5            Tartronic acid; 73      -0.348 -0.50700 -0.18900 1.91e-05
## 6           Dodecanoic acid; 54      -0.348 -0.50800 -0.18800 1.99e-05
## 7           Cholesterol, TMS; 23     -0.321 -0.47700 -0.16600 5.17e-05
## 8           Methionine, 2TMS; 16      0.326  0.16700  0.48600 6.33e-05
## 9             Oleic acid, TMS; 3     -0.319 -0.47900 -0.15900 9.77e-05
## 10          Citric acid, 4TMS; 6     -0.306 -0.46500 -0.14800 1.57e-04
## 11      Docosahexaenoic acid; 53     -0.294 -0.45300 -0.13600 2.79e-04
## 12             Glycine, 3TMS; 17     -0.290 -0.44900 -0.13100 3.59e-04
## 13             Leucine, 2TMS; 19      0.289  0.13000  0.44800 3.66e-04
## 14              Valine, 2TMS; 20      0.282  0.12500  0.44000 4.52e-04
## 15             Decanoic acid; 52     -0.281 -0.44000 -0.12100 5.69e-04
## 16          Stearic acid, TMS; 2     -0.274 -0.43400 -0.11400 8.04e-04
## 17         Aminomalonic acid; 45     -0.267 -0.42600 -0.10800 1.04e-03
## 18          Tridecanoic acid; 74     -0.264 -0.42400 -0.10300 1.31e-03
## 19         Palmitic acid, TMS; 5     -0.261 -0.42100 -0.10200 1.35e-03
## 20        Heptadecanoic acid; 61     -0.262 -0.42300 -0.10100 1.40e-03
## 21        Heptadecanoic acid; 60     -0.258 -0.41900 -0.09730 1.67e-03
## 22        Glutamic acid, 3TMS; 8      0.252  0.09370  0.41000 1.81e-03
## 23            Hydroxyproline; 64      0.252  0.09150  0.41200 2.10e-03
## 24          Isoleucine, 2TMS; 18      0.235  0.07700  0.39300 3.57e-03
## 25 2-hydroxy Isovaleric acid; 38      0.238  0.07740  0.39800 3.70e-03
## 26        Succinic acid, 2TMS; 7     -0.237 -0.39800 -0.07660 3.81e-03
## 27         Nonadecanoic acid; 66     -0.230 -0.39100 -0.06930 5.07e-03
## 28                  Tyrosine; 75     -0.214 -0.37400 -0.05310 9.14e-03
## 29                  Glycerol; 57     -0.198 -0.35900 -0.03680 1.61e-02
## 30            1-Monopalmitin; 37      0.168  0.00644  0.32900 4.15e-02
## 31        11-Eicosenoic acid; 35     -0.166 -0.32700 -0.00523 4.30e-02
## 32     Arachidonic acid, TMS; 24     -0.158 -0.31900  0.00320 5.47e-02
## 33             Nonanoic acid; 67     -0.157 -0.31800  0.00459 5.69e-02
## 34        Benzeneacetic acid; 47     -0.154 -0.31400  0.00649 6.00e-02
##       p.adj
## 1  7.96e-06
## 2  8.14e-05
## 3  8.14e-05
## 4  8.14e-05
## 5  2.48e-04
## 6  2.48e-04
## 7  5.53e-04
## 8  5.93e-04
## 9  8.14e-04
## 10 1.17e-03
## 11 1.90e-03
## 12 2.11e-03
## 13 2.11e-03
## 14 2.42e-03
## 15 2.84e-03
## 16 3.77e-03
## 17 4.58e-03
## 18 5.23e-03
## 19 5.23e-03
## 20 5.23e-03
## 21 5.95e-03
## 22 6.17e-03
```

```
## 23 6.83e-03
## 24 1.10e-02
## 25 1.10e-02
## 26 1.10e-02
## 27 1.41e-02
## 28 2.45e-02
## 29 4.15e-02
## 30 1.04e-01
## 31 1.04e-01
## 32 1.28e-01
## 33 1.29e-01
## 34 1.32e-01
## [1] ""
## [1] "Table: Hba1c_baseline"
## [1] "  (from model: "
## [1] "   ~ Amputation.at.DATE + Age.x + Gender.x + Hba1c_baseline"
## [1] "     + CALSBP + bmi + Smoking + Statin + log_Blood_TGA +"
## [1] "     Total_cholesterol + egfr)"
## [1] ""
##                           Name Coefficient     CI.L     CI.R  p.value  p.adj
## 1   Eicosapentaenoic acid; 55     -0.1300 -0.20000 -0.06040 0.000263 0.0112
## 2        Tridecanoic acid; 74     -0.1320 -0.20300 -0.06030 0.000300 0.0112
## 3         Arabinopyranose; 51      0.1260  0.05540  0.19700 0.000493 0.0123
## 4    Docosahexaenoic acid; 53     -0.1210 -0.19100 -0.05080 0.000743 0.0139
## 5           Glyceric acid; 30     -0.1140 -0.18400 -0.04380 0.001470 0.0209
## 6           Alanine, 2TMS; 25      0.1140  0.04300  0.18500 0.001670 0.0209
## 7            Ethanolamine; 56      0.1120  0.04050  0.18300 0.002130 0.0229
## 8            Valine, 2TMS; 20      0.1030  0.03320  0.17300 0.003850 0.0361
## 9           Decanoic acid; 52     -0.0960 -0.16700 -0.02530 0.007790 0.0650
## 10 4-Hydroxybutanoic acid; 43     -0.0884 -0.16000 -0.01710 0.015100 0.1130
## 11       Citric acid, 4TMS; 6      0.0818  0.01150  0.15200 0.022500 0.1430
## 12     11-Eicosenoic acid; 35     -0.0827 -0.15400 -0.01150 0.022900 0.1430
## 13       Myristoleic acid; 65     -0.0782 -0.14900 -0.00756 0.030000 0.1710
## 14             Lactic acid; 29     0.0778  0.00676  0.14900 0.031900 0.1710
## 15           Octanoic acid; 68     -0.0743 -0.14500 -0.00330 0.040300 0.1850
## 16           Proline, 2TMS; 21      0.0737  0.00309  0.14400 0.040800 0.1850
## 17     Glyceryl-glycoside; 59      0.0735  0.00266  0.14400 0.042000 0.1850
## [1] ""
## [1] "Table: CALSBP"
## [1] "  (from model: "
## [1] "   ~ Amputation.at.DATE + Age.x + Gender.x + Hba1c_baseline"
## [1] "     + CALSBP + bmi + Smoking + Statin + log_Blood_TGA +"
## [1] "     Total_cholesterol + egfr)"
## [1] ""
##                    Name Coefficient    CI.L   CI.R  p.value  p.adj
## 1 Myristoleic acid; 65     0.00824 0.00362 0.0129 0.000479 0.0359
## [1] ""
## [1] "Table: bmi"
## [1] "  (from model: "
## [1] "   ~ Amputation.at.DATE + Age.x + Gender.x + Hba1c_baseline"
## [1] "     + CALSBP + bmi + Smoking + Statin + log_Blood_TGA +"
## [1] "     Total_cholesterol + egfr)"
## [1] ""
##                               Name Coefficient      CI.L      CI.R  p.value
```

```
## 1          Glutamic acid, 3TMS; 8      0.0457  0.025600  0.065900 8.97e-06
## 2  2-Hydroxybutyric acid, 2TMS; 2      0.0408  0.020600  0.060900 7.38e-05
## 3                 Campesterol; 49     -0.0389 -0.059100 -0.018800 1.59e-04
## 4                 Lactic acid; 29      0.0303  0.009860  0.050800 3.70e-03
## 5                Pyruvic acid; 31     -0.0300 -0.050500 -0.009560 4.05e-03
## 6               Decanoic acid; 52     -0.0295 -0.049800 -0.009130 4.54e-03
## 7             1,3-Propanediol; 34     -0.0291 -0.049700 -0.008540 5.57e-03
## 8              Arachidic acid; 46     -0.0289 -0.049300 -0.008360 5.81e-03
## 9            alpha-Tocopherol; 26     -0.0262 -0.046400 -0.006050 1.09e-02
## 10 2,4-Dihydroxybutanoic acid; 28     -0.0255 -0.045200 -0.005670 1.17e-02
## 11          Pyroglutamic acid; 69     -0.0248 -0.045200 -0.004420 1.71e-02
## 12                    Ribitol; 70      0.0228  0.002320  0.043200 2.91e-02
## 13           Citric acid, 4TMS; 6     -0.0223 -0.042500 -0.002090 3.06e-02
## 14           Isoleucine, 2TMS; 18      0.0218  0.001660  0.041900 3.39e-02
## 15 alpha-ketoglutaric acid, TMS M     -0.0221 -0.042600 -0.001550 3.50e-02
## 16             Tartronic acid; 73     -0.0215 -0.041800 -0.001260 3.74e-02
## 17          Nonadecanoic acid; 66     -0.0211 -0.041600 -0.000609 4.36e-02
## 18         11-Eicosenoic acid; 35      0.0211  0.000574  0.041600 4.39e-02
##       p.adj
## 1  0.000673
## 2  0.002770
## 3  0.003980
## 4  0.054500
## 5  0.054500
## 6  0.054500
## 7  0.054500
## 8  0.054500
## 9  0.087800
## 10 0.087800
## 11 0.117000
## 12 0.175000
## 13 0.175000
## 14 0.175000
## 15 0.175000
## 16 0.175000
## 17 0.183000
## 18 0.183000
## [1] ""
## [1] "Table: Smoking"
## [1] "  (from model: "
## [1] "   ~ Amputation.at.DATE + Age.x + Gender.x + Hba1c_baseline"
## [1] "     + CALSBP + bmi + Smoking + Statin + log_Blood_TGA +"
## [1] "     Total_cholesterol + egfr)"
## [1] ""
##                               Name Coefficient    CI.L    CI.R  p.value   p.adj
## 1       Docosahexaenoic acid; 53      -0.381 -0.5710 -0.1910 8.86e-05 0.00408
## 2      3-Indolepropionic acid; 41      -0.378 -0.5700 -0.1860 1.18e-04 0.00408
## 3          Glutamic acid, 3TMS; 8       0.365  0.1760  0.5550 1.63e-04 0.00408
## 4              Tartronic acid; 73      -0.351 -0.5420 -0.1600 3.14e-04 0.00589
## 5               Glyceric acid; 30      -0.344 -0.5340 -0.1540 3.95e-04 0.00593
## 6                Valine, 2TMS; 20      -0.314 -0.5030 -0.1250 1.12e-03 0.01400
## 7            alpha-Tocopherol; 26      -0.306 -0.4960 -0.1170 1.57e-03 0.01690
## 8           Citric acid, 4TMS; 6      -0.268 -0.4590 -0.0780 5.74e-03 0.05040
## 9                 Campesterol; 49      -0.266 -0.4560 -0.0762 6.05e-03 0.05040
```

```
## 10         Benzeneacetic acid; 47      -0.265 -0.4570 -0.0726 6.94e-03 0.05210
## 11              Leucine, 2TMS; 19      -0.244 -0.4340 -0.0530 1.23e-02 0.08380
## 12            Malic acid, 3TMS; 11      -0.239 -0.4320 -0.0468 1.49e-02 0.09290
## 13 3,4-Dihydroxybutanoic acid; 27       0.223  0.0356  0.4110 1.98e-02 0.11400
## 14                    Ribitol; 70       0.218  0.0262  0.4110 2.60e-02 0.13900
## [1] ""
## [1] "Table: Statin"
## [1] "  (from model: "
## [1] "   ~ Amputation.at.DATE + Age.x + Gender.x + Hba1c_baseline"
## [1] "     + CALSBP + bmi + Smoking + Statin + log_Blood_TGA +"
## [1] "     Total_cholesterol + egfr)"
## [1] ""
##                        Name Coefficient    CI.L    CI.R  p.value  p.adj
## 1        L-5-Oxoproline; 63      -0.313 -0.4880 -0.1380 0.000459 0.0344
## 2           Campesterol; 49       0.241  0.0687  0.4130 0.006140 0.1880
## 3    Glutamic acid, 3TMS; 8       0.225  0.0530  0.3970 0.010400 0.1880
## 4     Aminomalonic acid; 45      -0.225 -0.3990 -0.0514 0.011100 0.1880
## 5 Arachidonic acid, TMS; 24       0.223  0.0482  0.3990 0.012500 0.1880
## [1] ""
## [1] "Table: log_Blood_TGA"
## [1] "  (from model: "
## [1] "   ~ Amputation.at.DATE + Age.x + Gender.x + Hba1c_baseline"
## [1] "     + CALSBP + bmi + Smoking + Statin + log_Blood_TGA +"
## [1] "     Total_cholesterol + egfr)"
## [1] ""
##                              Name Coefficient      CI.L      CI.R  p.value
## 1           Palmitic acid, TMS; 5       0.290  0.162000  0.419000 9.63e-06
## 2             Dodecanoic acid; 54       0.278  0.150000  0.407000 2.20e-05
## 3            Stearic acid, TMS; 2       0.275  0.146000  0.404000 2.88e-05
## 4               Octanoic acid; 68       0.262  0.133000  0.391000 6.89e-05
## 5               Decanoic acid; 52       0.261  0.132000  0.389000 7.01e-05
## 6              Arachidic acid; 46       0.249  0.120000  0.378000 1.61e-04
## 7  2-Hydroxybutyric acid, 2TMS; 2       0.226  0.099200  0.353000 4.87e-04
## 8              Oleic acid, TMS; 3       0.215  0.086300  0.344000 1.07e-03
## 9            Isoleucine, 2TMS; 18       0.202  0.075300  0.329000 1.81e-03
## 10              Valine, 2TMS; 20       0.199  0.072600  0.326000 2.06e-03
## 11            Arabinopyranose; 51       0.193  0.064300  0.322000 3.32e-03
## 12         Glyceryl-glycoside; 59       0.187  0.058500  0.315000 4.36e-03
## 13         Heptadecanoic acid; 60       0.180  0.050600  0.309000 6.41e-03
## 14                Lactic acid; 29       0.177  0.048400  0.306000 7.04e-03
## 15         Glutamic acid, 3TMS; 8       0.171  0.043600  0.298000 8.49e-03
## 16              Leucine, 2TMS; 19       0.168  0.040000  0.296000 1.01e-02
## 17         Heptadecanoic acid; 61       0.168  0.039200  0.298000 1.06e-02
## 18 3,4-Dihydroxybutanoic acid; 27       0.164  0.037600  0.289000 1.09e-02
## 19          Aminomalonic acid; 45      -0.165 -0.293000 -0.037000 1.16e-02
## 20               Pyruvic acid; 31       0.165  0.036100  0.294000 1.21e-02
## 21            Myristoleic acid; 65       0.161  0.033200  0.289000 1.37e-02
## 22 4-Hydroxybenzeneacetic acid; 4       0.156  0.030000  0.282000 1.53e-02
## 23     4-Hydroxybutanoic acid; 43       0.156  0.026300  0.285000 1.84e-02
## 24 alpha-ketoglutaric acid, TMS M       0.154  0.024500  0.283000 1.98e-02
## 25               Ribonic acid; 72       0.145  0.020000  0.270000 2.30e-02
## 26           Fumaric acid, 2TMS; 9       0.140  0.011300  0.269000 3.31e-02
## 27      Arachidonic acid, TMS; 24       0.139  0.009740  0.269000 3.51e-02
## 28              Proline, 2TMS; 21       0.135  0.006830  0.263000 3.90e-02
```

```
## 29           Malic acid, 3TMS; 11       0.129 -0.000308  0.258000 5.05e-02
## 30     3-Indolepropionic acid; 41      -0.128 -0.257000  0.000905 5.16e-02
## 31         Succinic acid, 2TMS; 7       0.126 -0.003570  0.255000 5.67e-02
## 32          Nonadecanoic acid; 66       0.124 -0.005350  0.253000 6.02e-02
## 33                    Ribitol; 71       0.119 -0.005960  0.243000 6.20e-02
## 34             Tartronic acid; 73      -0.122 -0.250000  0.006200 6.22e-02
## 35              Glycine, 3TMS; 17      -0.121 -0.249000  0.007080 6.41e-02
## 36              Glyceric acid; 30      -0.119 -0.247000  0.007860 6.59e-02
## 37            1,3-Propanediol; 34       0.117 -0.013000  0.246000 7.77e-02
## 38           Tridecanoic acid; 74       0.114 -0.015600  0.243000 8.49e-02
## 39 3-Hydroxybutyric acid, 2TMS; 1       0.114 -0.015800  0.244000 8.52e-02
##      p.adj
## 1  0.00072
## 2  0.00072
## 3  0.00072
## 4  0.00105
## 5  0.00105
## 6  0.00202
## 7  0.00522
## 8  0.01000
## 9  0.01510
## 10 0.01550
## 11 0.02260
## 12 0.02730
## 13 0.03700
## 14 0.03770
## 15 0.04250
## 16 0.04550
## 17 0.04550
## 18 0.04550
## 19 0.04560
## 20 0.04560
## 21 0.04880
## 22 0.05220
## 23 0.06000
## 24 0.06190
## 25 0.06890
## 26 0.09550
## 27 0.09750
## 28 0.10400
## 29 0.12900
## 30 0.12900
## 31 0.13700
## 32 0.13700
## 33 0.13700
## 34 0.13700
## 35 0.13700
## 36 0.13700
## 37 0.15700
## 38 0.16400
## 39 0.16400
## [1] ""
## [1] "Table: Total_cholesterol"
## [1] "  (from model: "
```

```
## [1] "   ~ Amputation.at.DATE + Age.x + Gender.x + Hba1c_baseline"
## [1] "     + CALSBP + bmi + Smoking + Statin + log_Blood_TGA +"
## [1] "     Total_cholesterol + egfr)"
## [1] ""
##                               Name Coefficient    CI.L      CI.R  p.value
## 1            Cholesterol, TMS; 23      0.4510  0.3580  0.544000 5.10e-21
## 2                 Campesterol; 49      0.3650  0.2700  0.459000 6.48e-14
## 3             alpha-Tocopherol; 26      0.3190  0.2240  0.414000 5.05e-11
## 4           Benzeneacetic acid; 47     -0.1880 -0.2840 -0.092100 1.23e-04
## 5            Linoleic acid, TMS; 4      0.1780  0.0826  0.274000 2.69e-04
## 6       4-Hydroxybutanoic acid; 43     -0.1750 -0.2710 -0.078700 3.72e-04
## 7                Proline, 2TMS; 21     -0.1730 -0.2680 -0.077400 3.88e-04
## 8                L-5-Oxoproline; 63     -0.1710 -0.2670 -0.074900 4.95e-04
## 9             Isoleucine, 2TMS; 18     -0.1680 -0.2620 -0.073400 5.03e-04
## 10               Glycine, 3TMS; 17     -0.1630 -0.2580 -0.067300 8.30e-04
## 11                    Tyrosine; 75     -0.1560 -0.2520 -0.060300 1.43e-03
## 12            Methionine, 2TMS; 16     -0.1500 -0.2450 -0.054200 2.13e-03
## 13 2,4-Dihydroxybutanoic acid; 28     -0.1440 -0.2360 -0.050700 2.45e-03
## 14              Arabinopyranose; 51     -0.1470 -0.2430 -0.051300 2.63e-03
## 15             Threonine, 3TMS; 12     -0.1430 -0.2390 -0.046500 3.65e-03
## 16       Docosahexaenoic acid; 53      0.1380  0.0431  0.233000 4.39e-03
## 17      Eicosapentaenoic acid; 55      0.1360  0.0420  0.231000 4.66e-03
## 18 4-Hydroxybenzeneacetic acid; 4     -0.1330 -0.2270 -0.038800 5.65e-03
## 19               Alanine, 2TMS; 25     -0.1320 -0.2280 -0.036400 6.86e-03
## 20          Glyceryl-glycoside; 59     -0.1320 -0.2270 -0.036000 6.99e-03
## 21        2-Palmitoylglycerol; 39      0.1300  0.0340  0.227000 8.05e-03
## 22                Serine, 3TMS; 14     -0.1280 -0.2230 -0.031700 9.13e-03
## 23               Hydroxylamine; 62     -0.1210 -0.2170 -0.024600 1.39e-02
## 24          3-Indoleacetic acid; 40     -0.1180 -0.2130 -0.022800 1.52e-02
## 25                     Ribitol; 71     -0.1100 -0.2030 -0.017700 1.96e-02
## 26         Phenylalanine, 2TMS; 13     -0.1110 -0.2070 -0.014600 2.40e-02
## 27                Pyruvic acid; 31     -0.1010 -0.1970 -0.005410 3.84e-02
## 28                Ribonic acid; 72     -0.0936 -0.1870 -0.000651 4.84e-02
## 29 3,4-Dihydroxybutanoic acid; 27     -0.0926 -0.1860  0.001050 5.26e-02
## 30             Malic acid, 3TMS; 11     -0.0948 -0.1910  0.001190 5.29e-02
## 31               Leucine, 2TMS; 19     -0.0928 -0.1880  0.002270 5.57e-02
## 32          Palmitic acid, TMS; 5      0.0915 -0.0039  0.187000 6.01e-02
## 33                Valine, 2TMS; 20     -0.0889 -0.1830  0.005320 6.44e-02
## 34                Ethanolamine; 56     -0.0860 -0.1820  0.010300 8.01e-02
##       p.adj
## 1  3.82e-19
## 2  2.43e-12
## 3  1.26e-09
## 4  2.31e-03
## 5  4.03e-03
## 6  4.16e-03
## 7  4.16e-03
## 8  4.19e-03
## 9  4.19e-03
## 10 6.22e-03
## 11 9.78e-03
## 12 1.33e-02
## 13 1.41e-02
## 14 1.41e-02
```

```
## 15 1.83e-02
## 16 2.06e-02
## 17 2.06e-02
## 18 2.35e-02
## 19 2.62e-02
## 20 2.62e-02
## 21 2.87e-02
## 22 3.11e-02
## 23 4.53e-02
## 24 4.75e-02
## 25 5.88e-02
## 26 6.93e-02
## 27 1.07e-01
## 28 1.30e-01
## 29 1.32e-01
## 30 1.32e-01
## 31 1.35e-01
## 32 1.41e-01
## 33 1.46e-01
## 34 1.77e-01
## [1] ""
## [1] "Table: egfr"
## [1] "  (from model: "
## [1] "   ~ Amputation.at.DATE + Age.x + Gender.x + Hba1c_baseline"
## [1] "     + CALSBP + bmi + Smoking + Statin + log_Blood_TGA +"
## [1] "     Total_cholesterol + egfr)"
## [1] ""
##                               Name Coefficient      CI.L      CI.R  p.value
## 1            Myo inositol 6TMS; 1    -0.01620 -0.019200 -0.013300 2.87e-26
## 2                     Ribitol; 71    -0.01570 -0.018700 -0.012800 1.07e-24
## 3                  Creatinine; 50    -0.01520 -0.018200 -0.012200 9.64e-23
## 4  2,4-Dihydroxybutanoic acid; 28    -0.01470 -0.017700 -0.011800 6.69e-22
## 5                Ribonic acid; 72    -0.01440 -0.017400 -0.011400 6.22e-21
## 6  3,4-Dihydroxybutanoic acid; 27    -0.01310 -0.016000 -0.010100 2.37e-17
## 7  4-Hydroxybenzeneacetic acid; 4    -0.01110 -0.014100 -0.008070 6.88e-13
## 8        4-Deoxytetronic acid; 33    -0.01100 -0.014000 -0.007960 1.54e-12
## 9        4-Deoxytetronic acid; 32    -0.01030 -0.013300 -0.007210 5.52e-11
## 10 2-Hydroxybutyric acid, 2TMS; 2     0.00959  0.006570  0.012600 5.84e-10
## 11           Isoleucine, 2TMS; 18     0.00957  0.006550  0.012600 6.12e-10
## 12               Valine, 2TMS; 20     0.00852  0.005510  0.011500 3.29e-08
## 13           Citric acid, 4TMS; 6    -0.00829 -0.011300 -0.005260 9.11e-08
## 14          Pyroglutamic acid; 69    -0.00786 -0.010900 -0.004800 4.95e-07
## 15              Hydroxyproline; 64    -0.00745 -0.010500 -0.004390 2.00e-06
## 16        3-Indoleacetic acid; 40    -0.00711 -0.010200 -0.004060 5.06e-06
## 17         Glyceryl-glycoside; 59    -0.00684 -0.009900 -0.003790 1.19e-05
## 18 4-Hydroxyphenyllactic acid; 44    -0.00611 -0.009170 -0.003060 9.10e-05
## 19               Serine, 3TMS; 14     0.00601  0.002950  0.009080 1.22e-04
## 20              Leucine, 2TMS; 19     0.00593  0.002890  0.008970 1.36e-04
## 21              Octanoic acid; 68     0.00597  0.002910  0.009040 1.37e-04
## 22           Stearic acid, TMS; 2     0.00575  0.002690  0.008810 2.36e-04
## 23           Methionine, 2TMS; 16     0.00521  0.002150  0.008260 8.35e-04
## 24              Glycine, 3TMS; 17    -0.00514 -0.008180 -0.002100 9.44e-04
## 25          Fumaric acid, 2TMS; 9    -0.00497 -0.008040 -0.001900 1.51e-03
## 26                   Tyrosine; 75     0.00484  0.001770  0.007910 2.01e-03
```

```
## 27      Eicosapentaenoic acid; 55     0.00442  0.001400  0.007440 4.10e-03
## 28  2-hydroxy Isovaleric acid; 38     0.00445  0.001380  0.007520 4.53e-03
## 29                  Glycerol; 57     0.00432  0.001240  0.007390 6.02e-03
## 30          Palmitic acid, TMS; 5     0.00425  0.001200  0.007310 6.29e-03
## 31           Cholesterol, TMS; 23     0.00405  0.001080  0.007030 7.52e-03
## 32           Malic acid, 3TMS; 11    -0.00395 -0.007020 -0.000881 1.17e-02
## 33         Benzeneacetic acid; 47    -0.00383 -0.006890 -0.000768 1.43e-02
## 34       Docosahexaenoic acid; 53     0.00370  0.000666  0.006730 1.69e-02
## 35              Glyceric acid; 30     0.00346  0.000428  0.006480 2.53e-02
## 36              Alanine, 2TMS; 25    -0.00335 -0.006410 -0.000283 3.23e-02
## 37                   Glycerol; 58    -0.00318 -0.006270 -0.000084 4.41e-02
##       p.adj
## 1  2.15e-24
## 2  4.00e-23
## 3  2.41e-21
## 4  1.25e-20
## 5  9.33e-20
## 6  2.96e-16
## 7  7.37e-12
## 8  1.44e-11
## 9  4.60e-10
## 10 4.17e-09
## 11 4.17e-09
## 12 2.05e-07
## 13 5.25e-07
## 14 2.65e-06
## 15 1.00e-05
## 16 2.37e-05
## 17 5.24e-05
## 18 3.79e-04
## 19 4.81e-04
## 20 4.89e-04
## 21 4.89e-04
## 22 8.06e-04
## 23 2.72e-03
## 24 2.95e-03
## 25 4.54e-03
## 26 5.79e-03
## 27 1.14e-02
## 28 1.21e-02
## 29 1.56e-02
## 30 1.57e-02
## 31 1.82e-02
## 32 2.74e-02
## 33 3.24e-02
## 34 3.72e-02
## 35 5.42e-02
## 36 6.73e-02
## 37 8.94e-02
```

###### 5.1.3.3 Table with All Metabolites

```
## [1] ""
## [1] "Table: Amputation.at.DATEJA"
## [1] "  (from model: "
## [1] "   ~ Amputation.at.DATE + Age.x + Gender.x + Hba1c_baseline"
## [1] "     + CALSBP + bmi + Smoking + Statin + log_Blood_TGA +"
## [1] "     Total_cholesterol + egfr)"
## [1] ""
##                               Name Coefficient    CI.L    CI.R p.value p.adj
## 1          Succinic acid, 2TMS; 7      0.5490  0.1240  0.9730  0.0113 0.450
## 2                    Glycerol; 57      0.5330  0.1070  0.9590  0.0141 0.450
## 3             Threonine, 3TMS; 12     -0.5130 -0.9380 -0.0880  0.0180 0.450
## 4         Phenylalanine, 2TMS; 13     -0.4800 -0.9060 -0.0545  0.0270 0.507
## 5          Glutamic acid, 3TMS; 8     -0.4460 -0.8640 -0.0287  0.0362 0.543
## 6                Ribonic acid; 72      0.3940 -0.0174  0.8050  0.0605 0.756
## 7            Stearic acid, TMS; 2      0.3580 -0.0647  0.7820  0.0968 0.861
## 8                    Tyrosine; 75     -0.3440 -0.7680  0.0804  0.1120 0.861
## 9               Proline, 2TMS; 21     -0.3240 -0.7450  0.0977  0.1320 0.861
## 10         Heptadecanoic acid; 61      0.3250 -0.0999  0.7490  0.1340 0.861
## 11 2-Hydroxybutyric acid, 2TMS; 2      0.3190 -0.0988  0.7360  0.1340 0.861
## 12           Methionine, 2TMS; 16     -0.3190 -0.7410  0.1020  0.1380 0.861
## 13           Myristoleic acid; 65      0.2980 -0.1240  0.7190  0.1660 0.863
## 14             Dodecanoic acid; 54      0.2860 -0.1360  0.7080  0.1840 0.863
## 15     4-Hydroxybutanoic acid; 43      0.2770 -0.1480  0.7020  0.2020 0.863
## 16 4-Hydroxybenzeneacetic acid; 4      0.2680 -0.1470  0.6830  0.2050 0.863
## 17                Campesterol; 49     -0.2690 -0.6870  0.1500  0.2080 0.863
## 18            Arabinopyranose; 51     -0.2690 -0.6930  0.1540  0.2130 0.863
## 19           Malic acid, 3TMS; 11      0.2580 -0.1660  0.6820  0.2330 0.863
## 20          Aminomalonic acid; 45     -0.2460 -0.6670  0.1760  0.2530 0.863
## 21          Pyroglutamic acid; 69      0.2460 -0.1760  0.6680  0.2540 0.863
## 22             Myo inositol 6TMS; 1      0.2380 -0.1710  0.6460  0.2540 0.863
## 23              Leucine, 2TMS; 19     -0.2260 -0.6460  0.1950  0.2930 0.863
## 24          Palmitic acid, TMS; 5      0.2210 -0.2010  0.6430  0.3040 0.863
## 25              Hydroxyproline; 64      0.2210 -0.2020  0.6450  0.3050 0.863
## 26 4-Hydroxyphenyllactic acid; 44      0.2170 -0.2060  0.6390  0.3150 0.863
## 27              Octanoic acid; 68      0.2160 -0.2080  0.6400  0.3180 0.863
## 28     3-Indolepropionic acid; 41     -0.2140 -0.6370  0.2100  0.3220 0.863
## 29              Decanoic acid; 52      0.1880 -0.2340  0.6100  0.3820 0.940
## 30         Heptadecanoic acid; 60      0.1860 -0.2390  0.6100  0.3920 0.940
## 31  2-hydroxy Isovaleric acid; 38     -0.1810 -0.6060  0.2430  0.4020 0.940
## 32               Valine, 2TMS; 20     -0.1760 -0.5930  0.2400  0.4060 0.940
## 33           Tridecanoic acid; 74      0.1720 -0.2530  0.5970  0.4280 0.940
## 34               Serine, 3TMS; 14     -0.1680 -0.5920  0.2550  0.4360 0.940
## 35        2-Palmitoylglycerol; 39     -0.1570 -0.5830  0.2690  0.4690 0.940
## 36              Hydroxylamine; 62      0.1490 -0.2760  0.5750  0.4920 0.940
## 37         11-Eicosenoic acid; 35      0.1450 -0.2800  0.5700  0.5040 0.940
## 38              Alanine, 2TMS; 25      0.1380 -0.2860  0.5620  0.5220 0.940
## 39             1-Monopalmitin; 37      0.1290 -0.2970  0.5560  0.5520 0.940
## 40             Arachidic acid; 46      0.1200 -0.3050  0.5440  0.5810 0.940
## 41                1-Dodecanol; 36      0.1200 -0.3070  0.5460  0.5820 0.940
## 42                Lactic acid; 29      0.1190 -0.3050  0.5430  0.5820 0.940
## 43           Citric acid, 4TMS; 6     -0.1170 -0.5360  0.3020  0.5840 0.940
## 44              Tartronic acid; 73     -0.1140 -0.5340  0.3060  0.5950 0.940
```

```
## 45                    Creatinine; 50      0.1110 -0.3040  0.5260  0.6000 0.940
## 46              Cholesterol, TMS; 23      0.1030 -0.3080  0.5140  0.6230 0.940
## 47                       Ribitol; 70      0.0993 -0.3240  0.5230  0.6460 0.940
## 48                       Ribitol; 71      0.0928 -0.3170  0.5030  0.6570 0.940
## 49              alpha-Tocopherol; 26     -0.0892 -0.5080  0.3290  0.6760 0.940
## 50         Eicosapentaenoic acid; 55     -0.0880 -0.5050  0.3290  0.6790 0.940
## 51                 Glycine, 3TMS; 17     -0.0863 -0.5070  0.3340  0.6880 0.940
## 52 3,4-Dihydroxybutanoic acid; 27      0.0756 -0.3380  0.4890  0.7200 0.940
## 53 alpha-ketoglutaric acid, TMS M     -0.0685 -0.4940  0.3570  0.7520 0.940
## 54            Benzeneacetic acid; 47     -0.0680 -0.4910  0.3550  0.7530 0.940
## 55          Docosahexaenoic acid; 53      0.0652 -0.3540  0.4840  0.7600 0.940
## 56                  Ethanolamine; 56      0.0659 -0.3600  0.4920  0.7610 0.940
## 57                Oleic acid, TMS; 3      0.0644 -0.3590  0.4880  0.7650 0.940
## 58         Arachidonic acid, TMS; 24      0.0647 -0.3610  0.4900  0.7660 0.940
## 59              Isoleucine, 2TMS; 18      0.0614 -0.3560  0.4790  0.7730 0.940
## 60             Nonadecanoic acid; 66      0.0616 -0.3640  0.4870  0.7760 0.940
## 61           3-Indoleacetic acid; 40     -0.0557 -0.4770  0.3660  0.7960 0.940
## 62                 Glyceric acid; 30     -0.0480 -0.4670  0.3710  0.8220 0.940
## 63          4-Deoxytetronic acid; 32      0.0477 -0.3740  0.4700  0.8250 0.940
## 64 2,4-Dihydroxybutanoic acid; 28      0.0461 -0.3640  0.4560  0.8260 0.940
## 65                  Pyruvic acid; 31      0.0475 -0.3770  0.4720  0.8260 0.940
## 66             Linoleic acid, TMS; 4      0.0420 -0.3820  0.4660  0.8460 0.940
## 67                 Nonanoic acid; 67      0.0415 -0.3850  0.4680  0.8480 0.940
## 68          4-Deoxytetronic acid; 33      0.0329 -0.3860  0.4510  0.8770 0.940
## 69            Glyceryl-glycoside; 59      0.0317 -0.3910  0.4540  0.8830 0.940
## 70                   Bisphenol A; 48     -0.0301 -0.4580  0.3970  0.8900 0.940
## 71 3-Hydroxybutyric acid, 2TMS; 1      0.0279 -0.3990  0.4550  0.8980 0.940
## 72               1,3-Propanediol; 34      0.0266 -0.4000  0.4530  0.9030 0.940
## 73                      Glycerol; 58     -0.0203 -0.4480  0.4070  0.9260 0.946
## 74             Fumaric acid, 2TMS; 9     -0.0180 -0.4420  0.4070  0.9340 0.946
## 75                 L-5-Oxoproline; 63     -0.0130 -0.4380  0.4120  0.9520 0.952
```

##### 5.1.4 Fully-Adjusted Model

```
## [1] "Fitting models:"
## [1] "~ Amputation.at.DATE + Age.x + Gender.x + Hba1c_baseline + CALSBP + bmi + Smoking + Statin + lo
## [1] ""
```

###### 5.1.4.1 Forest Plot of Model Coefficients

```
## Warning: Ignoring unknown aesthetics: x
## Ignoring unknown aesthetics: x
## Ignoring unknown aesthetics: x
## Ignoring unknown aesthetics: x
## Ignoring unknown aesthetics: x
## Ignoring unknown aesthetics: x
## Ignoring unknown aesthetics: x
## Ignoring unknown aesthetics: x
## Ignoring unknown aesthetics: x
## Ignoring unknown aesthetics: x
## Ignoring unknown aesthetics: x
```

```
## NULL
```

###### 5.1.4.2 Tables of Model Coefficients

```
## [1] ""
## [1] "Table: Amputation.at.DATEJA"
## [1] "  (from model: "
## [1] "   ~ Amputation.at.DATE + Age.x + Gender.x + Hba1c_baseline"
## [1] "     + CALSBP + bmi + Smoking + Statin + log_Blood_TGA +"
## [1] "     Total_cholesterol + egfr + logUAER)"
## [1] ""
## [1] "No significant associations at p.adj < 0.2"
## [1] ""
## [1] "Table: Age.x"
## [1] "  (from model: "
## [1] "   ~ Amputation.at.DATE + Age.x + Gender.x + Hba1c_baseline"
## [1] "     + CALSBP + bmi + Smoking + Statin + log_Blood_TGA +"
## [1] "     Total_cholesterol + egfr + logUAER)"
## [1] ""
##                               Name Coefficient      CI.L     CI.R  p.value
## 1       Eicosapentaenoic acid; 55     0.02550  1.82e-02 0.032900 1.40e-11
## 2        Docosahexaenoic acid; 53     0.01380  6.44e-03 0.021200 2.51e-04
## 3  alpha-ketoglutaric acid, TMS M     0.01270  5.16e-03 0.020200 9.69e-04
## 4                Pyruvic acid; 31     0.01190  4.42e-03 0.019400 1.87e-03
## 5            alpha-Tocopherol; 26     0.01120  3.84e-03 0.018600 2.90e-03
## 6            Malic acid, 3TMS; 11     0.01070  3.22e-03 0.018200 5.10e-03
## 7           Palmitic acid, TMS; 5     0.01060  3.19e-03 0.018100 5.18e-03
## 8               Decanoic acid; 52     0.01060  3.11e-03 0.018100 5.52e-03
## 9         3-Indoleacetic acid; 40     0.01000  2.60e-03 0.017500 8.19e-03
## 10          Nonadecanoic acid; 66     0.01000  2.51e-03 0.017600 9.00e-03
## 11 4-Hydroxybenzeneacetic acid; 4     0.00951  2.19e-03 0.016800 1.10e-02
## 12              Glyceric acid; 30     0.00940  2.02e-03 0.016800 1.26e-02
## 13                    Ribitol; 70     0.00921  1.71e-03 0.016700 1.62e-02
## 14             Oleic acid, TMS; 3     0.00915  1.66e-03 0.016600 1.67e-02
## 15           Myristoleic acid; 65     0.00879  1.33e-03 0.016200 2.10e-02
## 16          Glutamic acid, 3TMS; 8     0.00851  1.15e-03 0.015900 2.35e-02
## 17          Succinic acid, 2TMS; 7     0.00819  6.81e-04 0.015700 3.25e-02
## 18 2,4-Dihydroxybutanoic acid; 28     0.00780  5.96e-04 0.015000 3.38e-02
## 19         11-Eicosenoic acid; 35     0.00798  4.38e-04 0.015500 3.81e-02
## 20          Fumaric acid, 2TMS; 9     0.00782  3.19e-04 0.015300 4.10e-02
## 21           Stearic acid, TMS; 2     0.00780  3.04e-04 0.015300 4.14e-02
## 22                   Tyrosine; 75     0.00775  2.46e-04 0.015200 4.30e-02
## 23             Tartronic acid; 73     0.00756  1.32e-04 0.015000 4.61e-02
## 24         Heptadecanoic acid; 61     0.00750 -2.12e-05 0.015000 5.06e-02
## 25              Alanine, 2TMS; 25     0.00745 -4.02e-05 0.014900 5.12e-02
## 26             Dodecanoic acid; 54     0.00703 -4.51e-04 0.014500 6.55e-02
## 27           Myo inositol 6TMS; 1     0.00665 -5.10e-04 0.013800 6.87e-02
## 28              Hydroxylamine; 62    -0.00699 -1.45e-02 0.000554 6.93e-02
## 29                   Glycerol; 57     0.00697 -5.66e-04 0.014500 6.98e-02
##       p.adj
## 1  1.05e-09
## 2  9.43e-03
## 3  2.42e-02
## 4  3.51e-02
## 5  4.36e-02
## 6  5.18e-02
```

```
## 7  5.18e-02
## 8  5.18e-02
## 9  6.75e-02
## 10 6.75e-02
## 11 7.48e-02
## 12 7.88e-02
## 13 8.93e-02
## 14 8.93e-02
## 15 1.05e-01
## 16 1.10e-01
## 17 1.41e-01
## 18 1.41e-01
## 19 1.46e-01
## 20 1.46e-01
## 21 1.46e-01
## 22 1.46e-01
## 23 1.50e-01
## 24 1.54e-01
## 25 1.54e-01
## 26 1.81e-01
## 27 1.81e-01
## 28 1.81e-01
## 29 1.81e-01
## [1] ""
## [1] "Table: Gender.x"
## [1] "  (from model: "
## [1] "   ~ Amputation.at.DATE + Age.x + Gender.x + Hba1c_baseline"
## [1] "     + CALSBP + bmi + Smoking + Statin + log_Blood_TGA +"
## [1] "     Total_cholesterol + egfr + logUAER)"
## [1] ""
##                          Name Coefficient     CI.L     CI.R  p.value
## 1       4-Deoxytetronic acid; 33       0.404  0.23900  0.56900 1.64e-06
## 2            Tartronic acid; 73      -0.383 -0.54900 -0.21700 6.63e-06
## 3             Proline, 2TMS; 21       0.370  0.20400  0.53600 1.37e-05
## 4          Methionine, 2TMS; 16       0.369  0.20200  0.53500 1.49e-05
## 5             Glyceric acid; 30      -0.363 -0.52900 -0.19800 1.67e-05
## 6          Myristoleic acid; 65      -0.363 -0.53000 -0.19700 2.04e-05
## 7              Valine, 2TMS; 20       0.342  0.17800  0.50600 4.48e-05
## 8             Leucine, 2TMS; 19       0.319  0.15300  0.48500 1.67e-04
## 9          Cholesterol, TMS; 23      -0.306 -0.46800 -0.14400 2.13e-04
## 10           Oleic acid, TMS; 3      -0.302 -0.46900 -0.13400 4.20e-04
## 11          Dodecanoic acid; 54      -0.300 -0.46800 -0.13300 4.34e-04
## 12         Citric acid, 4TMS; 6      -0.285 -0.45000 -0.11900 7.49e-04
## 13            Glycine, 3TMS; 17      -0.277 -0.44400 -0.11100 1.10e-03
## 14         Isoleucine, 2TMS; 18       0.265  0.10000  0.43000 1.63e-03
## 15           Hydroxyproline; 64       0.259  0.09190  0.42700 2.43e-03
## 16 2-hydroxy Isovaleric acid; 38       0.258  0.09060  0.42600 2.56e-03
## 17         Stearic acid, TMS; 2      -0.256 -0.42400 -0.08900 2.71e-03
## 18            Decanoic acid; 52      -0.255 -0.42200 -0.08800 2.79e-03
## 19     Docosahexaenoic acid; 53      -0.248 -0.41300 -0.08260 3.31e-03
## 20         Aminomalonic acid; 45      -0.245 -0.41100 -0.07820 3.98e-03
## 21        Succinic acid, 2TMS; 7      -0.242 -0.41000 -0.07460 4.66e-03
## 22        Glutamic acid, 3TMS; 8       0.237  0.07200  0.40100 4.88e-03
## 23         Palmitic acid, TMS; 5      -0.229 -0.39600 -0.06200 7.21e-03
```

```
## 24          Tridecanoic acid; 74      -0.226 -0.39500 -0.05820 8.37e-03
## 25        Heptadecanoic acid; 61      -0.212 -0.38000 -0.04370 1.35e-02
## 26         Nonadecanoic acid; 66      -0.207 -0.37600 -0.03930 1.56e-02
## 27        Heptadecanoic acid; 60      -0.194 -0.36200 -0.02620 2.35e-02
## 28                  Glycerol; 57      -0.181 -0.35000 -0.01250 3.52e-02
## 29                  Tyrosine; 75      -0.173 -0.34100 -0.00516 4.34e-02
## 30        Benzeneacetic acid; 47      -0.163 -0.33000  0.00449 5.65e-02
## 31            1-Monopalmitin; 37       0.161 -0.00795  0.33000 6.18e-02
## 32     Arachidonic acid, TMS; 24      -0.148 -0.31600  0.02010 8.44e-02
##       p.adj
## 1  0.000123
## 2  0.000249
## 3  0.000250
## 4  0.000250
## 5  0.000250
## 6  0.000255
## 7  0.000480
## 8  0.001570
## 9  0.001780
## 10 0.002960
## 11 0.002960
## 12 0.004680
## 13 0.006320
## 14 0.008750
## 15 0.011600
## 16 0.011600
## 17 0.011600
## 18 0.011600
## 19 0.013100
## 20 0.014900
## 21 0.016600
## 22 0.016600
## 23 0.023500
## 24 0.026200
## 25 0.040600
## 26 0.045100
## 27 0.065400
## 28 0.094400
## 29 0.112000
## 30 0.141000
## 31 0.149000
## 32 0.198000
## [1] ""
## [1] "Table: Hba1c_baseline"
## [1] "  (from model: "
## [1] "   ~ Amputation.at.DATE + Age.x + Gender.x + Hba1c_baseline"
## [1] "     + CALSBP + bmi + Smoking + Statin + log_Blood_TGA +"
## [1] "     Total_cholesterol + egfr + logUAER)"
## [1] ""
##                          Name Coefficient     CI.L     CI.R  p.value  p.adj
## 1            Valine, 2TMS; 20      0.1340  0.06100  0.20700 0.000331 0.0129
## 2             Ethanolamine; 56      0.1350  0.05950  0.21000 0.000451 0.0129
## 3         Arabinopyranose; 51      0.1300  0.05510  0.20400 0.000672 0.0129
## 4           Alanine, 2TMS; 25      0.1280  0.05340  0.20300 0.000790 0.0129
```

```
## 5   Eicosapentaenoic acid; 55     -0.1250 -0.19800 -0.05150 0.000859 0.0129
## 6    Docosahexaenoic acid; 53     -0.1110 -0.18500 -0.03740 0.003160 0.0395
## 7        Citric acid, 4TMS; 6      0.1090  0.03510  0.18300 0.003840 0.0411
## 8  4-Hydroxybutanoic acid; 43     -0.0976 -0.17300 -0.02250 0.010900 0.0874
## 9           Decanoic acid; 52     -0.0961 -0.17100 -0.02160 0.011500 0.0874
## 10          Glyceric acid; 30     -0.0941 -0.16800 -0.02050 0.012200 0.0874
## 11       Tridecanoic acid; 74     -0.0953 -0.17000 -0.02030 0.012800 0.0874
## 12            Campesterol; 49      0.0866  0.01310  0.16000 0.020900 0.1310
## 13     Glyceryl-glycoside; 59      0.0836  0.00918  0.15800 0.027700 0.1510
## 14            Lactic acid; 29      0.0826  0.00791  0.15700 0.030200 0.1510
## 15          Leucine, 2TMS; 19      0.0819  0.00783  0.15600 0.030200 0.1510
## 16   4-Deoxytetronic acid; 33     -0.0778 -0.15100 -0.00433 0.038000 0.1690
## 17       Myristoleic acid; 65     -0.0786 -0.15300 -0.00426 0.038300 0.1690
## 18                Ribitol; 70     -0.0780 -0.15300 -0.00323 0.040900 0.1700
## [1] ""
## [1] "Table: CALSBP"
## [1] "  (from model: "
## [1] "   ~ Amputation.at.DATE + Age.x + Gender.x + Hba1c_baseline"
## [1] "     + CALSBP + bmi + Smoking + Statin + log_Blood_TGA +"
## [1] "     Total_cholesterol + egfr + logUAER)"
## [1] ""
##                   Name Coefficient    CI.L   CI.R p.value p.adj
## 1 Myristoleic acid; 65     0.00797 0.00306 0.0129 0.00148 0.111
## [1] ""
## [1] "Table: bmi"
## [1] "  (from model: "
## [1] "   ~ Amputation.at.DATE + Age.x + Gender.x + Hba1c_baseline"
## [1] "     + CALSBP + bmi + Smoking + Statin + log_Blood_TGA +"
## [1] "     Total_cholesterol + egfr + logUAER)"
## [1] ""
##                              Name Coefficient     CI.L      CI.R  p.value
## 1          Glutamic acid, 3TMS; 8      0.0446  0.02410  0.065100 2.14e-05
## 2  2-Hydroxybutyric acid, 2TMS; 2      0.0423  0.02180  0.062800 5.41e-05
## 3                 Campesterol; 49     -0.0400 -0.06060 -0.019500 1.34e-04
## 4               Decanoic acid; 52     -0.0354 -0.05620 -0.014600 8.63e-04
## 5                 Lactic acid; 29      0.0299  0.00899  0.050700 5.07e-03
## 6  2,4-Dihydroxybutanoic acid; 28     -0.0279 -0.04790 -0.007820 6.48e-03
## 7             1,3-Propanediol; 34     -0.0291 -0.05010 -0.008120 6.61e-03
## 8                Pyruvic acid; 31     -0.0288 -0.04980 -0.007930 6.91e-03
## 9            alpha-Tocopherol; 26     -0.0274 -0.04790 -0.006820 9.07e-03
## 10             Arachidic acid; 46     -0.0275 -0.04850 -0.006620 9.91e-03
## 11          Nonadecanoic acid; 66     -0.0256 -0.04650 -0.004660 1.66e-02
## 12          Pyroglutamic acid; 69     -0.0254 -0.04620 -0.004600 1.67e-02
## 13           Citric acid, 4TMS; 6     -0.0247 -0.04530 -0.004090 1.88e-02
## 14            Dodecanoic acid; 54     -0.0237 -0.04450 -0.002910 2.55e-02
## 15         11-Eicosenoic acid; 35      0.0236  0.00265  0.044600 2.73e-02
## 16           Isoleucine, 2TMS; 18      0.0225  0.00204  0.043000 3.12e-02
## 17 alpha-ketoglutaric acid, TMS M     -0.0229 -0.04390 -0.001980 3.20e-02
## 18             Tartronic acid; 73     -0.0224 -0.04310 -0.001680 3.41e-02
## 19              Glycine, 3TMS; 17     -0.0219 -0.04260 -0.001210 3.81e-02
## 20                    Ribitol; 70      0.0221  0.00121  0.043000 3.82e-02
## 21              Octanoic acid; 68     -0.0204 -0.04130  0.000426 5.49e-02
##      p.adj
## 1  0.00161
```

```
## 2  0.00203
## 3  0.00336
## 4  0.01620
## 5  0.06470
## 6  0.06470
## 7  0.06470
## 8  0.06470
## 9  0.07440
## 10 0.07440
## 11 0.10400
## 12 0.10400
## 13 0.10900
## 14 0.13600
## 15 0.13600
## 16 0.14100
## 17 0.14100
## 18 0.14200
## 19 0.14300
## 20 0.14300
## 21 0.19600
## [1] ""
## [1] "Table: Smoking"
## [1] "  (from model: "
## [1] "   ~ Amputation.at.DATE + Age.x + Gender.x + Hba1c_baseline"
## [1] "     + CALSBP + bmi + Smoking + Statin + log_Blood_TGA +"
## [1] "     Total_cholesterol + egfr + logUAER)"
## [1] ""
##                          Name Coefficient   CI.L    CI.R  p.value  p.adj
## 1  3-Indolepropionic acid; 41      -0.357 -0.556 -0.1580 0.000437 0.0201
## 2          Tartronic acid; 73      -0.340 -0.538 -0.1430 0.000739 0.0201
## 3    Docosahexaenoic acid; 53      -0.336 -0.532 -0.1400 0.000806 0.0201
## 4      Glutamic acid, 3TMS; 8       0.323  0.127  0.5190 0.001220 0.0229
## 5            Valine, 2TMS; 20      -0.311 -0.505 -0.1160 0.001800 0.0270
## 6        alpha-Tocopherol; 26      -0.297 -0.493 -0.1010 0.003020 0.0333
## 7           Glyceric acid; 30      -0.296 -0.492 -0.1000 0.003110 0.0333
## 8      Benzeneacetic acid; 47      -0.265 -0.464 -0.0661 0.009060 0.0750
## 9        Citric acid, 4TMS; 6      -0.261 -0.458 -0.0644 0.009300 0.0750
## 10          Leucine, 2TMS; 19      -0.259 -0.457 -0.0621 0.010000 0.0750
## 11       Malic acid, 3TMS; 11      -0.253 -0.452 -0.0537 0.012800 0.0876
## 12           Ribonic acid; 72      -0.233 -0.425 -0.0417 0.017000 0.1060
## 13            Campesterol; 49      -0.234 -0.430 -0.0387 0.018900 0.1090
## [1] ""
## [1] "Table: Statin"
## [1] "  (from model: "
## [1] "   ~ Amputation.at.DATE + Age.x + Gender.x + Hba1c_baseline"
## [1] "     + CALSBP + bmi + Smoking + Statin + log_Blood_TGA +"
## [1] "     Total_cholesterol + egfr + logUAER)"
## [1] ""
##                        Name Coefficient    CI.L    CI.R p.value  p.adj
## 1           Campesterol; 49       0.289  0.1070  0.4710 0.00187 0.0888
## 2        L-5-Oxoproline; 63      -0.281 -0.4660 -0.0951 0.00305 0.0888
## 3 Arachidonic acid, TMS; 24       0.276  0.0907  0.4620 0.00355 0.0888
## [1] ""
## [1] "Table: log_Blood_TGA"
```

```
## [1] "  (from model: "
## [1] "   ~ Amputation.at.DATE + Age.x + Gender.x + Hba1c_baseline"
## [1] "     + CALSBP + bmi + Smoking + Statin + log_Blood_TGA +"
## [1] "     Total_cholesterol + egfr + logUAER)"
## [1] ""
##                               Name Coefficient      CI.L     CI.R  p.value
## 1           Palmitic acid, TMS; 5       0.281  0.149000  0.41300 2.97e-05
## 2               Octanoic acid; 68       0.266  0.134000  0.39800 8.16e-05
## 3              Arachidic acid; 46       0.262  0.130000  0.39500 1.09e-04
## 4             Dodecanoic acid; 54       0.261  0.129000  0.39300 1.09e-04
## 5            Stearic acid, TMS; 2       0.258  0.126000  0.39100 1.31e-04
## 6  2-Hydroxybutyric acid, 2TMS; 2       0.237  0.107000  0.36700 3.57e-04
## 7               Decanoic acid; 52       0.239  0.107000  0.37100 3.86e-04
## 8              Oleic acid, TMS; 3       0.213  0.080400  0.34500 1.64e-03
## 9                 Lactic acid; 29       0.196  0.064000  0.32900 3.66e-03
## 10         Glyceryl-glycoside; 59       0.190  0.058200  0.32200 4.75e-03
## 11           Isoleucine, 2TMS; 18       0.187  0.056900  0.31700 4.85e-03
## 12               Valine, 2TMS; 20       0.186  0.056500  0.31500 4.91e-03
## 13         Heptadecanoic acid; 60       0.180  0.046800  0.31200 8.07e-03
## 14            Arabinopyranose; 51       0.173  0.041200  0.30600 1.02e-02
## 15 4-Hydroxybenzeneacetic acid; 4       0.166  0.036900  0.29600 1.18e-02
## 16           Myristoleic acid; 65       0.163  0.031200  0.29400 1.53e-02
## 17          Aminomalonic acid; 45      -0.161 -0.293000 -0.03000 1.61e-02
## 18               Ribonic acid; 72       0.155  0.028000  0.28300 1.68e-02
## 19 alpha-ketoglutaric acid, TMS M       0.161  0.028100  0.29400 1.76e-02
## 20         Glutamic acid, 3TMS; 8       0.157  0.027300  0.28700 1.78e-02
## 21               Pyruvic acid; 31       0.160  0.027300  0.29300 1.81e-02
## 22 3,4-Dihydroxybutanoic acid; 27       0.153  0.025000  0.28100 1.92e-02
## 23              Leucine, 2TMS; 19       0.156  0.024800  0.28700 1.98e-02
## 24          Fumaric acid, 2TMS; 9       0.154  0.021700  0.28600 2.26e-02
## 25         Heptadecanoic acid; 61       0.154  0.021100  0.28600 2.32e-02
## 26      Arachidonic acid, TMS; 24       0.149  0.016100  0.28200 2.80e-02
## 27           Malic acid, 3TMS; 11       0.139  0.006890  0.27100 3.92e-02
## 28          Nonadecanoic acid; 66       0.137  0.004320  0.27000 4.30e-02
## 29     4-Hydroxybutanoic acid; 43       0.133 -0.000334  0.26500 5.06e-02
## 30         Succinic acid, 2TMS; 7       0.131 -0.001280  0.26400 5.23e-02
## 31              Glyceric acid; 30      -0.129 -0.259000  0.00172 5.31e-02
## 32            1,3-Propanediol; 34       0.130 -0.002880  0.26400 5.52e-02
## 33              Proline, 2TMS; 21       0.127 -0.003950  0.25900 5.73e-02
## 34                    Ribitol; 71       0.121 -0.006290  0.24800 6.25e-02
## 35     3-Indolepropionic acid; 41      -0.121 -0.253000  0.01130 7.30e-02
## 36              Glycine, 3TMS; 17      -0.114 -0.245000  0.01740 8.91e-02
## 37             Tartronic acid; 73      -0.114 -0.245000  0.01760 8.97e-02
## 38       4-Deoxytetronic acid; 33       0.111 -0.019300  0.24100 9.52e-02
##      p.adj
## 1  0.00197
## 2  0.00197
## 3  0.00197
## 4  0.00197
## 5  0.00197
## 6  0.00413
## 7  0.00413
## 8  0.01540
## 9  0.03050
```

```
## 10 0.03070
## 11 0.03070
## 12 0.03070
## 13 0.04660
## 14 0.05460
## 15 0.05890
## 16 0.06460
## 17 0.06460
## 18 0.06460
## 19 0.06460
## 20 0.06460
## 21 0.06460
## 22 0.06460
## 23 0.06460
## 24 0.06950
## 25 0.06950
## 26 0.08070
## 27 0.10900
## 28 0.11500
## 29 0.12800
## 30 0.12800
## 31 0.12800
## 32 0.12900
## 33 0.13000
## 34 0.13800
## 35 0.15600
## 36 0.18200
## 37 0.18200
## 38 0.18800
## [1] ""
## [1] "Table: Total_cholesterol"
## [1] "  (from model: "
## [1] "   ~ Amputation.at.DATE + Age.x + Gender.x + Hba1c_baseline"
## [1] "     + CALSBP + bmi + Smoking + Statin + log_Blood_TGA +"
## [1] "     Total_cholesterol + egfr + logUAER)"
## [1] ""
##                               Name Coefficient      CI.L     CI.R  p.value
## 1            Cholesterol, TMS; 23      0.4510  0.356000  0.54600 2.96e-20
## 2                 Campesterol; 49      0.3730  0.276000  0.46900 5.72e-14
## 3            alpha-Tocopherol; 26      0.3230  0.227000  0.42000 6.93e-11
## 4          Benzeneacetic acid; 47     -0.1890 -0.287000 -0.09100 1.61e-04
## 5           Linoleic acid, TMS; 4      0.1810  0.083300  0.27900 2.94e-04
## 6      4-Hydroxybutanoic acid; 43     -0.1700 -0.269000 -0.07160 7.26e-04
## 7              L-5-Oxoproline; 63     -0.1690 -0.267000 -0.07040 7.82e-04
## 8            Isoleucine, 2TMS; 18     -0.1640 -0.261000 -0.06790 8.48e-04
## 9               Proline, 2TMS; 21     -0.1640 -0.261000 -0.06630 9.94e-04
## 10                   Tyrosine; 75     -0.1600 -0.258000 -0.06170 1.42e-03
## 11           Methionine, 2TMS; 16     -0.1570 -0.254000 -0.05930 1.64e-03
## 12      Eicosapentaenoic acid; 55      0.1530  0.057200  0.25000 1.80e-03
## 13              Glycine, 3TMS; 17     -0.1510 -0.248000 -0.05330 2.43e-03
## 14       Docosahexaenoic acid; 53      0.1480  0.051500  0.24500 2.69e-03
## 15            Threonine, 3TMS; 12     -0.1500 -0.248000 -0.05140 2.88e-03
## 16              Alanine, 2TMS; 25     -0.1380 -0.236000 -0.03970 5.93e-03
## 17            Arabinopyranose; 51     -0.1360 -0.234000 -0.03800 6.54e-03
```

```
## 18         Glyceryl-glycoside; 59     -0.1310 -0.229000 -0.03370 8.42e-03
## 19 4-Hydroxybenzeneacetic acid; 4     -0.1270 -0.223000 -0.03140 9.27e-03
## 20            Malic acid, 3TMS; 11     -0.1290 -0.227000 -0.03080 1.00e-02
## 21 2,4-Dihydroxybutanoic acid; 28     -0.1220 -0.216000 -0.02770 1.12e-02
## 22        2-Palmitoylglycerol; 39      0.1240  0.025500  0.22300 1.36e-02
## 23              Hydroxylamine; 62     -0.1230 -0.222000 -0.02450 1.44e-02
## 24               Serine, 3TMS; 14     -0.1220 -0.220000 -0.02420 1.46e-02
## 25        3-Indoleacetic acid; 40     -0.1160 -0.213000 -0.01820 2.01e-02
## 26                    Ribitol; 71     -0.1020 -0.196000 -0.00744 3.45e-02
## 27        Phenylalanine, 2TMS; 13     -0.1060 -0.205000 -0.00774 3.45e-02
## 28               Ribonic acid; 72     -0.0969 -0.191000 -0.00252 4.42e-02
## 29          Palmitic acid, TMS; 5      0.0971 -0.000468  0.19500 5.11e-02
## 30 3,4-Dihydroxybutanoic acid; 27     -0.0928 -0.188000  0.00217 5.55e-02
## 31               Pyruvic acid; 31     -0.0955 -0.194000  0.00274 5.67e-02
## 32               Ethanolamine; 56     -0.0934 -0.192000  0.00510 6.31e-02
## 33              Leucine, 2TMS; 19     -0.0914 -0.189000  0.00572 6.51e-02
##       p.adj
## 1  2.22e-18
## 2  2.15e-12
## 3  1.73e-09
## 4  3.01e-03
## 5  4.41e-03
## 6  7.95e-03
## 7  7.95e-03
## 8  7.95e-03
## 9  8.28e-03
## 10 1.07e-02
## 11 1.11e-02
## 12 1.12e-02
## 13 1.40e-02
## 14 1.44e-02
## 15 1.44e-02
## 16 2.78e-02
## 17 2.89e-02
## 18 3.51e-02
## 19 3.66e-02
## 20 3.75e-02
## 21 4.01e-02
## 22 4.56e-02
## 23 4.56e-02
## 24 4.56e-02
## 25 6.02e-02
## 26 9.59e-02
## 27 9.59e-02
## 28 1.18e-01
## 29 1.32e-01
## 30 1.37e-01
## 31 1.37e-01
## 32 1.48e-01
## 33 1.48e-01
## [1] ""
## [1] "Table: egfr"
## [1] "  (from model: "
## [1] "   ~ Amputation.at.DATE + Age.x + Gender.x + Hba1c_baseline"
```

```
## [1] "     + CALSBP + bmi + Smoking + Statin + log_Blood_TGA +"
## [1] "     Total_cholesterol + egfr + logUAER)"
## [1] ""
##                               Name Coefficient      CI.L      CI.R  p.value
## 1            Myo inositol 6TMS; 1    -0.01600 -1.92e-02 -1.28e-02 2.87e-22
## 2                     Ribitol; 71    -0.01510 -1.83e-02 -1.19e-02 6.56e-20
## 3                  Creatinine; 50    -0.01490 -1.82e-02 -1.17e-02 6.34e-19
## 4  2,4-Dihydroxybutanoic acid; 28    -0.01470 -1.79e-02 -1.15e-02 6.82e-19
## 5                Ribonic acid; 72    -0.01310 -1.63e-02 -9.91e-03 2.23e-15
## 6  3,4-Dihydroxybutanoic acid; 27    -0.01130 -1.45e-02 -8.06e-03 1.05e-11
## 7        4-Deoxytetronic acid; 33    -0.01120 -1.45e-02 -7.93e-03 2.88e-11
## 8  4-Hydroxybenzeneacetic acid; 4    -0.01090 -1.41e-02 -7.59e-03 9.16e-11
## 9            Isoleucine, 2TMS; 18     0.00980  6.52e-03  1.31e-02 5.71e-09
## 10       4-Deoxytetronic acid; 32    -0.00898 -1.23e-02 -5.67e-03 1.16e-07
## 11           Citric acid, 4TMS; 6    -0.00893 -1.22e-02 -5.63e-03 1.24e-07
## 12          Pyroglutamic acid; 69    -0.00862 -1.19e-02 -5.29e-03 4.11e-07
## 13 2-Hydroxybutyric acid, 2TMS; 2     0.00832  5.04e-03  1.16e-02 7.18e-07
## 14         3-Indoleacetic acid; 40    -0.00756 -1.09e-02 -4.24e-03 8.35e-06
## 15 4-Hydroxyphenyllactic acid; 44    -0.00756 -1.09e-02 -4.24e-03 8.70e-06
## 16               Valine, 2TMS; 20     0.00741  4.15e-03  1.07e-02 9.19e-06
## 17          Glyceryl-glycoside; 59    -0.00644 -9.77e-03 -3.12e-03 1.50e-04
## 18              Hydroxyproline; 64    -0.00623 -9.56e-03 -2.89e-03 2.62e-04
## 19              Leucine, 2TMS; 19     0.00599  2.68e-03  9.30e-03 3.95e-04
## 20              Glycine, 3TMS; 17    -0.00600 -9.32e-03 -2.69e-03 3.95e-04
## 21               Serine, 3TMS; 14     0.00604  2.70e-03  9.38e-03 4.01e-04
## 22          Fumaric acid, 2TMS; 9    -0.00586 -9.20e-03 -2.52e-03 5.96e-04
## 23            Stearic acid, TMS; 2     0.00540  2.06e-03  8.74e-03 1.54e-03
## 24      Eicosapentaenoic acid; 55     0.00501  1.73e-03  8.29e-03 2.75e-03
## 25           Methionine, 2TMS; 16     0.00459  1.27e-03  7.90e-03 6.74e-03
## 26                    Glycerol; 57     0.00429  9.35e-04  7.65e-03 1.23e-02
## 27               Octanoic acid; 68     0.00409  7.51e-04  7.42e-03 1.64e-02
## 28          Palmitic acid, TMS; 5     0.00407  7.46e-04  7.39e-03 1.64e-02
## 29          Benzeneacetic acid; 47    -0.00407 -7.41e-03 -7.37e-04 1.67e-02
## 30            Malic acid, 3TMS; 11    -0.00401 -7.35e-03 -6.76e-04 1.85e-02
## 31          Aminomalonic acid; 45    -0.00383 -7.15e-03 -5.17e-04 2.35e-02
## 32           Cholesterol, TMS; 23     0.00337  1.44e-04  6.59e-03 4.06e-02
## 33                    Tyrosine; 75     0.00334  1.18e-06  6.68e-03 4.99e-02
## 34              Alanine, 2TMS; 25    -0.00334 -6.68e-03 -2.94e-07 5.00e-02
## 35                    Glycerol; 58    -0.00313 -6.50e-03  2.44e-04 6.90e-02
## 36                 Campesterol; 49    -0.00303 -6.32e-03  2.51e-04 7.02e-02
## 37        Phenylalanine, 2TMS; 13    -0.00294 -6.29e-03  4.14e-04 8.58e-02
## 38  2-hydroxy Isovaleric acid; 38     0.00292 -4.28e-04  6.26e-03 8.74e-02
## 39        2-Palmitoylglycerol; 39     0.00285 -5.06e-04  6.21e-03 9.60e-02
##       p.adj
## 1  2.15e-20
## 2  2.46e-18
## 3  1.28e-17
## 4  1.28e-17
## 5  3.34e-14
## 6  1.32e-10
## 7  3.09e-10
## 8  8.59e-10
## 9  4.76e-08
## 10 8.45e-07
```

```
## 11 8.45e-07
## 12 2.57e-06
## 13 4.14e-06
## 14 4.31e-05
## 15 4.31e-05
## 16 4.31e-05
## 17 6.64e-04
## 18 1.09e-03
## 19 1.43e-03
## 20 1.43e-03
## 21 1.43e-03
## 22 2.03e-03
## 23 5.02e-03
## 24 8.58e-03
## 25 2.02e-02
## 26 3.54e-02
## 27 4.33e-02
## 28 4.33e-02
## 29 4.33e-02
## 30 4.62e-02
## 31 5.69e-02
## 32 9.51e-02
## 33 1.10e-01
## 34 1.10e-01
## 35 1.46e-01
## 36 1.46e-01
## 37 1.72e-01
## 38 1.72e-01
## 39 1.85e-01
## [1] ""
## [1] "Table: logUAER"
## [1] "  (from model: "
## [1] "   ~ Amputation.at.DATE + Age.x + Gender.x + Hba1c_baseline"
## [1] "     + CALSBP + bmi + Smoking + Statin + log_Blood_TGA +"
## [1] "     Total_cholesterol + egfr + logUAER)"
## [1] ""
##                              Name Coefficient   CI.L  CI.R  p.value  p.adj
## 1 3,4-Dihydroxybutanoic acid; 27      0.0733 0.0333 0.113 0.000337 0.0253
## 2       4-Deoxytetronic acid; 32      0.0638 0.0229 0.105 0.002270 0.0853
```

###### 5.1.4.3 Table with All Metabolites

```
## [1] ""
## [1] "Table: Amputation.at.DATEJA"
## [1] "  (from model: "
## [1] "   ~ Amputation.at.DATE + Age.x + Gender.x + Hba1c_baseline"
## [1] "     + CALSBP + bmi + Smoking + Statin + log_Blood_TGA +"
## [1] "     Total_cholesterol + egfr + logUAER)"
## [1] ""
##                               Name Coefficient    CI.L    CI.R p.value p.adj
## 1          Succinic acid, 2TMS; 7     0.55400  0.1290  0.9780  0.0107 0.509
## 2                    Glycerol; 57     0.52300  0.0963  0.9490  0.0163 0.509
## 3             Threonine, 3TMS; 12    -0.49200 -0.9170 -0.0663  0.0235 0.509
## 4         Phenylalanine, 2TMS; 13    -0.48000 -0.9060 -0.0543  0.0271 0.509
## 5          Glutamic acid, 3TMS; 8    -0.43800 -0.8550 -0.0213  0.0394 0.591
## 6                Ribonic acid; 72     0.37800 -0.0303  0.7870  0.0696 0.867
## 7            Stearic acid, TMS; 2     0.35300 -0.0707  0.7770  0.1020 0.867
## 8  2-Hydroxybutyric acid, 2TMS; 2     0.34100 -0.0755  0.7570  0.1090 0.867
## 9          Heptadecanoic acid; 61     0.33400 -0.0916  0.7590  0.1240 0.867
## 10                   Tyrosine; 75    -0.32900 -0.7540  0.0950  0.1280 0.867
## 11              Proline, 2TMS; 21    -0.31600 -0.7370  0.1050  0.1410 0.867
## 12           Methionine, 2TMS; 16    -0.31000 -0.7310  0.1120  0.1490 0.867
## 13            Myristoleic acid; 65     0.28600 -0.1360  0.7080  0.1840 0.867
## 14             Arabinopyranose; 51    -0.27200 -0.6960  0.1520  0.2080 0.867
## 15             Dodecanoic acid; 54     0.27100 -0.1520  0.6940  0.2090 0.867
## 16     4-Hydroxybutanoic acid; 43     0.27000 -0.1560  0.6960  0.2150 0.867
## 17                 Campesterol; 49    -0.26100 -0.6780  0.1560  0.2200 0.867
## 18           Pyroglutamic acid; 69     0.26200 -0.1610  0.6840  0.2240 0.867
## 19 4-Hydroxybenzeneacetic acid; 4     0.25500 -0.1590  0.6700  0.2270 0.867
## 20            Malic acid, 3TMS; 11     0.23900 -0.1850  0.6620  0.2700 0.867
## 21              Leucine, 2TMS; 19    -0.22600 -0.6460  0.1940  0.2920 0.867
## 22           Aminomalonic acid; 45    -0.22600 -0.6470  0.1960  0.2940 0.867
## 23              Hydroxyproline; 64     0.22500 -0.1990  0.6490  0.2970 0.867
## 24              Myo inositol 6TMS; 1     0.21400 -0.1920  0.6190  0.3010 0.867
## 25           Palmitic acid, TMS; 5     0.21500 -0.2070  0.6370  0.3170 0.867
## 26     3-Indolepropionic acid; 41    -0.21300 -0.6360  0.2110  0.3250 0.867
## 27               Octanoic acid; 68     0.21100 -0.2130  0.6340  0.3300 0.867
## 28          Heptadecanoic acid; 60     0.21000 -0.2150  0.6360  0.3320 0.867
## 29 4-Hydroxyphenyllactic acid; 44     0.20700 -0.2150  0.6300  0.3350 0.867
## 30             Tridecanoic acid; 74     0.19500 -0.2310  0.6210  0.3690 0.922
## 31  2-hydroxy Isovaleric acid; 38    -0.17500 -0.6000  0.2490  0.4180 0.969
## 32               Decanoic acid; 52     0.17300 -0.2500  0.5960  0.4220 0.969
## 33                Valine, 2TMS; 20    -0.16400 -0.5790  0.2510  0.4380 0.969
## 34                Serine, 3TMS; 14    -0.16600 -0.5900  0.2580  0.4430 0.969
## 35        2-Palmitoylglycerol; 39    -0.15800 -0.5840  0.2690  0.4680 0.969
## 36               Alanine, 2TMS; 25     0.15400 -0.2700  0.5780  0.4760 0.969
## 37               Hydroxylamine; 62     0.15200 -0.2740  0.5790  0.4840 0.969
## 38                 1-Dodecanol; 36     0.13700 -0.2900  0.5650  0.5290 0.969
## 39              Arachidic acid; 46     0.13600 -0.2890  0.5620  0.5300 0.969
## 40                 Lactic acid; 29     0.13100 -0.2930  0.5560  0.5430 0.969
## 41            Citric acid, 4TMS; 6    -0.12200 -0.5410  0.2970  0.5680 0.969
## 42         11-Eicosenoic acid; 35     0.12000 -0.3060  0.5470  0.5810 0.969
## 43                  Creatinine; 50     0.11400 -0.3000  0.5280  0.5880 0.969
## 44              1-Monopalmitin; 37     0.11700 -0.3110  0.5440  0.5930 0.969
```

```
## 45      Eicosapentaenoic acid; 55    -0.10500 -0.5220  0.3110  0.6190 0.969
## 46                    Ribitol; 70     0.10100 -0.3230  0.5260  0.6400 0.969
## 47          Cholesterol, TMS; 23     0.09560 -0.3140  0.5050  0.6470 0.969
## 48             Tartronic acid; 73    -0.09770 -0.5180  0.3230  0.6490 0.969
## 49      Arachidonic acid, TMS; 24     0.09520 -0.3310  0.5210  0.6610 0.969
## 50              Glycine, 3TMS; 17    -0.08960 -0.5110  0.3310  0.6760 0.969
## 51           alpha-Tocopherol; 26    -0.08260 -0.5000  0.3350  0.6980 0.969
## 52 alpha-ketoglutaric acid, TMS M    -0.07540 -0.5010  0.3500  0.7280 0.969
## 53       Docosahexaenoic acid; 53     0.07250 -0.3460  0.4910  0.7340 0.969
## 54         3-Indoleacetic acid; 40    -0.06790 -0.4890  0.3530  0.7520 0.969
## 55        Benzeneacetic acid; 47    -0.06620 -0.4900  0.3580  0.7590 0.969
## 56                    Ribitol; 71     0.06250 -0.3450  0.4700  0.7630 0.969
## 57               Ethanolamine; 56     0.06320 -0.3630  0.4890  0.7710 0.969
## 58           Isoleucine, 2TMS; 18     0.05270 -0.3640  0.4690  0.8040 0.969
## 59          Linoleic acid, TMS; 4     0.05130 -0.3730  0.4750  0.8130 0.969
## 60             Oleic acid, TMS; 3     0.05030 -0.3740  0.4740  0.8160 0.969
## 61               Pyruvic acid; 31     0.04530 -0.3800  0.4700  0.8350 0.969
## 62 3,4-Dihydroxybutanoic acid; 27     0.04320 -0.3680  0.4540  0.8370 0.969
## 63              Glyceric acid; 30    -0.03570 -0.4540  0.3820  0.8670 0.969
## 64          Nonadecanoic acid; 66     0.03520 -0.3900  0.4610  0.8710 0.969
## 65                   Glycerol; 58    -0.03410 -0.4620  0.3940  0.8760 0.969
## 66 3-Hydroxybutyric acid, 2TMS; 1     0.03040 -0.3980  0.4580  0.8890 0.969
## 67          Fumaric acid, 2TMS; 9    -0.02840 -0.4530  0.3960  0.8960 0.969
## 68            1,3-Propanediol; 34     0.02550 -0.4010  0.4530  0.9070 0.969
## 69              Nonanoic acid; 67     0.02370 -0.4040  0.4510  0.9130 0.969
## 70                Bisphenol A; 48    -0.02320 -0.4510  0.4050  0.9150 0.969
## 71             L-5-Oxoproline; 63    -0.01970 -0.4450  0.4060  0.9280 0.969
## 72       4-Deoxytetronic acid; 32     0.01880 -0.4010  0.4390  0.9300 0.969
## 73         Glyceryl-glycoside; 59     0.00789 -0.4150  0.4300  0.9710 0.978
## 74       4-Deoxytetronic acid; 33     0.00733 -0.4100  0.4240  0.9730 0.978
## 75 2,4-Dihydroxybutanoic acid; 28     0.00584 -0.4020  0.4130  0.9780 0.978
```

#### 5.2 Amputation from DATE

##### 5.2.1 Crude Model

###### 5.2.1.1 Forest Plot of Model Coefficients

```
## NULL
```

###### 5.2.1.2 Table with All Metabolites

| Name | exp(coef) | Lower 95 % | Upper 95 % | Pr(>\|z\|) | p.adj |
|---|---|---|---|---|---|
| 4-Hydroxybenzeneacetic acid | 1.97 | 1.28 | 3.01 | 0.00189 | 0.142 |
| Ribitol (2) | 1.53 | 1.02 | 2.3 | 0.0384 | 0.525 |
| Palmitic acid | 0.697 | 0.493 | 0.986 | 0.0416 | 0.525 |
| Stearic acid | 0.687 | 0.467 | 1.01 | 0.0572 | 0.525 |
| Ribonic acid | 1.5 | 0.98 | 2.28 | 0.0622 | 0.525 |
| 2-Hydroxybutyric acid | 0.747 | 0.543 | 1.03 | 0.0731 | 0.525 |
| Oleic acid | 0.703 | 0.474 | 1.04 | 0.0802 | 0.525 |
| 2,4-Dihydroxybutanoic acid | 1.39 | 0.959 | 2.01 | 0.0822 | 0.525 |
| 3,4-Dihydroxybutanoic acid | 1.37 | 0.943 | 2 | 0.0982 | 0.525 |
| Hydroxylamine | 0.805 | 0.615 | 1.05 | 0.113 | 0.525 |
| Ethanolamine | 1.35 | 0.928 | 1.97 | 0.116 | 0.525 |
| Decanoic acid | 0.787 | 0.582 | 1.07 | 0.121 | 0.525 |
| Arabinopyranose | 1.45 | 0.897 | 2.33 | 0.13 | 0.525 |
| 3-Hydroxybutyric acid | 0.757 | 0.523 | 1.1 | 0.14 | 0.525 |
| 3-Indolepropionic acid | 0.831 | 0.647 | 1.07 | 0.148 | 0.525 |
| 4-Deoxytetronic acid (1) | 1.35 | 0.9 | 2.01 | 0.148 | 0.525 |
| 2-hydroxy Isovaleric acid | 0.786 | 0.566 | 1.09 | 0.15 | 0.525 |
| Benzeneacetic acid | 1.38 | 0.883 | 2.14 | 0.158 | 0.525 |
| Glycerol (2) | 0.798 | 0.581 | 1.1 | 0.163 | 0.525 |
| Linoleic acid | 0.834 | 0.645 | 1.08 | 0.165 | 0.525 |
| Creatinine | 1.32 | 0.889 | 1.97 | 0.168 | 0.525 |
| Nonanoic acid | 0.813 | 0.605 | 1.09 | 0.168 | 0.525 |
| Arachidic acid | 0.77 | 0.531 | 1.12 | 0.169 | 0.525 |
| Glutamic acid | 1.32 | 0.887 | 1.96 | 0.171 | 0.525 |
| Tridecanoic acid | 0.822 | 0.62 | 1.09 | 0.175 | 0.525 |
| 11-Eicosenoic acid | 0.828 | 0.62 | 1.11 | 0.201 | 0.561 |
| 1,3-Propanediol | 0.828 | 0.618 | 1.11 | 0.205 | 0.561 |
| Lactic acid | 0.804 | 0.572 | 1.13 | 0.21 | 0.561 |
| L-5-Oxoproline | 0.823 | 0.591 | 1.14 | 0.247 | 0.621 |
| 1-Dodecanol | 0.821 | 0.586 | 1.15 | 0.252 | 0.621 |
| Heptadecanoic acid (2) | 0.819 | 0.58 | 1.16 | 0.257 | 0.621 |
| Tartronic acid | 0.829 | 0.588 | 1.17 | 0.286 | 0.671 |
| Methionine | 1.23 | 0.825 | 1.83 | 0.309 | 0.697 |
| Glycerol (1) | 0.886 | 0.7 | 1.12 | 0.316 | 0.697 |
| Phenylalanine | 1.21 | 0.826 | 1.77 | 0.33 | 0.702 |
| Alanine | 1.21 | 0.822 | 1.77 | 0.337 | 0.702 |
| Proline | 1.19 | 0.813 | 1.75 | 0.371 | 0.721 |
| Docosahexaenoic acid | 0.88 | 0.66 | 1.17 | 0.386 | 0.721 |
| Eicosapentaenoic acid | 1.19 | 0.802 | 1.77 | 0.387 | 0.721 |
| Myristoleic acid | 0.863 | 0.616 | 1.21 | 0.395 | 0.721 |
| Glyceryl-glycoside | 1.21 | 0.777 | 1.9 | 0.395 | 0.721 |
| Campesterol | 0.869 | 0.619 | 1.22 | 0.418 | 0.721 |
| Fumaric acid | 1.16 | 0.799 | 1.69 | 0.43 | 0.721 |
| 3-Indoleacetic acid | 1.17 | 0.794 | 1.71 | 0.433 | 0.721 |
| Dodecanoic acid | 0.868 | 0.605 | 1.24 | 0.441 | 0.721 |
| Bisphenol A | 0.869 | 0.601 | 1.26 | 0.456 | 0.721 |
| Glyceric acid | 0.88 | 0.628 | 1.23 | 0.458 | 0.721 |
| Isoleucine | 1.17 | 0.766 | 1.8 | 0.463 | 0.721 |
| Leucine | 1.16 | 0.77 | 1.76 | 0.473 | 0.721 |
| 4-Hydroxyphenyllactic acid | 1.16 | 0.769 | 1.75 | 0.48 | 0.721 |

| Name | exp(coef) | Lower 95 % | Upper 95 % | Pr(>\|z\|) | p.adj |
|---|---|---|---|---|---|
| 2-Palmitoylglycerol | 1.14 | 0.771 | 1.67 | 0.52 | 0.762 |
| Aminomalonic acid | 1.13 | 0.773 | 1.65 | 0.528 | 0.762 |
| Citric acid | 0.896 | 0.626 | 1.28 | 0.549 | 0.777 |
| Cholesterol | 0.896 | 0.619 | 1.3 | 0.56 | 0.777 |
| Ribitol (1) | 1.11 | 0.764 | 1.62 | 0.575 | 0.784 |
| 4-Deoxytetronic acid (2) | 0.908 | 0.642 | 1.29 | 0.587 | 0.787 |
| Succinic acid | 1.1 | 0.758 | 1.61 | 0.607 | 0.788 |
| Hydroxyproline | 1.1 | 0.755 | 1.61 | 0.609 | 0.788 |
| 1-Monopalmitin | 0.93 | 0.687 | 1.26 | 0.641 | 0.814 |
| Glycine | 1.09 | 0.752 | 1.57 | 0.655 | 0.818 |
| Octanoic acid | 0.942 | 0.679 | 1.31 | 0.721 | 0.879 |
| Pyruvic acid | 1.07 | 0.735 | 1.56 | 0.727 | 0.879 |
| Arachidonic acid | 1.06 | 0.723 | 1.56 | 0.758 | 0.886 |
| Valine | 0.948 | 0.661 | 1.36 | 0.771 | 0.886 |
| Pyroglutamic acid | 0.952 | 0.672 | 1.35 | 0.78 | 0.886 |
| Myo inositol | 1.05 | 0.725 | 1.53 | 0.784 | 0.886 |
| Nonadecanoic acid | 0.956 | 0.671 | 1.36 | 0.802 | 0.886 |
| Serine | 0.954 | 0.66 | 1.38 | 0.803 | 0.886 |
| Tyrosine | 1.05 | 0.696 | 1.57 | 0.827 | 0.899 |
| Malic acid | 1.04 | 0.713 | 1.51 | 0.843 | 0.904 |
| alpha-Tocopherol | 1.01 | 0.695 | 1.47 | 0.959 | 0.982 |
| 4-Hydroxybutanoic acid | 0.995 | 0.689 | 1.44 | 0.978 | 0.982 |
| Threonine | 0.996 | 0.686 | 1.44 | 0.981 | 0.982 |
| alpha-ketoglutaric acid | 0.996 | 0.694 | 1.43 | 0.981 | 0.982 |
| Heptadecanoic acid (1) | 0.996 | 0.69 | 1.44 | 0.982 | 0.982 |

###### 5.2.1.3 Top-Metabolite from Cross-Sectional Analysis

```
## Call:
## survival::coxph(formula = survival::Surv(time = Amputation.tdiff,
##     event = Amputation.from.DATE) ~ Ribonic_acid, data = data.km)
##
##   n= 614, number of events= 28
##    (23 observations deleted due to missingness)
##
##                coef exp(coef) se(coef)     z Pr(>|z|)
## Ribonic_acid 0.4095    1.5060   0.2195 1.865   0.0622 .
## ---
## Signif. codes:  0 '***' 0.001 '**' 0.01 '*' 0.05 '.' 0.1 ' ' 1
##
##              exp(coef) exp(-coef) lower .95 upper .95
## Ribonic_acid     1.506      0.664    0.9794     2.316
##
## Concordance= 0.611  (se = 0.05 )
## Likelihood ratio test= 3.7  on 1 df,   p=0.05
## Wald test            = 3.48  on 1 df,   p=0.06
## Score (logrank) test = 3.29  on 1 df,   p=0.07
```

###### 5.2.1.4 Kaplan-Maier Curve with Median Cutpoint

- Top metabolite from cross-sectional analysis

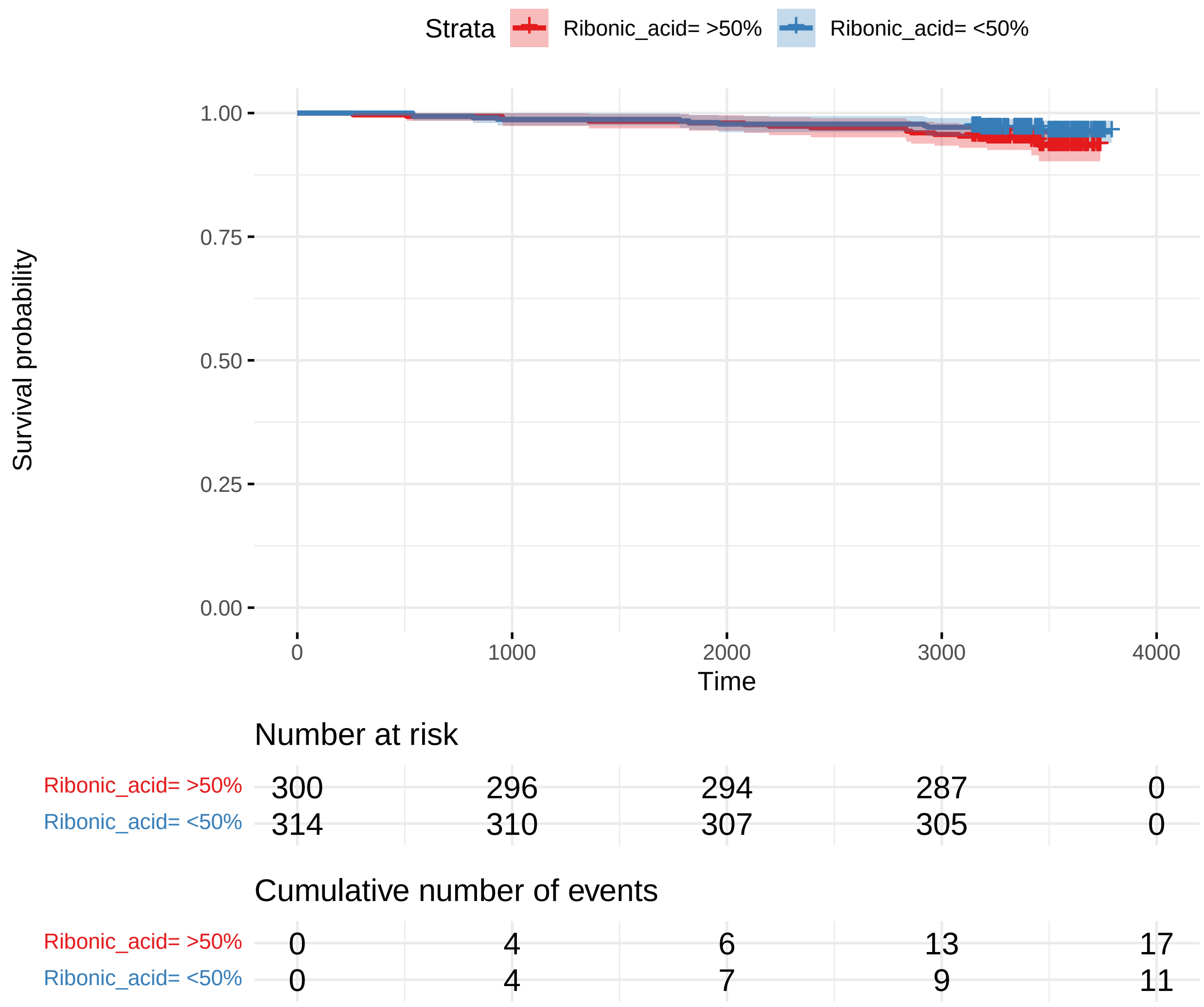

##### 5.2.2 Adusted Model

###### 5.2.2.1 Forest Plot of Model Coefficients

```
## NULL
```

###### 5.2.2.2 Table with All Metabolites

| Name | exp(coef) | Lower 95 % | Upper 95 % | Pr(>\|z\|) | p.adj |
|---|---|---|---|---|---|
| 2-Hydroxybutyric acid | 0.582 | 0.412 | 0.823 | 0.00216 | 0.162 |
| Palmitic acid | 0.632 | 0.456 | 0.876 | 0.00589 | 0.221 |
| 4-Hydroxybenzeneacetic acid | 1.73 | 1.08 | 2.76 | 0.0221 | 0.43 |
| Lactic acid | 0.7 | 0.511 | 0.958 | 0.026 | 0.43 |
| Oleic acid | 0.638 | 0.426 | 0.954 | 0.0286 | 0.43 |
| 3-Hydroxybutyric acid | 0.646 | 0.425 | 0.982 | 0.0408 | 0.46 |
| Stearic acid | 0.69 | 0.482 | 0.988 | 0.0429 | 0.46 |
| 4-Deoxytetronic acid (1) | 1.51 | 0.987 | 2.3 | 0.0573 | 0.537 |
| Glycerol (2) | 0.715 | 0.5 | 1.02 | 0.0669 | 0.557 |
| Aminomalonic acid | 1.44 | 0.94 | 2.21 | 0.094 | 0.615 |
| Glycerol (1) | 0.827 | 0.662 | 1.03 | 0.0951 | 0.615 |
| Benzeneacetic acid | 1.5 | 0.927 | 2.44 | 0.0983 | 0.615 |
| Myristoleic acid | 0.756 | 0.525 | 1.09 | 0.134 | 0.626 |
| Decanoic acid | 0.788 | 0.575 | 1.08 | 0.138 | 0.626 |
| Ribonic acid | 1.36 | 0.886 | 2.08 | 0.16 | 0.626 |
| Creatinine | 1.33 | 0.892 | 1.97 | 0.163 | 0.626 |
| Ribitol (2) | 1.37 | 0.878 | 2.12 | 0.166 | 0.626 |
| Hydroxylamine | 0.821 | 0.615 | 1.1 | 0.181 | 0.626 |
| Linoleic acid | 0.817 | 0.606 | 1.1 | 0.185 | 0.626 |
| 11-Eicosenoic acid | 0.819 | 0.607 | 1.1 | 0.189 | 0.626 |
| Methionine | 1.33 | 0.867 | 2.05 | 0.189 | 0.626 |
| 4-Deoxytetronic acid (2) | 0.781 | 0.538 | 1.13 | 0.192 | 0.626 |
| Ethanolamine | 1.29 | 0.882 | 1.87 | 0.192 | 0.626 |
| Arachidic acid | 0.772 | 0.51 | 1.17 | 0.222 | 0.685 |
| 2-hydroxy Isovaleric acid | 0.813 | 0.58 | 1.14 | 0.228 | 0.685 |
| Arabinopyranose | 1.33 | 0.814 | 2.18 | 0.254 | 0.695 |
| Glycine | 1.25 | 0.852 | 1.83 | 0.255 | 0.695 |
| 2,4-Dihydroxybutanoic acid | 1.26 | 0.838 | 1.9 | 0.265 | 0.695 |
| Dodecanoic acid | 0.809 | 0.556 | 1.18 | 0.269 | 0.695 |
| Campesterol | 0.824 | 0.561 | 1.21 | 0.324 | 0.79 |
| Heptadecanoic acid (2) | 0.848 | 0.606 | 1.19 | 0.337 | 0.79 |
| Nonanoic acid | 0.873 | 0.662 | 1.15 | 0.337 | 0.79 |
| Tridecanoic acid | 0.883 | 0.666 | 1.17 | 0.385 | 0.868 |
| L-5-Oxoproline | 0.847 | 0.579 | 1.24 | 0.394 | 0.868 |
| Valine | 0.857 | 0.572 | 1.28 | 0.453 | 0.947 |
| Eicosapentaenoic acid | 1.17 | 0.76 | 1.8 | 0.474 | 0.947 |
| Phenylalanine | 1.15 | 0.775 | 1.7 | 0.489 | 0.947 |
| 1,3-Propanediol | 0.911 | 0.687 | 1.21 | 0.518 | 0.947 |
| Docosahexaenoic acid | 0.901 | 0.651 | 1.25 | 0.529 | 0.947 |
| 3,4-Dihydroxybutanoic acid | 1.13 | 0.758 | 1.68 | 0.548 | 0.947 |
| 1-Monopalmitin | 0.912 | 0.671 | 1.24 | 0.555 | 0.947 |
| Citric acid | 0.893 | 0.603 | 1.32 | 0.572 | 0.947 |
| Bisphenol A | 0.895 | 0.6 | 1.34 | 0.588 | 0.947 |
| Threonine | 1.13 | 0.719 | 1.78 | 0.596 | 0.947 |
| 3-Indolepropionic acid | 0.917 | 0.663 | 1.27 | 0.602 | 0.947 |
| Glyceryl-glycoside | 1.13 | 0.699 | 1.82 | 0.62 | 0.947 |
| Serine | 1.1 | 0.725 | 1.68 | 0.644 | 0.947 |
| Succinic acid | 1.09 | 0.738 | 1.61 | 0.665 | 0.947 |
| Leucine | 1.1 | 0.7 | 1.72 | 0.688 | 0.947 |
| Alanine | 1.09 | 0.719 | 1.64 | 0.694 | 0.947 |

| Name | exp(coef) | Lower 95 % | Upper 95 % | Pr(>\|z\|) | p.adj |
|---|---|---|---|---|---|
| 4-Hydroxybutanoic acid | 1.08 | 0.736 | 1.58 | 0.695 | 0.947 |
| 1-Dodecanol | 0.933 | 0.649 | 1.34 | 0.706 | 0.947 |
| Isoleucine | 1.09 | 0.686 | 1.74 | 0.707 | 0.947 |
| Myo inositol | 0.932 | 0.637 | 1.36 | 0.716 | 0.947 |
| Cholesterol | 0.922 | 0.591 | 1.44 | 0.719 | 0.947 |
| 2-Palmitoylglycerol | 1.07 | 0.711 | 1.62 | 0.736 | 0.947 |
| 4-Hydroxyphenyllactic acid | 1.08 | 0.693 | 1.67 | 0.746 | 0.947 |
| Octanoic acid | 0.947 | 0.63 | 1.42 | 0.793 | 0.947 |
| Tartronic acid | 0.95 | 0.648 | 1.39 | 0.793 | 0.947 |
| Ribitol (1) | 0.951 | 0.65 | 1.39 | 0.798 | 0.947 |
| alpha-ketoglutaric acid | 0.962 | 0.708 | 1.31 | 0.802 | 0.947 |
| Fumaric acid | 1.05 | 0.71 | 1.56 | 0.803 | 0.947 |
| Glyceric acid | 1.04 | 0.746 | 1.45 | 0.811 | 0.947 |
| Arachidonic acid | 1.04 | 0.746 | 1.44 | 0.83 | 0.947 |
| Malic acid | 0.968 | 0.678 | 1.38 | 0.858 | 0.947 |
| Tyrosine | 1.04 | 0.694 | 1.55 | 0.862 | 0.947 |
| Nonadecanoic acid | 0.967 | 0.642 | 1.46 | 0.871 | 0.947 |
| Hydroxyproline | 1.03 | 0.691 | 1.55 | 0.872 | 0.947 |
| Pyruvic acid | 0.97 | 0.665 | 1.41 | 0.873 | 0.947 |
| Pyroglutamic acid | 0.973 | 0.658 | 1.44 | 0.89 | 0.947 |
| 3-Indoleacetic acid | 1.03 | 0.684 | 1.54 | 0.897 | 0.947 |
| Glutamic acid | 1.02 | 0.658 | 1.58 | 0.933 | 0.963 |
| Proline | 1.02 | 0.674 | 1.53 | 0.937 | 0.963 |
| Heptadecanoic acid (1) | 0.994 | 0.68 | 1.45 | 0.976 | 0.989 |
| alpha-Tocopherol | 1 | 0.671 | 1.49 | 0.997 | 0.997 |

###### 5.2.2.3 Top-Metabolite from Cross-Sectional Analysis

```
## Call:
## survival::coxph(formula = survival::Surv(time = Amputation.tdiff,
##     event = Amputation.from.DATE) ~ Ribonic_acid + Age.x + Gender.x +
##     Hba1c_baseline + CALSBP + bmi + Smoking + Statin + log_Blood_TGA +
##     Total_cholesterol, data = data.km)
##
##   n= 600, number of events= 28
##    (37 observations deleted due to missingness)
##
##                        coef exp(coef) se(coef)      z Pr(>|z|)
## Ribonic_acid       0.31105   1.36486  0.22151  1.404   0.1602
## Age.x              0.02766   1.02805  0.01868  1.481   0.1387
## Gender.x           0.61355   1.84697  0.42533  1.443   0.1492
## Hba1c_baseline     0.25261   1.28738  0.14517  1.740   0.0818 .
## CALSBP             0.01409   1.01419  0.01051  1.341   0.1800
## bmi                0.05329   1.05473  0.04632  1.150   0.2500
## Smoking            0.43811   1.54978  0.43587  1.005   0.3148
## Statin             0.67232   1.95877  0.52152  1.289   0.1973
## log_Blood_TGA      0.31301   1.36754  0.30302  1.033   0.3016
## Total_cholesterol -0.02248   0.97777  0.22169 -0.101   0.9192
## ---
## Signif. codes:  0 '***' 0.001 '**' 0.01 '*' 0.05 '.' 0.1 ' ' 1
##
##                   exp(coef) exp(-coef) lower .95 upper .95
## Ribonic_acid         1.3649     0.7327    0.8842     2.107
## Age.x                1.0280     0.9727    0.9911     1.066
## Gender.x             1.8470     0.5414    0.8024     4.251
## Hba1c_baseline       1.2874     0.7768    0.9686     1.711
## CALSBP               1.0142     0.9860    0.9935     1.035
## bmi                  1.0547     0.9481    0.9632     1.155
## Smoking              1.5498     0.6453    0.6596     3.641
## Statin               1.9588     0.5105    0.7048     5.444
## log_Blood_TGA        1.3675     0.7312    0.7551     2.477
## Total_cholesterol    0.9778     1.0227    0.6332     1.510
##
## Concordance= 0.764  (se = 0.036 )
## Likelihood ratio test= 26.61  on 10 df,   p=0.003
## Wald test            = 24.23  on 10 df,   p=0.007
## Score (logrank) test = 25.34  on 10 df,   p=0.005
```

###### 5.2.2.3.1 Forest Plot with Clinical Variables

- Top metabolite from cross-sectional analysis

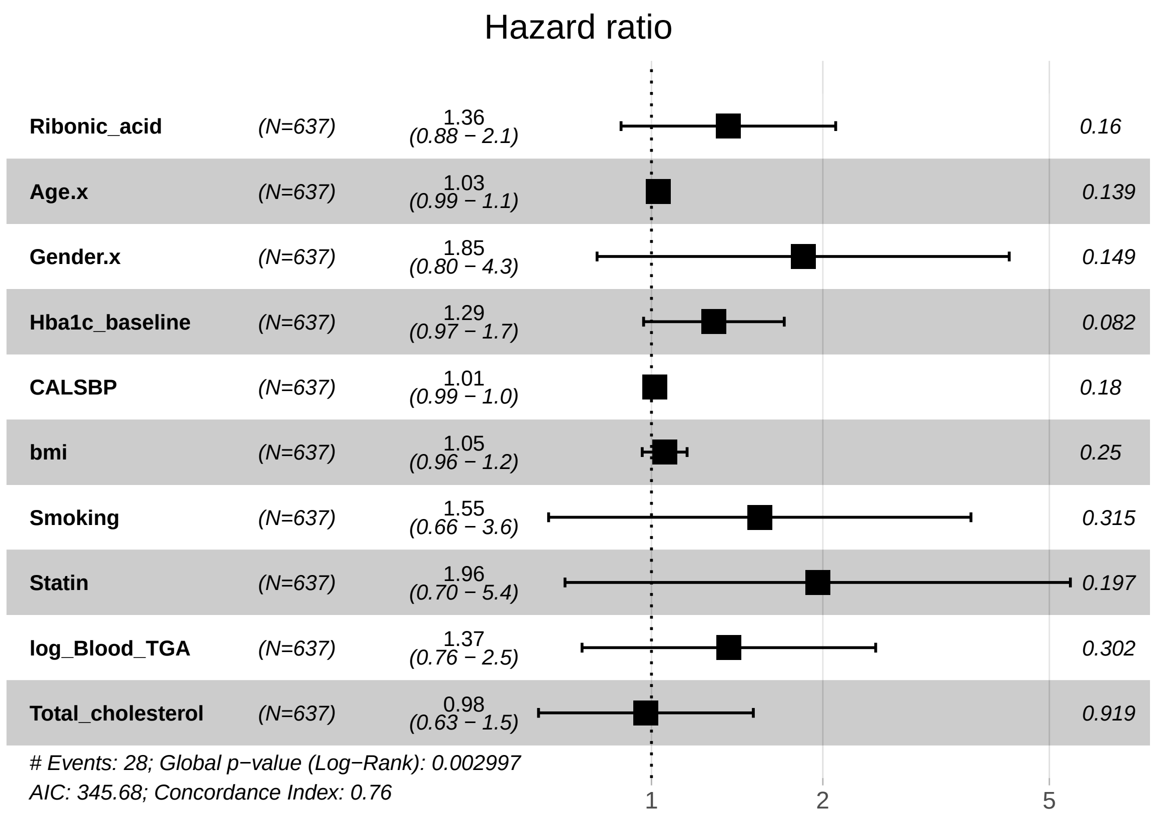

##### 5.2.3 Adusted Model with eGFR

###### 5.2.3.1 Forest Plot of Model Coefficients

```
## NULL
```

###### 5.2.3.2 Table with All Metabolites

| Name | exp(coef) | Lower 95 % | Upper 95 % | Pr(>\|z\|) | p.adj |
|---|---|---|---|---|---|
| Palmitic acid | 0.625 | 0.435 | 0.897 | 0.0109 | 0.434 |
| 2-Hydroxybutyric acid | 0.632 | 0.438 | 0.91 | 0.0137 | 0.434 |
| 4-Deoxytetronic acid (2) | 0.643 | 0.446 | 0.925 | 0.0174 | 0.434 |
| 3-Hydroxybutyric acid | 0.632 | 0.413 | 0.967 | 0.0343 | 0.489 |
| Oleic acid | 0.659 | 0.444 | 0.978 | 0.0384 | 0.489 |
| Lactic acid | 0.74 | 0.554 | 0.99 | 0.0423 | 0.489 |
| Glycerol (2) | 0.702 | 0.496 | 0.993 | 0.0456 | 0.489 |
| Myo inositol | 0.675 | 0.444 | 1.03 | 0.0653 | 0.612 |
| Stearic acid | 0.718 | 0.499 | 1.03 | 0.0748 | 0.624 |
| Methionine | 1.46 | 0.945 | 2.24 | 0.0888 | 0.666 |
| Aminomalonic acid | 1.43 | 0.924 | 2.22 | 0.108 | 0.726 |
| 4-Hydroxybenzeneacetic acid | 1.5 | 0.904 | 2.5 | 0.116 | 0.726 |
| Decanoic acid | 0.79 | 0.573 | 1.09 | 0.15 | 0.741 |
| Benzeneacetic acid | 1.39 | 0.867 | 2.24 | 0.17 | 0.741 |
| Ethanolamine | 1.32 | 0.887 | 1.96 | 0.172 | 0.741 |
| 11-Eicosenoic acid | 0.817 | 0.607 | 1.1 | 0.181 | 0.741 |
| Glycerol (1) | 0.861 | 0.688 | 1.08 | 0.192 | 0.741 |
| 4-Deoxytetronic acid (1) | 1.34 | 0.863 | 2.09 | 0.192 | 0.741 |
| Myristoleic acid | 0.794 | 0.56 | 1.13 | 0.196 | 0.741 |
| Linoleic acid | 0.829 | 0.62 | 1.11 | 0.204 | 0.741 |
| Nonanoic acid | 0.84 | 0.637 | 1.11 | 0.217 | 0.741 |
| Arachidic acid | 0.773 | 0.514 | 1.16 | 0.217 | 0.741 |
| Arabinopyranose | 1.35 | 0.817 | 2.23 | 0.243 | 0.772 |
| Dodecanoic acid | 0.808 | 0.559 | 1.17 | 0.254 | 0.772 |
| Hydroxylamine | 0.852 | 0.645 | 1.12 | 0.257 | 0.772 |
| Campesterol | 0.808 | 0.553 | 1.18 | 0.268 | 0.772 |
| Citric acid | 0.809 | 0.541 | 1.21 | 0.303 | 0.824 |
| Eicosapentaenoic acid | 1.26 | 0.808 | 1.97 | 0.308 | 0.824 |
| 2-hydroxy Isovaleric acid | 0.843 | 0.589 | 1.21 | 0.351 | 0.9 |
| Tridecanoic acid | 0.875 | 0.657 | 1.16 | 0.36 | 0.9 |
| L-5-Oxoproline | 0.841 | 0.573 | 1.23 | 0.375 | 0.904 |
| Heptadecanoic acid (2) | 0.863 | 0.619 | 1.2 | 0.386 | 0.904 |
| Isoleucine | 1.22 | 0.762 | 1.97 | 0.403 | 0.917 |
| 1,3-Propanediol | 0.893 | 0.66 | 1.21 | 0.46 | 0.977 |
| Glycine | 1.15 | 0.778 | 1.71 | 0.476 | 0.977 |
| Leucine | 1.16 | 0.754 | 1.78 | 0.503 | 0.977 |
| Serine | 1.15 | 0.755 | 1.74 | 0.522 | 0.977 |
| Pyroglutamic acid | 0.883 | 0.6 | 1.3 | 0.528 | 0.977 |
| Threonine | 1.14 | 0.733 | 1.79 | 0.554 | 0.977 |
| Creatinine | 1.13 | 0.745 | 1.72 | 0.563 | 0.977 |
| Phenylalanine | 1.11 | 0.755 | 1.65 | 0.584 | 0.977 |
| 1-Monopalmitin | 0.916 | 0.668 | 1.26 | 0.587 | 0.977 |
| Glyceric acid | 1.1 | 0.775 | 1.56 | 0.593 | 0.977 |
| Ribonic acid | 1.13 | 0.718 | 1.78 | 0.594 | 0.977 |
| Docosahexaenoic acid | 0.919 | 0.661 | 1.28 | 0.618 | 0.977 |
| Tyrosine | 1.11 | 0.719 | 1.72 | 0.634 | 0.977 |
| Bisphenol A | 0.91 | 0.613 | 1.35 | 0.639 | 0.977 |
| 2-Palmitoylglycerol | 1.1 | 0.725 | 1.67 | 0.653 | 0.977 |
| 4-Hydroxybutanoic acid | 1.09 | 0.743 | 1.59 | 0.67 | 0.977 |
| 3,4-Dihydroxybutanoic acid | 0.91 | 0.586 | 1.41 | 0.675 | 0.977 |

| Name | exp(coef) | Lower 95 % | Upper 95 % | Pr(>\|z\|) | p.adj |
|---|---|---|---|---|---|
| Ribitol (2) | 1.1 | 0.683 | 1.78 | 0.691 | 0.977 |
| Malic acid | 0.935 | 0.662 | 1.32 | 0.702 | 0.977 |
| Succinic acid | 1.07 | 0.722 | 1.59 | 0.732 | 0.977 |
| Arachidonic acid | 1.06 | 0.745 | 1.52 | 0.737 | 0.977 |
| alpha-ketoglutaric acid | 0.948 | 0.69 | 1.3 | 0.741 | 0.977 |
| Valine | 0.936 | 0.623 | 1.41 | 0.75 | 0.977 |
| 3-Indoleacetic acid | 0.935 | 0.611 | 1.43 | 0.758 | 0.977 |
| Hydroxyproline | 0.941 | 0.622 | 1.42 | 0.772 | 0.977 |
| 3-Indolepropionic acid | 0.954 | 0.689 | 1.32 | 0.776 | 0.977 |
| Ribitol (1) | 0.952 | 0.646 | 1.4 | 0.802 | 0.977 |
| Tartronic acid | 0.954 | 0.661 | 1.38 | 0.802 | 0.977 |
| 1-Dodecanol | 0.955 | 0.66 | 1.38 | 0.808 | 0.977 |
| Octanoic acid | 0.956 | 0.64 | 1.43 | 0.826 | 0.983 |
| Pyruvic acid | 0.966 | 0.662 | 1.41 | 0.855 | 0.988 |
| Proline | 0.969 | 0.65 | 1.45 | 0.878 | 0.988 |
| Cholesterol | 0.966 | 0.62 | 1.51 | 0.879 | 0.988 |
| Nonadecanoic acid | 0.975 | 0.654 | 1.45 | 0.899 | 0.988 |
| 2,4-Dihydroxybutanoic acid | 1.02 | 0.658 | 1.6 | 0.913 | 0.988 |
| Glutamic acid | 1.02 | 0.668 | 1.57 | 0.917 | 0.988 |
| 4-Hydroxyphenyllactic acid | 0.982 | 0.634 | 1.52 | 0.933 | 0.988 |
| Glyceryl-glycoside | 1.02 | 0.642 | 1.62 | 0.936 | 0.988 |
| Alanine | 1.01 | 0.672 | 1.52 | 0.961 | 0.992 |
| Fumaric acid | 0.992 | 0.668 | 1.47 | 0.968 | 0.992 |
| Heptadecanoic acid (1) | 0.998 | 0.69 | 1.44 | 0.992 | 0.992 |
| alpha-Tocopherol | 1 | 0.681 | 1.47 | 0.992 | 0.992 |

###### 5.2.3.3 Top-Metabolite from Cross-Sectional Analysis

```
## Call:
## survival::coxph(formula = survival::Surv(time = Amputation.tdiff,
##     event = Amputation.from.DATE) ~ Ribonic_acid + Age.x + Gender.x +
##     Hba1c_baseline + CALSBP + bmi + Smoking + Statin + log_Blood_TGA +
##     Total_cholesterol + egfr, data = data.km)
##
##   n= 598, number of events= 28
##    (39 observations deleted due to missingness)
##
##                         coef exp(coef)  se(coef)      z Pr(>|z|)
## Ribonic_acid       0.125827  1.134086  0.236202  0.533   0.5942
## Age.x              0.024095  1.024387  0.018670  1.291   0.1969
## Gender.x           0.737364  2.090418  0.430898  1.711   0.0870 .
## Hba1c_baseline     0.254205  1.289436  0.147353  1.725   0.0845 .
## CALSBP             0.012769  1.012850  0.010462  1.220   0.2223
## bmi                0.057599  1.059291  0.045924  1.254   0.2098
## Smoking            0.515193  1.673961  0.436684  1.180   0.2381
## Statin             0.579576  1.785282  0.522730  1.109   0.2675
## log_Blood_TGA      0.265712  1.304359  0.307168  0.865   0.3870
## Total_cholesterol  0.007319  1.007346  0.217403  0.034   0.9731
## egfr              -0.013544  0.986547  0.008162 -1.659   0.0970 .
## ---
## Signif. codes:  0 '***' 0.001 '**' 0.01 '*' 0.05 '.' 0.1 ' ' 1
##
##                   exp(coef) exp(-coef) lower .95 upper .95
## Ribonic_acid         1.1341     0.8818    0.7138     1.802
## Age.x                1.0244     0.9762    0.9876     1.063
## Gender.x             2.0904     0.4784    0.8984     4.864
## Hba1c_baseline       1.2894     0.7755    0.9660     1.721
## CALSBP               1.0129     0.9873    0.9923     1.034
## bmi                  1.0593     0.9440    0.9681     1.159
## Smoking              1.6740     0.5974    0.7113     3.940
## Statin               1.7853     0.5601    0.6409     4.973
## log_Blood_TGA        1.3044     0.7667    0.7144     2.382
## Total_cholesterol    1.0073     0.9927    0.6578     1.543
## egfr                 0.9865     1.0136    0.9709     1.002
##
## Concordance= 0.778  (se = 0.032 )
## Likelihood ratio test= 29.28  on 11 df,   p=0.002
## Wald test            = 25.78  on 11 df,   p=0.007
## Score (logrank) test = 28.46  on 11 df,   p=0.003
```

###### 5.2.3.3.1 Forest Plot with Clinical Variables

- Top metabolite from cross-sectional analysis

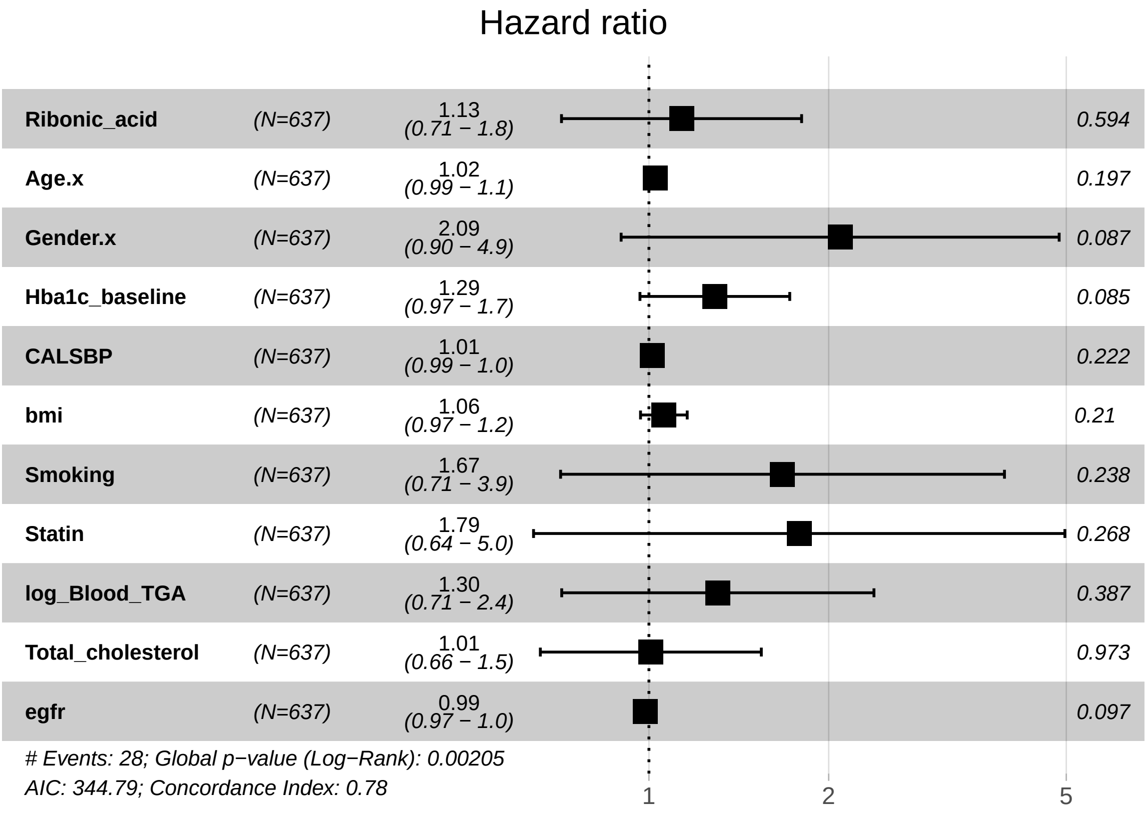

##### 5.2.4 Fully Adusted Model

###### 5.2.4.1 Forest Plot of Model Coefficients

```
## NULL
```

###### 5.2.4.2 Table with All Metabolites

| Name | exp(coef) | Lower 95 % | Upper 95 % | Pr(>\|z\|) | p.adj |
|---|---|---|---|---|---|
| Palmitic acid | 0.585 | 0.407 | 0.841 | 0.00381 | 0.268 |
| Glycerol (2) | 0.649 | 0.463 | 0.91 | 0.0123 | 0.268 |
| 3-Hydroxybutyric acid | 0.581 | 0.378 | 0.895 | 0.0136 | 0.268 |
| 4-Deoxytetronic acid (2) | 0.637 | 0.442 | 0.917 | 0.0151 | 0.268 |
| 2-Hydroxybutyric acid | 0.639 | 0.442 | 0.926 | 0.0179 | 0.268 |
| Oleic acid | 0.625 | 0.418 | 0.934 | 0.022 | 0.275 |
| Myo inositol | 0.628 | 0.417 | 0.946 | 0.0262 | 0.28 |
| Lactic acid | 0.732 | 0.549 | 0.976 | 0.0335 | 0.314 |
| Stearic acid | 0.686 | 0.474 | 0.994 | 0.0467 | 0.389 |
| Decanoic acid | 0.745 | 0.547 | 1.02 | 0.0628 | 0.471 |
| 11-Eicosenoic acid | 0.718 | 0.497 | 1.04 | 0.0782 | 0.533 |
| Methionine | 1.48 | 0.946 | 2.31 | 0.0865 | 0.541 |
| Nonanoic acid | 0.8 | 0.614 | 1.04 | 0.0976 | 0.563 |
| Aminomalonic acid | 1.43 | 0.911 | 2.25 | 0.12 | 0.595 |
| Dodecanoic acid | 0.762 | 0.531 | 1.09 | 0.138 | 0.595 |
| Ethanolamine | 1.36 | 0.902 | 2.04 | 0.143 | 0.595 |
| Myristoleic acid | 0.776 | 0.552 | 1.09 | 0.145 | 0.595 |
| Citric acid | 0.74 | 0.492 | 1.11 | 0.148 | 0.595 |
| Linoleic acid | 0.816 | 0.618 | 1.08 | 0.151 | 0.595 |
| Arachidic acid | 0.745 | 0.491 | 1.13 | 0.167 | 0.605 |
| 4-Hydroxybenzeneacetic acid | 1.42 | 0.857 | 2.36 | 0.173 | 0.605 |
| Benzeneacetic acid | 1.4 | 0.859 | 2.28 | 0.177 | 0.605 |
| 2-hydroxy Isovaleric acid | 0.801 | 0.556 | 1.15 | 0.234 | 0.737 |
| Campesterol | 0.791 | 0.537 | 1.17 | 0.236 | 0.737 |
| Glycerol (1) | 0.871 | 0.688 | 1.1 | 0.252 | 0.756 |
| L-5-Oxoproline | 0.808 | 0.553 | 1.18 | 0.271 | 0.782 |
| Heptadecanoic acid (2) | 0.833 | 0.595 | 1.17 | 0.288 | 0.799 |
| Hydroxylamine | 0.863 | 0.649 | 1.15 | 0.309 | 0.829 |
| Tridecanoic acid | 0.866 | 0.648 | 1.16 | 0.333 | 0.837 |
| 4-Deoxytetronic acid (1) | 1.25 | 0.796 | 1.96 | 0.335 | 0.837 |
| Arabinopyranose | 1.26 | 0.765 | 2.07 | 0.365 | 0.878 |
| 3,4-Dihydroxybutanoic acid | 0.829 | 0.531 | 1.29 | 0.408 | 0.878 |
| 1,3-Propanediol | 0.88 | 0.648 | 1.2 | 0.414 | 0.878 |
| Malic acid | 0.87 | 0.62 | 1.22 | 0.422 | 0.878 |
| Pyroglutamic acid | 0.855 | 0.581 | 1.26 | 0.425 | 0.878 |
| Serine | 1.19 | 0.775 | 1.81 | 0.434 | 0.878 |
| Eicosapentaenoic acid | 1.2 | 0.758 | 1.89 | 0.44 | 0.878 |
| Isoleucine | 1.2 | 0.753 | 1.91 | 0.445 | 0.878 |
| Threonine | 1.17 | 0.741 | 1.85 | 0.498 | 0.92 |
| Leucine | 1.15 | 0.752 | 1.75 | 0.526 | 0.92 |
| 1-Monopalmitin | 0.912 | 0.67 | 1.24 | 0.56 | 0.92 |
| Bisphenol A | 0.89 | 0.596 | 1.33 | 0.567 | 0.92 |
| Docosahexaenoic acid | 0.909 | 0.653 | 1.26 | 0.569 | 0.92 |
| Ribitol (1) | 0.894 | 0.607 | 1.32 | 0.569 | 0.92 |
| Phenylalanine | 1.12 | 0.753 | 1.66 | 0.58 | 0.92 |
| Fumaric acid | 0.902 | 0.606 | 1.34 | 0.613 | 0.92 |
| 3-Indoleacetic acid | 0.894 | 0.579 | 1.38 | 0.613 | 0.92 |
| Tyrosine | 1.12 | 0.715 | 1.76 | 0.619 | 0.92 |
| Glycine | 1.1 | 0.737 | 1.65 | 0.636 | 0.92 |
| 2-Palmitoylglycerol | 1.1 | 0.722 | 1.67 | 0.661 | 0.92 |

| Name | exp(coef) | Lower 95 % | Upper 95 % | Pr(>\|z\|) | p.adj |
|---|---|---|---|---|---|
| Valine | 0.913 | 0.606 | 1.38 | 0.663 | 0.92 |
| Creatinine | 1.1 | 0.723 | 1.66 | 0.667 | 0.92 |
| 1-Dodecanol | 0.922 | 0.634 | 1.34 | 0.669 | 0.92 |
| Cholesterol | 0.909 | 0.579 | 1.43 | 0.677 | 0.92 |
| Tartronic acid | 0.924 | 0.637 | 1.34 | 0.679 | 0.92 |
| 3-Indolepropionic acid | 0.94 | 0.68 | 1.3 | 0.706 | 0.92 |
| 2,4-Dihydroxybutanoic acid | 0.921 | 0.578 | 1.47 | 0.729 | 0.92 |
| Nonadecanoic acid | 0.934 | 0.633 | 1.38 | 0.733 | 0.92 |
| Glyceric acid | 1.06 | 0.736 | 1.53 | 0.752 | 0.92 |
| alpha-ketoglutaric acid | 0.95 | 0.687 | 1.31 | 0.755 | 0.92 |
| Glyceryl-glycoside | 0.934 | 0.599 | 1.46 | 0.764 | 0.92 |
| 4-Hydroxyphenyllactic acid | 0.934 | 0.59 | 1.48 | 0.772 | 0.92 |
| Arachidonic acid | 1.06 | 0.734 | 1.52 | 0.773 | 0.92 |
| Hydroxyproline | 0.944 | 0.618 | 1.44 | 0.788 | 0.923 |
| Octanoic acid | 0.955 | 0.65 | 1.4 | 0.813 | 0.938 |
| Ribonic acid | 1.04 | 0.665 | 1.62 | 0.868 | 0.967 |
| alpha-Tocopherol | 0.969 | 0.656 | 1.43 | 0.875 | 0.967 |
| Alanine | 1.03 | 0.68 | 1.57 | 0.882 | 0.967 |
| Proline | 0.974 | 0.65 | 1.46 | 0.898 | 0.967 |
| Pyruvic acid | 0.976 | 0.666 | 1.43 | 0.902 | 0.967 |
| 4-Hydroxybutanoic acid | 1.02 | 0.694 | 1.5 | 0.92 | 0.972 |
| Succinic acid | 1.02 | 0.682 | 1.51 | 0.939 | 0.978 |
| Glutamic acid | 1.01 | 0.657 | 1.56 | 0.952 | 0.978 |
| Heptadecanoic acid (1) | 0.994 | 0.682 | 1.45 | 0.974 | 0.986 |
| Ribitol (2) | 0.996 | 0.628 | 1.58 | 0.986 | 0.986 |

###### 5.2.4.3 Top-Metabolite from Cross-Sectional Analysis

```
## Call:
## survival::coxph(formula = survival::Surv(time = Amputation.tdiff,
##     event = Amputation.from.DATE) ~ Ribonic_acid + Age.x + Gender.x +
##     Hba1c_baseline + CALSBP + bmi + Smoking + Statin + log_Blood_TGA +
##     Total_cholesterol + egfr + logUAER, data = data.km)
##
##   n= 564, number of events= 27
##    (73 observations deleted due to missingness)
##
##                       coef exp(coef)  se(coef)      z Pr(>|z|)
## Ribonic_acid      0.038446  1.039194  0.230982  0.166   0.8678
## Age.x             0.020683  1.020899  0.020006  1.034   0.3012
## Gender.x          0.709210  2.032385  0.441730  1.606   0.1084
## Hba1c_baseline    0.284992  1.329752  0.149098  1.911   0.0559 .
## CALSBP            0.010211  1.010263  0.011381  0.897   0.3696
## bmi               0.046139  1.047220  0.047343  0.975   0.3298
## Smoking           0.492379  1.636205  0.457500  1.076   0.2818
## Statin            0.471402  1.602239  0.531747  0.887   0.3753
## log_Blood_TGA     0.192918  1.212784  0.315749  0.611   0.5412
## Total_cholesterol  0.004336  1.004345  0.220526  0.020   0.9843
## egfr             -0.016330  0.983803  0.008993 -1.816   0.0694 .
## logUAER           0.024631  1.024936  0.089073  0.277   0.7821
## ---
## Signif. codes:  0 '***' 0.001 '**' 0.01 '*' 0.05 '.' 0.1 ' ' 1
##
##                  exp(coef) exp(-coef) lower .95 upper .95
## Ribonic_acid         1.0392     0.9623    0.6608     1.634
## Age.x                1.0209     0.9795    0.9816     1.062
## Gender.x             2.0324     0.4920    0.8551     4.831
## Hba1c_baseline       1.3298     0.7520    0.9928     1.781
## CALSBP               1.0103     0.9898    0.9880     1.033
## bmi                  1.0472     0.9549    0.9544     1.149
## Smoking              1.6362     0.6112    0.6674     4.011
## Statin               1.6022     0.6241    0.5651     4.543
## log_Blood_TGA        1.2128     0.8245    0.6532     2.252
## Total_cholesterol    1.0043     0.9957    0.6519     1.547
## egfr                 0.9838     1.0165    0.9666     1.001
## logUAER              1.0249     0.9757    0.8608     1.220
##
## Concordance= 0.772  (se = 0.033 )
## Likelihood ratio test= 26.76  on 12 df,   p=0.008
## Wald test            = 24.21  on 12 df,   p=0.02
## Score (logrank) test = 26.96  on 12 df,   p=0.008
```

###### 5.2.4.3.1 Forest Plot with Clinical Variables

- Top metabolite from cross-sectional analysis

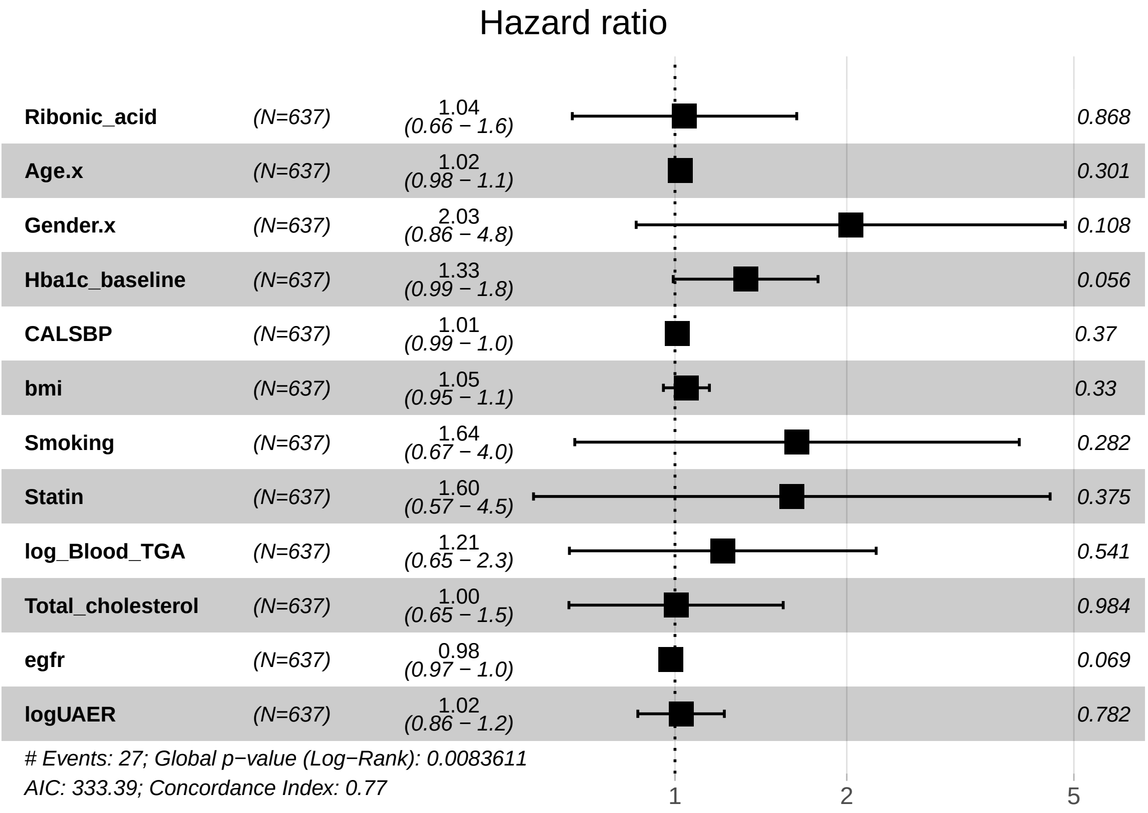

### 6 Ulcers

#### 6.1 Ulcer Diagnosis at DATE

##### 6.1.1 Crude Model

```
## [1] "Fitting models:"
## [1] "~ Ulcer.diagnosis.at.DATE"
## [1] ""
```

###### 6.1.1.1 Forest Plot of Model Coefficients

```
## NULL
```

###### 6.1.1.2 Tables of Model Coefficients

```
## [1] ""
## [1] "Table: Ulcer.diagnosis.at.DATEJA"
## [1] "  (from model: "
## [1] "   ~ Ulcer.diagnosis.at.DATE)"
## [1] ""
## [1] "No significant associations at p.adj < 0.2"
```

###### 6.1.1.3 Table with All Metabolites

```
## [1] ""
## [1] "Table: Ulcer.diagnosis.at.DATEJA"
## [1] "  (from model: "
## [1] "   ~ Ulcer.diagnosis.at.DATE)"
## [1] ""
##                               Name Coefficient    CI.L  CI.R p.value p.adj
## 1                      Ribitol; 71    0.563000  0.1070 1.020  0.0156 0.712
## 2             Stearic acid, TMS; 2    0.514000  0.0575 0.971  0.0273 0.712
## 3            Palmitic acid, TMS; 5    0.496000  0.0394 0.952  0.0333 0.712
## 4                     Glycerol; 58    0.441000 -0.0160 0.897  0.0586 0.712
## 5             Citric acid, 4TMS; 6   -0.421000 -0.8780 0.035  0.0704 0.712
## 6                 Ribonic acid; 72    0.415000 -0.0415 0.871  0.0748 0.712
## 7             Myristoleic acid; 65    0.413000 -0.0438 0.869  0.0764 0.712
## 8           Succinic acid, 2TMS; 7    0.365000 -0.0916 0.821  0.1170 0.712
## 9                 Serine, 3TMS; 14   -0.349000 -0.8050 0.108  0.1340 0.712
## 10            Malic acid, 3TMS; 11    0.346000 -0.1110 0.802  0.1380 0.712
## 11              L-5-Oxoproline; 63   -0.345000 -0.8010 0.112  0.1390 0.712
## 12           Nonadecanoic acid; 66    0.340000 -0.1160 0.797  0.1440 0.712
## 13                    Tyrosine; 75    0.332000 -0.1240 0.789  0.1540 0.712
## 14               Decanoic acid; 52    0.331000 -0.1260 0.787  0.1560 0.712
## 15 2-Hydroxybutyric acid, 2TMS; 2   -0.318000 -0.7750 0.138  0.1720 0.712
## 16              Arachidic acid; 46    0.313000 -0.1430 0.770  0.1790 0.712
## 17             1,3-Propanediol; 34    0.303000 -0.1530 0.760  0.1930 0.712
## 18          Heptadecanoic acid; 60    0.293000 -0.1640 0.749  0.2090 0.712
## 19  2-hydroxy Isovaleric acid; 38   -0.291000 -0.7470 0.166  0.2120 0.712
## 20            Myo inositol 6TMS; 1    0.287000 -0.1700 0.744  0.2180 0.712
## 21              Oleic acid, TMS; 3    0.287000 -0.1700 0.743  0.2180 0.712
## 22      Eicosapentaenoic acid; 55    0.275000 -0.1810 0.732  0.2370 0.712
## 23          Benzeneacetic acid; 47   -0.272000 -0.7280 0.185  0.2440 0.712
## 24               Proline, 2TMS; 21    0.269000 -0.1870 0.726  0.2470 0.712
## 25 alpha-ketoglutaric acid, TMS M    0.267000 -0.1900 0.723  0.2530 0.712
## 26            Methionine, 2TMS; 16    0.266000 -0.1910 0.722  0.2540 0.712
## 27               Nonanoic acid; 67    0.264000 -0.1920 0.721  0.2560 0.712
## 28          Heptadecanoic acid; 61    0.257000 -0.2000 0.713  0.2710 0.725
## 29           Aminomalonic acid; 45   -0.216000 -0.6730 0.240  0.3540 0.888
## 30 2,4-Dihydroxybutanoic acid; 28    0.215000 -0.2410 0.672  0.3550 0.888
## 31            Tridecanoic acid; 74    0.183000 -0.2730 0.640  0.4320 0.936
## 32            Cholesterol, TMS; 23   -0.178000 -0.6340 0.279  0.4450 0.936
## 33          11-Eicosenoic acid; 35    0.177000 -0.2800 0.633  0.4470 0.936
## 34               Glycine, 3TMS; 17   -0.175000 -0.6310 0.282  0.4530 0.936
## 35                Ethanolamine; 56    0.175000 -0.2820 0.631  0.4530 0.936
## 36                  Creatinine; 50    0.173000 -0.2830 0.630  0.4570 0.936
## 37            Isoleucine, 2TMS; 18   -0.165000 -0.6210 0.292  0.4800 0.936
## 38 3,4-Dihydroxybutanoic acid; 27    0.164000 -0.2930 0.620  0.4830 0.936
## 39              Dodecanoic acid; 54    0.162000 -0.2950 0.618  0.4870 0.936
## 40 4-Hydroxyphenyllactic acid; 44    0.155000 -0.3020 0.611  0.5070 0.950
## 41          Glyceryl-glycoside; 59   -0.141000 -0.5970 0.316  0.5460 0.955
## 42            Linoleic acid, TMS; 4   -0.134000 -0.5910 0.322  0.5640 0.955
## 43                 Pyruvic acid; 31    0.134000 -0.3230 0.590  0.5660 0.955
## 44 3-Hydroxybutyric acid, 2TMS; 1    0.130000 -0.3260 0.587  0.5760 0.955
## 45     4-Hydroxybutanoic acid; 43   -0.128000 -0.5850 0.328  0.5810 0.955
## 46           Pyroglutamic acid; 69    0.127000 -0.3290 0.584  0.5860 0.955
```

```
## 47                Campesterol; 49   -0.091400 -0.5480 0.365  0.6950 0.999
## 48           Arabinopyranose; 51    0.088100 -0.3680 0.545  0.7050 0.999
## 49                Bisphenol A; 48   -0.087600 -0.5440 0.369  0.7070 0.999
## 50             1-Monopalmitin; 37   -0.087400 -0.5440 0.369  0.7080 0.999
## 51              Octanoic acid; 68    0.084000 -0.3730 0.540  0.7180 0.999
## 52         Glutamic acid, 3TMS; 8    0.071300 -0.3850 0.528  0.7600 0.999
## 53                    Ribitol; 70    0.070400 -0.3860 0.527  0.7620 0.999
## 54        2-Palmitoylglycerol; 39    0.070400 -0.3860 0.527  0.7630 0.999
## 55       4-Deoxytetronic acid; 32    0.067300 -0.3890 0.524  0.7730 0.999
## 56        Phenylalanine, 2TMS; 13    0.066600 -0.3900 0.523  0.7750 0.999
## 57              Alanine, 2TMS; 25    0.059300 -0.3970 0.516  0.7990 0.999
## 58       4-Deoxytetronic acid; 33   -0.047800 -0.5040 0.409  0.8380 0.999
## 59              Glyceric acid; 30   -0.044100 -0.5010 0.412  0.8500 0.999
## 60          Fumaric acid, 2TMS; 9   -0.040100 -0.4970 0.416  0.8630 0.999
## 61             Tartronic acid; 73    0.039600 -0.4170 0.496  0.8650 0.999
## 62           alpha-Tocopherol; 26   -0.034400 -0.4910 0.422  0.8830 0.999
## 63      Arachidonic acid, TMS; 24    0.034100 -0.4220 0.491  0.8830 0.999
## 64       Docosahexaenoic acid; 53    0.031800 -0.4250 0.488  0.8910 0.999
## 65             Leucine, 2TMS; 19    0.031700 -0.4250 0.488  0.8920 0.999
## 66                1-Dodecanol; 36    0.028400 -0.4280 0.485  0.9030 0.999
## 67 4-Hydroxybenzeneacetic acid; 4   -0.025400 -0.4820 0.431  0.9130 0.999
## 68     3-Indolepropionic acid; 41   -0.022100 -0.4790 0.434  0.9240 0.999
## 69           Threonine, 3TMS; 12   -0.017700 -0.4740 0.439  0.9390 0.999
## 70              Valine, 2TMS; 20   -0.014300 -0.4710 0.442  0.9510 0.999
## 71             Hydroxyproline; 64   -0.008790 -0.4650 0.448  0.9700 0.999
## 72              Hydroxylamine; 62    0.004630 -0.4520 0.461  0.9840 0.999
## 73                   Glycerol; 57   -0.002280 -0.4590 0.454  0.9920 0.999
## 74                Lactic acid; 29    0.001630 -0.4550 0.458  0.9940 0.999
## 75        3-Indoleacetic acid; 40   -0.000194 -0.4570 0.456  0.9990 0.999
```

##### 6.1.2 Adjusted Model

```
## [1] "Fitting models:"
## [1] "~ Ulcer.diagnosis.at.DATE + Age.x + Gender.x + Hba1c_baseline + CALSBP + bmi + Smoking + Statin
## [1] ""
```

###### 6.1.2.1 Forest Plot of Model Coefficients

```
## Warning: Ignoring unknown aesthetics: x
## Ignoring unknown aesthetics: x
## Ignoring unknown aesthetics: x
## Ignoring unknown aesthetics: x
## Ignoring unknown aesthetics: x
## Ignoring unknown aesthetics: x
## Ignoring unknown aesthetics: x
## Ignoring unknown aesthetics: x
## Ignoring unknown aesthetics: x
```

```
## NULL
```

###### 6.1.2.2 Tables of Model Coefficients

```
## [1] ""
## [1] "Table: Ulcer.diagnosis.at.DATEJA"
## [1] "  (from model: "
## [1] "   ~ Ulcer.diagnosis.at.DATE + Age.x + Gender.x +"
## [1] "     Hba1c_baseline + CALSBP + bmi + Smoking + Statin +"
## [1] "     log_Blood_TGA + Total_cholesterol)"
## [1] ""
## [1] "No significant associations at p.adj < 0.2"
## [1] ""
## [1] "Table: Age.x"
## [1] "  (from model: "
## [1] "   ~ Ulcer.diagnosis.at.DATE + Age.x + Gender.x +"
## [1] "     Hba1c_baseline + CALSBP + bmi + Smoking + Statin +"
## [1] "     log_Blood_TGA + Total_cholesterol)"
## [1] ""
##                               Name Coefficient      CI.L      CI.R  p.value
## 1       Eicosapentaenoic acid; 55     0.02210  1.54e-02  2.88e-02 9.93e-11
## 2  2,4-Dihydroxybutanoic acid; 28     0.01750  1.08e-02  2.42e-02 3.01e-07
## 3  4-Hydroxybenzeneacetic acid; 4     0.01720  1.05e-02  2.39e-02 4.61e-07
## 4             Myo inositol 6TMS; 1     0.01520  8.49e-03  2.19e-02 8.79e-06
## 5                      Ribitol; 71     0.01430  7.63e-03  2.10e-02 2.74e-05
## 6        Docosahexaenoic acid; 53     0.01360  6.93e-03  2.03e-02 6.63e-05
## 7          3-Indoleacetic acid; 40     0.01360  6.89e-03  2.03e-02 6.99e-05
## 8                 Ribonic acid; 72     0.01200  5.35e-03  1.87e-02 4.24e-04
## 9            Aminomalonic acid; 45     0.01150  4.77e-03  1.82e-02 7.91e-04
## 10                     Ribitol; 70     0.01100  4.28e-03  1.77e-02 1.32e-03
## 11            Citric acid, 4TMS; 6     0.01080  4.14e-03  1.75e-02 1.51e-03
## 12 3,4-Dihydroxybutanoic acid; 27     0.01070  4.04e-03  1.74e-02 1.68e-03
## 13 alpha-ketoglutaric acid, TMS M     0.01070  4.01e-03  1.74e-02 1.72e-03
## 14            Malic acid, 3TMS; 11     0.01050  3.78e-03  1.72e-02 2.17e-03
## 15          Fumaric acid, 2TMS; 9     0.01050  3.78e-03  1.72e-02 2.18e-03
## 16 4-Hydroxyphenyllactic acid; 44     0.01020  3.53e-03  1.69e-02 2.76e-03
## 17                Pyruvic acid; 31     0.01020  3.52e-03  1.69e-02 2.79e-03
## 18            alpha-Tocopherol; 26     0.01020  3.48e-03  1.69e-02 2.89e-03
## 19               Decanoic acid; 52     0.01010  3.42e-03  1.68e-02 3.08e-03
## 20               Glyceric acid; 30     0.00987  3.17e-03  1.66e-02 3.87e-03
## 21          Succinic acid, 2TMS; 7     0.00951  2.81e-03  1.62e-02 5.39e-03
## 22               Glycine, 3TMS; 17     0.00948  2.79e-03  1.62e-02 5.51e-03
## 23           Pyroglutamic acid; 69     0.00896  2.27e-03  1.57e-02 8.69e-03
## 24               Alanine, 2TMS; 25     0.00865  1.95e-03  1.53e-02 1.13e-02
## 25          11-Eicosenoic acid; 35     0.00855  1.85e-03  1.52e-02 1.23e-02
## 26                  Creatinine; 50     0.00808  1.38e-03  1.48e-02 1.80e-02
## 27               Leucine, 2TMS; 19    -0.00779 -1.45e-02 -1.09e-03 2.26e-02
## 28                Valine, 2TMS; 20    -0.00777 -1.45e-02 -1.07e-03 2.29e-02
## 29             Dodecanoic acid; 54     0.00721  5.19e-04  1.39e-02 3.47e-02
## 30            Myristoleic acid; 65     0.00712  4.25e-04  1.38e-02 3.71e-02
## 31          Heptadecanoic acid; 61     0.00701  3.17e-04  1.37e-02 4.01e-02
## 32         Phenylalanine, 2TMS; 13     0.00683  1.32e-04  1.35e-02 4.57e-02
## 33           Benzeneacetic acid; 47     0.00681  1.15e-04  1.35e-02 4.62e-02
## 34            Isoleucine, 2TMS; 18    -0.00678 -1.35e-02 -7.99e-05 4.73e-02
## 35              Tartronic acid; 73     0.00665 -4.17e-05  1.33e-02 5.15e-02
## 36          Glutamic acid, 3TMS; 8     0.00644 -2.57e-04  1.31e-02 5.95e-02
```

```
## 37             Oleic acid, TMS; 3     0.00640 -2.98e-04  1.31e-02 6.11e-02
## 38              Hydroxylamine; 62    -0.00618 -1.29e-02  5.14e-04 7.04e-02
## 39                1-Dodecanol; 36    -0.00616 -1.29e-02  5.34e-04 7.13e-02
## 40             L-5-Oxoproline; 63     0.00604 -6.60e-04  1.27e-02 7.73e-02
## 41         Heptadecanoic acid; 60     0.00600 -6.99e-04  1.27e-02 7.92e-02
## 42            Threonine, 3TMS; 12    -0.00598 -1.27e-02  7.12e-04 7.99e-02
## 43           Nonadecanoic acid; 66     0.00584 -8.53e-04  1.25e-02 8.72e-02
## 44           Palmitic acid, TMS; 5     0.00569 -1.00e-03  1.24e-02 9.55e-02
##       p.adj
## 1  7.44e-09
## 2  1.13e-05
## 3  1.15e-05
## 4  1.65e-04
## 5  4.11e-04
## 6  7.49e-04
## 7  7.49e-04
## 8  3.97e-03
## 9  6.60e-03
## 10 9.89e-03
## 11 9.93e-03
## 12 9.93e-03
## 13 9.93e-03
## 14 1.09e-02
## 15 1.09e-02
## 16 1.21e-02
## 17 1.21e-02
## 18 1.21e-02
## 19 1.22e-02
## 20 1.45e-02
## 21 1.88e-02
## 22 1.88e-02
## 23 2.83e-02
## 24 3.54e-02
## 25 3.70e-02
## 26 5.20e-02
## 27 6.15e-02
## 28 6.15e-02
## 29 8.97e-02
## 30 9.28e-02
## 31 9.70e-02
## 32 1.04e-01
## 33 1.04e-01
## 34 1.04e-01
## 35 1.10e-01
## 36 1.24e-01
## 37 1.24e-01
## 38 1.37e-01
## 39 1.37e-01
## 40 1.43e-01
## 41 1.43e-01
## 42 1.43e-01
## 43 1.52e-01
## 44 1.63e-01
## [1] ""
```

```
## [1] "Table: Gender.x"
## [1] "  (from model: "
## [1] "   ~ Ulcer.diagnosis.at.DATE + Age.x + Gender.x +"
## [1] "     Hba1c_baseline + CALSBP + bmi + Smoking + Statin +"
## [1] "     log_Blood_TGA + Total_cholesterol)"
## [1] ""
##                              Name Coefficient    CI.L     CI.R  p.value    p.adj
## 1           Citric acid, 4TMS; 6      -0.395 -0.5520 -0.23700 8.90e-07 6.68e-05
## 2           Methionine, 2TMS; 16       0.381  0.2230  0.53800 2.16e-06 7.24e-05
## 3               Valine, 2TMS; 20       0.376  0.2180  0.53300 2.90e-06 7.24e-05
## 4              Proline, 2TMS; 21       0.366  0.2080  0.52300 5.26e-06 9.26e-05
## 5           Myristoleic acid; 65      -0.363 -0.5200 -0.20600 6.17e-06 9.26e-05
## 6              Leucine, 2TMS; 19       0.353  0.1960  0.51100 1.10e-05 1.37e-04
## 7              Glycine, 3TMS; 17      -0.348 -0.5050 -0.19000 1.50e-05 1.60e-04
## 8           Isoleucine, 2TMS; 18       0.345  0.1880  0.50300 1.71e-05 1.60e-04
## 9             Tartronic acid; 73      -0.343 -0.5010 -0.18600 1.93e-05 1.61e-04
## 10             Glyceric acid; 30      -0.332 -0.4900 -0.17500 3.52e-05 2.45e-04
## 11           Dodecanoic acid; 54      -0.332 -0.4890 -0.17400 3.59e-05 2.45e-04
## 12      4-Deoxytetronic acid; 33       0.313  0.1550  0.47000 9.97e-05 6.23e-04
## 13            Oleic acid, TMS; 3      -0.303 -0.4600 -0.14500 1.65e-04 9.54e-04
## 14         Aminomalonic acid; 45      -0.295 -0.4520 -0.13700 2.41e-04 1.29e-03
## 15 2-hydroxy Isovaleric acid; 38       0.288  0.1310  0.44600 3.34e-04 1.67e-03
## 16          Cholesterol, TMS; 23      -0.276 -0.4340 -0.11900 5.77e-04 2.71e-03
## 17          Tridecanoic acid; 74      -0.260 -0.4170 -0.10200 1.22e-03 5.06e-03
## 18         Glutamic acid, 3TMS; 8      0.259  0.1020  0.41600 1.27e-03 5.06e-03
## 19           Myo inositol 6TMS; 1      -0.257 -0.4150 -0.10000 1.35e-03 5.06e-03
## 20        Heptadecanoic acid; 60      -0.257 -0.4140 -0.09930 1.39e-03 5.06e-03
## 21      Docosahexaenoic acid; 53      -0.255 -0.4130 -0.09810 1.47e-03 5.06e-03
## 22             Decanoic acid; 52      -0.255 -0.4130 -0.09780 1.48e-03 5.06e-03
## 23        Heptadecanoic acid; 61      -0.248 -0.4060 -0.09100 1.98e-03 6.47e-03
## 24        Succinic acid, 2TMS; 7      -0.246 -0.4040 -0.08900 2.15e-03 6.72e-03
## 25         Nonadecanoic acid; 66      -0.225 -0.3820 -0.06770 5.08e-03 1.52e-02
## 26         Palmitic acid, TMS; 5      -0.213 -0.3700 -0.05530 8.10e-03 2.34e-02
## 27          Stearic acid, TMS; 2      -0.207 -0.3650 -0.04990 9.84e-03 2.73e-02
## 28        Benzeneacetic acid; 47      -0.195 -0.3520 -0.03740 1.53e-02 4.09e-02
## 29            Hydroxyproline; 64       0.182  0.0246  0.33900 2.34e-02 6.06e-02
## 30            1-Monopalmitin; 37       0.176  0.0189  0.33400 2.82e-02 7.04e-02
## 31        11-Eicosenoic acid; 35      -0.172 -0.3300 -0.01480 3.20e-02 7.75e-02
## 32              Ribonic acid; 72      -0.166 -0.3230 -0.00868 3.86e-02 8.89e-02
## 33                  Tyrosine; 75      -0.166 -0.3230 -0.00829 3.91e-02 8.89e-02
## 34             Nonanoic acid; 67      -0.162 -0.3200 -0.00499 4.32e-02 9.52e-02
## 35              Pyruvic acid; 31       0.150 -0.0069  0.30800 6.09e-02 1.31e-01
## 36     Arachidonic acid, TMS; 24      -0.143 -0.3000  0.01460 7.54e-02 1.57e-01
## 37                  Glycerol; 57      -0.139 -0.2960  0.01870 8.43e-02 1.71e-01
## [1] ""
## [1] "Table: Hba1c_baseline"
## [1] "  (from model: "
## [1] "   ~ Ulcer.diagnosis.at.DATE + Age.x + Gender.x +"
## [1] "     Hba1c_baseline + CALSBP + bmi + Smoking + Statin +"
## [1] "     log_Blood_TGA + Total_cholesterol)"
## [1] ""
##                         Name Coefficient     CI.L     CI.R  p.value  p.adj
## 1        Tridecanoic acid; 74     -0.1280 -0.19900 -0.05650 0.000440 0.0166
## 2         Arabinopyranose; 51      0.1260  0.05500  0.19700 0.000510 0.0166
```

```
## 3   Eicosapentaenoic acid; 55     -0.1240 -0.19500 -0.05240 0.000665 0.0166
## 4            Ethanolamine; 56      0.1160  0.04440  0.18700 0.001460 0.0232
## 5           Glyceric acid; 30     -0.1130 -0.18500 -0.04230 0.001770 0.0232
## 6    Docosahexaenoic acid; 53     -0.1130 -0.18400 -0.04190 0.001860 0.0232
## 7            Valine, 2TMS; 20      0.1090  0.03780  0.18000 0.002690 0.0288
## 8           Alanine, 2TMS; 25      0.1070  0.03610  0.17800 0.003130 0.0294
## 9           Decanoic acid; 52     -0.0945 -0.16600 -0.02330 0.009250 0.0770
## 10 4-Hydroxybutanoic acid; 43     -0.0898 -0.16100 -0.01860 0.013400 0.1010
## 11     11-Eicosenoic acid; 35     -0.0827 -0.15400 -0.01150 0.022800 0.1550
## 12            Lactic acid; 29      0.0801  0.00889  0.15100 0.027500 0.1720
## 13       Myristoleic acid; 65     -0.0766 -0.14800 -0.00542 0.034900 0.1910
## 14       Citric acid, 4TMS; 6      0.0754  0.00425  0.14700 0.037800 0.1910
## 15         Tartronic acid; 73     -0.0752 -0.14600 -0.00407 0.038300 0.1910
## [1] ""
## [1] "Table: CALSBP"
## [1] "  (from model: "
## [1] "   ~ Ulcer.diagnosis.at.DATE + Age.x + Gender.x +"
## [1] "     Hba1c_baseline + CALSBP + bmi + Smoking + Statin +"
## [1] "     log_Blood_TGA + Total_cholesterol)"
## [1] ""
##                   Name Coefficient     CI.L     CI.R  p.value  p.adj
## 1 Myristoleic acid; 65     0.00809  0.00341  0.01280 0.000695 0.0521
## 2    Glyceric acid; 30    -0.00666 -0.01130 -0.00199 0.005210 0.1960
## [1] ""
## [1] "Table: bmi"
## [1] "  (from model: "
## [1] "   ~ Ulcer.diagnosis.at.DATE + Age.x + Gender.x +"
## [1] "     Hba1c_baseline + CALSBP + bmi + Smoking + Statin +"
## [1] "     log_Blood_TGA + Total_cholesterol)"
## [1] ""
##                               Name Coefficient     CI.L     CI.R  p.value
## 1          Glutamic acid, 3TMS; 8      0.0450  0.02440  0.06560 1.89e-05
## 2                 Campesterol; 49     -0.0396 -0.06020 -0.01900 1.66e-04
## 3  2-Hydroxybutyric acid, 2TMS; 2      0.0385  0.01790  0.05910 2.54e-04
## 4                Pyruvic acid; 31     -0.0304 -0.05100 -0.00979 3.85e-03
## 5               Decanoic acid; 52     -0.0297 -0.05030 -0.00911 4.71e-03
## 6                 Lactic acid; 29      0.0295  0.00885  0.05010 5.09e-03
## 7              Arachidic acid; 46     -0.0290 -0.04960 -0.00841 5.79e-03
## 8             1,3-Propanediol; 34     -0.0285 -0.04910 -0.00785 6.80e-03
## 9            alpha-Tocopherol; 26     -0.0274 -0.04800 -0.00679 9.18e-03
## 10 2,4-Dihydroxybutanoic acid; 28     -0.0247 -0.04530 -0.00411 1.88e-02
## 11          Pyroglutamic acid; 69     -0.0243 -0.04500 -0.00374 2.06e-02
## 12           Citric acid, 4TMS; 6     -0.0236 -0.04420 -0.00297 2.49e-02
## 13                    Ribitol; 70      0.0231  0.00253  0.04380 2.78e-02
## 14 alpha-ketoglutaric acid, TMS M     -0.0220 -0.04260 -0.00138 3.65e-02
##      p.adj
## 1  0.00142
## 2  0.00622
## 3  0.00635
## 4  0.06200
## 5  0.06200
## 6  0.06200
## 7  0.06200
## 8  0.06380
```

```
## 9  0.07650
## 10 0.14100
## 11 0.14100
## 12 0.15600
## 13 0.16000
## 14 0.19500
## [1] ""
## [1] "Table: Smoking"
## [1] "  (from model: "
## [1] "   ~ Ulcer.diagnosis.at.DATE + Age.x + Gender.x +"
## [1] "     Hba1c_baseline + CALSBP + bmi + Smoking + Statin +"
## [1] "     log_Blood_TGA + Total_cholesterol)"
## [1] ""
##                               Name Coefficient    CI.L     CI.R  p.value   p.adj
## 1          Glutamic acid, 3TMS; 8       0.385  0.1920  0.57800 9.29e-05 0.00618
## 2      3-Indolepropionic acid; 41      -0.362 -0.5550 -0.17000 2.31e-04 0.00618
## 3        Docosahexaenoic acid; 53      -0.361 -0.5540 -0.16800 2.47e-04 0.00618
## 4              Tartronic acid; 73      -0.353 -0.5450 -0.16000 3.41e-04 0.00639
## 5               Glyceric acid; 30      -0.326 -0.5190 -0.13300 9.30e-04 0.01390
## 6            Citric acid, 4TMS; 6      -0.309 -0.5020 -0.11600 1.71e-03 0.02140
## 7            alpha-Tocopherol; 26      -0.293 -0.4860 -0.09990 2.93e-03 0.03140
## 8          Benzeneacetic acid; 47      -0.282 -0.4750 -0.08960 4.11e-03 0.03850
## 9                Ribonic acid; 72      -0.274 -0.4670 -0.08150 5.30e-03 0.04400
## 10           Malic acid, 3TMS; 11      -0.268 -0.4610 -0.07520 6.44e-03 0.04400
## 11               Valine, 2TMS; 20      -0.266 -0.4590 -0.07310 6.89e-03 0.04400
## 12                Campesterol; 49      -0.265 -0.4580 -0.07240 7.04e-03 0.04400
## 13       3-Indoleacetic acid; 40      -0.236 -0.4290 -0.04350 1.63e-02 0.09420
## 14 4-Hydroxyphenyllactic acid; 44      -0.228 -0.4210 -0.03480 2.07e-02 0.11100
## 15              Leucine, 2TMS; 19      -0.208 -0.4010 -0.01540 3.43e-02 0.15700
## 16                    Ribitol; 70       0.207  0.0144  0.40000 3.51e-02 0.15700
## 17         Heptadecanoic acid; 60      -0.207 -0.4000 -0.01400 3.55e-02 0.15700
## 18         Heptadecanoic acid; 61      -0.200 -0.3930 -0.00688 4.24e-02 0.17700
## [1] ""
## [1] "Table: Statin"
## [1] "  (from model: "
## [1] "   ~ Ulcer.diagnosis.at.DATE + Age.x + Gender.x +"
## [1] "     Hba1c_baseline + CALSBP + bmi + Smoking + Statin +"
## [1] "     log_Blood_TGA + Total_cholesterol)"
## [1] ""
##                               Name Coefficient    CI.L    CI.R  p.value  p.adj
## 1              L-5-Oxoproline; 63      -0.323 -0.4970 -0.1490 0.000279 0.0209
## 2                 Campesterol; 49       0.255  0.0812  0.4290 0.004060 0.1520
## 3       Arachidonic acid, TMS; 24       0.213  0.0388  0.3870 0.016500 0.1900
## 4              Arachidic acid; 46      -0.212 -0.3860 -0.0379 0.017000 0.1900
## 5          Glutamic acid, 3TMS; 8       0.211  0.0372  0.3850 0.017400 0.1900
## 6          Heptadecanoic acid; 61      -0.210 -0.3840 -0.0359 0.018100 0.1900
## 7            Cholesterol, TMS; 23      -0.208 -0.3820 -0.0334 0.019500 0.1900
## 8                     Ribitol; 70       0.203  0.0292  0.3770 0.022100 0.1900
## 9           Aminomalonic acid; 45      -0.200 -0.3740 -0.0256 0.024500 0.1900
## 10            Dodecanoic acid; 54      -0.197 -0.3710 -0.0231 0.026400 0.1900
## 11          Linoleic acid, TMS; 4      -0.192 -0.3660 -0.0182 0.030400 0.1900
## 12                    Ribitol; 71       0.191  0.0173  0.3660 0.031200 0.1900
## 13               Valine, 2TMS; 20      -0.189 -0.3640 -0.0153 0.033000 0.1900
## 14 2,4-Dihydroxybutanoic acid; 28       0.187  0.0128  0.3610 0.035400 0.1900
```

```
## [1] ""
## [1] "Table: log_Blood_TGA"
## [1] "  (from model: "
## [1] "   ~ Ulcer.diagnosis.at.DATE + Age.x + Gender.x +"
## [1] "     Hba1c_baseline + CALSBP + bmi + Smoking + Statin +"
## [1] "     log_Blood_TGA + Total_cholesterol)"
## [1] ""
##                               Name Coefficient     CI.L     CI.R  p.value
## 1            Dodecanoic acid; 54       0.275  0.14600  0.40400 2.86e-05
## 2          Palmitic acid, TMS; 5       0.274  0.14500  0.40300 3.23e-05
## 3              Decanoic acid; 52       0.254  0.12500  0.38300 1.17e-04
## 4           Stearic acid, TMS; 2       0.253  0.12400  0.38200 1.23e-04
## 5             Arachidic acid; 46       0.249  0.12000  0.37800 1.55e-04
## 6              Octanoic acid; 68       0.236  0.10700  0.36500 3.34e-04
## 7  3,4-Dihydroxybutanoic acid; 27       0.229  0.10000  0.35800 5.01e-04
## 8                Ribonic acid; 72       0.220  0.09120  0.34900 8.21e-04
## 9          Glyceryl-glycoside; 59       0.220  0.09090  0.34900 8.36e-04
## 10 4-Hydroxybenzeneacetic acid; 4       0.213  0.08380  0.34200 1.22e-03
## 11             Oleic acid, TMS; 3       0.209  0.07990  0.33800 1.50e-03
## 12                    Ribitol; 71       0.197  0.06800  0.32600 2.76e-03
## 13            Arabinopyranose; 51       0.192  0.06250  0.32100 3.62e-03
## 14         Heptadecanoic acid; 60       0.184  0.05500  0.31300 5.19e-03
## 15 2-Hydroxybutyric acid, 2TMS; 2       0.181  0.05150  0.31000 6.09e-03
## 16                Lactic acid; 29       0.171  0.04240  0.30000 9.21e-03
## 17 2,4-Dihydroxybutanoic acid; 28       0.171  0.04170  0.30000 9.52e-03
## 18         Heptadecanoic acid; 61       0.169  0.04000  0.29800 1.02e-02
## 19          Fumaric acid, 2TMS; 9       0.163  0.03430  0.29200 1.31e-02
## 20         Glutamic acid, 3TMS; 8       0.160  0.03140  0.28900 1.48e-02
## 21               Pyruvic acid; 31       0.160  0.03070  0.28900 1.53e-02
## 22           Aminomalonic acid; 45      -0.157 -0.28600 -0.02810 1.70e-02
## 23          Myristoleic acid; 65       0.157  0.02800  0.28600 1.71e-02
## 24          Isoleucine, 2TMS; 18       0.156  0.02750  0.28500 1.74e-02
## 25               Valine, 2TMS; 20       0.156  0.02720  0.28500 1.76e-02
## 26     4-Hydroxybutanoic acid; 43       0.156  0.02700  0.28500 1.78e-02
## 27           Myo inositol 6TMS; 1       0.156  0.02660  0.28500 1.81e-02
## 28       4-Deoxytetronic acid; 33       0.153  0.02410  0.28200 2.00e-02
## 29          Malic acid, 3TMS; 11       0.152  0.02330  0.28100 2.07e-02
## 30 alpha-ketoglutaric acid, TMS M       0.152  0.02280  0.28100 2.11e-02
## 31     3-Indolepropionic acid; 41      -0.142 -0.27100 -0.01280 3.13e-02
## 32         Succinic acid, 2TMS; 7       0.138  0.00943  0.26700 3.54e-02
## 33              Glyceric acid; 30      -0.138 -0.26700 -0.00915 3.58e-02
## 34              Leucine, 2TMS; 19       0.137  0.00848  0.26600 3.67e-02
## 35              Proline, 2TMS; 21       0.135  0.00604  0.26400 4.02e-02
## 36      Arachidonic acid, TMS; 24       0.135  0.00559  0.26400 4.09e-02
## 37          Nonadecanoic acid; 66       0.124 -0.00490  0.25300 5.94e-02
## 38             Tartronic acid; 73      -0.123 -0.25200  0.00570 6.10e-02
## 39             1,3-Propanediol; 34       0.117 -0.01230  0.24600 7.63e-02
## 40               Serine, 3TMS; 14      -0.116 -0.24500  0.01290 7.77e-02
## 41 3-Hydroxybutyric acid, 2TMS; 1       0.115 -0.01440  0.24400 8.16e-02
## 42          Tridecanoic acid; 74       0.114 -0.01480  0.24300 8.28e-02
## 43        3-Indoleacetic acid; 40       0.112 -0.01690  0.24100 8.85e-02
## 44              Nonanoic acid; 67       0.107 -0.02220  0.23600 1.05e-01
##      p.adj
## 1  0.00121
```

```
## 2  0.00121
## 3  0.00230
## 4  0.00230
## 5  0.00232
## 6  0.00417
## 7  0.00537
## 8  0.00696
## 9  0.00696
## 10 0.00918
## 11 0.01020
## 12 0.01720
## 13 0.02090
## 14 0.02780
## 15 0.03040
## 16 0.04200
## 17 0.04200
## 18 0.04270
## 19 0.05030
## 20 0.05030
## 21 0.05030
## 22 0.05030
## 23 0.05030
## 24 0.05030
## 25 0.05030
## 26 0.05030
## 27 0.05030
## 28 0.05260
## 29 0.05260
## 30 0.05260
## 31 0.07560
## 32 0.08100
## 33 0.08100
## 34 0.08100
## 35 0.08520
## 36 0.08520
## 37 0.12000
## 38 0.12000
## 39 0.14600
## 40 0.14600
## 41 0.14800
## 42 0.14800
## 43 0.15400
## 44 0.17900
## [1] ""
## [1] "Table: Total_cholesterol"
## [1] "  (from model: "
## [1] "   ~ Ulcer.diagnosis.at.DATE + Age.x + Gender.x +"
## [1] "     Hba1c_baseline + CALSBP + bmi + Smoking + Statin +"
## [1] "     log_Blood_TGA + Total_cholesterol)"
## [1] ""
##                               Name Coefficient      CI.L     CI.R  p.value
## 1            Cholesterol, TMS; 23      0.4640  0.367000  0.56000 4.49e-21
## 2                 Campesterol; 49      0.3700  0.274000  0.46700 5.31e-14
## 3            alpha-Tocopherol; 26      0.3290  0.232000  0.42500 2.38e-11
```

```
## 4          Benzeneacetic acid; 47     -0.1900 -0.286000 -0.09340 1.14e-04
## 5      4-Hydroxybutanoic acid; 43     -0.1820 -0.278000 -0.08560 2.15e-04
## 6           Linoleic acid, TMS; 4      0.1820  0.085200  0.27800 2.22e-04
## 7               Proline, 2TMS; 21     -0.1740 -0.271000 -0.07800 3.93e-04
## 8  2,4-Dihydroxybutanoic acid; 28     -0.1700 -0.267000 -0.07400 5.30e-04
## 9               Glycine, 3TMS; 17     -0.1690 -0.265000 -0.07250 5.93e-04
## 10             L-5-Oxoproline; 63     -0.1610 -0.257000 -0.06430 1.09e-03
## 11 4-Hydroxybenzeneacetic acid; 4     -0.1550 -0.252000 -0.05880 1.61e-03
## 12                   Ribitol; 71     -0.1490 -0.246000 -0.05280 2.42e-03
## 13       Docosahexaenoic acid; 53      0.1490  0.052200  0.24500 2.52e-03
## 14      Eicosapentaenoic acid; 55      0.1480  0.051600  0.24400 2.61e-03
## 15                  Tyrosine; 75     -0.1460 -0.243000 -0.05000 2.92e-03
## 16           Isoleucine, 2TMS; 18     -0.1460 -0.243000 -0.04970 2.97e-03
## 17         Glyceryl-glycoside; 59     -0.1430 -0.240000 -0.04690 3.57e-03
## 18            Arabinopyranose; 51     -0.1420 -0.239000 -0.04580 3.83e-03
## 19              Alanine, 2TMS; 25     -0.1420 -0.238000 -0.04530 3.96e-03
## 20        2-Palmitoylglycerol; 39      0.1380  0.041700  0.23400 4.99e-03
## 21           Methionine, 2TMS; 16     -0.1380 -0.234000 -0.04130 5.11e-03
## 22            Threonine, 3TMS; 12     -0.1340 -0.230000 -0.03760 6.44e-03
## 23               Ribonic acid; 72     -0.1330 -0.230000 -0.03700 6.70e-03
## 24        3-Indoleacetic acid; 40     -0.1290 -0.226000 -0.03280 8.65e-03
## 25              Hydroxylamine; 62     -0.1250 -0.221000 -0.02870 1.10e-02
## 26 3,4-Dihydroxybutanoic acid; 27     -0.1170 -0.214000 -0.02090 1.71e-02
## 27           Malic acid, 3TMS; 11     -0.1090 -0.206000 -0.01300 2.62e-02
## 28        Phenylalanine, 2TMS; 13     -0.1090 -0.205000 -0.01210 2.74e-02
## 29               Serine, 3TMS; 14     -0.1080 -0.204000 -0.01140 2.83e-02
## 30               Pyruvic acid; 31     -0.1010 -0.197000 -0.00441 4.04e-02
## 31           Myo inositol 6TMS; 1     -0.0997 -0.196000 -0.00326 4.28e-02
## 32 2-Hydroxybutyric acid, 2TMS; 2      0.0962 -0.000213  0.19300 5.05e-02
## 33          Palmitic acid, TMS; 5      0.0933 -0.003080  0.19000 5.78e-02
## 34     3-Indolepropionic acid; 41      0.0861 -0.010300  0.18200 8.01e-02
## 35            1,3-Propanediol; 34     -0.0858 -0.182000  0.01060 8.10e-02
## 36               Ethanolamine; 56     -0.0827 -0.179000  0.01370 9.27e-02
##       p.adj
## 1  3.37e-19
## 2  1.99e-12
## 3  5.95e-10
## 4  2.14e-03
## 5  2.78e-03
## 6  2.78e-03
## 7  4.21e-03
## 8  4.94e-03
## 9  4.94e-03
## 10 8.15e-03
## 11 1.10e-02
## 12 1.39e-02
## 13 1.39e-02
## 14 1.39e-02
## 15 1.39e-02
## 16 1.39e-02
## 17 1.56e-02
## 18 1.56e-02
## 19 1.56e-02
## 20 1.82e-02
```

```
## 21 1.82e-02
## 22 2.19e-02
## 23 2.19e-02
## 24 2.70e-02
## 25 3.30e-02
## 26 4.94e-02
## 27 7.28e-02
## 28 7.33e-02
## 29 7.33e-02
## 30 1.01e-01
## 31 1.03e-01
## 32 1.18e-01
## 33 1.31e-01
## 34 1.74e-01
## 35 1.74e-01
## 36 1.93e-01
```

###### 6.1.2.3 Table with All Metabolites

```
## [1] ""
## [1] "Table: Ulcer.diagnosis.at.DATEJA"
## [1] "  (from model: "
## [1] "   ~ Ulcer.diagnosis.at.DATE + Age.x + Gender.x +"
## [1] "     Hba1c_baseline + CALSBP + bmi + Smoking + Statin +"
## [1] "     log_Blood_TGA + Total_cholesterol)"
## [1] ""
##                                Name Coefficient   CI.L    CI.R p.value p.adj
## 1            Citric acid, 4TMS; 6   -0.567000 -1.050 -0.0878  0.0204 0.986
## 2  2-Hydroxybutyric acid, 2TMS; 2   -0.425000 -0.904  0.0546  0.0825 0.986
## 3   2-hydroxy Isovaleric acid; 38   -0.357000 -0.837  0.1220  0.1440 0.986
## 4            Cholesterol, TMS; 23   -0.343000 -0.823  0.1360  0.1610 0.986
## 5          Benzeneacetic acid; 47   -0.339000 -0.819  0.1400  0.1650 0.986
## 6                     Ribitol; 71    0.338000 -0.141  0.8180  0.1670 0.986
## 7                    Tyrosine; 75    0.335000 -0.144  0.8140  0.1710 0.986
## 8                Serine, 3TMS; 14   -0.326000 -0.805  0.1540  0.1830 0.986
## 9                    Glycerol; 58    0.314000 -0.165  0.7940  0.1990 0.986
## 10              Nonanoic acid; 67    0.303000 -0.176  0.7820  0.2160 0.986
## 11              L-5-Oxoproline; 63  -0.296000 -0.776  0.1830  0.2260 0.986
## 12          Palmitic acid, TMS; 5    0.286000 -0.194  0.7650  0.2430 0.986
## 13          Nonadecanoic acid; 66    0.282000 -0.197  0.7620  0.2490 0.986
## 14         Glyceryl-glycoside; 59   -0.267000 -0.747  0.2120  0.2750 0.986
## 15       4-Deoxytetronic acid; 33   -0.267000 -0.746  0.2120  0.2750 0.986
## 16           Myristoleic acid; 65    0.258000 -0.221  0.7380  0.2910 0.986
## 17         Heptadecanoic acid; 61    0.252000 -0.227  0.7320  0.3020 0.986
## 18           alpha-Tocopherol; 26   -0.249000 -0.728  0.2310  0.3090 0.986
## 19           Malic acid, 3TMS; 11    0.234000 -0.245  0.7140  0.3380 0.986
## 20               Ribonic acid; 72    0.234000 -0.246  0.7130  0.3400 0.986
## 21           Methionine, 2TMS; 16    0.230000 -0.249  0.7100  0.3460 0.986
## 22 4-Hydroxybenzeneacetic acid; 4   -0.226000 -0.705  0.2540  0.3560 0.986
## 23           Stearic acid, TMS; 2    0.224000 -0.256  0.7030  0.3600 0.986
## 24          Aminomalonic acid; 45   -0.205000 -0.684  0.2740  0.4020 0.986
## 25            1,3-Propanediol; 34    0.200000 -0.280  0.6790  0.4140 0.986
## 26                    Ribitol; 70   -0.197000 -0.677  0.2820  0.4200 0.986
## 27              Glyceric acid; 30   -0.191000 -0.670  0.2890  0.4360 0.986
## 28           Tridecanoic acid; 74    0.187000 -0.292  0.6670  0.4440 0.986
## 29         Glutamic acid, 3TMS; 8   -0.178000 -0.657  0.3020  0.4680 0.986
## 30          Fumaric acid, 2TMS; 9   -0.177000 -0.656  0.3030  0.4700 0.986
## 31             1-Monopalmitin; 37   -0.176000 -0.655  0.3040  0.4730 0.986
## 32       Docosahexaenoic acid; 53   -0.172000 -0.652  0.3070  0.4820 0.986
## 33           Isoleucine, 2TMS; 18   -0.170000 -0.650  0.3090  0.4860 0.986
## 34                Campesterol; 49   -0.170000 -0.650  0.3090  0.4870 0.986
## 35 2,4-Dihydroxybutanoic acid; 28   -0.154000 -0.634  0.3250  0.5280 0.986
## 36 alpha-ketoglutaric acid, TMS M    0.152000 -0.327  0.6310  0.5340 0.986
## 37               Ethanolamine; 56    0.152000 -0.328  0.6310  0.5350 0.986
## 38              Proline, 2TMS; 21    0.151000 -0.329  0.6300  0.5380 0.986
## 39         Heptadecanoic acid; 60    0.144000 -0.335  0.6240  0.5550 0.986
## 40         3-Indoleacetic acid; 40  -0.144000 -0.623  0.3350  0.5560 0.986
## 41         11-Eicosenoic acid; 35    0.142000 -0.337  0.6210  0.5620 0.986
## 42             Arachidic acid; 46    0.142000 -0.337  0.6210  0.5620 0.986
## 43         Succinic acid, 2TMS; 7    0.139000 -0.340  0.6190  0.5690 0.986
## 44               Hydroxylamine; 62   0.130000 -0.350  0.6090  0.5960 0.986
```

```
## 45                Bisphenol A; 48   -0.123000 -0.603  0.3560  0.6140 0.986
## 46              Decanoic acid; 52    0.120000 -0.359  0.6000  0.6230 0.986
## 47           Myo inositol 6TMS; 1    0.117000 -0.362  0.5960  0.6320 0.986
## 48             Tartronic acid; 73   -0.107000 -0.587  0.3720  0.6610 0.986
## 49      4-Hydroxybutanoic acid; 43   -0.106000 -0.585  0.3740  0.6650 0.986
## 50              Glycine, 3TMS; 17   -0.102000 -0.582  0.3770  0.6750 0.986
## 51           Linoleic acid, TMS; 4   -0.102000 -0.582  0.3770  0.6760 0.986
## 52             Oleic acid, TMS; 3    0.098700 -0.381  0.5780  0.6870 0.986
## 53 3,4-Dihydroxybutanoic acid; 27   -0.090600 -0.570  0.3890  0.7110 0.986
## 54        2-Palmitoylglycerol; 39   -0.090300 -0.570  0.3890  0.7120 0.986
## 55                1-Dodecanol; 36    0.086100 -0.393  0.5660  0.7250 0.986
## 56 4-Hydroxyphenyllactic acid; 44   -0.082500 -0.562  0.3970  0.7360 0.986
## 57           Threonine, 3TMS; 12    0.071200 -0.408  0.5510  0.7710 0.997
## 58      Eicosapentaenoic acid; 55    0.060800 -0.419  0.5400  0.8040 0.997
## 59              Octanoic acid; 68    0.060000 -0.419  0.5390  0.8060 0.997
## 60               Valine, 2TMS; 20   -0.052900 -0.532  0.4270  0.8290 0.997
## 61                 Creatinine; 50    0.049400 -0.430  0.5290  0.8400 0.997
## 62              Hydroxyproline; 64   -0.047300 -0.527  0.4320  0.8470 0.997
## 63              Alanine, 2TMS; 25   -0.046500 -0.526  0.4330  0.8490 0.997
## 64      Arachidonic acid, TMS; 24   -0.040900 -0.520  0.4390  0.8670 0.997
## 65              Leucine, 2TMS; 19    0.037100 -0.442  0.5170  0.8790 0.997
## 66        Phenylalanine, 2TMS; 13   -0.026600 -0.506  0.4530  0.9130 0.997
## 67                Lactic acid; 29   -0.026500 -0.506  0.4530  0.9140 0.997
## 68               Pyruvic acid; 31    0.024500 -0.455  0.5040  0.9200 0.997
## 69 3-Hydroxybutyric acid, 2TMS; 1    0.020800 -0.459  0.5000  0.9320 0.997
## 70       4-Deoxytetronic acid; 32   -0.014700 -0.494  0.4650  0.9520 0.997
## 71            Arabinopyranose; 51   -0.010700 -0.490  0.4690  0.9650 0.997
## 72           Pyroglutamic acid; 69    0.009160 -0.470  0.4890  0.9700 0.997
## 73                   Glycerol; 57   -0.006500 -0.486  0.4730  0.9790 0.997
## 74             Dodecanoic acid; 54    0.005040 -0.474  0.4850  0.9840 0.997
## 75     3-Indolepropionic acid; 41   -0.000676 -0.480  0.4790  0.9980 0.998
```

##### 6.1.3 Adjusted Model with eGFR

```
## [1] "Fitting models:"
## [1] "~ Ulcer.diagnosis.at.DATE + Age.x + Gender.x + Hba1c_baseline + CALSBP + bmi + Smoking + Statin
## [1] ""
```

###### 6.1.3.1 Forest Plot of Model Coefficients

```
## Warning: Ignoring unknown aesthetics: x
## Ignoring unknown aesthetics: x
## Ignoring unknown aesthetics: x
## Ignoring unknown aesthetics: x
## Ignoring unknown aesthetics: x
## Ignoring unknown aesthetics: x
## Ignoring unknown aesthetics: x
## Ignoring unknown aesthetics: x
## Ignoring unknown aesthetics: x
## Ignoring unknown aesthetics: x
```

```
## NULL
```

###### 6.1.3.2 Tables of Model Coefficients

```
## [1] ""
## [1] "Table: Ulcer.diagnosis.at.DATEJA"
## [1] "  (from model: "
## [1] "   ~ Ulcer.diagnosis.at.DATE + Age.x + Gender.x +"
## [1] "     Hba1c_baseline + CALSBP + bmi + Smoking + Statin +"
## [1] "     log_Blood_TGA + Total_cholesterol + egfr)"
## [1] ""
## [1] "No significant associations at p.adj < 0.2"
## [1] ""
## [1] "Table: Age.x"
## [1] "  (from model: "
## [1] "   ~ Ulcer.diagnosis.at.DATE + Age.x + Gender.x +"
## [1] "     Hba1c_baseline + CALSBP + bmi + Smoking + Statin +"
## [1] "     log_Blood_TGA + Total_cholesterol + egfr)"
## [1] ""
##                                Name Coefficient      CI.L     CI.R  p.value
## 1       Eicosapentaenoic acid; 55     0.02460  1.78e-02 0.031400 2.11e-12
## 2        Docosahexaenoic acid; 53     0.01540  8.55e-03 0.022200 1.07e-05
## 3               Glyceric acid; 30     0.01220  5.34e-03 0.019000 4.85e-04
## 4            alpha-Tocopherol; 26     0.01170  4.91e-03 0.018600 7.66e-04
## 5                Pyruvic acid; 31     0.01150  4.57e-03 0.018400 1.15e-03
## 6               Decanoic acid; 52     0.01140  4.52e-03 0.018300 1.18e-03
## 7  alpha-ketoglutaric acid, TMS M     0.01120  4.26e-03 0.018100 1.58e-03
## 8  4-Hydroxybenzeneacetic acid; 4     0.01030  3.56e-03 0.017100 2.81e-03
## 9           Aminomalonic acid; 45     0.01030  3.46e-03 0.017200 3.24e-03
## 10        3-Indoleacetic acid; 40     0.00999  3.11e-03 0.016900 4.43e-03
## 11 2,4-Dihydroxybutanoic acid; 28     0.00902  2.33e-03 0.015700 8.28e-03
## 12                    Ribitol; 70     0.00928  2.36e-03 0.016200 8.59e-03
## 13                   Tyrosine; 75     0.00859  1.66e-03 0.015500 1.52e-02
## 14          Palmitic acid, TMS; 5     0.00828  1.40e-03 0.015200 1.84e-02
## 15         Succinic acid, 2TMS; 7     0.00820  1.25e-03 0.015100 2.07e-02
## 16           Malic acid, 3TMS; 11     0.00811  1.18e-03 0.015000 2.18e-02
## 17         11-Eicosenoic acid; 35     0.00812  1.17e-03 0.015100 2.20e-02
## 18           Myristoleic acid; 65     0.00793  1.05e-03 0.014800 2.38e-02
## 19          Fumaric acid, 2TMS; 9     0.00760  6.75e-04 0.014500 3.15e-02
## 20         Heptadecanoic acid; 61     0.00761  6.72e-04 0.014500 3.16e-02
## 21         Glutamic acid, 3TMS; 8     0.00747  6.41e-04 0.014300 3.20e-02
## 22            Dodecanoic acid; 54     0.00747  5.82e-04 0.014400 3.36e-02
## 23                   Glycerol; 57     0.00739  4.26e-04 0.014300 3.75e-02
## 24             Oleic acid, TMS; 3     0.00721  2.96e-04 0.014100 4.10e-02
## 25              Alanine, 2TMS; 25     0.00717  2.47e-04 0.014100 4.24e-02
## 26              L-5-Oxoproline; 63     0.00715  2.22e-04 0.014100 4.31e-02
## 27             Tartronic acid; 73     0.00706  1.96e-04 0.013900 4.38e-02
## 28 4-Hydroxyphenyllactic acid; 44     0.00688 -2.07e-05 0.013800 5.07e-02
## 29              Octanoic acid; 68     0.00687 -4.75e-05 0.013800 5.16e-02
## 30               Hydroxylamine; 62   -0.00679 -1.37e-02 0.000160 5.55e-02
## 31              Glycine, 3TMS; 17     0.00670 -1.68e-04 0.013600 5.59e-02
## 32        2-Palmitoylglycerol; 39     0.00657 -3.80e-04 0.013500 6.39e-02
## 33              Hydroxyproline; 64   -0.00630 -1.32e-02 0.000618 7.42e-02
## 34        Phenylalanine, 2TMS; 13     0.00633 -6.27e-04 0.013300 7.45e-02
## 35          Nonadecanoic acid; 66     0.00631 -6.37e-04 0.013300 7.50e-02
## 36            Citric acid, 4TMS; 6     0.00614 -6.88e-04 0.013000 7.79e-02
```

```
## 37          Stearic acid, TMS; 2     0.00615 -7.59e-04 0.013100 8.10e-02
## 38        Heptadecanoic acid; 60     0.00610 -8.32e-04 0.013000 8.45e-02
## 39          Methionine, 2TMS; 16     0.00598 -9.07e-04 0.012900 8.87e-02
## 40          Myo inositol 6TMS; 1     0.00572 -9.46e-04 0.012400 9.26e-02
##       p.adj
## 1  1.58e-10
## 2  4.03e-04
## 3  1.21e-02
## 4  1.44e-02
## 5  1.48e-02
## 6  1.48e-02
## 7  1.69e-02
## 8  2.63e-02
## 9  2.70e-02
## 10 3.33e-02
## 11 5.37e-02
## 12 5.37e-02
## 13 8.76e-02
## 14 9.69e-02
## 15 9.69e-02
## 16 9.69e-02
## 17 9.69e-02
## 18 9.93e-02
## 19 1.14e-01
## 20 1.14e-01
## 21 1.14e-01
## 22 1.14e-01
## 23 1.22e-01
## 24 1.22e-01
## 25 1.22e-01
## 26 1.22e-01
## 27 1.22e-01
## 28 1.33e-01
## 29 1.33e-01
## 30 1.35e-01
## 31 1.35e-01
## 32 1.50e-01
## 33 1.61e-01
## 34 1.61e-01
## 35 1.61e-01
## 36 1.62e-01
## 37 1.64e-01
## 38 1.67e-01
## 39 1.71e-01
## 40 1.74e-01
## [1] ""
## [1] "Table: Gender.x"
## [1] "  (from model: "
## [1] "   ~ Ulcer.diagnosis.at.DATE + Age.x + Gender.x +"
## [1] "     Hba1c_baseline + CALSBP + bmi + Smoking + Statin +"
## [1] "     log_Blood_TGA + Total_cholesterol + egfr)"
## [1] ""
##                             Name Coefficient    CI.L     CI.R  p.value    p.adj
## 1       4-Deoxytetronic acid; 33       0.435  0.2780  0.59300 7.07e-08 5.30e-06
```

```
## 2          Myristoleic acid; 65      -0.379 -0.5380 -0.22000 3.25e-06 8.88e-05
## 3             Glyceric acid; 30      -0.375 -0.5330 -0.21700 3.55e-06 8.88e-05
## 4             Proline, 2TMS; 21       0.363  0.2040  0.52300 7.87e-06 1.48e-04
## 5            Tartronic acid; 73      -0.350 -0.5080 -0.19100 1.65e-05 2.47e-04
## 6           Dodecanoic acid; 54      -0.340 -0.4990 -0.18100 2.98e-05 3.72e-04
## 7          Cholesterol, TMS; 23      -0.315 -0.4700 -0.16000 6.97e-05 7.47e-04
## 8            Oleic acid, TMS; 3      -0.318 -0.4780 -0.15800 9.75e-05 9.14e-04
## 9          Methionine, 2TMS; 16       0.314  0.1550  0.47300 1.14e-04 9.48e-04
## 10         Citric acid, 4TMS; 6      -0.302 -0.4600 -0.14400 1.78e-04 1.34e-03
## 11     Docosahexaenoic acid; 53      -0.291 -0.4490 -0.13300 3.21e-04 2.09e-03
## 12            Glycine, 3TMS; 17      -0.291 -0.4500 -0.13200 3.35e-04 2.09e-03
## 13            Leucine, 2TMS; 19       0.282  0.1230  0.44100 5.00e-04 2.88e-03
## 14             Valine, 2TMS; 20       0.277  0.1200  0.43400 5.55e-04 2.98e-03
## 15            Decanoic acid; 52      -0.277 -0.4360 -0.11800 6.60e-04 3.30e-03
## 16        Aminomalonic acid; 45      -0.272 -0.4310 -0.11200 8.34e-04 3.91e-03
## 17          Stearic acid, TMS; 2     -0.267 -0.4270 -0.10700 1.08e-03 4.78e-03
## 18         Tridecanoic acid; 74      -0.261 -0.4210 -0.10100 1.43e-03 5.67e-03
## 19        Palmitic acid, TMS; 5      -0.258 -0.4180 -0.09920 1.48e-03 5.67e-03
## 20           Hydroxyproline; 64       0.259  0.0992  0.41900 1.51e-03 5.67e-03
## 21       Heptadecanoic acid; 61      -0.256 -0.4160 -0.09540 1.79e-03 6.38e-03
## 22       Heptadecanoic acid; 60      -0.254 -0.4150 -0.09390 1.90e-03 6.49e-03
## 23       Glutamic acid, 3TMS; 8       0.241  0.0831  0.39900 2.79e-03 9.10e-03
## 24          Isoleucine, 2TMS; 18      0.238  0.0804  0.39500 3.09e-03 9.66e-03
## 25 2-hydroxy Isovaleric acid; 38      0.236  0.0763  0.39700 3.82e-03 1.15e-02
## 26        Nonadecanoic acid; 66      -0.232 -0.3920 -0.07110 4.70e-03 1.36e-02
## 27                 Tyrosine; 75      -0.228 -0.3880 -0.06730 5.39e-03 1.50e-02
## 28       Succinic acid, 2TMS; 7      -0.223 -0.3840 -0.06250 6.49e-03 1.74e-02
## 29                 Glycerol; 57      -0.183 -0.3440 -0.02190 2.60e-02 6.72e-02
## 30           1-Monopalmitin; 37       0.174  0.0124  0.33500 3.48e-02 8.70e-02
## 31       11-Eicosenoic acid; 35      -0.164 -0.3240 -0.00299 4.59e-02 1.11e-01
## 32            Nonanoic acid; 67      -0.159 -0.3200  0.00199 5.29e-02 1.24e-01
## 33    Arachidonic acid, TMS; 24      -0.156 -0.3160  0.00513 5.78e-02 1.31e-01
## 34        Benzeneacetic acid; 47     -0.151 -0.3110  0.00832 6.32e-02 1.39e-01
## [1] ""
## [1] "Table: Hba1c_baseline"
## [1] "  (from model: "
## [1] "   ~ Ulcer.diagnosis.at.DATE + Age.x + Gender.x +"
## [1] "     Hba1c_baseline + CALSBP + bmi + Smoking + Statin +"
## [1] "     log_Blood_TGA + Total_cholesterol + egfr)"
## [1] ""
##                          Name Coefficient     CI.L    CI.R  p.value  p.adj
## 1   Eicosapentaenoic acid; 55     -0.1310 -0.20100 -0.0615 0.000235 0.0117
## 2        Tridecanoic acid; 74     -0.1310 -0.20200 -0.0599 0.000312 0.0117
## 3         Arabinopyranose; 51      0.1250  0.05350  0.1960 0.000595 0.0149
## 4    Docosahexaenoic acid; 53     -0.1200 -0.19000 -0.0497 0.000821 0.0154
## 5           Alanine, 2TMS; 25      0.1150  0.04420  0.1860 0.001490 0.0188
## 6           Glyceric acid; 30     -0.1140 -0.18400 -0.0435 0.001510 0.0188
## 7            Ethanolamine; 56      0.1120  0.04040  0.1830 0.002170 0.0232
## 8            Valine, 2TMS; 20      0.1020  0.03200  0.1710 0.004280 0.0401
## 9           Decanoic acid; 52     -0.0952 -0.16600 -0.0246 0.008290 0.0691
## 10 4-Hydroxybutanoic acid; 43     -0.0860 -0.15700 -0.0147 0.018100 0.1340
## 11       Citric acid, 4TMS; 6      0.0835  0.01340  0.1540 0.019600 0.1340
## 12     11-Eicosenoic acid; 35     -0.0823 -0.15400 -0.0110 0.023600 0.1480
## 13            Lactic acid; 29      0.0787  0.00766  0.1500 0.029900 0.1720
```

```
## 14       Myristoleic acid; 65     -0.0772 -0.14800 -0.0066 0.032100 0.1720
## 15     Glyceryl-glycoside; 59      0.0750  0.00421  0.1460 0.037900 0.1890
## [1] ""
## [1] "Table: CALSBP"
## [1] "  (from model: "
## [1] "   ~ Ulcer.diagnosis.at.DATE + Age.x + Gender.x +"
## [1] "     Hba1c_baseline + CALSBP + bmi + Smoking + Statin +"
## [1] "     log_Blood_TGA + Total_cholesterol + egfr)"
## [1] ""
##                   Name Coefficient    CI.L   CI.R p.value  p.adj
## 1 Myristoleic acid; 65     0.00823 0.00361 0.0129 0.00049 0.0368
## [1] ""
## [1] "Table: bmi"
## [1] "  (from model: "
## [1] "   ~ Ulcer.diagnosis.at.DATE + Age.x + Gender.x +"
## [1] "     Hba1c_baseline + CALSBP + bmi + Smoking + Statin +"
## [1] "     log_Blood_TGA + Total_cholesterol + egfr)"
## [1] ""
##                              Name Coefficient      CI.L      CI.R  p.value
## 1          Glutamic acid, 3TMS; 8      0.0456  0.025300  0.065800 1.06e-05
## 2                 Campesterol; 49     -0.0393 -0.059600 -0.019100 1.45e-04
## 3  2-Hydroxybutyric acid, 2TMS; 2      0.0392  0.019000  0.059400 1.48e-04
## 4                 Lactic acid; 29      0.0301  0.009600  0.050700 4.04e-03
## 5                Pyruvic acid; 31     -0.0300 -0.050500 -0.009430 4.26e-03
## 6               Decanoic acid; 52     -0.0292 -0.049600 -0.008770 5.11e-03
## 7              Arachidic acid; 46     -0.0285 -0.049000 -0.007890 6.71e-03
## 8             1,3-Propanediol; 34     -0.0284 -0.049000 -0.007770 6.99e-03
## 9            alpha-Tocopherol; 26     -0.0270 -0.047200 -0.006740 9.01e-03
## 10 2,4-Dihydroxybutanoic acid; 28     -0.0264 -0.046200 -0.006560 9.12e-03
## 11          Pyroglutamic acid; 69     -0.0252 -0.045600 -0.004740 1.58e-02
## 12           Citric acid, 4TMS; 6     -0.0244 -0.044700 -0.004170 1.81e-02
## 13             Tartronic acid; 73     -0.0218 -0.042200 -0.001480 3.56e-02
## 14                    Ribitol; 70      0.0219  0.001410  0.042400 3.62e-02
## 15           Isoleucine, 2TMS; 18      0.0213  0.001130  0.041500 3.85e-02
## 16 alpha-ketoglutaric acid, TMS M     -0.0214 -0.042000 -0.000853 4.12e-02
## 17         11-Eicosenoic acid; 35      0.0214  0.000851  0.042000 4.13e-02
##       p.adj
## 1  0.000791
## 2  0.003690
## 3  0.003690
## 4  0.063800
## 5  0.063800
## 6  0.063800
## 7  0.065500
## 8  0.065500
## 9  0.068400
## 10 0.068400
## 11 0.108000
## 12 0.113000
## 13 0.182000
## 14 0.182000
## 15 0.182000
## 16 0.182000
## 17 0.182000
```

```
## [1] ""
## [1] "Table: Smoking"
## [1] "  (from model: "
## [1] "   ~ Ulcer.diagnosis.at.DATE + Age.x + Gender.x +"
## [1] "     Hba1c_baseline + CALSBP + bmi + Smoking + Statin +"
## [1] "     log_Blood_TGA + Total_cholesterol + egfr)"
## [1] ""
##                              Name Coefficient    CI.L     CI.R  p.value   p.adj
## 1        Docosahexaenoic acid; 53      -0.382 -0.5720 -0.19200 8.46e-05 0.00349
## 2          Glutamic acid, 3TMS; 8       0.375  0.1850  0.56400 1.11e-04 0.00349
## 3      3-Indolepropionic acid; 41      -0.374 -0.5660 -0.18200 1.39e-04 0.00349
## 4              Tartronic acid; 73      -0.349 -0.5390 -0.15800 3.45e-04 0.00622
## 5               Glyceric acid; 30      -0.342 -0.5320 -0.15200 4.15e-04 0.00622
## 6                Valine, 2TMS; 20      -0.311 -0.5000 -0.12200 1.26e-03 0.01580
## 7            alpha-Tocopherol; 26      -0.304 -0.4940 -0.11400 1.71e-03 0.01830
## 8            Citric acid, 4TMS; 6      -0.264 -0.4530 -0.07390 6.50e-03 0.05600
## 9                 Campesterol; 49      -0.260 -0.4500 -0.07030 7.25e-03 0.05600
## 10         Benzeneacetic acid; 47      -0.262 -0.4540 -0.07010 7.47e-03 0.05600
## 11           Malic acid, 3TMS; 11      -0.245 -0.4380 -0.05270 1.26e-02 0.08560
## 12              Leucine, 2TMS; 19      -0.240 -0.4300 -0.04890 1.38e-02 0.08630
## 13 3,4-Dihydroxybutanoic acid; 27       0.223  0.0350  0.41000 2.01e-02 0.11600
## 14                    Ribitol; 70       0.217  0.0252  0.40900 2.67e-02 0.14300
## 15         Heptadecanoic acid; 60      -0.203 -0.3950 -0.01010 3.91e-02 0.18400
## 16         Heptadecanoic acid; 61      -0.202 -0.3940 -0.00890 4.03e-02 0.18400
## 17               Ribonic acid; 72      -0.193 -0.3800 -0.00702 4.20e-02 0.18400
## 18        3-Indoleacetic acid; 40      -0.196 -0.3870 -0.00486 4.45e-02 0.18400
## 19 4-Hydroxyphenyllactic acid; 44      -0.195 -0.3860 -0.00295 4.66e-02 0.18400
## 20           Isoleucine, 2TMS; 18      -0.187 -0.3760  0.00224 5.28e-02 0.19800
## [1] ""
## [1] "Table: Statin"
## [1] "  (from model: "
## [1] "   ~ Ulcer.diagnosis.at.DATE + Age.x + Gender.x +"
## [1] "     Hba1c_baseline + CALSBP + bmi + Smoking + Statin +"
## [1] "     log_Blood_TGA + Total_cholesterol + egfr)"
## [1] ""
##                 Name Coefficient   CI.L   CI.R  p.value p.adj
## 1 L-5-Oxoproline; 63      -0.302 -0.478 -0.126 0.000761 0.057
## [1] ""
## [1] "Table: log_Blood_TGA"
## [1] "  (from model: "
## [1] "   ~ Ulcer.diagnosis.at.DATE + Age.x + Gender.x +"
## [1] "     Hba1c_baseline + CALSBP + bmi + Smoking + Statin +"
## [1] "     log_Blood_TGA + Total_cholesterol + egfr)"
## [1] ""
##                              Name Coefficient      CI.L      CI.R  p.value
## 1           Palmitic acid, TMS; 5       0.293  0.164000  4.21e-01 8.18e-06
## 2             Dodecanoic acid; 54       0.280  0.151000  4.08e-01 2.01e-05
## 3            Stearic acid, TMS; 2       0.278  0.149000  4.07e-01 2.41e-05
## 4               Octanoic acid; 68       0.264  0.135000  3.93e-01 6.33e-05
## 5               Decanoic acid; 52       0.262  0.134000  3.90e-01 6.42e-05
## 6             Arachidic acid; 46       0.250  0.121000  3.80e-01 1.51e-04
## 7  2-Hydroxybutyric acid, 2TMS; 2       0.226  0.099400  3.53e-01 4.81e-04
## 8              Oleic acid, TMS; 3       0.216  0.087000  3.45e-01 1.03e-03
## 9            Isoleucine, 2TMS; 18       0.202  0.075200  3.29e-01 1.82e-03
```

```
## 10              Valine, 2TMS; 20       0.198  0.071700  3.25e-01 2.16e-03
## 11           Arabinopyranose; 51       0.192  0.062800  3.21e-01 3.58e-03
## 12         Glyceryl-glycoside; 59      0.186  0.057500  3.14e-01 4.58e-03
## 13         Heptadecanoic acid; 60      0.181  0.052000  3.11e-01 6.00e-03
## 14                Lactic acid; 29      0.178  0.048900  3.07e-01 6.89e-03
## 15         Heptadecanoic acid; 61      0.171  0.041800  3.00e-01 9.51e-03
## 16         Glutamic acid, 3TMS; 8      0.168  0.040600  2.95e-01 9.76e-03
## 17              Leucine, 2TMS; 19      0.167  0.039100  2.95e-01 1.05e-02
## 18           Aminomalonic acid; 45    -0.167 -0.296000 -3.91e-02 1.06e-02
## 19 3,4-Dihydroxybutanoic acid; 27      0.163  0.037400  2.89e-01 1.10e-02
## 20               Pyruvic acid; 31      0.166  0.036500  2.95e-01 1.20e-02
## 21            Myristoleic acid; 65     0.164  0.035700  2.92e-01 1.23e-02
## 22 4-Hydroxybenzeneacetic acid; 4      0.156  0.030200  2.83e-01 1.52e-02
## 23     4-Hydroxybutanoic acid; 43      0.157  0.027200  2.86e-01 1.78e-02
## 24 alpha-ketoglutaric acid, TMS M      0.154  0.024700  2.83e-01 1.96e-02
## 25               Ribonic acid; 72      0.148  0.022500  2.73e-01 2.08e-02
## 26          Fumaric acid, 2TMS; 9      0.139  0.010400  2.69e-01 3.43e-02
## 27      Arachidonic acid, TMS; 24      0.139  0.009900  2.69e-01 3.49e-02
## 28             Proline, 2TMS; 21       0.134  0.005670  2.62e-01 4.07e-02
## 29           Malic acid, 3TMS; 11      0.131  0.001750  2.60e-01 4.70e-02
## 30     3-Indolepropionic acid; 41     -0.129 -0.258000 -6.46e-05 4.99e-02
## 31         Succinic acid, 2TMS; 7      0.129 -0.000522  2.58e-01 5.09e-02
## 32          Nonadecanoic acid; 66      0.125 -0.003940  2.55e-01 5.74e-02
## 33                    Ribitol; 71      0.120 -0.004490  2.45e-01 5.88e-02
## 34             Tartronic acid; 73     -0.123 -0.251000  5.21e-03 6.01e-02
## 35              Glycine, 3TMS; 17     -0.122 -0.250000  6.12e-03 6.20e-02
## 36              Glyceric acid; 30     -0.120 -0.248000  6.94e-03 6.38e-02
## 37            1,3-Propanediol; 34      0.118 -0.012100  2.47e-01 7.54e-02
## 38            Tridecanoic acid; 74     0.115 -0.014100  2.44e-01 8.07e-02
## 39 3-Hydroxybutyric acid, 2TMS; 1      0.114 -0.015600  2.44e-01 8.47e-02
##       p.adj
## 1  0.000603
## 2  0.000603
## 3  0.000603
## 4  0.000963
## 5  0.000963
## 6  0.001890
## 7  0.005150
## 8  0.009660
## 9  0.015200
## 10 0.016200
## 11 0.024400
## 12 0.028600
## 13 0.034600
## 14 0.036900
## 15 0.043600
## 16 0.043600
## 17 0.043600
## 18 0.043600
## 19 0.043600
## 20 0.043800
## 21 0.043800
## 22 0.051700
## 23 0.057900
```

```
## 24 0.061300
## 25 0.062300
## 26 0.096900
## 27 0.096900
## 28 0.109000
## 29 0.121000
## 30 0.123000
## 31 0.123000
## 32 0.133000
## 33 0.133000
## 34 0.133000
## 35 0.133000
## 36 0.133000
## 37 0.153000
## 38 0.159000
## 39 0.163000
## [1] ""
## [1] "Table: Total_cholesterol"
## [1] "  (from model: "
## [1] "    ~ Ulcer.diagnosis.at.DATE + Age.x + Gender.x +"
## [1] "      Hba1c_baseline + CALSBP + bmi + Smoking + Statin +"
## [1] "      log_Blood_TGA + Total_cholesterol + egfr)"
## [1] ""
##                               Name Coefficient    CI.L     CI.R  p.value
## 1            Cholesterol, TMS; 23      0.4550  0.3620  0.54800 2.75e-21
## 2                 Campesterol; 49      0.3710  0.2760  0.46600 2.81e-14
## 3            alpha-Tocopherol; 26      0.3240  0.2290  0.41800 2.81e-11
## 4          Benzeneacetic acid; 47     -0.1810 -0.2770 -0.08560 2.11e-04
## 5           Linoleic acid, TMS; 4      0.1790  0.0833  0.27500 2.55e-04
## 6      4-Hydroxybutanoic acid; 43     -0.1770 -0.2730 -0.08030 3.33e-04
## 7               Proline, 2TMS; 21     -0.1710 -0.2660 -0.07540 4.58e-04
## 8            Isoleucine, 2TMS; 18     -0.1670 -0.2610 -0.07230 5.51e-04
## 9              L-5-Oxoproline; 63     -0.1660 -0.2630 -0.07020 7.07e-04
## 10              Glycine, 3TMS; 17     -0.1590 -0.2550 -0.06400 1.06e-03
## 11                   Tyrosine; 75     -0.1580 -0.2540 -0.06150 1.33e-03
## 12           Methionine, 2TMS; 16     -0.1500 -0.2450 -0.05410 2.16e-03
## 13 2,4-Dihydroxybutanoic acid; 28     -0.1400 -0.2330 -0.04740 3.10e-03
## 14             Arabinopyranose; 51     -0.1440 -0.2400 -0.04760 3.39e-03
## 15        Docosahexaenoic acid; 53      0.1390  0.0444  0.23400 4.04e-03
## 16       Eicosapentaenoic acid; 55      0.1360  0.0415  0.23100 4.81e-03
## 17            Threonine, 3TMS; 12     -0.1380 -0.2340 -0.04110 5.22e-03
## 18 4-Hydroxybenzeneacetic acid; 4     -0.1310 -0.2250 -0.03740 6.19e-03
## 19              Alanine, 2TMS; 25     -0.1330 -0.2290 -0.03690 6.71e-03
## 20        2-Palmitoylglycerol; 39      0.1330  0.0369  0.23000 6.76e-03
## 21         Glyceryl-glycoside; 59     -0.1270 -0.2230 -0.03150 9.23e-03
## 22               Hydroxylamine; 62     -0.1250 -0.2210 -0.02820 1.13e-02
## 23               Serine, 3TMS; 14     -0.1210 -0.2170 -0.02500 1.35e-02
## 24                    Ribitol; 71     -0.1150 -0.2080 -0.02240 1.50e-02
## 25         3-Indoleacetic acid; 40     -0.1140 -0.2100 -0.01900 1.88e-02
## 26               Ribonic acid; 72     -0.1010 -0.1940 -0.00745 3.43e-02
## 27        Phenylalanine, 2TMS; 13     -0.1040 -0.2010 -0.00770 3.43e-02
## 28               Pyruvic acid; 31     -0.1020 -0.1990 -0.00629 3.68e-02
## 29            Malic acid, 3TMS; 11     -0.1010 -0.1970 -0.00500 3.92e-02
## 30 3,4-Dihydroxybutanoic acid; 27     -0.0908 -0.1850  0.00293 5.76e-02
```

```
## 31             Leucine, 2TMS; 19     -0.0912 -0.1860  0.00409 6.07e-02
## 32              Ethanolamine; 56     -0.0893 -0.1860  0.00718 6.96e-02
## 33              Valine, 2TMS; 20     -0.0867 -0.1810  0.00763 7.16e-02
## 34           1,3-Propanediol; 34     -0.0852 -0.1820  0.01150 8.40e-02
## 35          Palmitic acid, TMS; 5      0.0840 -0.0115  0.18000 8.48e-02
## 36     3-Indolepropionic acid; 41      0.0828 -0.0131  0.17900 9.06e-02
##       p.adj
## 1  2.06e-19
## 2  1.05e-12
## 3  7.02e-10
## 4  3.83e-03
## 5  3.83e-03
## 6  4.16e-03
## 7  4.91e-03
## 8  5.16e-03
## 9  5.89e-03
## 10 7.99e-03
## 11 9.06e-03
## 12 1.35e-02
## 13 1.79e-02
## 14 1.81e-02
## 15 2.02e-02
## 16 2.25e-02
## 17 2.30e-02
## 18 2.53e-02
## 19 2.53e-02
## 20 2.53e-02
## 21 3.30e-02
## 22 3.87e-02
## 23 4.40e-02
## 24 4.68e-02
## 25 5.64e-02
## 26 9.54e-02
## 27 9.54e-02
## 28 9.86e-02
## 29 1.01e-01
## 30 1.44e-01
## 31 1.47e-01
## 32 1.63e-01
## 33 1.63e-01
## 34 1.82e-01
## 35 1.82e-01
## 36 1.89e-01
## [1] ""
## [1] "Table: egfr"
## [1] "  (from model: "
## [1] "   ~ Ulcer.diagnosis.at.DATE + Age.x + Gender.x +"
## [1] "     Hba1c_baseline + CALSBP + bmi + Smoking + Statin +"
## [1] "     log_Blood_TGA + Total_cholesterol + egfr)"
## [1] ""
##                              Name Coefficient      CI.L      CI.R  p.value
## 1            Myo inositol 6TMS; 1    -0.01650 -0.019400 -1.36e-02 9.38e-28
## 2                     Ribitol; 71    -0.01580 -0.018700 -1.29e-02 1.87e-25
## 3                  Creatinine; 50    -0.01540 -0.018300 -1.24e-02 1.09e-23
```

```
## 4  2,4-Dihydroxybutanoic acid; 28    -0.01490 -0.017800 -1.19e-02 8.90e-23
## 5                Ribonic acid; 72    -0.01480 -0.017800 -1.19e-02 1.36e-22
## 6  3,4-Dihydroxybutanoic acid; 27    -0.01320 -0.016100 -1.02e-02 4.21e-18
## 7  4-Hydroxybenzeneacetic acid; 4    -0.01150 -0.014400 -8.51e-03 4.94e-14
## 8        4-Deoxytetronic acid; 33    -0.01110 -0.014100 -8.13e-03 4.24e-13
## 9        4-Deoxytetronic acid; 32    -0.01030 -0.013400 -7.33e-03 2.23e-11
## 10           Isoleucine, 2TMS; 18     0.00947  0.006490  1.24e-02 5.60e-10
## 11 2-Hydroxybutyric acid, 2TMS; 2     0.00911  0.006130  1.21e-02 2.43e-09
## 12               Valine, 2TMS; 20     0.00873  0.005760  1.17e-02 9.77e-09
## 13           Citric acid, 4TMS; 6    -0.00831 -0.011300 -5.33e-03 5.46e-08
## 14          Pyroglutamic acid; 69    -0.00817 -0.011200 -5.15e-03 1.24e-07
## 15              Hydroxyproline; 64    -0.00774 -0.010800 -4.72e-03 5.73e-07
## 16        3-Indoleacetic acid; 40    -0.00709 -0.010100 -4.08e-03 4.00e-06
## 17         Glyceryl-glycoside; 59    -0.00696 -0.009970 -3.95e-03 6.32e-06
## 18 4-Hydroxyphenyllactic acid; 44    -0.00641 -0.009420 -3.39e-03 3.30e-05
## 19              Leucine, 2TMS; 19     0.00622  0.003220  9.22e-03 5.07e-05
## 20               Serine, 3TMS; 14     0.00614  0.003120  9.16e-03 7.07e-05
## 21              Octanoic acid; 68     0.00574  0.002710  8.77e-03 2.06e-04
## 22           Methionine, 2TMS; 16     0.00566  0.002640  8.67e-03 2.37e-04
## 23           Stearic acid, TMS; 2     0.00539  0.002360  8.41e-03 4.87e-04
## 24                   Tyrosine; 75     0.00535  0.002320  8.38e-03 5.49e-04
## 25              Glycine, 3TMS; 17    -0.00507 -0.008080 -2.07e-03 9.48e-04
## 26          Fumaric acid, 2TMS; 9    -0.00501 -0.008040 -1.97e-03 1.22e-03
## 27      Eicosapentaenoic acid; 55     0.00455  0.001570  7.53e-03 2.77e-03
## 28  2-hydroxy Isovaleric acid; 38     0.00458  0.001550  7.61e-03 3.06e-03
## 29           Malic acid, 3TMS; 11    -0.00421 -0.007230 -1.18e-03 6.54e-03
## 30          Palmitic acid, TMS; 5     0.00407  0.001060  7.08e-03 8.10e-03
## 31           Cholesterol, TMS; 23     0.00385  0.000918  6.78e-03 1.01e-02
## 32         Benzeneacetic acid; 47    -0.00384 -0.006860 -8.21e-04 1.27e-02
## 33                   Glycerol; 57     0.00368  0.000637  6.73e-03 1.78e-02
## 34       Docosahexaenoic acid; 53     0.00358  0.000589  6.57e-03 1.90e-02
## 35              Alanine, 2TMS; 25    -0.00353 -0.006560 -5.06e-04 2.22e-02
## 36              Glyceric acid; 30     0.00347  0.000481  6.46e-03 2.29e-02
## 37                   Glycerol; 58    -0.00307 -0.006130 -2.35e-05 4.83e-02
##       p.adj
## 1  7.03e-26
## 2  7.02e-24
## 3  2.73e-22
## 4  1.67e-21
## 5  2.03e-21
## 6  5.27e-17
## 7  5.29e-13
## 8  3.97e-12
## 9  1.85e-10
## 10 4.20e-09
## 11 1.65e-08
## 12 6.11e-08
## 13 3.15e-07
## 14 6.62e-07
## 15 2.87e-06
## 16 1.88e-05
## 17 2.79e-05
## 18 1.37e-04
## 19 2.00e-04
```

```
## 20 2.65e-04
## 21 7.36e-04
## 22 8.09e-04
## 23 1.59e-03
## 24 1.72e-03
## 25 2.84e-03
## 26 3.52e-03
## 27 7.69e-03
## 28 8.21e-03
## 29 1.69e-02
## 30 2.02e-02
## 31 2.44e-02
## 32 2.98e-02
## 33 4.05e-02
## 34 4.19e-02
## 35 4.75e-02
## 36 4.77e-02
## 37 9.78e-02
```

###### 6.1.3.3 Table with All Metabolites

```
## [1] ""
## [1] "Table: Ulcer.diagnosis.at.DATEJA"
## [1] "  (from model: "
## [1] "   ~ Ulcer.diagnosis.at.DATE + Age.x + Gender.x +"
## [1] "     Hba1c_baseline + CALSBP + bmi + Smoking + Statin +"
## [1] "     log_Blood_TGA + Total_cholesterol + egfr)"
## [1] ""
##                               Name Coefficient   CI.L   CI.R p.value p.adj
## 1            Citric acid, 4TMS; 6    -0.62200 -1.090 -0.151 0.00962 0.722
## 2  2-Hydroxybutyric acid, 2TMS; 2    -0.36400 -0.833  0.106 0.12900 0.927
## 3                     Tyrosine; 75    0.36700 -0.110  0.844 0.13200 0.927
## 4          Benzeneacetic acid; 47    -0.36300 -0.839  0.112 0.13400 0.927
## 5        4-Deoxytetronic acid; 33    -0.33900 -0.809  0.131 0.15800 0.927
## 6   2-hydroxy Isovaleric acid; 38    -0.32800 -0.805  0.149 0.17700 0.927
## 7            Cholesterol, TMS; 23    -0.31600 -0.778  0.145 0.17900 0.927
## 8          Glyceryl-glycoside; 59    -0.31500 -0.789  0.160 0.19300 0.927
## 9           Palmitic acid, TMS; 5     0.31000 -0.164  0.784 0.20000 0.927
## 10 4-Hydroxybenzeneacetic acid; 4    -0.29900 -0.765  0.168 0.20900 0.927
## 11              Nonanoic acid; 67     0.29800 -0.181  0.777 0.22300 0.927
## 12                   Glycerol; 58     0.29400 -0.186  0.774 0.23000 0.927
## 13               Serine, 3TMS; 14    -0.28800 -0.764  0.188 0.23500 0.927
## 14             L-5-Oxoproline; 63    -0.28400 -0.761  0.193 0.24300 0.927
## 15         Nonadecanoic acid; 66      0.28100 -0.197  0.759 0.24900 0.927
## 16           Myristoleic acid; 65     0.26700 -0.207  0.741 0.26900 0.927
## 17           Methionine, 2TMS; 16     0.26400 -0.210  0.738 0.27500 0.927
## 18 2,4-Dihydroxybutanoic acid; 28    -0.25100 -0.712  0.209 0.28500 0.927
## 19            Stearic acid, TMS; 2    0.25700 -0.219  0.733 0.29000 0.927
## 20          Heptadecanoic acid; 61    0.25400 -0.223  0.732 0.29700 0.927
## 21                    Ribitol; 71     0.23800 -0.223  0.698 0.31200 0.927
## 22           alpha-Tocopherol; 26    -0.23400 -0.704  0.236 0.32900 0.927
## 23          Aminomalonic acid; 45    -0.21800 -0.692  0.256 0.36700 0.927
## 24           Fumaric acid, 2TMS; 9   -0.20900 -0.686  0.268 0.39100 0.927
## 25                    Ribitol; 70    -0.20800 -0.684  0.268 0.39200 0.927
## 26           Malic acid, 3TMS; 11     0.20600 -0.271  0.683 0.39700 0.927
## 27             1,3-Propanediol; 34    0.19900 -0.280  0.679 0.41500 0.927
## 28         3-Indoleacetic acid; 40   -0.19300 -0.666  0.281 0.42400 0.927
## 29            Tridecanoic acid; 74    0.19000 -0.287  0.668 0.43500 0.927
## 30 3,4-Dihydroxybutanoic acid; 27    -0.17700 -0.642  0.288 0.45600 0.927
## 31                Campesterol; 49    -0.17700 -0.648  0.293 0.46000 0.927
## 32              Glyceric acid; 30    -0.16900 -0.640  0.301 0.48000 0.927
## 33             1-Monopalmitin; 37    -0.17200 -0.652  0.307 0.48100 0.927
## 34          Glutamic acid, 3TMS; 8   -0.16700 -0.637  0.303 0.48700 0.927
## 35               Ethanolamine; 56     0.16200 -0.317  0.641 0.50600 0.927
## 36 alpha-ketoglutaric acid, TMS M     0.15500 -0.323  0.633 0.52400 0.927
## 37       Docosahexaenoic acid; 53    -0.14700 -0.618  0.323 0.53900 0.927
## 38              Proline, 2TMS; 21     0.14400 -0.329  0.618 0.55000 0.927
## 39             Arachidic acid; 46     0.14300 -0.335  0.621 0.55700 0.927
## 40               Ribonic acid; 72     0.13600 -0.326  0.599 0.56300 0.927
## 41          Heptadecanoic acid; 60    0.13900 -0.338  0.617 0.56700 0.927
## 42              Glycine, 3TMS; 17    -0.13600 -0.609  0.337 0.57200 0.927
## 43         11-Eicosenoic acid; 35     0.13700 -0.341  0.615 0.57400 0.927
## 44              Decanoic acid; 52     0.13100 -0.343  0.605 0.58800 0.927
```

```
## 45 4-Hydroxyphenyllactic acid; 44    -0.12800 -0.603  0.348 0.59800 0.927
## 46         Succinic acid, 2TMS; 7     0.12800 -0.350  0.606 0.59900 0.927
## 47              Hydroxylamine; 62     0.12500 -0.354  0.604 0.60900 0.927
## 48                Bisphenol A; 48    -0.11800 -0.599  0.362 0.63000 0.927
## 49           Isoleucine, 2TMS; 18    -0.11200 -0.581  0.357 0.63900 0.927
## 50             Tartronic acid; 73    -0.11100 -0.583  0.362 0.64700 0.927
## 51             Oleic acid, TMS; 3     0.10800 -0.368  0.584 0.65600 0.927
## 52     4-Hydroxybutanoic acid; 43    -0.10700 -0.586  0.371 0.66000 0.927
## 53             Hydroxyproline; 64    -0.09880 -0.575  0.378 0.68400 0.927
## 54              Octanoic acid; 68     0.09680 -0.380  0.573 0.69000 0.927
## 55          Linoleic acid, TMS; 4    -0.09540 -0.572  0.381 0.69500 0.927
## 56      Eicosapentaenoic acid; 55     0.09060 -0.378  0.560 0.70500 0.927
## 57                1-Dodecanol; 36     0.08810 -0.392  0.568 0.71900 0.927
## 58       4-Deoxytetronic acid; 32    -0.08410 -0.558  0.390 0.72800 0.927
## 59           Threonine, 3TMS; 12     0.08020 -0.398  0.559 0.74200 0.927
## 60        2-Palmitoylglycerol; 39    -0.07690 -0.556  0.402 0.75300 0.927
## 61              Leucine, 2TMS; 19     0.07460 -0.398  0.547 0.75700 0.927
## 62              Alanine, 2TMS; 25    -0.07230 -0.549  0.404 0.76600 0.927
## 63                 Creatinine; 50    -0.05080 -0.518  0.416 0.83100 0.982
## 64           Pyroglutamic acid; 69    -0.04420 -0.519  0.431 0.85500 0.982
## 65        Phenylalanine, 2TMS; 13    -0.04150 -0.521  0.438 0.86500 0.982
## 66      Arachidonic acid, TMS; 24    -0.03440 -0.513  0.444 0.88800 0.982
## 67               Pyruvic acid; 31     0.02930 -0.448  0.506 0.90400 0.982
## 68 3-Hydroxybutyric acid, 2TMS; 1     0.02010 -0.460  0.500 0.93400 0.982
## 69                Lactic acid; 29    -0.01800 -0.495  0.459 0.94100 0.982
## 70                   Glycerol; 57     0.01560 -0.464  0.495 0.94900 0.982
## 71     3-Indolepropionic acid; 41     0.01460 -0.461  0.491 0.95200 0.982
## 72            Dodecanoic acid; 54     0.01190 -0.463  0.487 0.96100 0.982
## 73           Myo inositol 6TMS; 1     0.00929 -0.450  0.468 0.96800 0.982
## 74             Arabinopyranose; 51    -0.00936 -0.486  0.467 0.96900 0.982
## 75               Valine, 2TMS; 20     0.00142 -0.466  0.469 0.99500 0.995
```

##### 6.1.4 Fully-Adjusted Model

```
## [1] "Fitting models:"
## [1] "~ Ulcer.diagnosis.at.DATE + Age.x + Gender.x + Hba1c_baseline + CALSBP + bmi + Smoking + Statin
## [1] ""
```

###### 6.1.4.1 Forest Plot of Model Coefficients

```
## Warning: Ignoring unknown aesthetics: x
## Ignoring unknown aesthetics: x
## Ignoring unknown aesthetics: x
## Ignoring unknown aesthetics: x
## Ignoring unknown aesthetics: x
## Ignoring unknown aesthetics: x
## Ignoring unknown aesthetics: x
## Ignoring unknown aesthetics: x
## Ignoring unknown aesthetics: x
## Ignoring unknown aesthetics: x
## Ignoring unknown aesthetics: x
```

```
## NULL
```

###### 6.1.4.2 Tables of Model Coefficients

```
## [1] ""
## [1] "Table: Ulcer.diagnosis.at.DATEJA"
## [1] "  (from model: "
## [1] "   ~ Ulcer.diagnosis.at.DATE + Age.x + Gender.x +"
## [1] "     Hba1c_baseline + CALSBP + bmi + Smoking + Statin +"
## [1] "     log_Blood_TGA + Total_cholesterol + egfr + logUAER)"
## [1] ""
## [1] "No significant associations at p.adj < 0.2"
## [1] ""
## [1] "Table: Age.x"
## [1] "  (from model: "
## [1] "   ~ Ulcer.diagnosis.at.DATE + Age.x + Gender.x +"
## [1] "     Hba1c_baseline + CALSBP + bmi + Smoking + Statin +"
## [1] "     log_Blood_TGA + Total_cholesterol + egfr + logUAER)"
## [1] ""
##                               Name Coefficient      CI.L     CI.R  p.value
## 1       Eicosapentaenoic acid; 55     0.02540  0.018000 0.032700 2.18e-11
## 2        Docosahexaenoic acid; 53     0.01390  0.006530 0.021400 2.32e-04
## 3  alpha-ketoglutaric acid, TMS M     0.01250  0.004970 0.020100 1.16e-03
## 4                Pyruvic acid; 31     0.01200  0.004480 0.019600 1.80e-03
## 5             alpha-Tocopherol; 26     0.01150  0.004120 0.018900 2.30e-03
## 6               Decanoic acid; 52     0.01060  0.003080 0.018100 5.70e-03
## 7             Malic acid, 3TMS; 11     0.01060  0.003080 0.018100 5.74e-03
## 8            Palmitic acid, TMS; 5     0.01040  0.002950 0.017900 6.31e-03
## 9          3-Indoleacetic acid; 40     0.01030  0.002840 0.017800 6.83e-03
## 10 4-Hydroxybenzeneacetic acid; 4     0.01010  0.002760 0.017400 7.05e-03
## 11              Glyceric acid; 30     0.00952  0.002120 0.016900 1.18e-02
## 12                    Ribitol; 70     0.00957  0.002050 0.017100 1.26e-02
## 13           Nonadecanoic acid; 66     0.00954  0.002000 0.017100 1.32e-02
## 14              Oleic acid, TMS; 3     0.00911  0.001600 0.016600 1.75e-02
## 15            Myristoleic acid; 65     0.00872  0.001230 0.016200 2.24e-02
## 16          Glutamic acid, 3TMS; 8     0.00845  0.001060 0.015800 2.51e-02
## 17 2,4-Dihydroxybutanoic acid; 28     0.00818  0.000965 0.015400 2.63e-02
## 18          Succinic acid, 2TMS; 7     0.00835  0.000811 0.015900 3.00e-02
## 19           Fumaric acid, 2TMS; 9     0.00804  0.000517 0.015600 3.62e-02
## 20          11-Eicosenoic acid; 35     0.00787  0.000312 0.015400 4.13e-02
## 21            Stearic acid, TMS; 2     0.00775  0.000236 0.015300 4.32e-02
## 22               Alanine, 2TMS; 25     0.00772  0.000200 0.015200 4.42e-02
## 23              Tartronic acid; 73     0.00757  0.000111 0.015000 4.67e-02
## 24         Heptadecanoic acid; 61     0.00736 -0.000182 0.014900 5.58e-02
## 25                   Glycerol; 57     0.00737 -0.000202 0.014900 5.64e-02
## 26             Dodecanoic acid; 54     0.00724 -0.000268 0.014700 5.87e-02
## 27            Myo inositol 6TMS; 1     0.00691 -0.000268 0.014100 5.92e-02
## 28                   Tyrosine; 75     0.00699 -0.000537 0.014500 6.87e-02
## 29              Hydroxylamine; 62    -0.00701 -0.014600 0.000561 6.95e-02
##       p.adj
## 1  1.64e-09
## 2  8.69e-03
## 3  2.91e-02
## 4  3.37e-02
## 5  3.45e-02
## 6  5.29e-02
```

```
## 7  5.29e-02
## 8  5.29e-02
## 9  5.29e-02
## 10 5.29e-02
## 11 7.61e-02
## 12 7.61e-02
## 13 7.61e-02
## 14 9.38e-02
## 15 1.12e-01
## 16 1.16e-01
## 17 1.16e-01
## 18 1.25e-01
## 19 1.43e-01
## 20 1.51e-01
## 21 1.51e-01
## 22 1.51e-01
## 23 1.52e-01
## 24 1.64e-01
## 25 1.64e-01
## 26 1.64e-01
## 27 1.64e-01
## 28 1.80e-01
## 29 1.80e-01
## [1] ""
## [1] "Table: Gender.x"
## [1] "  (from model: "
## [1] "   ~ Ulcer.diagnosis.at.DATE + Age.x + Gender.x +"
## [1] "     Hba1c_baseline + CALSBP + bmi + Smoking + Statin +"
## [1] "     log_Blood_TGA + Total_cholesterol + egfr + logUAER)"
## [1] ""
##                             Name Coefficient     CI.L     CI.R  p.value
## 1       4-Deoxytetronic acid; 33       0.406  0.24200  0.57100 1.38e-06
## 2             Tartronic acid; 73      -0.385 -0.55100 -0.21900 5.58e-06
## 3              Glyceric acid; 30      -0.364 -0.52800 -0.19900 1.57e-05
## 4              Proline, 2TMS; 21       0.360  0.19400  0.52600 2.21e-05
## 5           Methionine, 2TMS; 16       0.359  0.19300  0.52500 2.40e-05
## 6           Myristoleic acid; 65      -0.357 -0.52300 -0.19000 2.76e-05
## 7               Valine, 2TMS; 20       0.338  0.17400  0.50100 5.36e-05
## 8              Leucine, 2TMS; 19       0.313  0.14700  0.47900 2.18e-04
## 9           Cholesterol, TMS; 23      -0.301 -0.46300 -0.14000 2.54e-04
## 10            Oleic acid, TMS; 3      -0.301 -0.46800 -0.13400 4.28e-04
## 11           Dodecanoic acid; 54      -0.293 -0.46000 -0.12600 5.89e-04
## 12          Citric acid, 4TMS; 6      -0.285 -0.45000 -0.12000 7.15e-04
## 13             Glycine, 3TMS; 17      -0.279 -0.44500 -0.11300 1.00e-03
## 14          Isoleucine, 2TMS; 18       0.268  0.10300  0.43200 1.43e-03
## 15            Hydroxyproline; 64       0.266  0.09910  0.43400 1.82e-03
## 16 2-hydroxy Isovaleric acid; 38       0.255  0.08780  0.42300 2.83e-03
## 17             Decanoic acid; 52      -0.251 -0.41800 -0.08420 3.20e-03
## 18         Aminomalonic acid; 45      -0.250 -0.41600 -0.08360 3.24e-03
## 19      Docosahexaenoic acid; 53      -0.246 -0.41100 -0.08070 3.54e-03
## 20          Stearic acid, TMS; 2      -0.248 -0.41500 -0.08100 3.65e-03
## 21        Glutamic acid, 3TMS; 8       0.226  0.06140  0.39000 7.14e-03
## 22        Succinic acid, 2TMS; 7      -0.228 -0.39600 -0.06060 7.66e-03
## 23         Palmitic acid, TMS; 5      -0.225 -0.39100 -0.05830 8.17e-03
```

```
## 24          Tridecanoic acid; 74      -0.222 -0.39000 -0.05440 9.48e-03
## 25         Nonadecanoic acid; 66      -0.209 -0.37700 -0.04130 1.46e-02
## 26        Heptadecanoic acid; 61      -0.204 -0.37200 -0.03670 1.69e-02
## 27        Heptadecanoic acid; 60      -0.190 -0.35700 -0.02170 2.69e-02
## 28                  Tyrosine; 75      -0.184 -0.35200 -0.01700 3.09e-02
## 29                  Glycerol; 57      -0.167 -0.33500  0.00184 5.26e-02
## 30            1-Monopalmitin; 37       0.165 -0.00352  0.33400 5.50e-02
## 31        Benzeneacetic acid; 47      -0.162 -0.32900  0.00461 5.67e-02
##       p.adj
## 1  0.000104
## 2  0.000209
## 3  0.000345
## 4  0.000345
## 5  0.000345
## 6  0.000345
## 7  0.000574
## 8  0.002050
## 9  0.002110
## 10 0.003210
## 11 0.004020
## 12 0.004470
## 13 0.005780
## 14 0.007650
## 15 0.009080
## 16 0.013300
## 17 0.013500
## 18 0.013500
## 19 0.013700
## 20 0.013700
## 21 0.025500
## 22 0.026100
## 23 0.026600
## 24 0.029600
## 25 0.043900
## 26 0.048900
## 27 0.074600
## 28 0.082700
## 29 0.136000
## 30 0.137000
## 31 0.137000
## [1] ""
## [1] "Table: Hba1c_baseline"
## [1] "  (from model: "
## [1] "   ~ Ulcer.diagnosis.at.DATE + Age.x + Gender.x +"
## [1] "     Hba1c_baseline + CALSBP + bmi + Smoking + Statin +"
## [1] "     log_Blood_TGA + Total_cholesterol + egfr + logUAER)"
## [1] ""
##                          Name Coefficient     CI.L     CI.R  p.value  p.adj
## 1            Valine, 2TMS; 20      0.1330  0.06010  0.20600 0.000363 0.0118
## 2            Ethanolamine; 56      0.1350  0.05950  0.21000 0.000448 0.0118
## 3           Alanine, 2TMS; 25      0.1290  0.05470  0.20400 0.000696 0.0118
## 4   Eicosapentaenoic acid; 55     -0.1260 -0.19900 -0.05250 0.000783 0.0118
## 5         Arabinopyranose; 51      0.1280  0.05340  0.20300 0.000788 0.0118
## 6    Docosahexaenoic acid; 53     -0.1100 -0.18400 -0.03680 0.003310 0.0384
```

```
## 7        Citric acid, 4TMS; 6      0.1100  0.03590  0.18300 0.003590 0.0384
## 8           Decanoic acid; 52     -0.0953 -0.17000 -0.02080 0.012200 0.0921
## 9           Glyceric acid; 30     -0.0941 -0.16800 -0.02050 0.012300 0.0921
## 10 4-Hydroxybutanoic acid; 43     -0.0955 -0.17100 -0.02050 0.012700 0.0921
## 11       Tridecanoic acid; 74     -0.0945 -0.16900 -0.01950 0.013500 0.0921
## 12            Campesterol; 49      0.0853  0.01190  0.15900 0.022800 0.1420
## 13     Glyceryl-glycoside; 59      0.0845  0.01020  0.15900 0.025900 0.1490
## 14            Lactic acid; 29      0.0834  0.00870  0.15800 0.028700 0.1540
## 15           Leucine, 2TMS; 19     0.0803  0.00633  0.15400 0.033400 0.1670
## 16   4-Deoxytetronic acid; 33     -0.0771 -0.15000 -0.00363 0.039700 0.1830
## 17       Myristoleic acid; 65     -0.0773 -0.15200 -0.00297 0.041500 0.1830
## 18                Ribitol; 70     -0.0767 -0.15100 -0.00201 0.044200 0.1840
## [1] ""
## [1] "Table: CALSBP"
## [1] "  (from model: "
## [1] "   ~ Ulcer.diagnosis.at.DATE + Age.x + Gender.x +"
## [1] "     Hba1c_baseline + CALSBP + bmi + Smoking + Statin +"
## [1] "     log_Blood_TGA + Total_cholesterol + egfr + logUAER)"
## [1] ""
##                   Name Coefficient    CI.L   CI.R p.value p.adj
## 1 Myristoleic acid; 65     0.00793 0.00302 0.0129 0.00158 0.118
## [1] ""
## [1] "Table: bmi"
## [1] "  (from model: "
## [1] "   ~ Ulcer.diagnosis.at.DATE + Age.x + Gender.x +"
## [1] "     Hba1c_baseline + CALSBP + bmi + Smoking + Statin +"
## [1] "     log_Blood_TGA + Total_cholesterol + egfr + logUAER)"
## [1] ""
##                              Name Coefficient      CI.L     CI.R  p.value
## 1          Glutamic acid, 3TMS; 8      0.0442  0.023600  0.06480 2.76e-05
## 2                 Campesterol; 49     -0.0405 -0.061100 -0.01990 1.22e-04
## 3  2-Hydroxybutyric acid, 2TMS; 2      0.0402  0.019600  0.06080 1.30e-04
## 4               Decanoic acid; 52     -0.0351 -0.056000 -0.01430 9.86e-04
## 5  2,4-Dihydroxybutanoic acid; 28     -0.0290 -0.049100 -0.00888 4.76e-03
## 6                 Lactic acid; 29      0.0299  0.008900  0.05080 5.27e-03
## 7                Pyruvic acid; 31     -0.0290 -0.050000 -0.00803 6.78e-03
## 8            alpha-Tocopherol; 26     -0.0284 -0.049000 -0.00773 7.08e-03
## 9             1,3-Propanediol; 34     -0.0285 -0.049600 -0.00740 8.15e-03
## 10             Arachidic acid; 46     -0.0271 -0.048100 -0.00611 1.14e-02
## 11           Citric acid, 4TMS; 6     -0.0266 -0.047300 -0.00595 1.16e-02
## 12          Pyroglutamic acid; 69     -0.0258 -0.046600 -0.00489 1.56e-02
## 13            Dodecanoic acid; 54     -0.0240 -0.044900 -0.00308 2.46e-02
## 14          Nonadecanoic acid; 66     -0.0241 -0.045100 -0.00309 2.46e-02
## 15         11-Eicosenoic acid; 35      0.0241  0.003020  0.04520 2.50e-02
## 16             Tartronic acid; 73     -0.0225 -0.043300 -0.00172 3.38e-02
## 17              Glycine, 3TMS; 17     -0.0224 -0.043200 -0.00160 3.48e-02
## 18 alpha-ketoglutaric acid, TMS M     -0.0225 -0.043600 -0.00151 3.57e-02
## 19          Isoleucine, 2TMS; 18      0.0218  0.001180  0.04230 3.82e-02
## 20                    Ribitol; 70      0.0211  0.000183  0.04210 4.80e-02
##      p.adj
## 1  0.00207
## 2  0.00324
## 3  0.00324
## 4  0.01850
```

```
## 5  0.06580
## 6  0.06580
## 7  0.06630
## 8  0.06630
## 9  0.06790
## 10 0.07930
## 11 0.07930
## 12 0.09740
## 13 0.12500
## 14 0.12500
## 15 0.12500
## 16 0.14900
## 17 0.14900
## 18 0.14900
## 19 0.15100
## 20 0.18000
## [1] ""
## [1] "Table: Smoking"
## [1] "  (from model: "
## [1] "   ~ Ulcer.diagnosis.at.DATE + Age.x + Gender.x +"
## [1] "     Hba1c_baseline + CALSBP + bmi + Smoking + Statin +"
## [1] "     log_Blood_TGA + Total_cholesterol + egfr + logUAER)"
## [1] ""
##                           Name Coefficient   CI.L    CI.R  p.value  p.adj
## 1  3-Indolepropionic acid; 41      -0.352 -0.551 -0.1530 0.000524 0.0167
## 2    Docosahexaenoic acid; 53      -0.338 -0.534 -0.1420 0.000740 0.0167
## 3          Tartronic acid; 73      -0.338 -0.536 -0.1410 0.000782 0.0167
## 4      Glutamic acid, 3TMS; 8       0.332  0.136  0.5280 0.000891 0.0167
## 5            Valine, 2TMS; 20      -0.307 -0.502 -0.1120 0.002010 0.0301
## 6        alpha-Tocopherol; 26      -0.297 -0.493 -0.1010 0.002990 0.0331
## 7           Glyceric acid; 30      -0.296 -0.492 -0.1000 0.003090 0.0331
## 8      Benzeneacetic acid; 47      -0.266 -0.465 -0.0674 0.008700 0.0748
## 9        Citric acid, 4TMS; 6      -0.262 -0.458 -0.0655 0.008980 0.0748
## 10       Malic acid, 3TMS; 11      -0.257 -0.456 -0.0580 0.011400 0.0790
## 11          Leucine, 2TMS; 19      -0.254 -0.451 -0.0569 0.011600 0.0790
## 12           Ribonic acid; 72      -0.242 -0.433 -0.0500 0.013500 0.0842
## 13            Campesterol; 49      -0.229 -0.425 -0.0339 0.021500 0.1240
## [1] ""
## [1] "Table: Statin"
## [1] "  (from model: "
## [1] "   ~ Ulcer.diagnosis.at.DATE + Age.x + Gender.x +"
## [1] "     Hba1c_baseline + CALSBP + bmi + Smoking + Statin +"
## [1] "     log_Blood_TGA + Total_cholesterol + egfr + logUAER)"
## [1] ""
##                         Name Coefficient    CI.L    CI.R p.value p.adj
## 1           Campesterol; 49       0.294  0.1110  0.4760 0.00161 0.107
## 2 Arachidonic acid, TMS; 24       0.278  0.0913  0.4640 0.00353 0.107
## 3        L-5-Oxoproline; 63      -0.272 -0.4580 -0.0854 0.00427 0.107
## 4    Glutamic acid, 3TMS; 8       0.241  0.0583  0.4230 0.00974 0.183
## [1] ""
## [1] "Table: log_Blood_TGA"
## [1] "  (from model: "
## [1] "   ~ Ulcer.diagnosis.at.DATE + Age.x + Gender.x +"
## [1] "     Hba1c_baseline + CALSBP + bmi + Smoking + Statin +"
```

```
## [1] "     log_Blood_TGA + Total_cholesterol + egfr + logUAER)"
## [1] ""
##                                 Name Coefficient      CI.L     CI.R  p.value
## 1          Palmitic acid, TMS; 5       0.283  0.151000  0.41400 2.70e-05
## 2              Octanoic acid; 68       0.267  0.135000  0.39900 7.69e-05
## 3            Dodecanoic acid; 54       0.262  0.130000  0.39400 1.02e-04
## 4             Arachidic acid; 46       0.263  0.131000  0.39600 1.04e-04
## 5           Stearic acid, TMS; 2       0.260  0.128000  0.39300 1.18e-04
## 6  2-Hydroxybutyric acid, 2TMS; 2       0.238  0.108000  0.36700 3.41e-04
## 7              Decanoic acid; 52       0.240  0.108000  0.37200 3.67e-04
## 8             Oleic acid, TMS; 3       0.213  0.080700  0.34500 1.62e-03
## 9                 Lactic acid; 29       0.197  0.064600  0.32900 3.57e-03
## 10         Glyceryl-glycoside; 59       0.189  0.057700  0.32100 4.85e-03
## 11           Isoleucine, 2TMS; 18       0.187  0.056900  0.31700 4.86e-03
## 12               Valine, 2TMS; 20       0.185  0.055700  0.31500 5.09e-03
## 13         Heptadecanoic acid; 60       0.181  0.047900  0.31400 7.68e-03
## 14            Arabinopyranose; 51       0.172  0.039800  0.30400 1.08e-02
## 15 4-Hydroxybenzeneacetic acid; 4       0.167  0.037500  0.29600 1.15e-02
## 16           Myristoleic acid; 65       0.164  0.032800  0.29600 1.44e-02
## 17          Aminomalonic acid; 45      -0.163 -0.294000 -0.03130 1.53e-02
## 18               Ribonic acid; 72       0.157  0.029800  0.28500 1.57e-02
## 19 alpha-ketoglutaric acid, TMS M       0.161  0.027800  0.29300 1.78e-02
## 20               Pyruvic acid; 31       0.160  0.027400  0.29300 1.80e-02
## 21 3,4-Dihydroxybutanoic acid; 27       0.153  0.024800  0.28100 1.93e-02
## 22         Glutamic acid, 3TMS; 8       0.155  0.024900  0.28500 1.96e-02
## 23              Leucine, 2TMS; 19       0.155  0.023800  0.28600 2.06e-02
## 24         Heptadecanoic acid; 61       0.156  0.023000  0.28800 2.15e-02
## 25          Fumaric acid, 2TMS; 9       0.154  0.021300  0.28600 2.29e-02
## 26      Arachidonic acid, TMS; 24       0.149  0.016400  0.28200 2.77e-02
## 27            Malic acid, 3TMS; 11       0.140  0.008280  0.27300 3.73e-02
## 28          Nonadecanoic acid; 66       0.138  0.005230  0.27100 4.16e-02
## 29         Succinic acid, 2TMS; 7       0.134  0.001190  0.26700 4.80e-02
## 30     4-Hydroxybutanoic acid; 43       0.134  0.000579  0.26700 4.90e-02
## 31              Glyceric acid; 30      -0.129 -0.259000  0.00136 5.24e-02
## 32            1,3-Propanediol; 34       0.131 -0.002490  0.26400 5.44e-02
## 33              Proline, 2TMS; 21       0.126 -0.005160  0.25800 5.97e-02
## 34                    Ribitol; 71       0.121 -0.005610  0.24800 6.10e-02
## 35     3-Indolepropionic acid; 41      -0.122 -0.254000  0.01040 7.10e-02
## 36              Glycine, 3TMS; 17      -0.115 -0.246000  0.01680 8.73e-02
## 37             Tartronic acid; 73      -0.114 -0.245000  0.01710 8.82e-02
## 38       4-Deoxytetronic acid; 33       0.110 -0.019700  0.24000 9.63e-02
##      p.adj
## 1  0.00177
## 2  0.00177
## 3  0.00177
## 4  0.00177
## 5  0.00177
## 6  0.00394
## 7  0.00394
## 8  0.01510
## 9  0.02970
## 10 0.03180
## 11 0.03180
## 12 0.03180
```

```
## 13 0.04430
## 14 0.05730
## 15 0.05730
## 16 0.06520
## 17 0.06520
## 18 0.06520
## 19 0.06670
## 20 0.06670
## 21 0.06670
## 22 0.06670
## 23 0.06730
## 24 0.06730
## 25 0.06880
## 26 0.07980
## 27 0.10400
## 28 0.11200
## 29 0.12300
## 30 0.12300
## 31 0.12700
## 32 0.12800
## 33 0.13500
## 34 0.13500
## 35 0.15200
## 36 0.17900
## 37 0.17900
## 38 0.19000
## [1] ""
## [1] "Table: Total_cholesterol"
## [1] "  (from model: "
## [1] "   ~ Ulcer.diagnosis.at.DATE + Age.x + Gender.x +"
## [1] "     Hba1c_baseline + CALSBP + bmi + Smoking + Statin +"
## [1] "     log_Blood_TGA + Total_cholesterol + egfr + logUAER)"
## [1] ""
##                               Name Coefficient     CI.L      CI.R  p.value
## 1            Cholesterol, TMS; 23      0.4550  0.36000  0.549000 1.60e-20
## 2                 Campesterol; 49      0.3790  0.28200  0.475000 2.60e-14
## 3            alpha-Tocopherol; 26      0.3290  0.23200  0.425000 3.62e-11
## 4           Linoleic acid, TMS; 4      0.1820  0.08430  0.281000 2.76e-04
## 5          Benzeneacetic acid; 47     -0.1820 -0.28000 -0.084100 2.78e-04
## 6      4-Hydroxybutanoic acid; 43     -0.1720 -0.27100 -0.073200 6.55e-04
## 7            Isoleucine, 2TMS; 18     -0.1620 -0.25800 -0.065400 1.02e-03
## 8              L-5-Oxoproline; 63     -0.1640 -0.26300 -0.065800 1.09e-03
## 9               Proline, 2TMS; 21     -0.1620 -0.26000 -0.064400 1.15e-03
## 10                   Tyrosine; 75     -0.1620 -0.26000 -0.063500 1.27e-03
## 11           Methionine, 2TMS; 16     -0.1560 -0.25300 -0.058200 1.77e-03
## 12      Eicosapentaenoic acid; 55      0.1540  0.05720  0.250000 1.80e-03
## 13       Docosahexaenoic acid; 53      0.1480  0.05120  0.245000 2.76e-03
## 14              Glycine, 3TMS; 17     -0.1470 -0.24500 -0.049800 3.08e-03
## 15            Threonine, 3TMS; 12     -0.1440 -0.24200 -0.045000 4.34e-03
## 16              Alanine, 2TMS; 25     -0.1380 -0.23600 -0.039700 5.96e-03
## 17           Malic acid, 3TMS; 11     -0.1350 -0.23300 -0.037100 6.93e-03
## 18            Arabinopyranose; 51     -0.1320 -0.23000 -0.033600 8.54e-03
## 19 4-Hydroxybenzeneacetic acid; 4     -0.1260 -0.22200 -0.029800 1.02e-02
## 20         Glyceryl-glycoside; 59     -0.1260 -0.22400 -0.028500 1.14e-02
```

```
## 21         2-Palmitoylglycerol; 39      0.1280  0.02880  0.226000 1.14e-02
## 22               Hydroxylamine; 62     -0.1270 -0.22600 -0.027800 1.21e-02
## 23 2,4-Dihydroxybutanoic acid; 28     -0.1180 -0.21200 -0.023300 1.46e-02
## 24                Serine, 3TMS; 14     -0.1150 -0.21300 -0.016400 2.22e-02
## 25         3-Indoleacetic acid; 40     -0.1110 -0.20900 -0.013500 2.57e-02
## 26                     Ribitol; 71     -0.1050 -0.19900 -0.011000 2.86e-02
## 27                Ribonic acid; 72     -0.1030 -0.19800 -0.008590 3.25e-02
## 28         Phenylalanine, 2TMS; 13     -0.0996 -0.19800 -0.000782 4.82e-02
## 29                Ethanolamine; 56     -0.0964 -0.19500  0.002240 5.54e-02
## 30                Pyruvic acid; 31     -0.0955 -0.19400  0.002940 5.72e-02
## 31 3,4-Dihydroxybutanoic acid; 27     -0.0888 -0.18400  0.006250 6.71e-02
## 32           Palmitic acid, TMS; 5      0.0899 -0.00784  0.188000 7.14e-02
## 33               Leucine, 2TMS; 19     -0.0887 -0.18600  0.008620 7.40e-02
## 34     3-Indolepropionic acid; 41      0.0861 -0.01200  0.184000 8.54e-02
## 35 2-Hydroxybutyric acid, 2TMS; 2      0.0830 -0.01340  0.179000 9.14e-02
##       p.adj
## 1  1.20e-18
## 2  9.76e-13
## 3  9.04e-10
## 4  4.17e-03
## 5  4.17e-03
## 6  8.18e-03
## 7  9.52e-03
## 8  9.52e-03
## 9  9.52e-03
## 10 9.52e-03
## 11 1.13e-02
## 12 1.13e-02
## 13 1.59e-02
## 14 1.65e-02
## 15 2.17e-02
## 16 2.79e-02
## 17 3.06e-02
## 18 3.56e-02
## 19 4.04e-02
## 20 4.08e-02
## 21 4.08e-02
## 22 4.11e-02
## 23 4.76e-02
## 24 6.95e-02
## 25 7.71e-02
## 26 8.26e-02
## 27 9.04e-02
## 28 1.29e-01
## 29 1.43e-01
## 30 1.43e-01
## 31 1.62e-01
## 32 1.67e-01
## 33 1.68e-01
## 34 1.88e-01
## 35 1.96e-01
## [1] ""
## [1] "Table: egfr"
## [1] "  (from model: "
```

```
## [1] "   ~ Ulcer.diagnosis.at.DATE + Age.x + Gender.x +"
## [1] "     Hba1c_baseline + CALSBP + bmi + Smoking + Statin +"
## [1] "     log_Blood_TGA + Total_cholesterol + egfr + logUAER)"
## [1] ""
##                               Name Coefficient      CI.L      CI.R  p.value
## 1           Myo inositol 6TMS; 1    -0.01630 -1.94e-02 -0.013100 2.37e-23
## 2                    Ribitol; 71    -0.01520 -1.83e-02 -0.012000 2.04e-20
## 3                 Creatinine; 50    -0.01510 -1.83e-02 -0.011900 1.31e-19
## 4  2,4-Dihydroxybutanoic acid; 28    -0.01480 -1.79e-02 -0.011600 2.20e-19
## 5                Ribonic acid; 72    -0.01350 -1.67e-02 -0.010400 1.51e-16
## 6  3,4-Dihydroxybutanoic acid; 27    -0.01140 -1.46e-02 -0.008180 4.28e-12
## 7        4-Deoxytetronic acid; 33    -0.01130 -1.45e-02 -0.008020 1.47e-11
## 8  4-Hydroxybenzeneacetic acid; 4    -0.01120 -1.44e-02 -0.007960 1.51e-11
## 9           Isoleucine, 2TMS; 18     0.00971  6.47e-03  0.013000 5.35e-09
## 10       4-Deoxytetronic acid; 32    -0.00902 -1.23e-02 -0.005750 7.44e-08
## 11           Citric acid, 4TMS; 6    -0.00886 -1.21e-02 -0.005600 1.11e-07
## 12          Pyroglutamic acid; 69    -0.00893 -1.22e-02 -0.005630 1.20e-07
## 13 2-Hydroxybutyric acid, 2TMS; 2     0.00786  4.62e-03  0.011100 2.14e-06
## 14 4-Hydroxyphenyllactic acid; 44    -0.00781 -1.11e-02 -0.004520 3.51e-06
## 15              Valine, 2TMS; 20     0.00760  4.36e-03  0.010800 4.39e-06
## 16        3-Indoleacetic acid; 40    -0.00751 -1.08e-02 -0.004230 7.60e-06
## 17         Glyceryl-glycoside; 59    -0.00650 -9.79e-03 -0.003210 1.12e-04
## 18             Hydroxyproline; 64    -0.00650 -9.81e-03 -0.003200 1.18e-04
## 19             Leucine, 2TMS; 19     0.00625  2.97e-03  0.009520 1.88e-04
## 20              Serine, 3TMS; 14     0.00618  2.88e-03  0.009490 2.51e-04
## 21             Glycine, 3TMS; 17    -0.00592 -9.20e-03 -0.002640 4.14e-04
## 22          Fumaric acid, 2TMS; 9    -0.00585 -9.16e-03 -0.002550 5.31e-04
## 23      Eicosapentaenoic acid; 55     0.00514  1.90e-03  0.008390 1.90e-03
## 24           Stearic acid, TMS; 2     0.00502  1.72e-03  0.008330 2.91e-03
## 25          Methionine, 2TMS; 16     0.00497  1.69e-03  0.008250 3.04e-03
## 26          Malic acid, 3TMS; 11    -0.00426 -7.56e-03 -0.000953 1.16e-02
## 27         Benzeneacetic acid; 47    -0.00405 -7.35e-03 -0.000749 1.62e-02
## 28          Palmitic acid, TMS; 5     0.00386  5.72e-04  0.007150 2.14e-02
## 29              Octanoic acid; 68     0.00386  5.57e-04  0.007160 2.20e-02
## 30                   Tyrosine; 75     0.00377  4.67e-04  0.007080 2.53e-02
## 31                   Glycerol; 57     0.00369  3.62e-04  0.007020 2.98e-02
## 32          Aminomalonic acid; 45    -0.00360 -6.88e-03 -0.000317 3.16e-02
## 33             Alanine, 2TMS; 25    -0.00353 -6.84e-03 -0.000226 3.63e-02
## 34           Cholesterol, TMS; 23     0.00322  2.88e-05  0.006400 4.80e-02
## 35  2-hydroxy Isovaleric acid; 38     0.00308 -2.27e-04  0.006390 6.79e-02
## 36                   Glycerol; 58    -0.00304 -6.38e-03  0.000292 7.37e-02
## 37        2-Palmitoylglycerol; 39     0.00302 -3.04e-04  0.006340 7.50e-02
## 38                Campesterol; 49    -0.00276 -6.01e-03  0.000494 9.65e-02
## 39         Succinic acid, 2TMS; 7    -0.00280 -6.11e-03  0.000520 9.84e-02
## 40         Glutamic acid, 3TMS; 8     0.00270 -5.53e-04  0.005950 1.04e-01
##       p.adj
## 1  1.78e-21
## 2  7.63e-19
## 3  3.29e-18
## 4  4.12e-18
## 5  2.26e-15
## 6  5.35e-11
## 7  1.41e-10
## 8  1.41e-10
```

```
## 9  4.46e-08
## 10 5.58e-07
## 11 7.52e-07
## 12 7.52e-07
## 13 1.24e-05
## 14 1.88e-05
## 15 2.20e-05
## 16 3.56e-05
## 17 4.93e-04
## 18 4.93e-04
## 19 7.43e-04
## 20 9.40e-04
## 21 1.48e-03
## 22 1.81e-03
## 23 6.20e-03
## 24 9.09e-03
## 25 9.11e-03
## 26 3.34e-02
## 27 4.50e-02
## 28 5.70e-02
## 29 5.70e-02
## 30 6.34e-02
## 31 7.21e-02
## 32 7.41e-02
## 33 8.24e-02
## 34 1.06e-01
## 35 1.46e-01
## 36 1.52e-01
## 37 1.52e-01
## 38 1.89e-01
## 39 1.89e-01
## 40 1.95e-01
## [1] ""
## [1] "Table: logUAER"
## [1] "  (from model: "
## [1] "   ~ Ulcer.diagnosis.at.DATE + Age.x + Gender.x +"
## [1] "     Hba1c_baseline + CALSBP + bmi + Smoking + Statin +"
## [1] "     log_Blood_TGA + Total_cholesterol + egfr + logUAER)"
## [1] ""
##                              Name Coefficient   CI.L  CI.R  p.value  p.adj
## 1 3,4-Dihydroxybutanoic acid; 27      0.0744 0.0344 0.114 0.000272 0.0204
## 2       4-Deoxytetronic acid; 32      0.0644 0.0234 0.105 0.002080 0.0781
```

###### 6.1.4.3 Table with All Metabolites

```
## [1] ""
## [1] "Table: Ulcer.diagnosis.at.DATEJA"
## [1] "  (from model: "
## [1] "   ~ Ulcer.diagnosis.at.DATE + Age.x + Gender.x +"
## [1] "     Hba1c_baseline + CALSBP + bmi + Smoking + Statin +"
## [1] "     log_Blood_TGA + Total_cholesterol + egfr + logUAER)"
## [1] ""
##                               Name Coefficient    CI.L    CI.R p.value p.adj
## 1            Citric acid, 4TMS; 6    -0.52900 -1.0100 -0.0442  0.0325 0.980
## 2  2-Hydroxybutyric acid, 2TMS; 2    -0.45800 -0.9400  0.0235  0.0623 0.980
## 3          Nonadecanoic acid; 66     0.40000 -0.0923  0.8930  0.1110 0.980
## 4                   Tyrosine; 75     0.39400 -0.0973  0.8860  0.1160 0.980
## 5         Benzeneacetic acid; 47    -0.38800 -0.8790  0.1020  0.1210 0.980
## 6               Serine, 3TMS; 14    -0.36400 -0.8550  0.1270  0.1460 0.980
## 7         Glyceryl-glycoside; 59    -0.33500 -0.8240  0.1540  0.1800 0.980
## 8                   Glycerol; 58     0.33500 -0.1610  0.8310  0.1850 0.980
## 9            Cholesterol, TMS; 23    -0.31800 -0.7920  0.1550  0.1880 0.980
## 10 4-Hydroxybenzeneacetic acid; 4    -0.30600 -0.7860  0.1740  0.2110 0.980
## 11 3,4-Dihydroxybutanoic acid; 27    -0.29500 -0.7710  0.1800  0.2230 0.980
## 12 2,4-Dihydroxybutanoic acid; 28    -0.29200 -0.7630  0.1800  0.2250 0.980
## 13        Heptadecanoic acid; 61     0.30100 -0.1920  0.7940  0.2310 0.980
## 14          Palmitic acid, TMS; 5     0.29500 -0.1940  0.7840  0.2370 0.980
## 15           alpha-Tocopherol; 26    -0.27700 -0.7600  0.2070  0.2620 0.980
## 16              Nonanoic acid; 67     0.28300 -0.2120  0.7790  0.2620 0.980
## 17      4-Deoxytetronic acid; 33    -0.27200 -0.7550  0.2110  0.2690 0.980
## 18             L-5-Oxoproline; 63    -0.27400 -0.7670  0.2180  0.2750 0.980
## 19  2-hydroxy Isovaleric acid; 38    -0.26600 -0.7580  0.2260  0.2890 0.980
## 20       3-Indoleacetic acid; 40    -0.23900 -0.7270  0.2490  0.3360 0.980
## 21           Stearic acid, TMS; 2     0.24000 -0.2520  0.7310  0.3390 0.980
## 22           Malic acid, 3TMS; 11     0.22900 -0.2620  0.7200  0.3610 0.980
## 23         Myristoleic acid; 65     0.22200 -0.2670  0.7110  0.3730 0.980
## 24                   Ribitol; 70    -0.22200 -0.7130  0.2690  0.3760 0.980
## 25        Glutamic acid, 3TMS; 8    -0.20800 -0.6910  0.2760  0.3990 0.980
## 26          Tridecanoic acid; 74     0.20700 -0.2860  0.7010  0.4090 0.980
## 27          Methionine, 2TMS; 16     0.19900 -0.2900  0.6870  0.4250 0.980
## 28        Succinic acid, 2TMS; 7     0.19600 -0.2970  0.6890  0.4350 0.980
## 29          Aminomalonic acid; 45    -0.19400 -0.6820  0.2940  0.4350 0.980
## 30                   Ribitol; 71     0.18700 -0.2840  0.6580  0.4370 0.980
## 31          Isoleucine, 2TMS; 18    -0.19100 -0.6740  0.2910  0.4370 0.980
## 32         Fumaric acid, 2TMS; 9    -0.18600 -0.6770  0.3060  0.4580 0.980
## 33               Campesterol; 49    -0.17300 -0.6560  0.3100  0.4830 0.980
## 34           1,3-Propanediol; 34     0.17300 -0.3220  0.6670  0.4940 0.980
## 35        Heptadecanoic acid; 60     0.17200 -0.3210  0.6650  0.4950 0.980
## 36      4-Deoxytetronic acid; 32    -0.15500 -0.6420  0.3320  0.5320 0.980
## 37        11-Eicosenoic acid; 35     0.15000 -0.3440  0.6440  0.5520 0.980
## 38             Proline, 2TMS; 21     0.14800 -0.3400  0.6360  0.5530 0.980
## 39             Glycine, 3TMS; 17    -0.14600 -0.6340  0.3420  0.5570 0.980
## 40              Ethanolamine; 56     0.14600 -0.3470  0.6400  0.5610 0.980
## 41            1-Monopalmitin; 37    -0.14700 -0.6420  0.3490  0.5620 0.980
## 42            Arachidic acid; 46     0.14300 -0.3500  0.6350  0.5700 0.980
## 43               Bisphenol A; 48    -0.13200 -0.6280  0.3640  0.6010 0.980
## 44            Hydroxyproline; 64    -0.12500 -0.6160  0.3670  0.6190 0.980
```

```
## 45          Linoleic acid, TMS; 4    -0.11500 -0.6060  0.3760  0.6450 0.980
## 46              Glyceric acid; 30    -0.11200 -0.5960  0.3720  0.6500 0.980
## 47              Alanine, 2TMS; 25    -0.11300 -0.6040  0.3780  0.6520 0.980
## 48              Decanoic acid; 52     0.10800 -0.3820  0.5970  0.6660 0.980
## 49               Ribonic acid; 72     0.10100 -0.3720  0.5750  0.6750 0.980
## 50              Octanoic acid; 68     0.10400 -0.3870  0.5950  0.6780 0.980
## 51              Hydroxylamine; 62     0.10200 -0.3920  0.5970  0.6850 0.980
## 52     4-Hydroxybutanoic acid; 43    -0.09950 -0.5930  0.3940  0.6930 0.980
## 53        2-Palmitoylglycerol; 39    -0.09410 -0.5880  0.4000  0.7090 0.980
## 54                1-Dodecanol; 36     0.08940 -0.4060  0.5850  0.7230 0.980
## 55 alpha-ketoglutaric acid, TMS M     0.08120 -0.4120  0.5740  0.7470 0.980
## 56          Myo inositol 6TMS; 1    -0.07490 -0.5440  0.3950  0.7540 0.980
## 57      Eicosapentaenoic acid; 55     0.07480 -0.4070  0.5570  0.7610 0.980
## 58 4-Hydroxyphenyllactic acid; 44    -0.07380 -0.5630  0.4160  0.7680 0.980
## 59                Creatinine; 50    -0.07050 -0.5500  0.4090  0.7730 0.980
## 60            Oleic acid, TMS; 3     0.06250 -0.4280  0.5530  0.8030 0.980
## 61            Tartronic acid; 73    -0.05800 -0.5450  0.4290  0.8150 0.980
## 62            Arabinopyranose; 51    -0.05120 -0.5420  0.4400  0.8380 0.980
## 63       Docosahexaenoic acid; 53    -0.04800 -0.5320  0.4360  0.8460 0.980
## 64        Phenylalanine, 2TMS; 13    -0.04120 -0.5360  0.4530  0.8700 0.980
## 65               Valine, 2TMS; 20    -0.03910 -0.5200  0.4420  0.8730 0.980
## 66               Pyruvic acid; 31    -0.03800 -0.5310  0.4550  0.8800 0.980
## 67          Pyroglutamic acid; 69    -0.03640 -0.5260  0.4530  0.8840 0.980
## 68      Arachidonic acid, TMS; 24    -0.03520 -0.5290  0.4580  0.8890 0.980
## 69                Lactic acid; 29     0.03010 -0.4610  0.5220  0.9040 0.983
## 70     3-Indolepropionic acid; 41     0.02100 -0.4700  0.5120  0.9330 0.992
## 71 3-Hydroxybutyric acid, 2TMS; 1    -0.01670 -0.5130  0.4790  0.9470 0.992
## 72            Threonine, 3TMS; 12     0.01040 -0.4830  0.5040  0.9670 0.992
## 73              Leucine, 2TMS; 19     0.00692 -0.4800  0.4940  0.9780 0.992
## 74                   Glycerol; 57    -0.00309 -0.4980  0.4920  0.9900 0.992
## 75             Dodecanoic acid; 54    -0.00240 -0.4930  0.4880  0.9920 0.992
```

#### 6.2 Ulcer Diagnosis from DATE

##### 6.2.1 Crude Model

###### 6.2.1.1 Forest Plot of Model Coefficients

```
## Warning: Ignoring unknown aesthetics: x
```

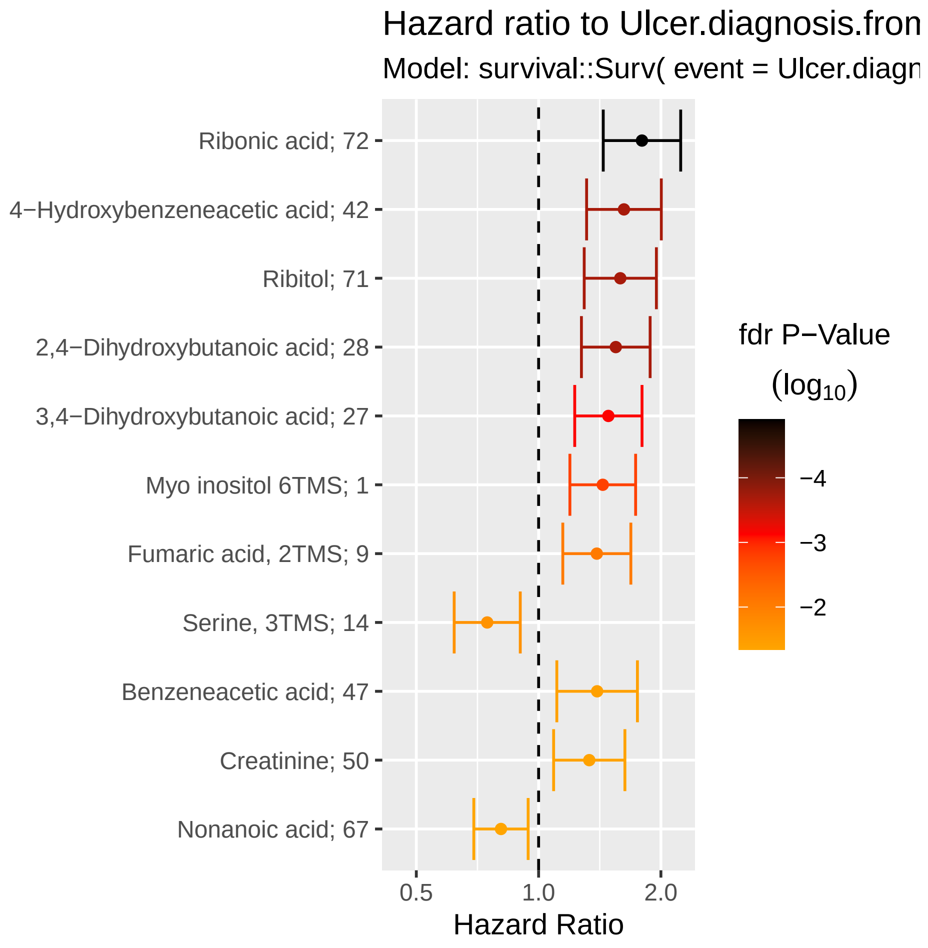

###### 6.2.1.2 Table with All Metabolites

| Name | exp(coef) | Lower 95 % | Upper 95 % | Pr(>\|z\|) | p.adj |
|---|---|---|---|---|---|
| Ribonic acid | 1.8 | 1.44 | 2.24 | 1.69e-07 | 1.27e-05 |
| 4-Hydroxybenzeneacetic acid | 1.62 | 1.31 | 2 | 7.56e-06 | 0.000201 |
| Ribitol (2) | 1.59 | 1.29 | 1.95 | 9.19e-06 | 0.000201 |
| 2,4-Dihydroxybutanoic acid | 1.55 | 1.27 | 1.88 | 1.07e-05 | 0.000201 |
| 3,4-Dihydroxybutanoic acid | 1.48 | 1.23 | 1.8 | 4.85e-05 | 0.000728 |
| Myo inositol | 1.44 | 1.19 | 1.73 | 0.000132 | 0.00164 |
| Fumaric acid | 1.39 | 1.15 | 1.69 | 0.000801 | 0.00858 |
| Serine | 0.747 | 0.62 | 0.901 | 0.00229 | 0.0215 |
| Benzeneacetic acid | 1.39 | 1.11 | 1.75 | 0.00444 | 0.037 |
| Creatinine | 1.33 | 1.09 | 1.63 | 0.00536 | 0.0402 |
| Nonanoic acid | 0.808 | 0.693 | 0.942 | 0.00664 | 0.0453 |
| Tyrosine | 0.834 | 0.728 | 0.956 | 0.00895 | 0.056 |
| 4-Hydroxyphenyllactic acid | 1.29 | 1.04 | 1.61 | 0.0225 | 0.121 |
| Alanine | 1.26 | 1.03 | 1.53 | 0.0227 | 0.121 |
| 3-Indolepropionic acid | 0.861 | 0.753 | 0.983 | 0.0269 | 0.132 |
| 4-Deoxytetronic acid (1) | 1.25 | 1.02 | 1.53 | 0.0284 | 0.132 |
| Malic acid | 1.24 | 1.02 | 1.52 | 0.03 | 0.132 |
| Arabinopyranose | 1.28 | 1.02 | 1.6 | 0.0356 | 0.148 |
| Hydroxyproline | 1.22 | 1.01 | 1.49 | 0.0431 | 0.156 |
| Valine | 0.824 | 0.684 | 0.994 | 0.0432 | 0.156 |
| Threonine | 0.864 | 0.75 | 0.996 | 0.0442 | 0.156 |
| Succinic acid | 1.22 | 1 | 1.47 | 0.0458 | 0.156 |
| Glyceryl-glycoside | 1.25 | 0.995 | 1.58 | 0.0553 | 0.18 |
| Glycine | 1.2 | 0.991 | 1.44 | 0.0626 | 0.196 |
| 1,3-Propanediol | 0.872 | 0.745 | 1.02 | 0.0845 | 0.253 |
| Citric acid | 1.18 | 0.975 | 1.43 | 0.0895 | 0.258 |
| Leucine | 0.867 | 0.732 | 1.03 | 0.099 | 0.275 |
| Nonadecanoic acid | 0.88 | 0.753 | 1.03 | 0.108 | 0.29 |
| Eicosapentaenoic acid | 1.16 | 0.954 | 1.42 | 0.136 | 0.351 |
| 3-Indoleacetic acid | 1.16 | 0.952 | 1.41 | 0.142 | 0.352 |
| 2-Hydroxybutyric acid | 0.871 | 0.722 | 1.05 | 0.146 | 0.352 |
| Pyroglutamic acid | 1.16 | 0.938 | 1.43 | 0.171 | 0.396 |
| Arachidic acid | 0.872 | 0.715 | 1.06 | 0.174 | 0.396 |
| Cholesterol | 0.885 | 0.733 | 1.07 | 0.204 | 0.449 |
| Linoleic acid | 0.906 | 0.774 | 1.06 | 0.224 | 0.473 |
| Pyruvic acid | 1.13 | 0.929 | 1.36 | 0.227 | 0.473 |
| Hydroxylamine | 0.906 | 0.763 | 1.08 | 0.262 | 0.532 |
| Tartronic acid | 0.906 | 0.758 | 1.08 | 0.28 | 0.552 |
| Glycerol (1) | 1.13 | 0.899 | 1.43 | 0.289 | 0.556 |
| 3-Hydroxybutyric acid | 0.904 | 0.747 | 1.09 | 0.3 | 0.562 |
| alpha-ketoglutaric acid | 1.11 | 0.902 | 1.36 | 0.328 | 0.599 |
| Methionine | 0.915 | 0.763 | 1.1 | 0.337 | 0.601 |
| Decanoic acid | 0.918 | 0.768 | 1.1 | 0.347 | 0.604 |
| Glycerol (2) | 1.09 | 0.899 | 1.32 | 0.381 | 0.636 |
| Ethanolamine | 1.09 | 0.899 | 1.32 | 0.382 | 0.636 |
| Phenylalanine | 0.929 | 0.773 | 1.12 | 0.428 | 0.698 |
| Lactic acid | 1.08 | 0.889 | 1.3 | 0.45 | 0.704 |
| Glyceric acid | 0.933 | 0.78 | 1.12 | 0.45 | 0.704 |
| Isoleucine | 0.934 | 0.777 | 1.12 | 0.469 | 0.718 |
| Glutamic acid | 1.07 | 0.877 | 1.29 | 0.524 | 0.776 |

| Name | exp(coef) | Lower 95 % | Upper 95 % | Pr(>\|z\|) | p.adj |
|---|---|---|---|---|---|
| alpha-Tocopherol | 1.06 | 0.876 | 1.29 | 0.528 | 0.776 |
| 4-Deoxytetronic acid (2) | 1.06 | 0.875 | 1.29 | 0.538 | 0.776 |
| Octanoic acid | 0.95 | 0.801 | 1.13 | 0.56 | 0.79 |
| 1-Monopalmitin | 1.06 | 0.868 | 1.29 | 0.576 | 0.79 |
| 2-Palmitoylglycerol | 1.05 | 0.873 | 1.27 | 0.58 | 0.79 |
| Campesterol | 1.05 | 0.866 | 1.27 | 0.626 | 0.838 |
| Oleic acid | 0.96 | 0.795 | 1.16 | 0.67 | 0.882 |
| 1-Dodecanol | 0.964 | 0.801 | 1.16 | 0.696 | 0.883 |
| 2-hydroxy Isovaleric acid | 0.964 | 0.802 | 1.16 | 0.699 | 0.883 |
| Myristoleic acid | 1.04 | 0.858 | 1.25 | 0.706 | 0.883 |
| 11-Eicosenoic acid | 1.03 | 0.854 | 1.25 | 0.734 | 0.903 |
| Stearic acid | 0.973 | 0.803 | 1.18 | 0.779 | 0.943 |
| Docosahexaenoic acid | 0.979 | 0.817 | 1.17 | 0.814 | 0.962 |
| 4-Hydroxybutanoic acid | 1.02 | 0.846 | 1.23 | 0.828 | 0.962 |
| Palmitic acid | 0.98 | 0.809 | 1.19 | 0.833 | 0.962 |
| Aminomalonic acid | 0.988 | 0.82 | 1.19 | 0.899 | 0.997 |
| Heptadecanoic acid (1) | 0.99 | 0.821 | 1.19 | 0.916 | 0.997 |
| L-5-Oxoproline | 1.01 | 0.836 | 1.21 | 0.935 | 0.997 |
| Ribitol (1) | 0.994 | 0.815 | 1.21 | 0.951 | 0.997 |
| Dodecanoic acid | 1.01 | 0.834 | 1.21 | 0.952 | 0.997 |
| Bisphenol A | 0.995 | 0.825 | 1.2 | 0.958 | 0.997 |
| Heptadecanoic acid (2) | 1 | 0.834 | 1.21 | 0.968 | 0.997 |
| Proline | 0.997 | 0.827 | 1.2 | 0.971 | 0.997 |
| Tridecanoic acid | 1 | 0.832 | 1.2 | 0.996 | 0.997 |
| Arachidonic acid | 1 | 0.826 | 1.21 | 0.997 | 0.997 |

###### 6.2.1.3 Top Metabolite

```
## Call:
## survival::coxph(formula = survival::Surv(time = Ulcer.diagnosis.tdiff,
##     event = Ulcer.diagnosis.from.DATE) ~ Ribonic_acid, data = data.km)
##
##   n= 618, number of events= 108
##    (19 observations deleted due to missingness)
##
##                coef exp(coef) se(coef)     z Pr(>|z|)
## Ribonic_acid 0.5957    1.8142   0.1139 5.231 1.69e-07 ***
## ---
## Signif. codes:  0 '***' 0.001 '**' 0.01 '*' 0.05 '.' 0.1 ' ' 1
##
##              exp(coef) exp(-coef) lower .95 upper .95
## Ribonic_acid     1.814     0.5512     1.451     2.268
##
## Concordance= 0.647  (se = 0.026 )
## Likelihood ratio test= 28.99  on 1 df,   p=7e-08
## Wald test            = 27.36  on 1 df,   p=2e-07
## Score (logrank) test = 24.86  on 1 df,   p=6e-07
```

###### 6.2.1.4 Kaplan-Maier Curve with Median Cutpoint

- Top metabolite

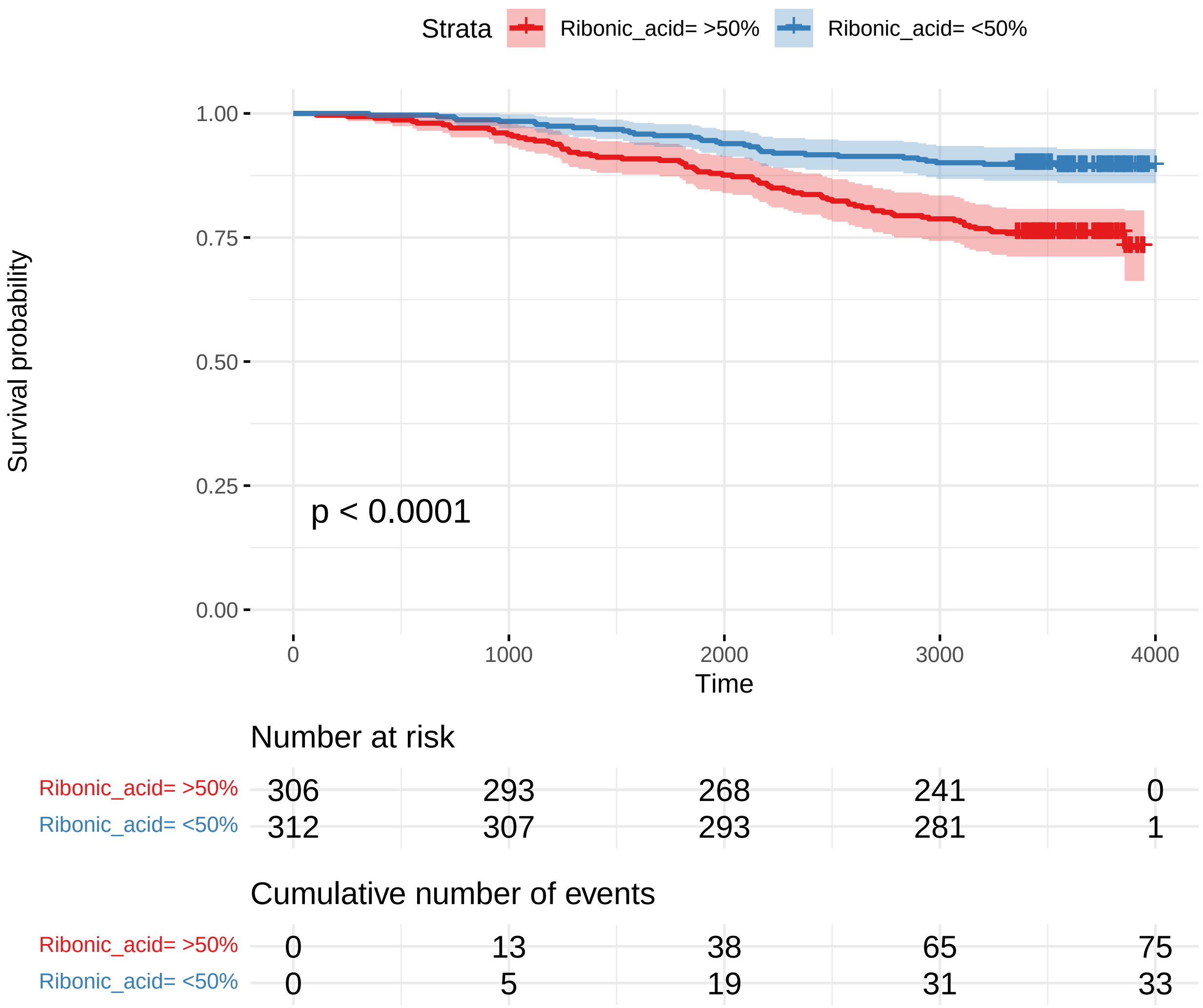

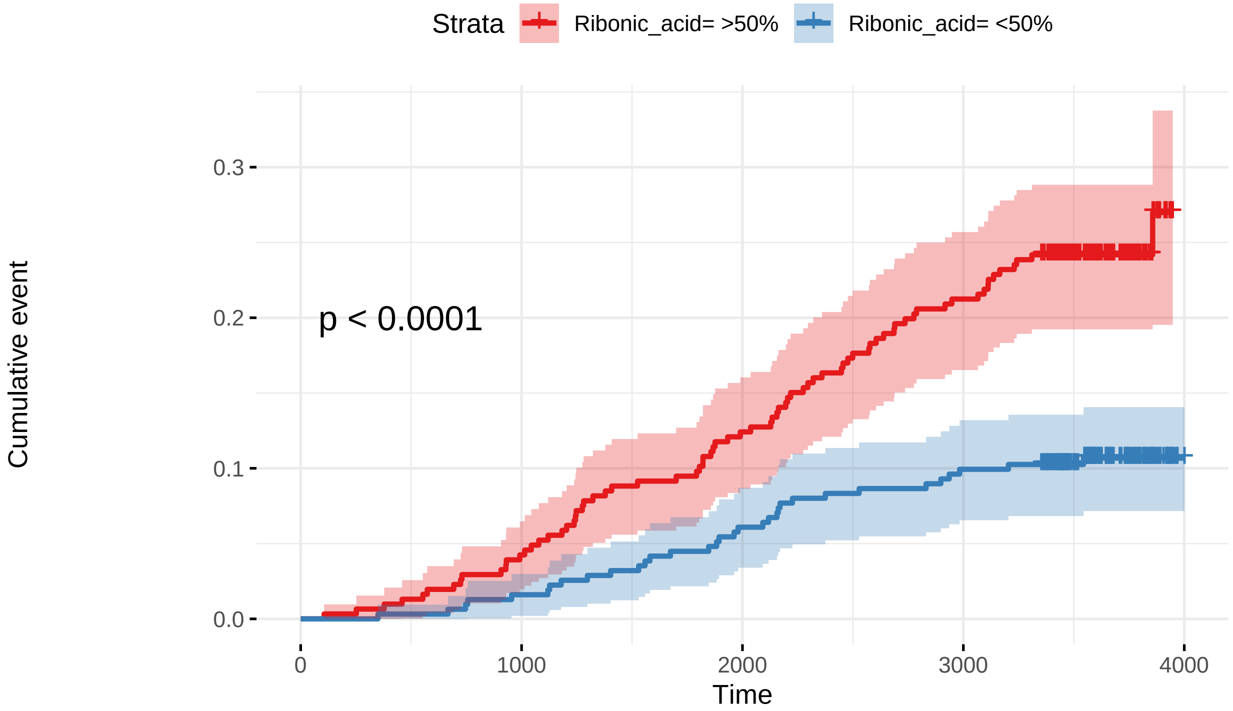

Number at risk

| | 0 | 1000 | 2000 | 3000 | 4000 |
|---|---|---|---|---|---|
| Ribonic_acid= >50% | 306 | 293 | 268 | 241 | 0 |
| Ribonic_acid= <50% | 312 | 307 | 293 | 281 | 1 |

Cumulative number of events

| | 0 | 1000 | 2000 | 3000 | 4000 |
|---|---|---|---|---|---|
| Ribonic_acid= >50% | 0 | 13 | 38 | 65 | 75 |
| Ribonic_acid= <50% | 0 | 5 | 19 | 31 | 33 |

##### 6.2.2 Adusted Model

###### 6.2.2.1 Forest Plot of Model Coefficients

```
## Warning: Ignoring unknown aesthetics: x
```

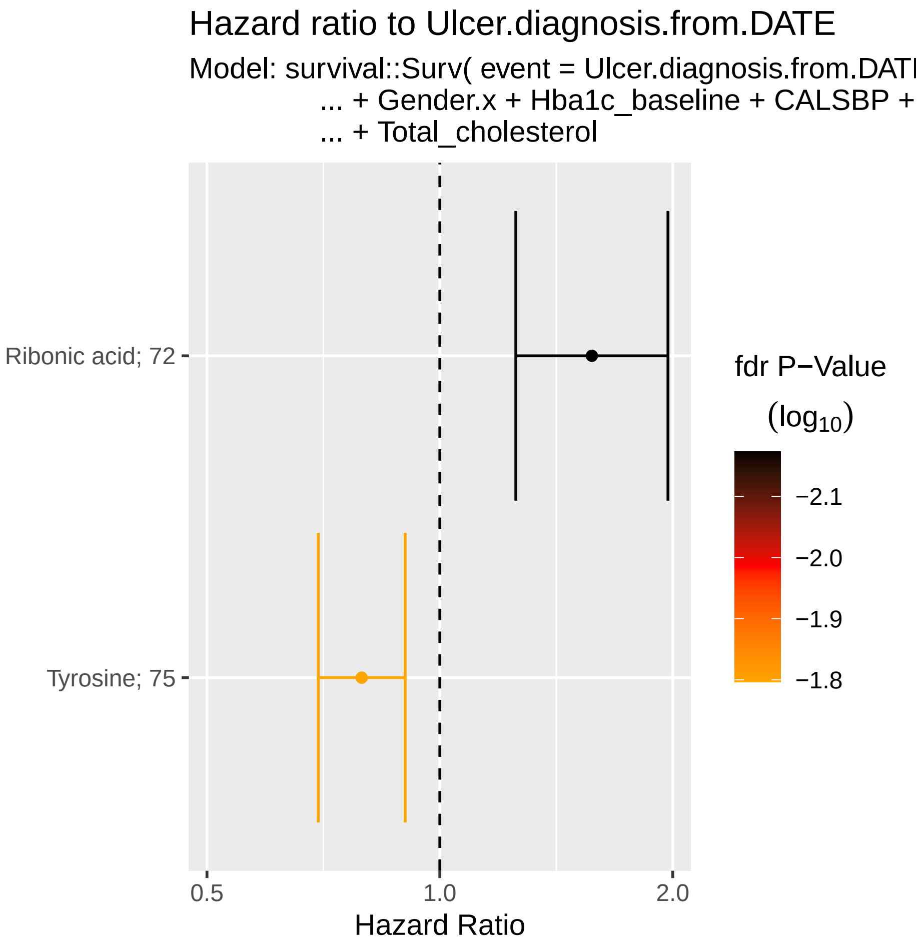

###### 6.2.2.2 Table with All Metabolites

| Name | exp(coef) | Lower 95 % | Upper 95 % | Pr(>\|z\|) | p.adj |
|---|---|---|---|---|---|
| Ribonic acid | 1.57 | 1.25 | 1.97 | 8.95e-05 | 0.00671 |
| Tyrosine | 0.793 | 0.696 | 0.902 | 0.000425 | 0.0159 |
| 2,4-Dihydroxybutanoic acid | 1.35 | 1.09 | 1.67 | 0.00583 | 0.146 |
| Fumaric acid | 1.3 | 1.06 | 1.59 | 0.0104 | 0.157 |
| Valine | 0.773 | 0.631 | 0.946 | 0.0125 | 0.157 |
| Nonanoic acid | 0.829 | 0.715 | 0.963 | 0.014 | 0.157 |
| Ribitol (2) | 1.31 | 1.05 | 1.62 | 0.0172 | 0.157 |
| 4-Hydroxybenzeneacetic acid | 1.32 | 1.05 | 1.65 | 0.0173 | 0.157 |
| 2-Hydroxybutyric acid | 0.791 | 0.648 | 0.966 | 0.0214 | 0.157 |
| Myo inositol | 1.25 | 1.03 | 1.52 | 0.0241 | 0.157 |
| Benzeneacetic acid | 1.29 | 1.03 | 1.63 | 0.0289 | 0.157 |
| Glycine | 1.24 | 1.02 | 1.51 | 0.0295 | 0.157 |
| 4-Deoxytetronic acid (1) | 1.26 | 1.02 | 1.55 | 0.03 | 0.157 |
| 3,4-Dihydroxybutanoic acid | 1.25 | 1.02 | 1.54 | 0.0303 | 0.157 |
| Serine | 0.8 | 0.652 | 0.98 | 0.0314 | 0.157 |
| Leucine | 0.848 | 0.727 | 0.99 | 0.0368 | 0.173 |
| Nonadecanoic acid | 0.831 | 0.696 | 0.992 | 0.0406 | 0.179 |
| Creatinine | 1.23 | 1 | 1.5 | 0.0456 | 0.19 |
| Phenylalanine | 0.834 | 0.693 | 1 | 0.0539 | 0.213 |
| Succinic acid | 1.2 | 0.986 | 1.47 | 0.0689 | 0.259 |
| Proline | 0.842 | 0.686 | 1.03 | 0.102 | 0.349 |
| Malic acid | 1.18 | 0.965 | 1.45 | 0.106 | 0.349 |
| Ribitol (1) | 0.851 | 0.699 | 1.04 | 0.107 | 0.349 |
| Hydroxyproline | 1.18 | 0.958 | 1.45 | 0.12 | 0.374 |
| Arabinopyranose | 1.19 | 0.948 | 1.51 | 0.131 | 0.394 |
| 3-Hydroxybutyric acid | 0.863 | 0.705 | 1.05 | 0.15 | 0.433 |
| Isoleucine | 0.873 | 0.72 | 1.06 | 0.17 | 0.462 |
| 4-Hydroxyphenyllactic acid | 1.18 | 0.932 | 1.48 | 0.172 | 0.462 |
| Threonine | 0.886 | 0.743 | 1.06 | 0.179 | 0.462 |
| Citric acid | 1.15 | 0.935 | 1.41 | 0.187 | 0.469 |
| Alanine | 1.15 | 0.931 | 1.41 | 0.197 | 0.477 |
| Glutamic acid | 0.882 | 0.725 | 1.07 | 0.208 | 0.488 |
| 1,3-Propanediol | 0.909 | 0.779 | 1.06 | 0.229 | 0.52 |
| Methionine | 0.884 | 0.721 | 1.09 | 0.239 | 0.528 |
| Hydroxylamine | 0.912 | 0.764 | 1.09 | 0.303 | 0.637 |
| Decanoic acid | 0.912 | 0.764 | 1.09 | 0.306 | 0.637 |
| Glyceryl-glycoside | 1.12 | 0.885 | 1.42 | 0.34 | 0.685 |
| Pyroglutamic acid | 1.11 | 0.89 | 1.38 | 0.356 | 0.685 |
| 4-Hydroxybutanoic acid | 1.09 | 0.901 | 1.33 | 0.361 | 0.685 |
| Eicosapentaenoic acid | 1.11 | 0.889 | 1.38 | 0.365 | 0.685 |
| Arachidic acid | 0.91 | 0.738 | 1.12 | 0.375 | 0.687 |
| 4-Deoxytetronic acid (2) | 0.92 | 0.75 | 1.13 | 0.426 | 0.758 |
| 3-Indolepropionic acid | 0.936 | 0.792 | 1.11 | 0.435 | 0.758 |
| L-5-Oxoproline | 1.08 | 0.877 | 1.34 | 0.458 | 0.781 |
| Tridecanoic acid | 1.07 | 0.894 | 1.27 | 0.474 | 0.789 |
| 1-Dodecanol | 1.07 | 0.882 | 1.3 | 0.491 | 0.8 |
| Glycerol (2) | 1.07 | 0.875 | 1.31 | 0.509 | 0.812 |
| Tartronic acid | 0.937 | 0.768 | 1.14 | 0.521 | 0.814 |
| 1-Monopalmitin | 1.07 | 0.858 | 1.34 | 0.534 | 0.814 |
| Ethanolamine | 1.06 | 0.87 | 1.29 | 0.56 | 0.814 |

| Name | exp(coef) | Lower 95 % | Upper 95 % | Pr(>\|z\|) | p.adj |
|---|---|---|---|---|---|
| Aminomalonic acid | 1.06 | 0.869 | 1.29 | 0.564 | 0.814 |
| Glycerol (1) | 1.07 | 0.849 | 1.35 | 0.564 | 0.814 |
| Octanoic acid | 0.946 | 0.773 | 1.16 | 0.593 | 0.839 |
| Bisphenol A | 1.05 | 0.866 | 1.27 | 0.616 | 0.84 |
| alpha-Tocopherol | 1.06 | 0.851 | 1.31 | 0.616 | 0.84 |
| Cholesterol | 1.05 | 0.841 | 1.32 | 0.654 | 0.875 |
| 2-Palmitoylglycerol | 1.04 | 0.855 | 1.27 | 0.672 | 0.875 |
| Oleic acid | 0.96 | 0.788 | 1.17 | 0.683 | 0.875 |
| Linoleic acid | 0.966 | 0.808 | 1.15 | 0.703 | 0.875 |
| Campesterol | 1.04 | 0.841 | 1.29 | 0.709 | 0.875 |
| 11-Eicosenoic acid | 1.04 | 0.851 | 1.27 | 0.711 | 0.875 |
| Palmitic acid | 0.967 | 0.796 | 1.17 | 0.733 | 0.883 |
| Heptadecanoic acid (1) | 0.969 | 0.802 | 1.17 | 0.741 | 0.883 |
| Docosahexaenoic acid | 0.97 | 0.799 | 1.18 | 0.759 | 0.889 |
| 3-Indoleacetic acid | 0.97 | 0.784 | 1.2 | 0.776 | 0.893 |
| alpha-ketoglutaric acid | 1.03 | 0.852 | 1.24 | 0.786 | 0.893 |
| Myristoleic acid | 0.976 | 0.795 | 1.2 | 0.813 | 0.903 |
| Dodecanoic acid | 0.977 | 0.8 | 1.19 | 0.819 | 0.903 |
| Arachidonic acid | 0.981 | 0.814 | 1.18 | 0.841 | 0.915 |
| Heptadecanoic acid (2) | 1.02 | 0.839 | 1.23 | 0.867 | 0.929 |
| 2-hydroxy Isovaleric acid | 1.01 | 0.835 | 1.23 | 0.881 | 0.931 |
| Glyceric acid | 1.01 | 0.844 | 1.21 | 0.909 | 0.947 |
| Lactic acid | 1.01 | 0.824 | 1.24 | 0.928 | 0.953 |
| Stearic acid | 1 | 0.821 | 1.22 | 0.982 | 0.995 |
| Pyruvic acid | 0.999 | 0.825 | 1.21 | 0.995 | 0.995 |

###### 6.2.2.3 Top Metabolite

```
## Call:
## survival::coxph(formula = survival::Surv(time = Ulcer.diagnosis.tdiff,
##     event = Ulcer.diagnosis.from.DATE) ~ Ribonic_acid + Age.x +
##     Gender.x + Hba1c_baseline + CALSBP + bmi + Smoking + Statin +
##     log_Blood_TGA + Total_cholesterol, data = data.km)
##
##   n= 605, number of events= 107
##    (32 observations deleted due to missingness)
##
##                        coef  exp(coef)   se(coef)      z Pr(>|z|)
## Ribonic_acid       0.4601710  1.5843449  0.1174692  3.917 8.95e-05 ***
## Age.x              0.0434494  1.0444071  0.0091261  4.761 1.93e-06 ***
## Gender.x           0.3421315  1.4079455  0.2055785  1.664 0.096065 .
## Hba1c_baseline     0.3097510  1.3630857  0.0865588  3.579 0.000346 ***
## CALSBP            -0.0009112  0.9990892  0.0055210 -0.165 0.868913
## bmi                0.0167344  1.0168752  0.0243790  0.686 0.492446
## Smoking            0.3277397  1.3878276  0.2411257  1.359 0.174081
## Statin             0.7673117  2.1539680  0.2623612  2.925 0.003449 **
## log_Blood_TGA      0.0691174  1.0715620  0.1705221  0.405 0.685236
## Total_cholesterol -0.0901358  0.9138071  0.1237872 -0.728 0.466521
## ---
## Signif. codes:  0 '***' 0.001 '**' 0.01 '*' 0.05 '.' 0.1 ' ' 1
##
##                   exp(coef) exp(-coef) lower .95 upper .95
## Ribonic_acid         1.5843     0.6312    1.2585     1.995
## Age.x                1.0444     0.9575    1.0259     1.063
## Gender.x             1.4079     0.7103    0.9410     2.107
## Hba1c_baseline       1.3631     0.7336    1.1504     1.615
## CALSBP               0.9991     1.0009    0.9883     1.010
## bmi                  1.0169     0.9834    0.9694     1.067
## Smoking              1.3878     0.7206    0.8651     2.226
## Statin               2.1540     0.4643    1.2880     3.602
## log_Blood_TGA        1.0716     0.9332    0.7671     1.497
## Total_cholesterol    0.9138     1.0943    0.7169     1.165
##
## Concordance= 0.753  (se = 0.019 )
## Likelihood ratio test= 90.56  on 10 df,   p=4e-15
## Wald test            = 79.09  on 10 df,   p=8e-13
## Score (logrank) test = 81.22  on 10 df,   p=3e-13
```

###### 6.2.2.3.1 Forest Plot with Clinical Variables

- Top metabolite

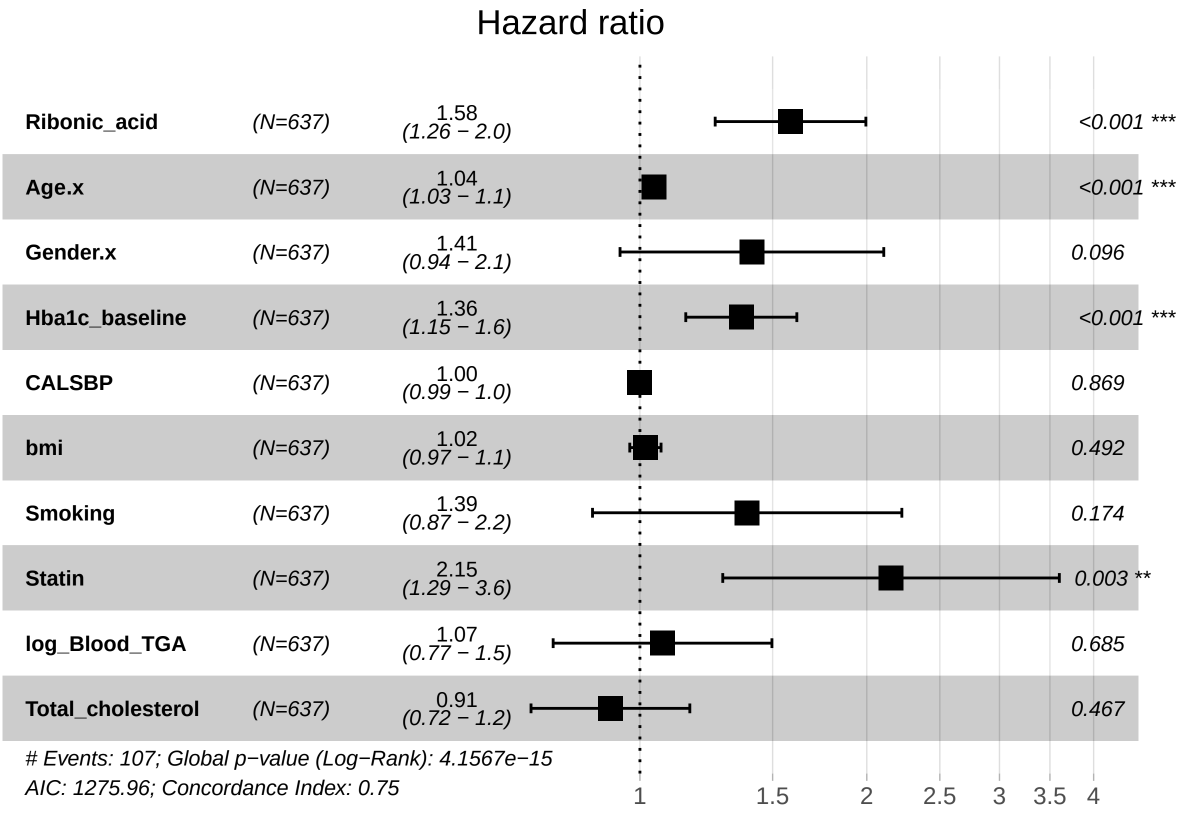

##### 6.2.3 Adjusted Model with eGFR

###### 6.2.3.1 Forest Plot of Model Coefficients

```
## NULL
```

###### 6.2.3.2 Table with All Metabolites

| Name | exp(coef) | Lower 95 % | Upper 95 % | Pr(>\|z\|) | p.adj |
|---|---|---|---|---|---|
| Nonanoic acid | 0.797 | 0.687 | 0.926 | 0.00293 | 0.115 |
| Tyrosine | 0.816 | 0.713 | 0.934 | 0.00308 | 0.115 |
| Ribonic acid | 1.38 | 1.07 | 1.79 | 0.0121 | 0.302 |
| 4-Deoxytetronic acid (2) | 0.779 | 0.635 | 0.955 | 0.0164 | 0.308 |
| Phenylalanine | 0.817 | 0.679 | 0.982 | 0.0316 | 0.395 |
| Fumaric acid | 1.24 | 1.02 | 1.52 | 0.0346 | 0.395 |
| Nonadecanoic acid | 0.831 | 0.698 | 0.989 | 0.0369 | 0.395 |
| Ribitol (1) | 0.834 | 0.684 | 1.02 | 0.0709 | 0.596 |
| Proline | 0.834 | 0.683 | 1.02 | 0.074 | 0.596 |
| Serine | 0.833 | 0.679 | 1.02 | 0.0794 | 0.596 |
| Valine | 0.842 | 0.686 | 1.03 | 0.101 | 0.617 |
| Benzeneacetic acid | 1.21 | 0.961 | 1.51 | 0.105 | 0.617 |
| Glycine | 1.17 | 0.961 | 1.43 | 0.116 | 0.617 |
| 3-Hydroxybutyric acid | 0.853 | 0.699 | 1.04 | 0.12 | 0.617 |
| 1,3-Propanediol | 0.881 | 0.749 | 1.04 | 0.124 | 0.617 |
| Arabinopyranose | 1.2 | 0.947 | 1.52 | 0.132 | 0.617 |
| Succinic acid | 1.17 | 0.951 | 1.43 | 0.14 | 0.618 |
| Eicosapentaenoic acid | 1.18 | 0.941 | 1.48 | 0.152 | 0.634 |
| Leucine | 0.901 | 0.774 | 1.05 | 0.176 | 0.697 |
| 2-Hydroxybutyric acid | 0.872 | 0.709 | 1.07 | 0.193 | 0.725 |
| Threonine | 0.892 | 0.746 | 1.07 | 0.211 | 0.754 |
| 3-Indoleacetic acid | 0.877 | 0.702 | 1.1 | 0.249 | 0.804 |
| 4-Deoxytetronic acid (1) | 1.13 | 0.915 | 1.41 | 0.252 | 0.804 |
| 2,4-Dihydroxybutanoic acid | 1.15 | 0.901 | 1.46 | 0.269 | 0.804 |
| 4-Hydroxybenzeneacetic acid | 1.14 | 0.898 | 1.45 | 0.281 | 0.804 |
| Glutamic acid | 0.899 | 0.74 | 1.09 | 0.282 | 0.804 |
| 4-Hydroxybutanoic acid | 1.11 | 0.914 | 1.35 | 0.292 | 0.804 |
| Decanoic acid | 0.91 | 0.763 | 1.09 | 0.3 | 0.804 |
| Malic acid | 1.11 | 0.905 | 1.36 | 0.318 | 0.821 |
| Glycerol (1) | 1.12 | 0.891 | 1.4 | 0.334 | 0.821 |
| Arachidic acid | 0.908 | 0.741 | 1.11 | 0.351 | 0.821 |
| Hydroxylamine | 0.923 | 0.78 | 1.09 | 0.352 | 0.821 |
| Hydroxyproline | 1.1 | 0.892 | 1.37 | 0.361 | 0.821 |
| Cholesterol | 1.11 | 0.883 | 1.39 | 0.376 | 0.83 |
| L-5-Oxoproline | 1.09 | 0.884 | 1.34 | 0.418 | 0.873 |
| 1-Dodecanol | 1.08 | 0.892 | 1.32 | 0.419 | 0.873 |
| 2-Palmitoylglycerol | 1.08 | 0.885 | 1.32 | 0.446 | 0.9 |
| Alanine | 1.08 | 0.879 | 1.33 | 0.456 | 0.9 |
| 2-hydroxy Isovaleric acid | 1.08 | 0.876 | 1.32 | 0.483 | 0.911 |
| 4-Hydroxyphenyllactic acid | 1.08 | 0.858 | 1.37 | 0.502 | 0.911 |
| Glyceric acid | 1.07 | 0.879 | 1.29 | 0.513 | 0.911 |
| alpha-Tocopherol | 1.07 | 0.867 | 1.32 | 0.529 | 0.911 |
| Ribitol (2) | 1.08 | 0.847 | 1.38 | 0.531 | 0.911 |
| 1-Monopalmitin | 1.07 | 0.86 | 1.34 | 0.536 | 0.911 |
| 3,4-Dihydroxybutanoic acid | 1.07 | 0.854 | 1.35 | 0.548 | 0.911 |
| Tridecanoic acid | 1.05 | 0.884 | 1.25 | 0.572 | 0.911 |
| Ethanolamine | 1.06 | 0.864 | 1.3 | 0.576 | 0.911 |
| Methionine | 0.945 | 0.767 | 1.16 | 0.592 | 0.911 |
| Bisphenol A | 1.05 | 0.868 | 1.28 | 0.595 | 0.911 |
| Tartronic acid | 0.951 | 0.783 | 1.16 | 0.612 | 0.918 |

| Name | exp(coef) | Lower 95 % | Upper 95 % | Pr(>\|z\|) | p.adj |
|---|---|---|---|---|---|
| Creatinine | 1.05 | 0.857 | 1.29 | 0.63 | 0.92 |
| Heptadecanoic acid (1) | 0.962 | 0.801 | 1.16 | 0.681 | 0.92 |
| Isoleucine | 0.961 | 0.788 | 1.17 | 0.692 | 0.92 |
| Citric acid | 1.04 | 0.843 | 1.29 | 0.701 | 0.92 |
| 3-Indolepropionic acid | 0.967 | 0.816 | 1.15 | 0.702 | 0.92 |
| Aminomalonic acid | 1.04 | 0.848 | 1.27 | 0.708 | 0.92 |
| Oleic acid | 0.964 | 0.796 | 1.17 | 0.711 | 0.92 |
| Linoleic acid | 0.968 | 0.813 | 1.15 | 0.712 | 0.92 |
| Dodecanoic acid | 0.966 | 0.792 | 1.18 | 0.729 | 0.92 |
| Stearic acid | 1.03 | 0.849 | 1.26 | 0.736 | 0.92 |
| Myo inositol | 1.03 | 0.818 | 1.3 | 0.791 | 0.943 |
| 11-Eicosenoic acid | 1.03 | 0.843 | 1.25 | 0.792 | 0.943 |
| Glyceryl-glycoside | 1.03 | 0.816 | 1.29 | 0.827 | 0.943 |
| Heptadecanoic acid (2) | 1.02 | 0.843 | 1.24 | 0.832 | 0.943 |
| Octanoic acid | 0.981 | 0.805 | 1.19 | 0.845 | 0.943 |
| Lactic acid | 1.02 | 0.838 | 1.24 | 0.846 | 0.943 |
| Glycerol (2) | 1.02 | 0.835 | 1.24 | 0.851 | 0.943 |
| Myristoleic acid | 0.984 | 0.81 | 1.2 | 0.873 | 0.943 |
| Campesterol | 1.02 | 0.823 | 1.25 | 0.88 | 0.943 |
| alpha-ketoglutaric acid | 1.01 | 0.838 | 1.23 | 0.893 | 0.943 |
| Arachidonic acid | 0.988 | 0.813 | 1.2 | 0.903 | 0.943 |
| Pyroglutamic acid | 0.987 | 0.792 | 1.23 | 0.906 | 0.943 |
| Palmitic acid | 0.996 | 0.814 | 1.22 | 0.973 | 0.995 |
| Docosahexaenoic acid | 1 | 0.821 | 1.22 | 0.982 | 0.995 |
| Pyruvic acid | 1 | 0.827 | 1.21 | 0.995 | 0.995 |

###### 6.2.3.3 Top Metabolite from Adjusted Model

```
## Call:
## survival::coxph(formula = survival::Surv(time = Ulcer.diagnosis.tdiff,
##     event = Ulcer.diagnosis.from.DATE) ~ Ribonic_acid + Age.x +
##     Gender.x + Hba1c_baseline + CALSBP + bmi + Smoking + Statin +
##     log_Blood_TGA + Total_cholesterol + egfr, data = data.km)
##
##   n= 603, number of events= 107
##    (34 observations deleted due to missingness)
##
##                        coef exp(coef)  se(coef)      z Pr(>|z|)
## Ribonic_acid       0.331041  1.392417  0.131879  2.510 0.012067 *
## Age.x              0.040851  1.041696  0.009169  4.455 8.38e-06 ***
## Gender.x           0.407541  1.503116  0.207463  1.964 0.049484 *
## Hba1c_baseline     0.299112  1.348661  0.087588  3.415 0.000638 ***
## CALSBP            -0.001878  0.998124  0.005517 -0.340 0.733554
## bmi                0.016694  1.016835  0.024309  0.687 0.492229
## Smoking            0.342762  1.408834  0.241142  1.421 0.155196
## Statin             0.710802  2.035622  0.264929  2.683 0.007297 **
## log_Blood_TGA      0.042170  1.043072  0.170503  0.247 0.804654
## Total_cholesterol -0.076283  0.926554  0.122820 -0.621 0.534535
## egfr              -0.008234  0.991800  0.004232 -1.946 0.051697 .
## ---
## Signif. codes:  0 '***' 0.001 '**' 0.01 '*' 0.05 '.' 0.1 ' ' 1
##
##                   exp(coef) exp(-coef) lower .95 upper .95
## Ribonic_acid         1.3924     0.7182    1.0753     1.803
## Age.x                1.0417     0.9600    1.0231     1.061
## Gender.x             1.5031     0.6653    1.0009     2.257
## Hba1c_baseline       1.3487     0.7415    1.1359     1.601
## CALSBP               0.9981     1.0019    0.9874     1.009
## bmi                  1.0168     0.9834    0.9695     1.066
## Smoking              1.4088     0.7098    0.8782     2.260
## Statin               2.0356     0.4913    1.2111     3.421
## log_Blood_TGA        1.0431     0.9587    0.7468     1.457
## Total_cholesterol    0.9266     1.0793    0.7283     1.179
## egfr                 0.9918     1.0083    0.9836     1.000
##
## Concordance= 0.761  (se = 0.019 )
## Likelihood ratio test= 94.16  on 11 df,   p=3e-15
## Wald test            = 82.66  on 11 df,   p=5e-13
## Score (logrank) test = 88.44  on 11 df,   p=3e-14
```

###### 6.2.3.3.1 Forest Plot with Clinical Variables

- Top metabolite from adjusted model

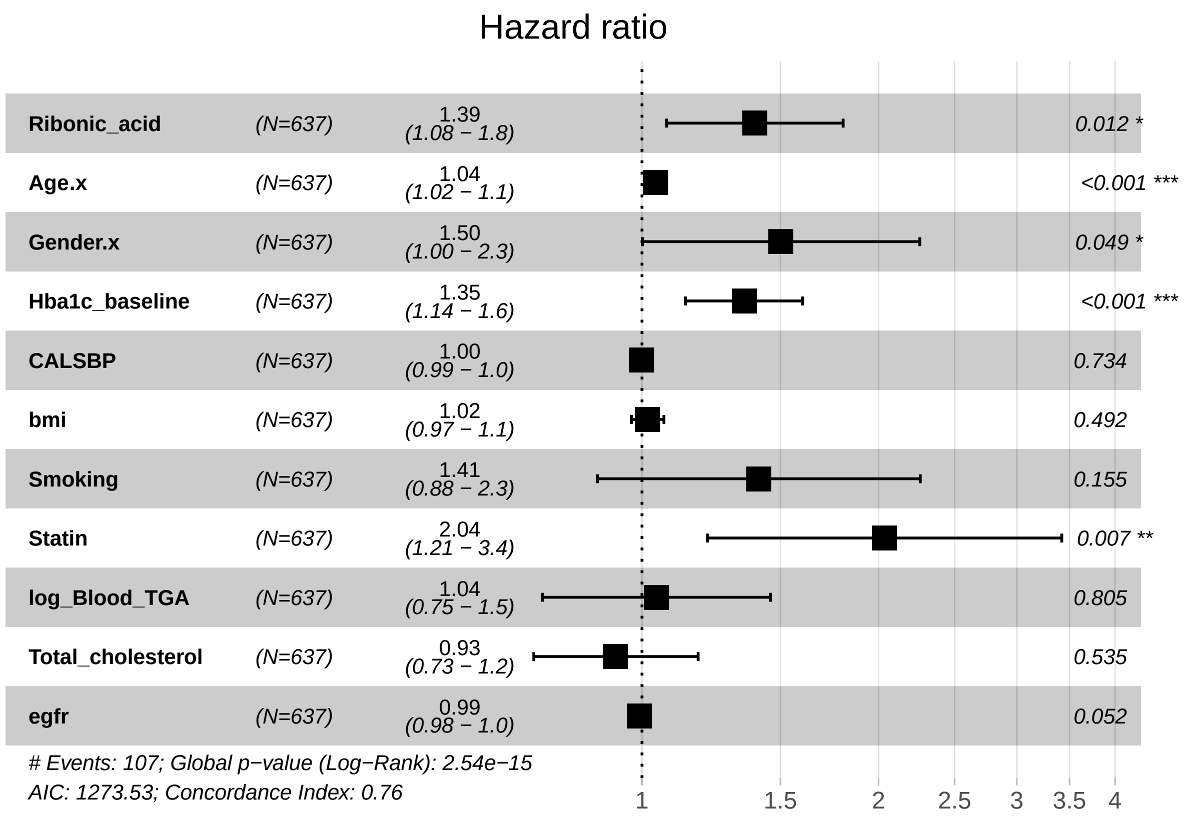

##### 6.2.4 Fully Adjusted Model

###### 6.2.4.1 Forest Plot of Model Coefficients

```
## NULL
```

###### 6.2.4.2 Table with All Metabolites

| Name | exp(coef) | Lower 95 % | Upper 95 % | Pr(>\|z\|) | p.adj |
|---|---|---|---|---|---|
| Tyrosine | 0.81 | 0.707 | 0.928 | 0.00233 | 0.12 |
| Nonanoic acid | 0.798 | 0.687 | 0.927 | 0.00321 | 0.12 |
| Ribonic acid | 1.37 | 1.06 | 1.77 | 0.0154 | 0.385 |
| Nonadecanoic acid | 0.816 | 0.685 | 0.971 | 0.022 | 0.394 |
| 4-Deoxytetronic acid (2) | 0.791 | 0.643 | 0.973 | 0.0263 | 0.394 |
| Phenylalanine | 0.814 | 0.675 | 0.982 | 0.0315 | 0.394 |
| Ribitol (1) | 0.813 | 0.666 | 0.991 | 0.0405 | 0.434 |
| Fumaric acid | 1.21 | 0.989 | 1.48 | 0.0641 | 0.543 |
| Valine | 0.821 | 0.666 | 1.01 | 0.0651 | 0.543 |
| Proline | 0.834 | 0.682 | 1.02 | 0.0755 | 0.547 |
| Serine | 0.832 | 0.677 | 1.02 | 0.0802 | 0.547 |
| 3-Hydroxybutyric acid | 0.84 | 0.687 | 1.03 | 0.0892 | 0.549 |
| Succinic acid | 1.19 | 0.97 | 1.46 | 0.0952 | 0.549 |
| 1,3-Propanediol | 0.882 | 0.75 | 1.04 | 0.131 | 0.69 |
| Benzeneacetic acid | 1.19 | 0.946 | 1.5 | 0.138 | 0.69 |
| Arabinopyranose | 1.18 | 0.932 | 1.5 | 0.168 | 0.721 |
| Leucine | 0.901 | 0.775 | 1.05 | 0.176 | 0.721 |
| Glycine | 1.15 | 0.938 | 1.4 | 0.181 | 0.721 |
| 3-Indoleacetic acid | 0.859 | 0.685 | 1.08 | 0.189 | 0.721 |
| Malic acid | 1.15 | 0.932 | 1.42 | 0.192 | 0.721 |
| Decanoic acid | 0.897 | 0.752 | 1.07 | 0.227 | 0.811 |
| Threonine | 0.9 | 0.751 | 1.08 | 0.253 | 0.812 |
| Arachidic acid | 0.891 | 0.724 | 1.1 | 0.276 | 0.812 |
| Glycerol (1) | 1.13 | 0.899 | 1.43 | 0.291 | 0.812 |
| 2-Hydroxybutyric acid | 0.893 | 0.723 | 1.1 | 0.294 | 0.812 |
| 4-Hydroxybutanoic acid | 1.11 | 0.91 | 1.35 | 0.303 | 0.812 |
| Eicosapentaenoic acid | 1.13 | 0.896 | 1.42 | 0.306 | 0.812 |
| Glutamic acid | 0.903 | 0.74 | 1.1 | 0.311 | 0.812 |
| 4-Deoxytetronic acid (1) | 1.12 | 0.897 | 1.39 | 0.324 | 0.812 |
| Hydroxyproline | 1.11 | 0.898 | 1.38 | 0.325 | 0.812 |
| Hydroxylamine | 0.923 | 0.777 | 1.1 | 0.361 | 0.845 |
| Cholesterol | 1.11 | 0.881 | 1.39 | 0.381 | 0.845 |
| Alanine | 1.09 | 0.888 | 1.35 | 0.398 | 0.845 |
| 4-Hydroxybenzeneacetic acid | 1.11 | 0.874 | 1.4 | 0.401 | 0.845 |
| L-5-Oxoproline | 1.09 | 0.886 | 1.35 | 0.404 | 0.845 |
| 2,4-Dihydroxybutanoic acid | 1.11 | 0.868 | 1.42 | 0.406 | 0.845 |
| 4-Hydroxyphenyllactic acid | 1.09 | 0.854 | 1.38 | 0.501 | 0.891 |
| 2-Palmitoylglycerol | 1.07 | 0.876 | 1.31 | 0.508 | 0.891 |
| 1-Dodecanol | 1.07 | 0.877 | 1.3 | 0.516 | 0.891 |
| Ethanolamine | 1.07 | 0.868 | 1.31 | 0.535 | 0.891 |
| 2-hydroxy Isovaleric acid | 1.07 | 0.866 | 1.32 | 0.538 | 0.891 |
| Tartronic acid | 0.941 | 0.774 | 1.14 | 0.542 | 0.891 |
| Tridecanoic acid | 1.05 | 0.884 | 1.25 | 0.564 | 0.891 |
| 1-Monopalmitin | 1.07 | 0.857 | 1.33 | 0.568 | 0.891 |
| Heptadecanoic acid (1) | 0.948 | 0.787 | 1.14 | 0.573 | 0.891 |
| Methionine | 0.947 | 0.767 | 1.17 | 0.609 | 0.891 |
| 3-Indolepropionic acid | 0.957 | 0.809 | 1.13 | 0.61 | 0.891 |
| Isoleucine | 0.95 | 0.781 | 1.16 | 0.611 | 0.891 |
| 3,4-Dihydroxybutanoic acid | 1.06 | 0.841 | 1.34 | 0.614 | 0.891 |
| Dodecanoic acid | 0.951 | 0.78 | 1.16 | 0.619 | 0.891 |

| Name | exp(coef) | Lower 95 % | Upper 95 % | Pr(>\|z\|) | p.adj |
|---|---|---|---|---|---|
| Creatinine | 1.05 | 0.857 | 1.29 | 0.62 | 0.891 |
| Glyceric acid | 1.05 | 0.864 | 1.28 | 0.62 | 0.891 |
| alpha-Tocopherol | 1.05 | 0.852 | 1.3 | 0.629 | 0.891 |
| Lactic acid | 1.04 | 0.856 | 1.27 | 0.681 | 0.946 |
| Oleic acid | 0.965 | 0.795 | 1.17 | 0.714 | 0.951 |
| Ribitol (2) | 1.05 | 0.82 | 1.33 | 0.719 | 0.951 |
| Bisphenol A | 1.04 | 0.851 | 1.26 | 0.727 | 0.951 |
| Linoleic acid | 0.972 | 0.815 | 1.16 | 0.747 | 0.951 |
| alpha-ketoglutaric acid | 1.03 | 0.847 | 1.25 | 0.768 | 0.951 |
| Stearic acid | 1.03 | 0.843 | 1.25 | 0.783 | 0.951 |
| Octanoic acid | 0.973 | 0.801 | 1.18 | 0.785 | 0.951 |
| Aminomalonic acid | 1.03 | 0.837 | 1.27 | 0.786 | 0.951 |
| Myristoleic acid | 0.976 | 0.804 | 1.18 | 0.805 | 0.958 |
| Glyceryl-glycoside | 0.978 | 0.778 | 1.23 | 0.845 | 0.96 |
| Citric acid | 1.02 | 0.824 | 1.27 | 0.851 | 0.96 |
| Pyroglutamic acid | 0.98 | 0.786 | 1.22 | 0.858 | 0.96 |
| Glycerol (2) | 1.02 | 0.831 | 1.24 | 0.88 | 0.96 |
| Palmitic acid | 0.986 | 0.804 | 1.21 | 0.896 | 0.96 |
| Heptadecanoic acid (2) | 0.989 | 0.816 | 1.2 | 0.913 | 0.96 |
| Docosahexaenoic acid | 0.99 | 0.813 | 1.21 | 0.923 | 0.96 |
| Myo inositol | 1.01 | 0.8 | 1.28 | 0.932 | 0.96 |
| 11-Eicosenoic acid | 0.991 | 0.801 | 1.23 | 0.935 | 0.96 |
| Campesterol | 1.01 | 0.813 | 1.25 | 0.944 | 0.96 |
| Pyruvic acid | 1.01 | 0.829 | 1.22 | 0.959 | 0.96 |
| Arachidonic acid | 0.995 | 0.816 | 1.21 | 0.96 | 0.96 |

###### 6.2.4.3 Top Metabolites

###### 6.2.4.3.1 Top Metabolite – Tyrosine

```
## Call:
## survival::coxph(formula = survival::Surv(time = Ulcer.diagnosis.tdiff,
##     event = Ulcer.diagnosis.from.DATE) ~ Tyrosine + Age.x + Gender.x +
##     Hba1c_baseline + CALSBP + bmi + Smoking + Statin + log_Blood_TGA +
##     Total_cholesterol + egfr + logUAER, data = data.km)
##
##   n= 570, number of events= 105
##    (67 observations deleted due to missingness)
##
##                        coef exp(coef)  se(coef)      z Pr(>|z|)
## Tyrosine          -0.211095  0.809697  0.069329 -3.045  0.00233 **
## Age.x              0.532079  1.702469  0.125699  4.233 2.31e-05 ***
## Gender.x           0.391308  1.478913  0.215163  1.819  0.06896 .
## Hba1c_baseline     0.392627  1.480866  0.100879  3.892 9.94e-05 ***
## CALSBP            -0.042728  0.958172  0.105902 -0.403  0.68660
## bmi                0.003430  1.003436  0.101722  0.034  0.97310
## Smoking            0.250030  1.284064  0.246942  1.013  0.31130
## Statin             0.620318  1.859519  0.267533  2.319  0.02041 *
## log_Blood_TGA      0.061086  1.062991  0.119191  0.513  0.60830
## Total_cholesterol -0.137230  0.871770  0.109713 -1.251  0.21101
## egfr              -0.362850  0.695691  0.119620 -3.033  0.00242 **
## logUAER           -0.007858  0.992173  0.110637 -0.071  0.94338
## ---
## Signif. codes:  0 '***' 0.001 '**' 0.01 '*' 0.05 '.' 0.1 ' ' 1
##
##                   exp(coef) exp(-coef) lower .95 upper .95
## Tyrosine             0.8097     1.2350    0.7068    0.9275
## Age.x                1.7025     0.5874    1.3307    2.1781
## Gender.x             1.4789     0.6762    0.9701    2.2547
## Hba1c_baseline       1.4809     0.6753    1.2152    1.8046
## CALSBP               0.9582     1.0437    0.7786    1.1792
## bmi                  1.0034     0.9966    0.8221    1.2248
## Smoking              1.2841     0.7788    0.7914    2.0835
## Statin               1.8595     0.5378    1.1007    3.1414
## log_Blood_TGA        1.0630     0.9407    0.8415    1.3427
## Total_cholesterol    0.8718     1.1471    0.7031    1.0809
## egfr                 0.6957     1.4374    0.5503    0.8795
## logUAER              0.9922     1.0079    0.7988    1.2324
##
## Concordance= 0.753  (se = 0.019 )
## Likelihood ratio test= 87.75  on 12 df,   p=1e-13
## Wald test            = 79.74  on 12 df,   p=5e-12
## Score (logrank) test = 85.52  on 12 df,   p=4e-13
```

Forest Plot with Clinical Variables

- Tyrosine

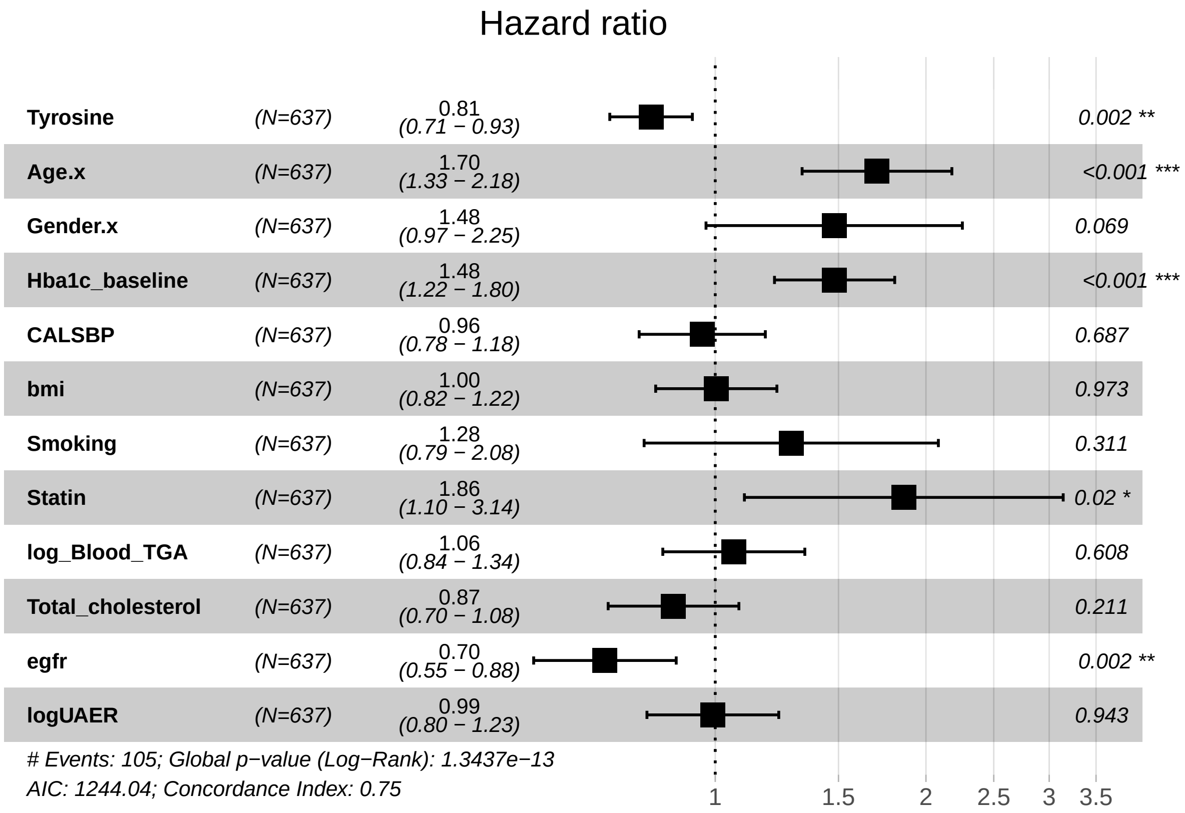

Kaplan-Maier Curve with Median Cutpoint

- Tyrosine

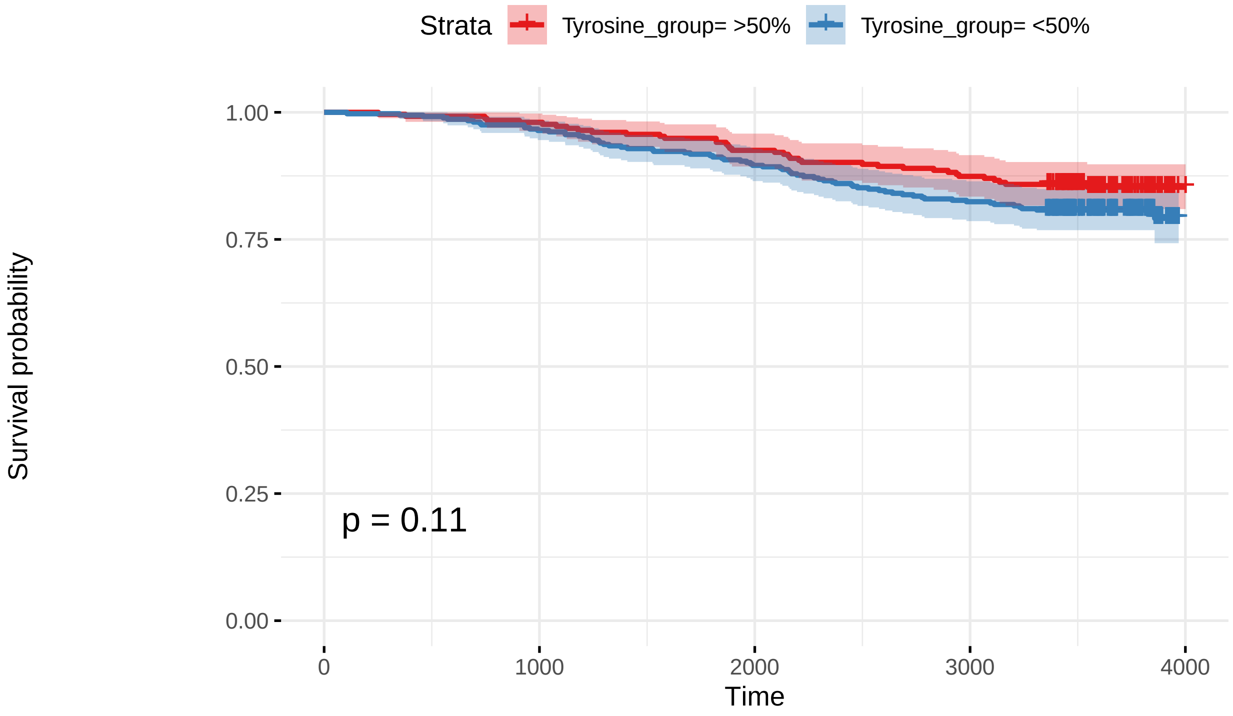

Number at risk

| | 0 | 1000 | 2000 | 3000 | 4000 |
|---|---|---|---|---|---|
| Tyrosine_group= >50% | 254 | 249 | 235 | 222 | 1 |
| Tyrosine_group= <50% | 364 | 351 | 326 | 300 | 0 |

Cumulative number of events

| | 0 | 1000 | 2000 | 3000 | 4000 |
|---|---|---|---|---|---|
| Tyrosine_group= >50% | 0 | 5 | 19 | 32 | 37 |
| Tyrosine_group= <50% | 0 | 13 | 38 | 64 | 71 |

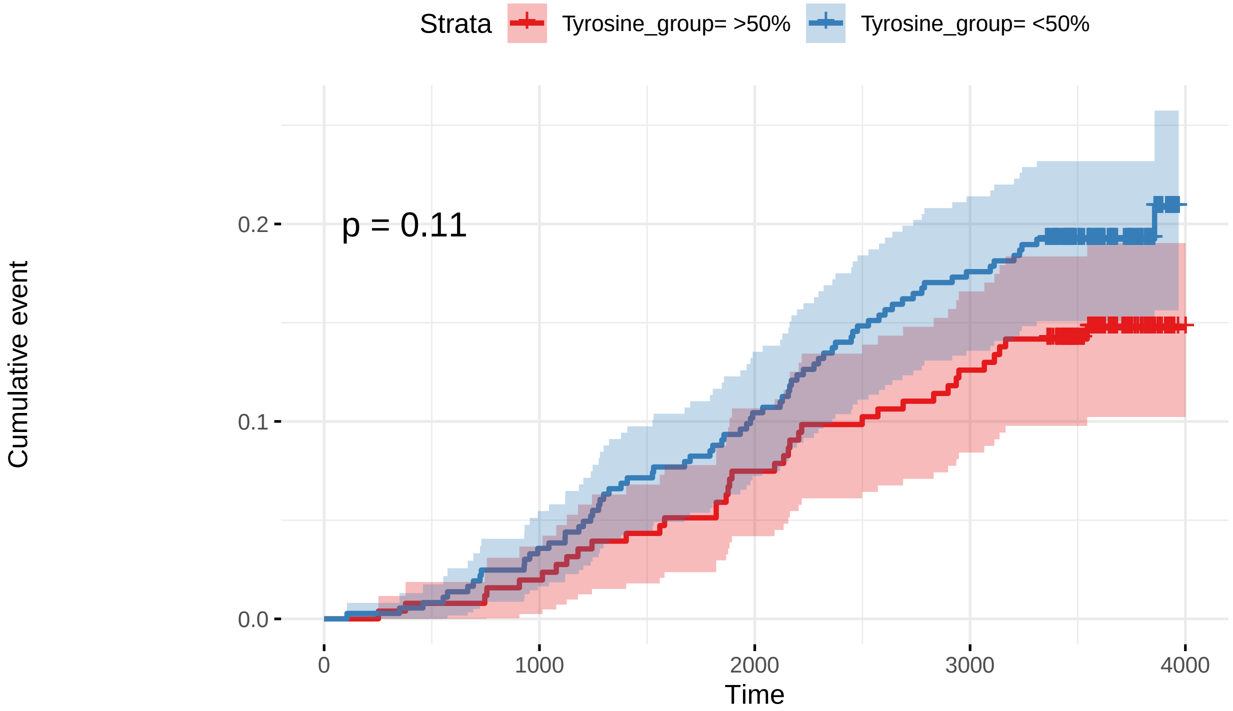

Number at risk

| | 0 | 1000 | 2000 | 3000 | 4000 |
|---|---|---|---|---|---|
| Tyrosine_group= >50% | 254 | 249 | 235 | 222 | 1 |
| Tyrosine_group= <50% | 364 | 351 | 326 | 300 | 0 |

Cumulative number of events

| | 0 | 1000 | 2000 | 3000 | 4000 |
|---|---|---|---|---|---|
| Tyrosine_group= >50% | 0 | 5 | 19 | 32 | 37 |
| Tyrosine_group= <50% | 0 | 13 | 38 | 64 | 71 |

```
##
##   1
## 517
```

```
##
## (-9.21,-0.29] (-0.29,0.155] (0.155,0.541]  (0.541,1.57]
##           159           210           108           159
```

```
##
## 4th Quartile 3rd Quartile 2nd Quartile 1st Quartile
##          159          108          210          159
```

```
## Loading required package: ggplot2
```

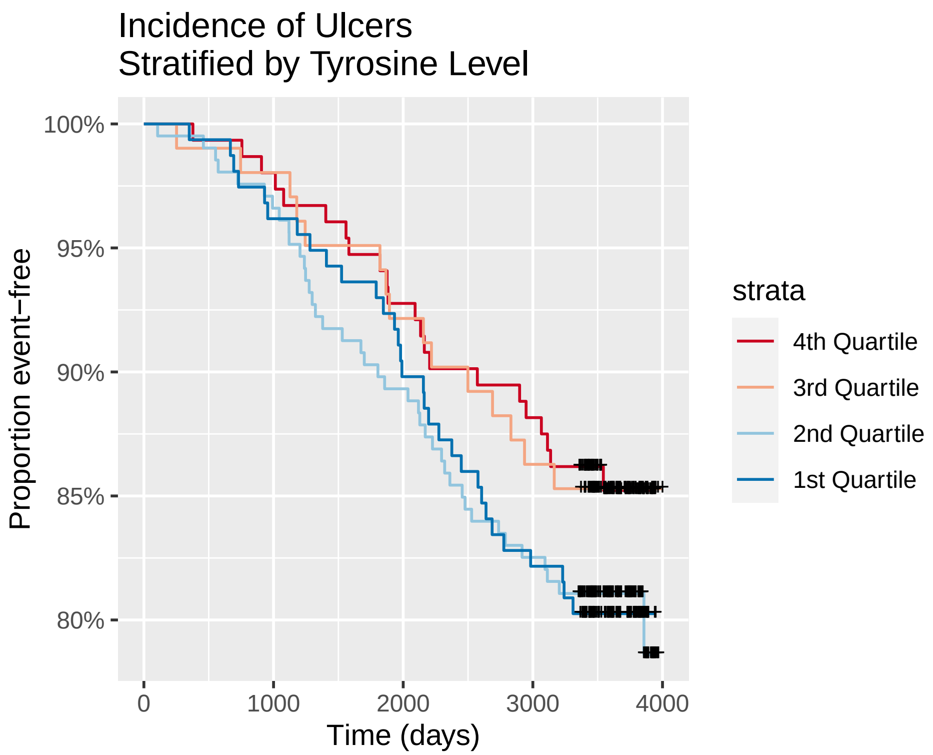

###### 6.2.4.3.2 Top Metabolite from Adjusted Model – Ribonic Acid

```
## Call:
## survival::coxph(formula = survival::Surv(time = Ulcer.diagnosis.tdiff,
##     event = Ulcer.diagnosis.from.DATE) ~ Ribonic_acid + Age.x +
##     Gender.x + Hba1c_baseline + CALSBP + bmi + Smoking + Statin +
##     log_Blood_TGA + Total_cholesterol + egfr + logUAER, data = data.km)
##
##   n= 570, number of events= 105
##    (67 observations deleted due to missingness)
##
##                       coef exp(coef) se(coef)      z Pr(>|z|)
## Ribonic_acid       0.31444   1.36949  0.12977  2.423 0.015392 *
## Age.x              0.46314   1.58905  0.12302  3.765 0.000167 ***
## Gender.x           0.45362   1.57400  0.21598  2.100 0.035701 *
## Hba1c_baseline     0.37866   1.46033  0.10263  3.690 0.000224 ***
## CALSBP            -0.03533   0.96529  0.10325 -0.342 0.732234
## bmi                0.04687   1.04799  0.10025  0.468 0.640089
## Smoking            0.32928   1.38997  0.24962  1.319 0.187116
## Statin             0.65246   1.92026  0.26774  2.437 0.014811 *
## log_Blood_TGA      0.01055   1.01061  0.12009  0.088 0.929969
## Total_cholesterol -0.05763   0.94400  0.10868 -0.530 0.595947
## egfr              -0.24726   0.78094  0.13022 -1.899 0.057587 .
## logUAER           -0.01014   0.98992  0.11117 -0.091 0.927351
## ---
## Signif. codes:  0 '***' 0.001 '**' 0.01 '*' 0.05 '.' 0.1 ' ' 1
##
##                   exp(coef) exp(-coef) lower .95 upper .95
## Ribonic_acid         1.3695     0.7302    1.0619     1.766
## Age.x                1.5891     0.6293    1.2486     2.022
## Gender.x             1.5740     0.6353    1.0308     2.403
## Hba1c_baseline       1.4603     0.6848    1.1943     1.786
## CALSBP               0.9653     1.0360    0.7884     1.182
## bmi                  1.0480     0.9542    0.8610     1.276
## Smoking              1.3900     0.7194    0.8522     2.267
## Statin               1.9203     0.5208    1.1362     3.245
## log_Blood_TGA        1.0106     0.9895    0.7987     1.279
## Total_cholesterol    0.9440     1.0593    0.7629     1.168
## egfr                 0.7809     1.2805    0.6050     1.008
## logUAER              0.9899     1.0102    0.7961     1.231
##
## Concordance= 0.755  (se = 0.02 )
## Likelihood ratio test= 86.86  on 12 df,   p=2e-13
## Wald test            = 78.01  on 12 df,   p=1e-11
## Score (logrank) test = 82.79  on 12 df,   p=1e-12
```

Forest Plot with Clinical Variables

- Top metabolite from adjusted model

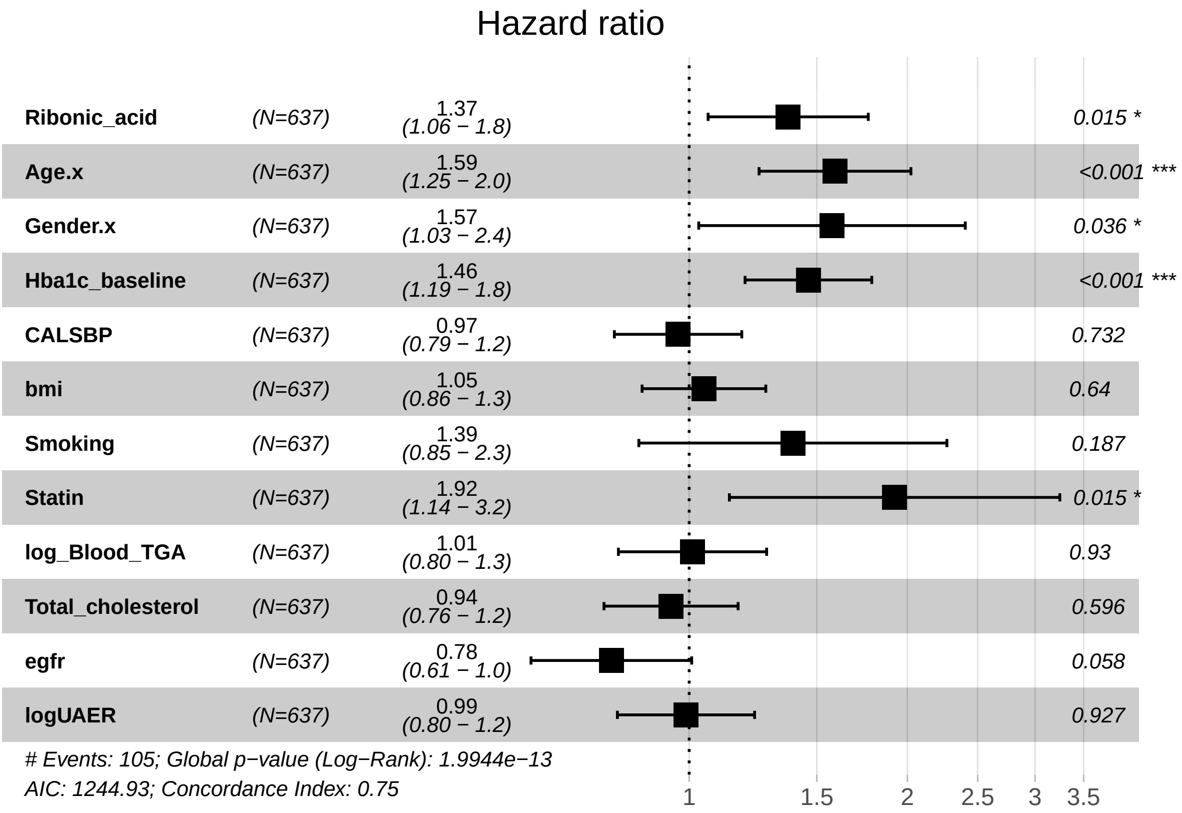

##### 6.2.5 Compilation

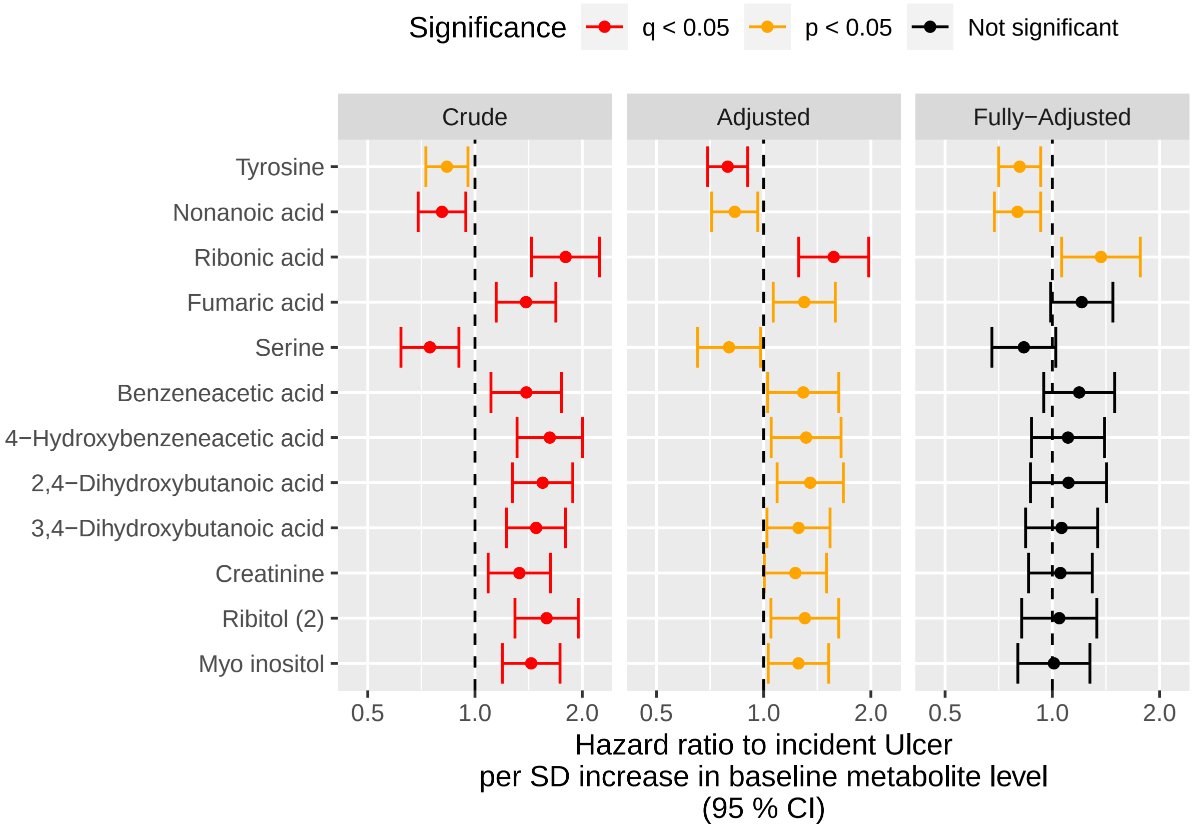

### 7 Integration of Results

#### 7.1 Crude Models

##### 7.1.1 Cross-Sectional

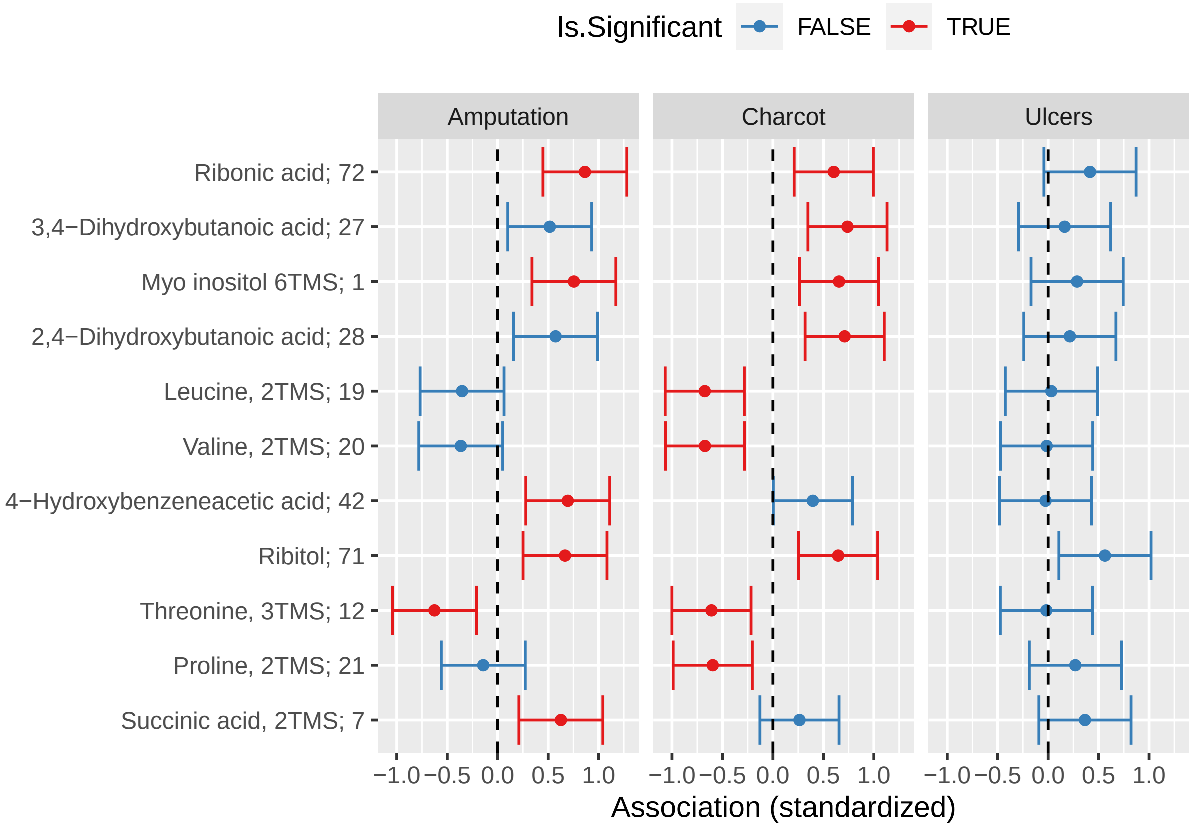

##### 7.1.2 Longitudinal

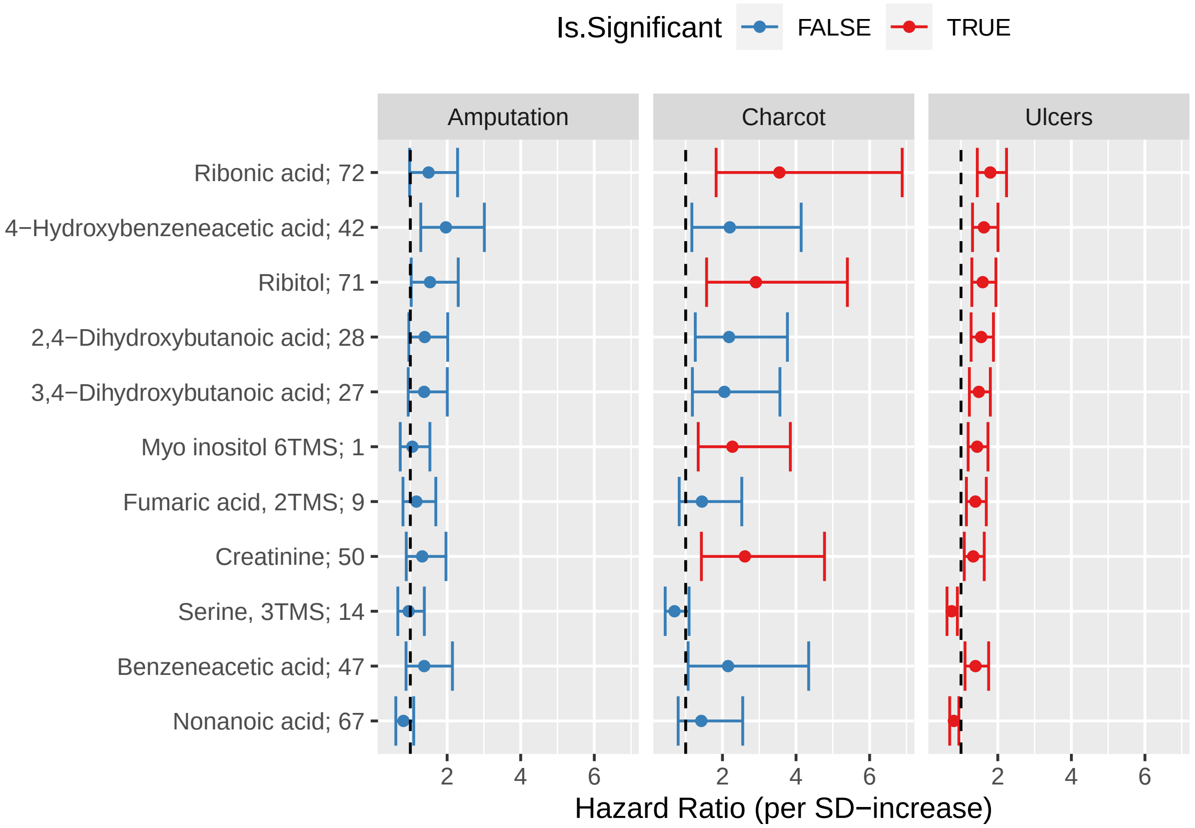

#### 7.2 Adjusted Models

##### 7.2.1 Cross-Sectional

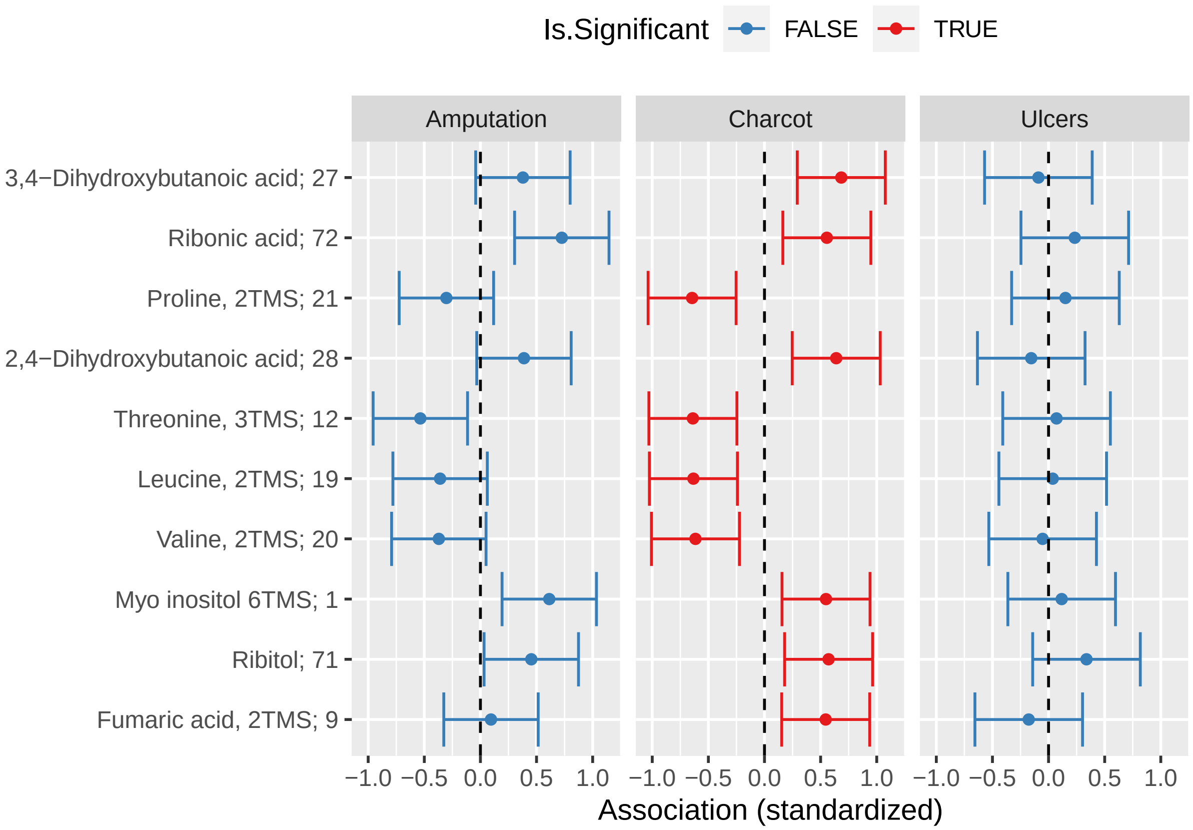

##### 7.2.2 Longitudinal

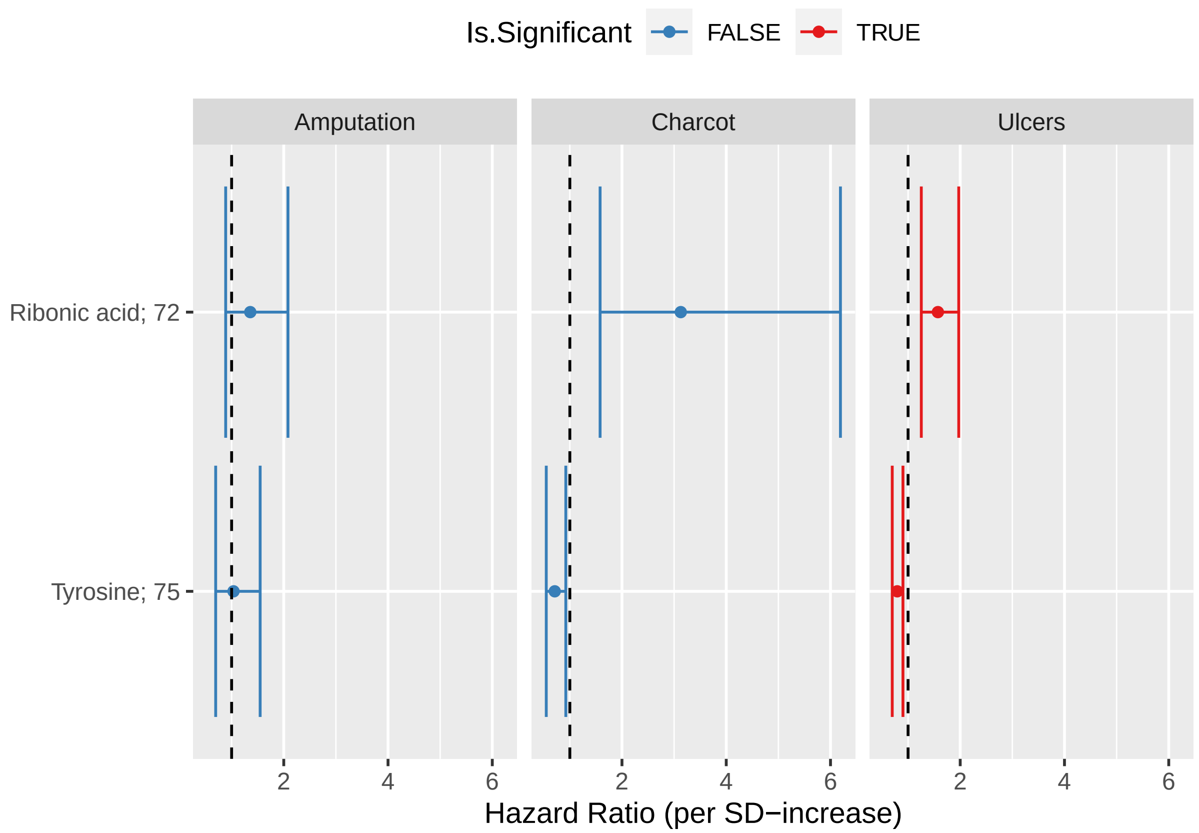
